# JAX-CNV: A whole genome sequencing-based algorithm for copy number detection at clinical grade level

**DOI:** 10.1101/2021.03.16.21252173

**Authors:** Wan-Ping Lee, Qihui Zhu, Xiaofei Yang, Silvia Liu, Eliza Cerveira, Mallory Ryan, Adam Mil-Homens, Lauren Bellfy, Kai Ye, Chengsheng Zhang, Charles Lee

## Abstract

We aimed to develop a whole genome sequencing (WGS)-based copy number variant (CNV) calling algorithm with the potential of replacing chromosomal microarray assay (CMA) for clinical diagnosis. JAX-CNV is thus developed for CNV detection from WGS. The performance of this CNV calling algorithm was evaluated in a blinded manner on 31 samples and compared to the results of clinically-validated CMAs. Comparing to 112 CNVs reported by clinically-validated CMAs of the 31 samples, JAX-CNV is 100% recalling them. Besides, JAX-CNV identified an average of 30 CNVs per individual that is an approximately seven-fold increase compared to calls of clinically-validated CMAs. Experimental validation of 24 randomly selected CNVs, showed one false positive (i.e., a false discovery rate of 4.17%). A robustness test on lower-coverage data revealed a 100% sensitivity for CNVs greater than 300 kb (the current threshold for College of American Pathologists) down to 10x coverage. For CNVs greater than 50 kb, sensitivities were 100% for coverages deeper than 20x, 97% for 15x, and 95% for 10x. We developed a WGS-based CNV pipeline, including this newly developed CNV caller JAX-CNV, and found it capable of detecting CMA reported CNVs at 100% sensitivity with about 4% false discovery rate. We propose that JAX-CNV could be further examined in a multi-institutional study to justify the transition of first-tier genetic testing from CMAs to WGS. JAX-CNV is available on https://github.com/TheJacksonLaboratory/JAX-CNV.

## Introduction

Copy number variants (CNVs) are known to play key roles in human evolution, genomic diversity, and disease susceptibility [1–5]. In addition, copy number changes have been reported to cause microdeletion and microduplication syndromes such as Williams syndrome, Prader-Willi syndrome, Angelman syndrome, and DiGeorge syndrome [5–9]. Various technologies including fluorescence *in situ* hybridization (FISH), PCR-based assays, chromosomal microarray assays (CMAs), and next-generation sequencing (NGS) have been developed in research and clinical laboratories for CNV detection. Since 2010, CMAs have been considered the first-tier test for patients with unexplained developmental delay or intellectual disability, autism spectrum disorders, and congenital anomalies [10,11].

Over the past decade, advances in NGS technologies have brought unprecedented improvements in throughput, speed, and cost of DNA sequencing. These improvements make whole genome sequencing (WGS) feasible for broad use in research, with its ability to detect many types of genetic variations, and promise to offer the potential of a single test that captures nearly all genomic variations in an unbiased manner [12–14]. Although several WGS-based CNV calling algorithms were developed [15–21], none of them has been widely accepted for clinical applications because of the high false discovery rate, often substantially greater than 5%.

In addition to developing new CNV-calling algorithms, integrative pipelines that combine multiple CNV-calling algorithms to improve accuracy and overcome limitations of individual performance are commonly used. For example, Zhou et al. [22] developed a method to integrate callsets of CNVnator [19] and Lumpy[16]. Noll et al. proposed SKALD [23], which is based on consensus, filtered calls from BreakDancer [24] and GenomeSTRiP [25]. Trost et al. [26] composed a pipeline that employs CNVnator and ERDS [27] for CNV identification. However, most of those pipelines are not open source, and applied filters that were specifically developed for individual projects and were not standardized. Thus, it is unknown whether the sensitivity and specificity of the pipelines are comparable to the standards in clinical diagnosis. Given current limitations, an algorithmic pipeline with sufficient sensitivity and specificity for clinical application is demanded.

Here, we present JAX-CNV, a newly developed WGS-based CNV calling algorithm. An evaluation of its performance was performed on WGS data from 31 patient samples and compared to callsets of the clinically-validated CMA at the Jackson Laboratory for Genomic Medicine (JAX-GM). The result suggests that JAX-CNV has high sensitivity (100%) necessary for diagnostic decisions and a low false discovery rate (4%). This algorithm could serve as a basis for the use of WGS, as a replacement for array-based clinical genetic testing.

## Methods

**Figure 1A** shows the overview of the WGS data analysis that includes three major steps, pre-processing, alignment and CNV calling.

**Figure 1:**
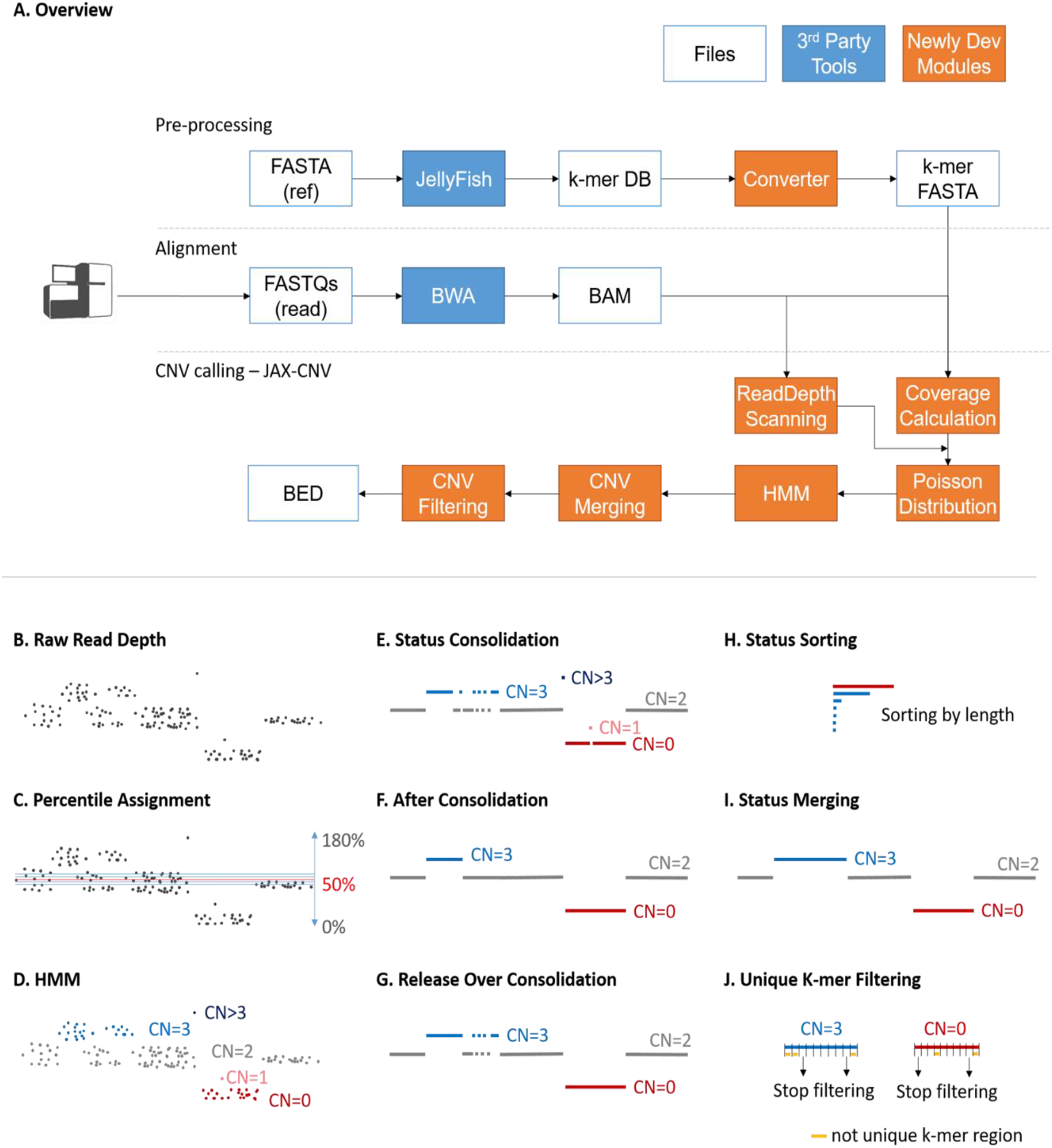
A. Overview of the CNV calling pipeline consists of three major steps: pre-processing, alignment and CNV calling. B-J are details of CNV calling approach.

### WGS Analysis Workflow: Pre-process

The pre-processing steps for a given reference genome (such as GRCh37, GRCh38, or other versions of human reference genomes) include BWA index (v0.7.15) and JELLYFISH [28] count (v2.2.6). BWA index creates the required files for BWA alignment, while JELLYFISH calculates the counts of each 25-mer of the reference genome and generated k-mer database (k-mer DB in **Figure 1A**) to indicate mappabilities of regions. This is the similar idea proposed in CLImAT-HET [29], using k-mer for low mappability region identification. Theoretically, larger k will increase the uniqueness of a k-mer against the given reference genome. However, that also requires larger computational resource to process larger k-mer. The experiment of different k-mer (k from 5 to 40) shows that 25-mer holds 76% unique k-mer (Supplementary Materials and Methods: K-mer selection) and maintains the final sensitivity. Using 30-mer could hold 3% more unique k-mer, but the final results were not improved by using 30-mer. To efficiently compress the file size, each number in JELLYFISH k-mer DB was converted to a character by calculating its log2 value added by 33 because ASCII code 33 in decimal is the first printable and visually character. For example, if a 25-mer has only one position in the genome, the log2(1) value is zero. For converting the zero to the first printable ASCII code, 33 was added to the original log2 value (k-mer FASTA in **Figure 1A**). BWA index, JELLYFISH count, and k-mer DB conversions may take 190, 105, and 403 minutes, respectively.

### WGS Analysis Workflow: Alignment

The analysis could start at raw FASTQ files or BWA aligned BAM/CRAM files. If FASTQ is given, FASTQC (v0.11.5) would be applied additionally in the pipeline for quality control. Then, BWA mem (v0.7.15) is employed for mapping reads against a given human reference genome. Once alignments are obtained (from our pipeline or given by users), alignments were sorted by SAMTOOLS and used as the input file of JAX-CNV. **Figure 1B** illustrated read depths after alignment.

### WGS Analysis Workflow: CNV calling: coverage calculation

JAX-CNV uses the log2(25-mer) FASTA-format file from the pre-processing step to scan the high-confident mapping (unique) regions for an accurate coverage calculation. A region is considered a high-confident mapping region when each 25-mer count in the region is one and the size of the region is larger than 20 kb. For each chromosome, we calculated an average coverage based on those 20 high-confident regions on a given BAM/CRAM of a sample. Once average coverages of all chromosomes were obtained, the interquartile range was applied to filter outliers. Outliers could indicate trisomy, monosomy, and other gross chromosome number anomalies of a sample. Then, an overall coverage of the sequenced sample was calculated based on average coverages of all chromosomes excluding the outliers. Using the interquartile range method, we were able to detect aneuploidies. For those aneuploidies, we will not detect any smaller CNVs on the respective chromosomes in the further steps.

### WGS Analysis Workflow: CNV calling: HMM

The overall coverage of the sequenced sample is used as the baseline and set to 50% percentile (**Figure 1C**). Next, we scanned the BAM/CRAM by shifting bins (the default bin size is set to 50 bp and it is user-adjustable by “−-bin”), and assigned a percentile for each bin according to the difference between the overall coverage and the read depth of the bin. The percentiles normally range from 0% to 180%, and 180% - 200% are reserved for copy numbers larger than two. For example, if the overall coverage is 50x and a read depth of a bin is 100x, the percentile of the bin will be 100% (100 / 50 ⨯ 50%; **Figure 1C**). Then, a Hidden Markov Model (HMM) with a Poisson distribution [30] was applied to convert the percentile of each bin to one of the five HMM CNV statuses: CN=0 (loss), CN=1 (loss), CN=2 (normal), CN=3 (gain) and CN>3 (gain) (**Figure 1D**). Afterwards, if the CNV statuses of the two adjacent bins are the same, we merged them into a segment (**Figure 1E**).

### WGS Analysis Workflow: CNV calling: CNV merging

Since the default bin size is set to 50 bp which is small compared to CNVs that we expect to detect, oscillations of CNV statuses could happen frequently as illustrated in **Figure 1E**. There is a small CN=1 status inside a large CN=0 region. Using larger bins may resolve oscillations, but it decreases sensitivity. To maintain high sensitivity, we set the default bin size is 50 bp, that is user-adjustable by “--bin”, followed by a merging step. The merging step is necessary to mitigate oscillations caused by uneven read depth problem that is common in WGS data. To show oscillations of CNV statuses and uneven read depth problem, Supplementary Figure S1 gives coverage, log2(25-mer), and ratio of low-quality alignments of the 112 truth CNVs.

If the length of a CNV status is shorter than 5 kb, then it will be absorbed by the previous status (**Figure 1F**) to form a cluster regardless of the CNV type (loss or gain). This status consolidation may be too aggressive to merge too many other CNV types together into a cluster, leading to incorrect results. Thus, for each status consolidation, if the original CNV type of a cluster covers less than 80% of the total length of the current cluster, the merging stops and reinstates the original status (**Figure 1G**). This approach also prevents small statuses to merge large statuses.

After the completion of merging, all CNV regions were sorted by their respective lengths (**Figure 1H**). From the largest to the smallest, each CNV region checked other CNVs of the same type (loss or gain) downstream and upstream from the nearest to the farthest coordinates for further clustering (**Figure 1I**). The above procedure stops when encountering a CNV with the different type (loss or gain). This step allows larger CNVs to cross normal status (CN=2) and to merge smaller CNVs nearby. Stating from the largest CNV enables larger CNVs having more opportunity to cross normal statuses and merge with other CNVs with the same type. Candidate CNVs were then generated.

### WGS Analysis Workflow: CNV filtering

For each candidate CNV obtained from the previous step, CNV calling: CNV merging, we divided it into ten bins of equal length. Each bin was assigned a uniqueness value corresponding to the count of unique k-mers (i.e., the number of 25-mers that has a unique position in the reference genome). Starting from the bin with the smallest coordinate (the most left) to the bin with the largest coordinate, we filtered a bin if the uniqueness value is low (percentage of unique k-mers is lower than 60% by default; user-adjustable by “--unique_kmer”). The procedure stops after encountering a low uniqueness bin (**Figure 1J**). The above procedure was repeated from the bin with the largest coordinate (the most right) to the bin with the smallest coordinate. This approach helps us to trim tails of CNVs in low-confident mapping regions.

Then, the density-based spatial clustering of applications with noise (DBSCAN) algorithm [31] was further employed to cluster the candidate CNV fragments as the final step to making a CNV call. For this final step, using DBSCAN helps us to have a global view of CNV regions and consolidate them better. Specifically, we first sorted the candidate CNV fragments based on their coordinates. Then, we separated the fragments into different raw clusters by two conditions: (1) the distances of any two continuous fragments are < 3 Mb, and (2) the types (loss or gain) of all fragments located in the raw cluster region. Next, for each raw cluster, we calculated the distance of every continuous fragment pairs. The mean distance of the raw cluster was also calculated. The DBSCAN function (in DBSCAN R package) was applied on the distance matrix of each raw cluster. The procedure ended when the cluster results are not change (Supplementary Materials and Methods: DBSCAN). We reported CNVs for a given individual in a BED-format file.

### Assessment of Pathogenicity of a CNV

The criteria and guidelines used for classification and interpretation of CNVs were published previously [10,11,32]. In general, a CNV is classified as pathogenic if it (1) overlaps genomic coordinates for a well-known deletion or duplication syndrome; (2) contains disease genes reported in GeneReviews (https://www.ncbi.nlm.nih.gov/books/NBK1116/), OMIM (https://www.omim.org/), or DECIPHER (https://decipher.sanger.ac.uk/disorders#syndromes/overview).

### Droplet Digital PCR (ddPCR) Validation

Droplet Digital PCR (ddPCR) assays were performed to examine the accuracy of the genomic aberrations detected by the JAX-GM clinical CMA platform (Supplementary Materials and Methods: Droplet Digital PCR (ddPCR) Validation) and the JAX-CNV algorithm. The customized assays utilized primers designed by Primer3Plus [33], based on the GRCh38 assembly. All primer pairs were tested for their uniqueness across the human genome using In-Silico PCR from UCSC Genome Browser. A BLAT search was also performed at the same time to make sure all primer candidates only hit one site of the human genome. Lastly, the NCBI 1000 Genomes Browser was used to check if there were any single nucleotide variations (SNVs) in the primer or probe-binding region. All primers and probes used in this study were listed in Supplementary Table S1.

## Results and Discussion

### Dataset

Currently, the microarray proficiency test offered by the College of American Pathologists (CAP) requests participating clinical laboratories to report CNVs greater than 300 kb [32]. We selected 31 samples associated with various constitutional disorders (i.e., DiGeorge syndrome, Williams syndrome, Cri-du-chat syndrome, Smith-Magenis syndrome, Wolf-Hirschhorn syndrome, Miller-Dieker Lissencephaly syndrome, Tetralogy of Fallot syndrome, 1p deletion syndrome, and Angelman syndrome) from the Coriell Institute. For 22 of these 31 samples, the Coriell Institute reports a total of 45 CNVs (25 deletions and 20 duplications, ranging from 101 kb to 94 Mb in size) (**Table 1**; Supplementary Table S2).

**Table 1.** A comparison of CNV calls of JAX-CNV on the 45 Coriell-registered CNVs. In the Pathogenic Annotation column, ‘G’, ‘D’ and ‘M’ mean annotations from GeneReviews, Decipher, and OMIM databases, respectively. In the JAX-CNV columns, ‘+’ denotes CNVs captured by the methods/coverages, and ‘*’ denotes that CNVs are not 50% reciprocal overlapping but recovered in manual review. Shadowed cells mean no call.

| Coriell IDs | Coriell Description | CNV Region | CNV Type | Pathogenic Annotation | JAX-CNV |  |  |  |  |
| --- | --- | --- | --- | --- | --- | --- | --- | --- | --- |
|  |  |  |  |  | Original_coverage (30-48x) | 30x | 20x | 15x | 10x |
| GM02820 | Chromosome aberration | 9p24.3p13.3 (34.5 Mb) | DUP | G/M | + | + | + | + | + |
|  |  | 12q24.32 q24.33 (7.3 Mb) | DEL | G/M | + | + | + | + | + |
| GM03997 | Derivative chromosome | 5q35.1 (130 Kb) | DUP | M | + | + | + | + | + |
|  |  | 12p13.33p12.2 (20.8 Mb) | DUP | G/D/M | + | + | + | + | + |
|  |  | 12q24.33 (623 Kb) | DEL | G/M | + | + | + | + | + |
| GM05876 | DiGeorge syndrome | 22q11.21 (1.4 Mb) | DEL | G/D/M | + | + | + | + | + |
| GM09025 | Ring chromosome | 16q24.2 (383 Kb) | DUP | G/M | + | + | + | + | + |
|  |  | 22q13.31q13.33 (2.9 Mb) | DEL | G/D/M | + | + | + | + | + |
| GM09209 | Miller-Dieker Lissencephaly syndrome | 17p13.3 (5.9 Mb) | DEL | G/D/M | + | + | + | + | + |
| GM09687 | Recombinant chromosome | 16p13.3 (1.1 Mb) | DEL | G/D/M | + | + | + | + | + |
|  |  | 16q22.1q24.3 (20 Mb) | DUP | G/M | + | + | + | + | + |
| GM09711 | Dicentric chromosome | 2q13 (140 Kb) | DUP | G/M | + | + | + | + | * |
|  |  | 13q11q34 (94 Mb) | DUP | G/M | + | + | + | + | + |
|  |  | 13q34 (1.7 Mb) | DEL | M | + | + | + | + | + |
| GM10946 | Recombinant chromosome | 6p21.2p21.1 (964 Kb) | DUP | G/M | + | + | + | + | + |
|  |  | 6p12.3 (780 Kb) | DUP |  | + | + | + | + | + |
|  |  | 6q14.1q16.3 (25 Mb) | DEL | G/M | + | + | + | + | + |
| GM11428 | Duplicated chromosome | 3p26.3p26.2 (5.3 Mb) | DEL | G/M | + | + | + | + | + |
|  |  | 3q22.1q26.1 (29.8 Mb) | DUP | G/D/M | + | + | + | + | + |
|  |  | 3q26.1 (112.8 Kb) | DEL |  | + | + | + | + | + |
|  |  | 3q26.1q29 (35.2 Mb) | DUP | G/M | + | + | + | + | + |
| GM11516 | Angelman syndrome | 15q11.2q13.1 (7 Mb) | DEL | G/D/M | + | + | + | + | + |
| GM13480 | Williams syndrome | 7q11.23 (1.6 Mb) | DEL | G/D/M | + | + | + | + | + |
|  |  | 9p24.1 (107.6 Kb) | DUP |  | + | + | + | + | + |
| GM13590 | Duplicated chromosome | 2q11.2q21.1 (33.6 Mb) | DUP | G/M | + | + | + | + | + |
|  |  | 2q37.3 (119 Kb) | DEL |  | + | + | + | + | + |
|  |  | 4q31.22 (101.5 Kb) | DEL | M | + | + | + | + | + |
|  |  | 9p13.3 (120.3 Kb) | DUP | G/M | + | + | + | * | * |
|  |  | 17q11.1 (101 Kb) | DUP | M | + | + | + | + | + |
| GM13946 | Williams syndrome | 7q11.23q11.23 (1.6 Mb) | DEL | G/D/M | + | + | + | + | + |
| GM14164 | Tetralogy of fallot | 13q14.2 (47.9 Mb) | DEL | G/M | + | + | + | + | + |
|  |  | 22q11.21 (148.8 Kb) | DUP | M | + | + | + | + |  |
| GM16580 | 18p deletion syndrome | 18p11.32 (1.6 Mb) | DEL | M | + | + | + | + | + |
|  |  | 18q21.33q23 (13.5 Mb) | DUP | M | + | + | + | + | + |
|  |  | 18q23 (4.0 Mb) | DEL | G/M | + | + | + | + | + |
| GM16593 | Cri-du-chat syndrome | 5p15.3 (14.7 Mb) | DEL | G/M | + | + | + | + | + |
|  |  | 14q24.3 (2.7 Mb) | DEL | M | + | + | + | + | + |
| GM18828 | Chromosome aberration | 1q31.3 (118 Kb) | DUP | G/M | + | + | + | + |  |
|  |  | 4p16.1 (140 Kb) | DUP | M | + | + | + | + | + |
| GM20200 | Isodicentric chromosome | 1q31.3 (103 Kb) | DEL | G/M | + | + | + | + | + |
|  |  | 15q11.1q13.1 (8.5 Mb) | DUP | G/D/M | + | + | + | + | + |
| GM20375 | Angelman syndrome | 15q11.2q13.1 (4.9 Mb) | DEL | G/D/M | + | + | + | + | + |
| GM20743 | Smith-Magenis syndrome | 17p11.2 (2.1 Mb) | DEL | G/D/M | + | + | + | + | + |
| GM22569 | 1p deletion syndrome | 1p36.33 (5.5 Mb) | DEL | G/M | + | + | + | + | + |
| GM22601 | Wolf-Hirschhorn syndrome | 4p16.3 (25.0 Mb) | DEL | G/D/M | + | + | + | + | + |

These 31 samples were also examined with a clinically-validated Affymetrix CytoScan HD platform (Affymetrix, Santa Clara, CA) for the detection of chromosomal imbalances following the standard operating procedures of the CLIA-certified laboratory at JAX-GM. The CMA data analysis was performed using the software supplied by the vendor (ChAS v3.3, Methods: Affymetrix CytoScan HD Analysis Flow). The JAX-GM clinically-validated CMA platform reported an additional 67 CNVs among these 31 individuals, using a size cutoff of 50 kb (Figure 2, Supplementary Table S3). In total, those 112 CNVs (65 deletions and 47 duplications, ranging from 51 kb to 94 Mb in size) were used as the truth set and set an initial baseline for sensitivity analysis. Of note, 70 of the 112 CNVs were considered to be pathogenic based on the previously-published criteria and guidelines [32,34] (41 of 45 Coriell registered CNVs and 29 of the 67 additional CNVs detected by the JAX-GM CMA).

**Figure 2.**
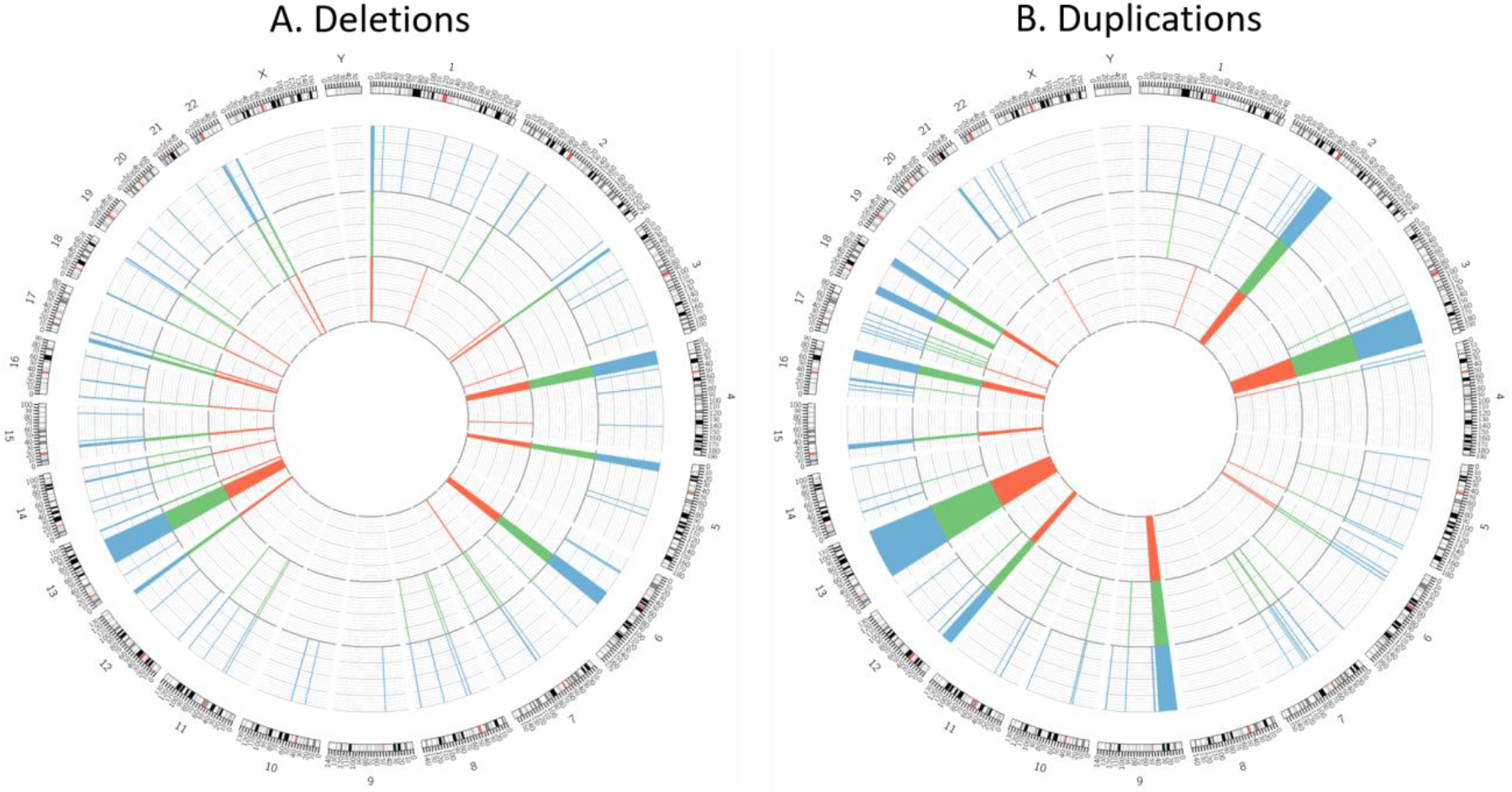
Distribution of CNVs in the 31 test samples. The sample were identified by three different methods (Coriell Institute CMA, red inner circle; JAX-GM CMA, green middle circle, and the JAX-CNV algorithm, blue outer circle). The JAX-CNV accurately detected all Coriell-registered and CMA-reported CNVs from WGS data of the 31 individuals.

### Sensitivity Assessment

We performed WGS on these 31 samples by Illumina paired-end sequencing at 30x - 48x coverage with the read length of 2×150bp (Supplementary Table S5) in a blinded study manner. BWA was applied for short read alignment against GRCh38 human reference genome (chr1-22, X, Y, and M), followed by analysis using the JAX-CNV algorithm for CNV identification. JAX-CNV accurately detected all 112 CNVs of the truth set from WGS data (**Table 1**, Figure 2). Figure 2 is a figure to show aggregated CNVs of all samples, and the locations of CNVs of each sample are given in Supplementary Table S4 and Figure S2. Of note, due to the different resolutions of CMAs and WGS, there were three deletions ranging from 104 kb to 291 kb in size (GM20375 and GM20743×2) and three duplications ranging from 105 kb to 292 kb in size (GM09687, GM13480 and GM20743) that did not meet the benchmark of 50% reciprocal overlap with the JAX-CNV calls, but they were still located in the same regions with either smaller or larger size ones (Supplementary Figure S3).

Figure 3A summarizes the calls from JAX-CNV and compares these calls with the truth set. JAX-CNV detected an additional 747 more CNVs than the array-based technology, and 89% of these calls were less than 300 kb in size and 50% were less than 100 kb in size. Figure 3B further summarizes CNV calls of each sample. JAX-CNV used 119 calls to identify 112 CNVs in the truth set. For example, for a 20.8 Mb duplication at chromosome regions 12p13 - 12p12 in GM03997, JAX-CNV made two CNV calls (12.3 Mb and 8.5 Mb) across the 20.8 Mb duplicated region (Supplementary Figure S4).

**Figure 3.**
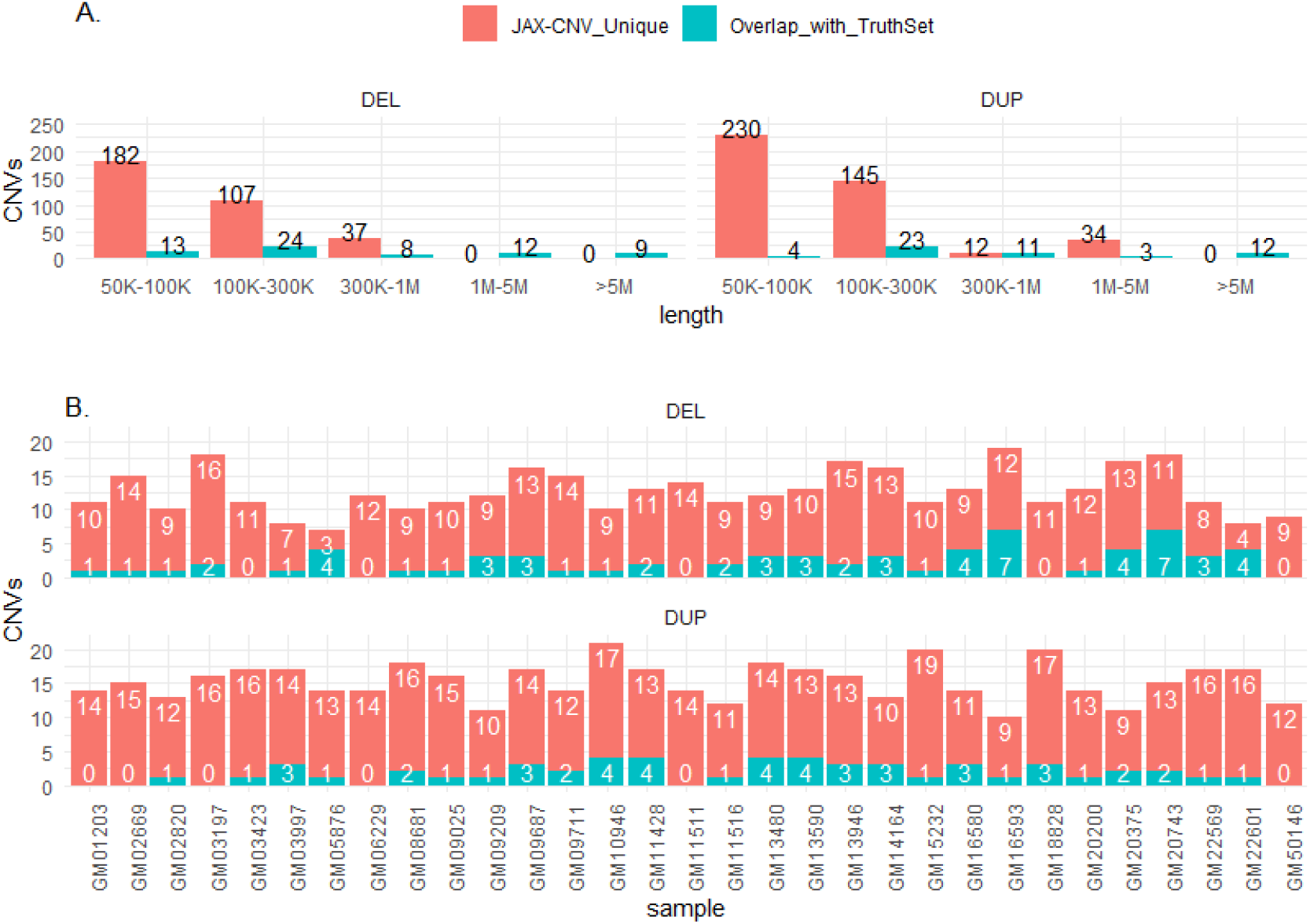
The concordance of CNV calls between CNV calls made with the JAX-CNV algorithm on WGS data compared to the array-based truth set. **A**. The unique JAX-CNV calls (in red), and CNV calls overlapping with the truth set (in green), as a function of CNV sizes. **B**. Concordance of CNV calls per sample.

### False Discovery Rate Assessment

As described above, JAX-CNV detected 747 more CNVs (across 31 individuals), which resulted in an approximately seven-fold increase of CNV calls compared to the truth set (Supplementary Table S4). To assess the accuracy of those additional calls, we randomly selected two samples, GM05876 and GM09209, for experimental validation. Compared to the truth set, there were 8 concordant CNV calls and 16 novel CNV calls in the two samples. Fourteen of the 16 novel CNV calls were validated by ddPCR (Supplementary Table S6). Among the invalidated CNV calls, one 1450.3 kb loss at 21p11 was inconclusive due to an unclear separation of positive and negative droplet clusters in the ddPCR assay, and one duplication at 16p11 (encompassing a segmental duplication and a simple repeat-rich region) was found to be a false positive from JAX-CNV. Of note, 28 among the 31 samples, JAX-CNV reports duplications at 16p11 because of noise signal in the region. We did not simply filter the region because JAX-GM CMA identified a duplication in the region for GM16580. As a result, there were 15 CNVs (out of 24, greater than 50 kb in size), that were missed by the clinically-validated CMA and one false positive CNV call among the 24 randomly chosen JAX-CNV-based CNV calls. This represents a 100% sensitivity for CNV calls greater than 50 kb with a false discovery rate of 4.17% (1/24) for JAX-CNV calls.

### CNV Detection on Low-Coverage WGS

Although the cost for NGS sequencing has dropped rapidly, its price still remains a big concern when WGS is considered as a first-tier assay in clinical diagnostics. To address this issue and assess the ability of JAX-CNV to accurately detect CNVs on low-coverage WGS data sets, we down-sampled the WGS data and assessed CNV-calling sensitivity of JAX-CNV. These 31 test samples were originally sequenced at coverages ranging from 30x to 48x (Supplementary Table S5). The simulation of different coverages was performed by SAMBAMBA [35] on the aligned BAM files. The test series consisted of down-sampling to 30x, 20x, 15x and 10x from the original 30x - 48x WGS coverage data. JAX-CNV was then applied on WGS data of different coverage depths. The assessments on 9x and 8x were also performed; however, the loss of sensitivity on those coverages is not accepted. Thus, 10x is the lowest coverage that JAX-CNV could maintain the sensitivity.

Among the 112 CNVs of the truth set, 50 were larger than 300 kb (CAP standard cutoff size). Even when the coverage was reduced down to 10x, JAX-CNV remained 100% sensitive for detecting CNVs greater than 300 kb (Figure 4). At a sequencing coverage of 15x, JAX-CNV failed to identify a 79.9 kb duplication at chromosome region 17q21.31 of GM05876 (Supplementary Figure S5). At a sequencing coverage of 10x, JAX-CNV failed to additionally identify a 55 kb deletion at chromosome region 11p11.12 of GM09687 and five duplications, ranging from 52 kb to 204 kb in size (Figure 4, Supplementary Figure S6). In summary, for deletions larger than 50 kb, the sensitivities are 100% (30x, 20x and 15x) and 98.5% (10x), while for duplications larger than 50 kb, the sensitivities are 100% (30x and 20x), 97.9% (15x) and 89.4% (10x).

**Figure 4.**
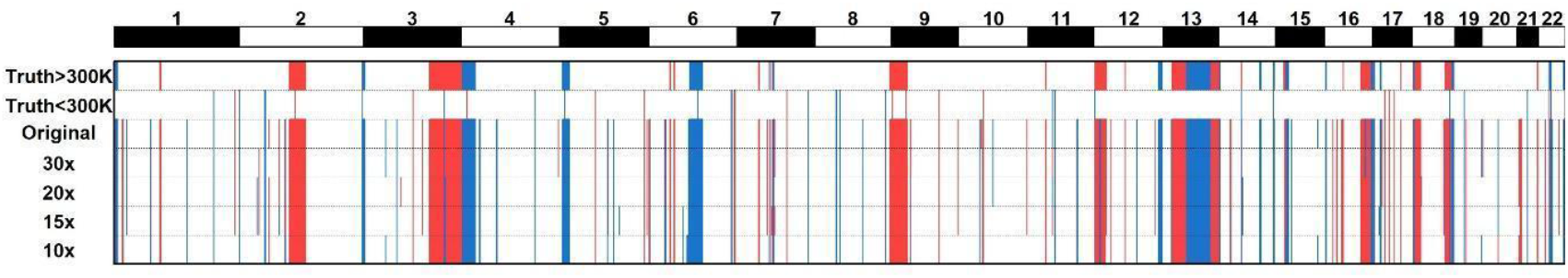
CNV detection on WGS data at five different sequencing coverages. The top two panels are the truth set using 300 kb as the cutoff. Red-colored bars indicate deletions while blue-colored bars indicate duplications.

### Comparison with Other WGS-Based Algorithms

We compared JAX-CNV to other CNV calling algorithms, Manta [15], Lumpy [16], Delly [17], CNVnator [19] and cn.MOPS [18], using the same WGS data. The scripts of performing those algorithms can be found in Supplementary Materials and Methods Scripts. In addition, we also tested combined methods (pipelines) for detection of CNVs, such as FusorSV [36] and MetaSV [37]. The sensitivity assessment of these two pipelines failed since they do not fully support GRCh38 or running time was longer than two days.

For the 112 CNVs of the truth set, Manta, Lumpy, Delly, CNVnator and cn.MOPS identified 66, 61, 77, 100 and 20 of the CNVs, that yields sensitivities of 59%, 54%, 69%, 89%, and 18%, respectively (Supplementary Figure S7). Some of the algorithms also incorrectly identified the CNV type (e.g., a deletion instead of a duplication). In general, read-depth-based algorithms, such as JAX-CNV and CNVnator, have greater sensitivity in detecting genomic imbalances. Other algorithms that primarily use paired-end, split-read or combinations of these strategies clearly showed lower sensitivity in identifying large chromosomal imbalances. Moreover, Manta, Lumpy and Delly identified tens of thousands of CNVs (39781, 35886, and 350183) greater than 50 kb in size (Supplementary Figure S7). Such big amount of reported CNVs poses a huge challenge for efficiently conducting clinical diagnosis and decision-making with these CNV algorithms.

With respect to speed, JAX-CNV took less than an hour to finish CNV detection for a given sample (range of 29 - 47 minutes for each sample) by using a single thread. Comparatively, Manta, Lumpy, Delly, CNVnator and cn.MOPS took 6 hours, 6.1 hours, 8.2 hours, 1.1 hours and 1 hour, respectively, to finish calling CNVs for a single sample. Analyses with JAX-CNV were completed using 4.5 GB of memory.

### Discussion

Since WGS-based assays are able to detect all types of genetic variations (SNVs, INDELs, and SVs), it has the potential to eventually supplant karyotyping, CMAs, and exome sequencing for disease diagnosis. Despite the growing applications of WGS-based assays for the detection of SNVs and INDELs in clinical settings [38–41], it still remains challenging to reliably detect CNVs for clinical diagnostics. While a number of CNV detection pipelines have been developed for research studies and further employed for clinical applications [26,34,42], none of them has been widely used for clinical diagnosis.

For research purpose pipelines, sacrificing specificity for sensitivity is beneficial to avoid missing potential CNVs. However, reporting hundreds, or even thousands of chromosomal aberrations is impractical for clinical diagnosis both due to time constraints and the complexity of evaluating a large number of CNVs with unknown significance. Without careful clinical considerations, it is impossible for those tools to meet the sensitivity, specificity and turnaround time requirements for disease diagnosis. Thus, there is still a need to develop novel or ameliorate existing bioinformatics tools and/or pipelines to improve the accuracy and turnaround time of the WGS-based assay for clinical applications. Hence, we developed a WGS-based CNV caller, JAX-CNV. The performance assessment showed a 100% sensitivity for CNVs larger than 50 kb.

Currently, the HMM in JAX-CNV has five statuses of CNVs: CN=0 (loss), CN=1 (loss), CN=2 (normal), CN=3 (gain) and CN>3 (gain). Thus, JAX-CNV can only indicate that a CNV status is larger than three if it is. To indicate the exact copy number that greater than three, JAX-CNV needs to check BAMs/CRAMs for read depth of the detected CNV region and detect the exact copy number in the region.

Since pathogenic chromosomal abnormalities are not limited to CNVs. Other structural variants (SVs) including translocations and inversions can also cause diseases [32]. We are currently developing new modules based on the established pipeline to accurately identify translocations and inversions. Detecting translocations and inversions is more difficult than detecting CNVs and a major reliance on the read depth signal as employed in the current CNV caller will not be effective. Thus, we are considering inclusion of paired-end alignment distance and orientation as inputs for identifying translocations and inversions in the next version of this pipeline. Breakpoints of inversions may be associated with deletions and further increase the detection difficulty. Nevertheless, previous studies have successfully detected translocations and inversions in WGS data [43]. Thus, we believe that with a careful design that reflects advanced knowledge of SVs, our future pipeline will become a comprehensive SV caller for clinical applications.

JAX-CNV is a newly developed WGS-based CNV algorithm for detecting deletions and duplications that are larger than 50 kb. The results obtained from the 31 Coriell samples demonstrated a 100% concordance between JAX-CNV calls, and the calls registered by Coriell and calls made by a clinical CMA platform. In addition to the high sensitivity and specificity, JAX-CNV is easy-to-use, stable, robust, and fast for detecting CNVs in WGS data. JAX-CNV requires 4.5 GB of memory and finishes CNV detection for a single sample in less than an hour. JAX-CNV meets the sensitivity, specificity, reproducibility, and speed requirements necessary for clinical applications, and demonstrates the potential to supplant CMA-based methods as the first-tier diagnostic assay.

## Data Availability

All the test samples were bought from Coriell Institute and the analysis tool is available on github.

https://github.com/TheJacksonLaboratory/JAX-CNV

## Data Availability

## Code Availability

Project name: JAX-CNV

Project home page: https://github.com/TheJacksonLaboratory/JAX-CNV

Operating system(s): Linux

Programing language: C++

Other requirements: N/A

License: https://github.com/TheJacksonLaboratory/JAX-CNV/blob/master/LICENSE

Any restrictions to use by non-academics: No restrictions

## CRediT (Contributor Roles Taxonomy) author statement

**Wan-Ping Lee**: Conceptualization, Methodology, Software, Formal analysis, Investigation, Data Curation, Writing - Original Draft, Writing - Review & Editing, Visualization, **Qihui Zhu**: Validation, Data Curation, Writing - Original Draft, Writing - Review & Editing, **Xiaofei Yang**: Methodology, Software, **Silvia Liu**: Methodology, Visualization, **Eliza Cerveira**: Validation, Data Curation, Writing - Original Draft, **Mallory Ryan**: Validation, Data Curation, **Adam Mil-Homens**: Validation, Data Curation, **Lauren Bellfy**: Validation, Data Curation, **Kai Ye**: Resources, Supervision, **Chengsheng Zhang**: Conceptualization, Methodology, Validation, Resources, Writing - Original Draft, Supervision, **Charles Lee**: Conceptualization, Methodology, Resources, Writing - Original Draft, Writing - Review & Editing, Supervision, Project administration, Funding acquisition.

## Acknowledgements

We would like to thank Drs. Zirui Dong and Yukyung Jun for their works on Figures 2 and 4. We would also like to thank Drs. Jee Young Kwon and Won Yeong Kang for their work on manuscript revision.

This study is supported in part by the operational funds from The First Affiliated Hospital of the Xi’an Jiaotong University. WL was supported by the National Science Foundation of China (61901352) and is supported by National Institute of Health, USA (U24AG041689 and U54AG052427). XY and KY are supported by the National Science Foundation of China (61702406 and 31671372), the National Science and Technology Major Project of China (grant number 2018ZX10302205), the National Key R&D Program of China (2018YFC0910400 and 2017YFC0907500), and the General Financial Grant from the China Postdoctoral Science Foundation (2017M623178). CL was a distinguished Ewha Womans University Professor supported in part by the Ewha Womans University Research grant of 2018-2019.

