## Supplemental Figures for "JAX-CNV: A whole genome sequencing-based algorithm for copy number detection at clinical grade level"

1. Precision Medicine Center, The First Affiliated Hospital of Xi’an Jiaotong University, Xi'an 710061, China
2. The Jackson Laboratory for Genomic Medicine, Farmington, CT 06032, USA
3. School of Cyber Science and Engineering, Xi'an Jiaotong University, Xi'an 710049, China
4. Department of Pathology and Laboratory Medicine, Perelman School of Medicine, University of Pennsylvania, Philadelphia, PA 19104, USA
5. School of Computer Science and Technology, Faculty of Electronic and Information Engineering, Xi'an Jiaotong University, Xi'an 710049, China
6. MOE Key Lab for Intelligent Networks & Networks Security, Faculty of Electronic and Information Engineering, Xi'an Jiaotong University, Xi'an 710049, China
7. Department of Life Sciences, Ewha Womans University, Seoul 03760, South Korea
8. Corresponding authors

*: These authors contributed equally to this work.

### Figure S1

112 sub figures of Figure S1 for 112 CNVs in the truth set.

Each sub figure consists of three panels that are, from the top to bottom, (1) Read Depth colored gray, (2) log2(k-mer_count) colored red, and (3) Ratio of low-quality alignment colored blue.

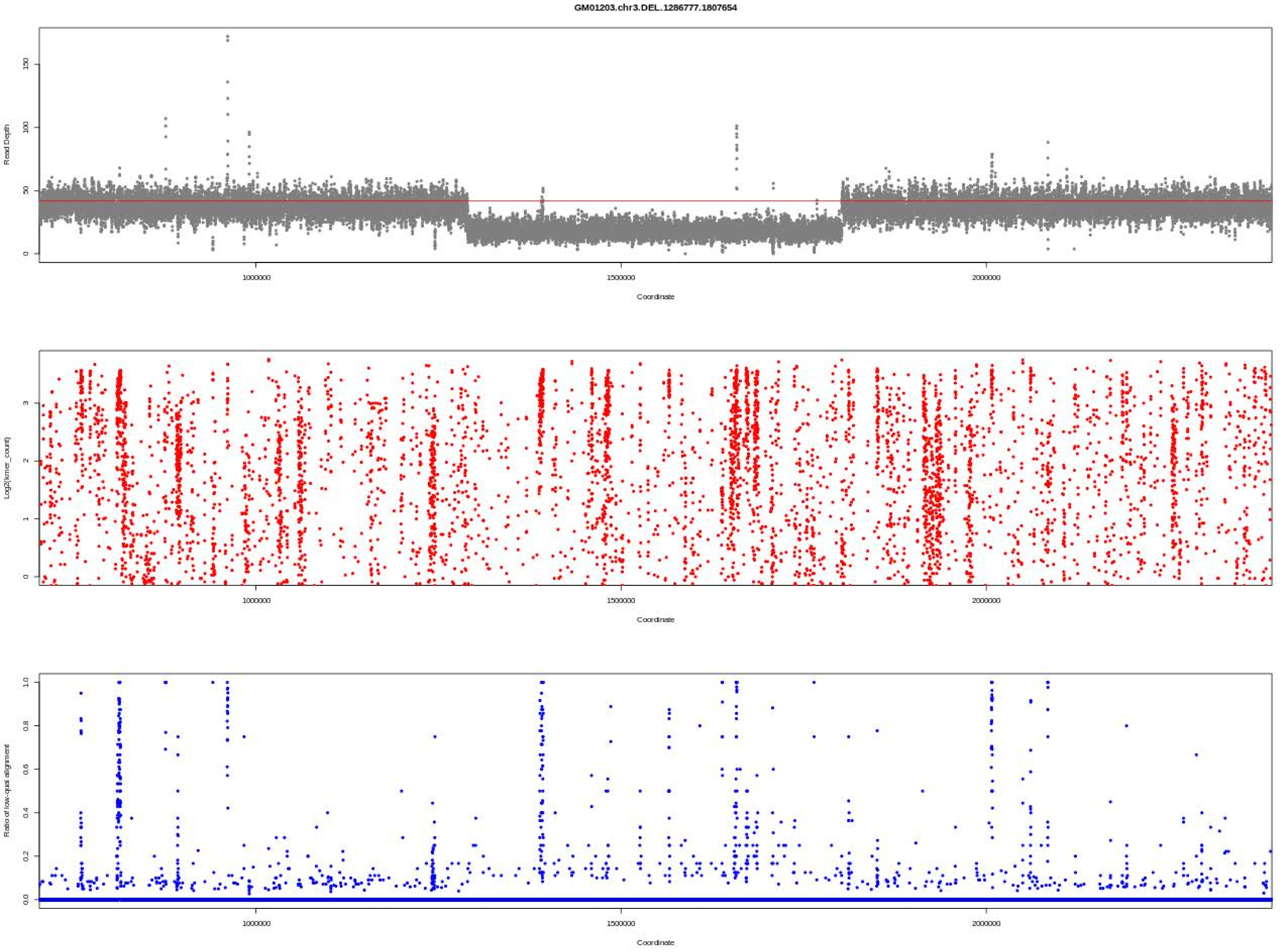
GM01203.chr3.DEL.1286777.1807654
Read Depth
Log2(kmer_count)
Ratio of low-qual alignment
Coordinate
1000000
1500000
2000000

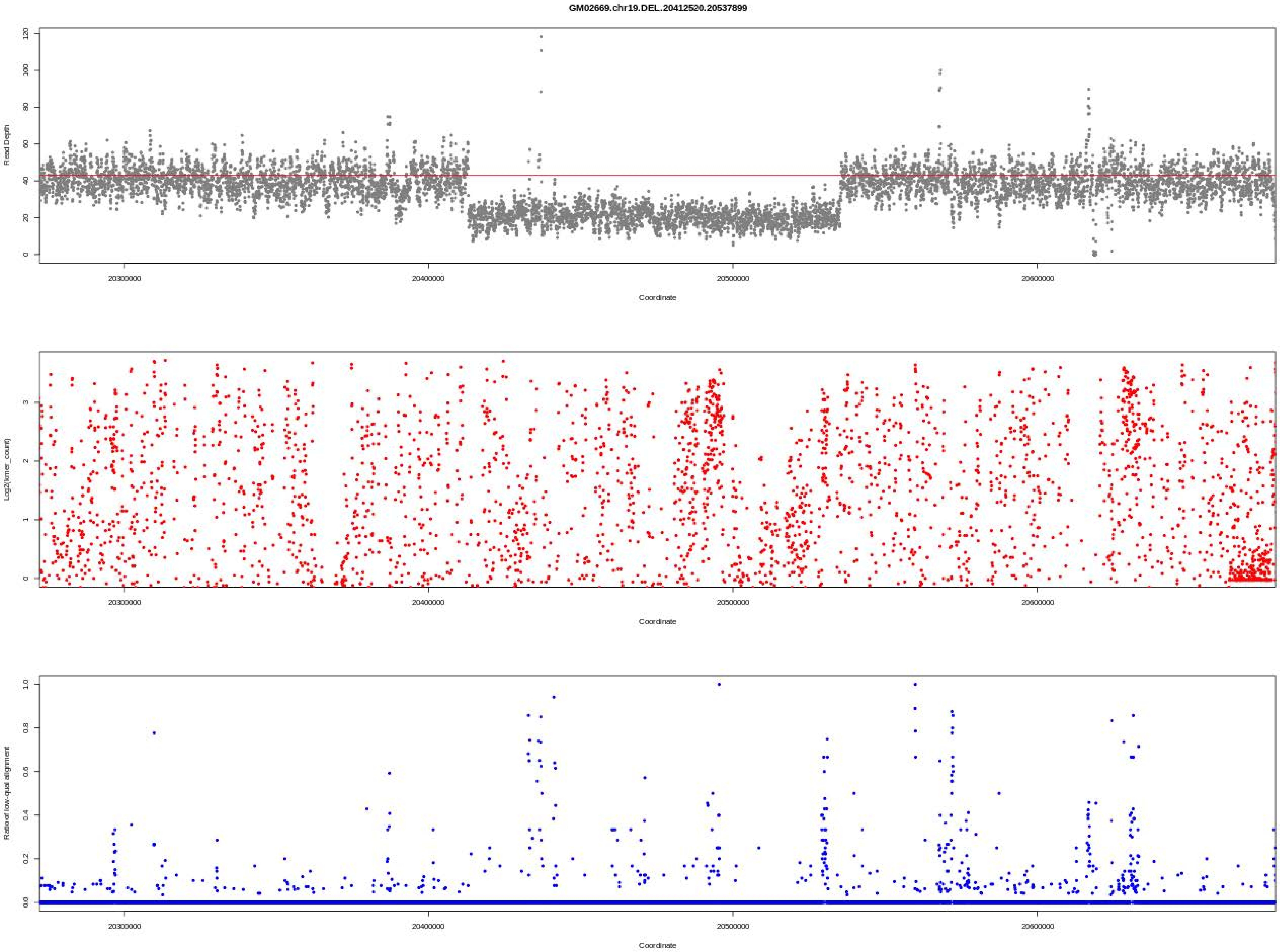
GM02669.chr19.DEL..20412520.20537899
Read Depth
Coordinate
Log2(kmer_count)
Coordinate
Ratio of low-qual alignment
Coordinate
20300000
20400000
20500000
20600000

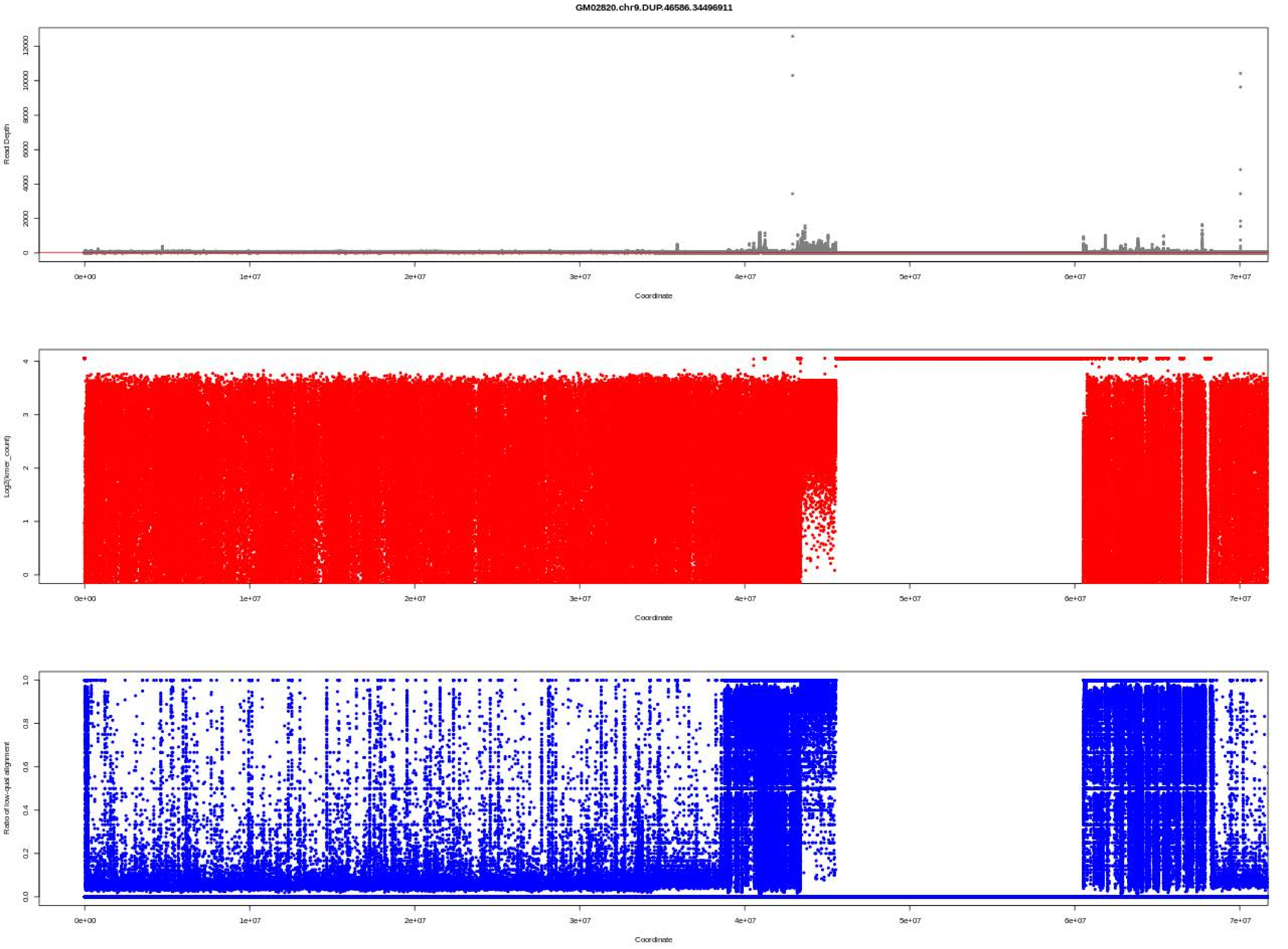
GM02820.chr9.DUP.46586.34496911
Read Depth
Coordinate
Log2(kmer_count)
Coordinate
Ratio of low-qual alignment
Coordinate

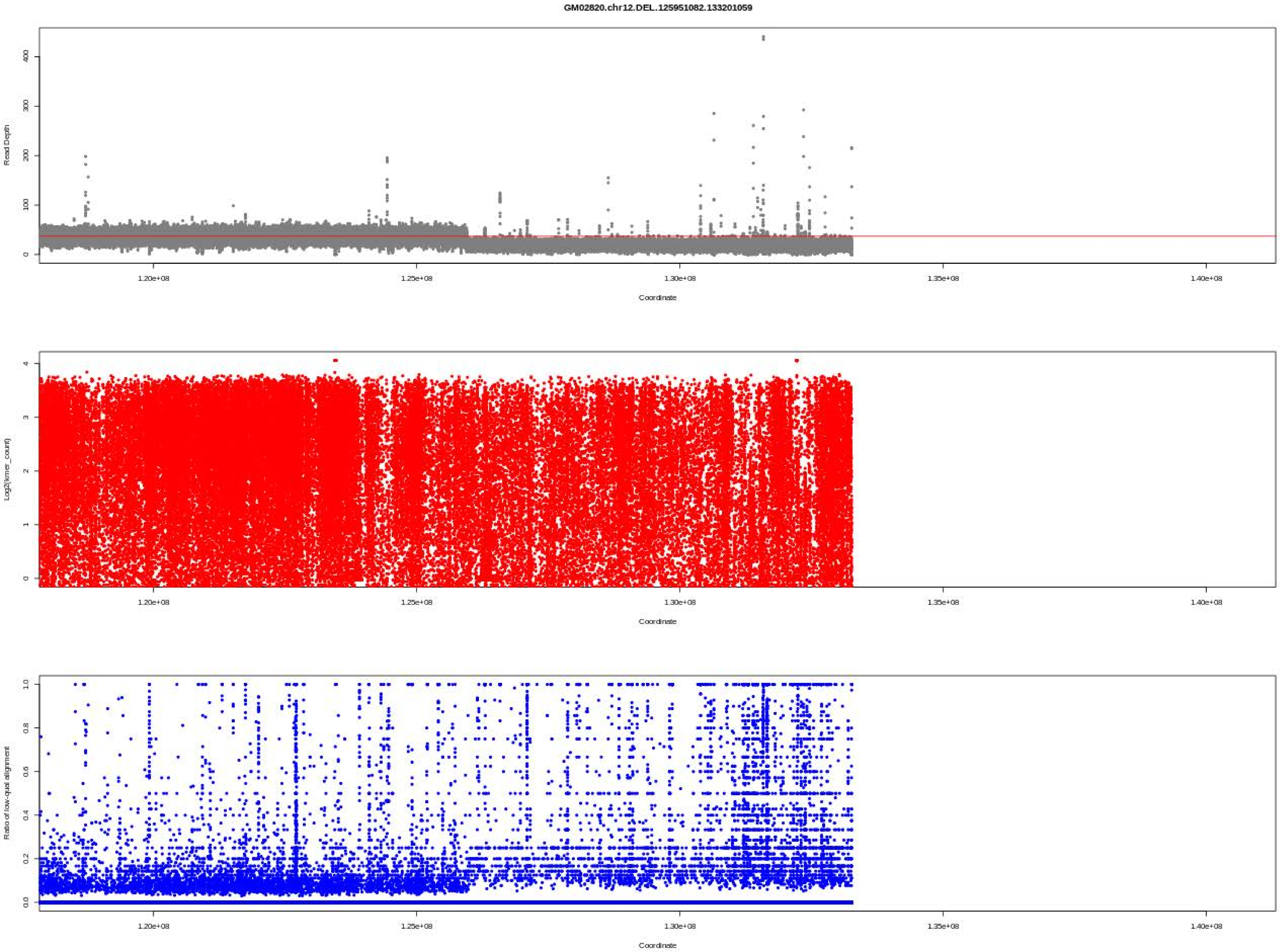
GM02820.chr12.DEL.125951082.133201059
Read Depth
0
100
200
300
400
Log2(kmer_count)
0
1
2
3
4
Ratio of low-qual alignment
0.0
0.2
0.4
0.6
0.8
1.0
1.20e+08
1.25e+08
1.30e+08
1.35e+08
1.40e+08
Coordinate

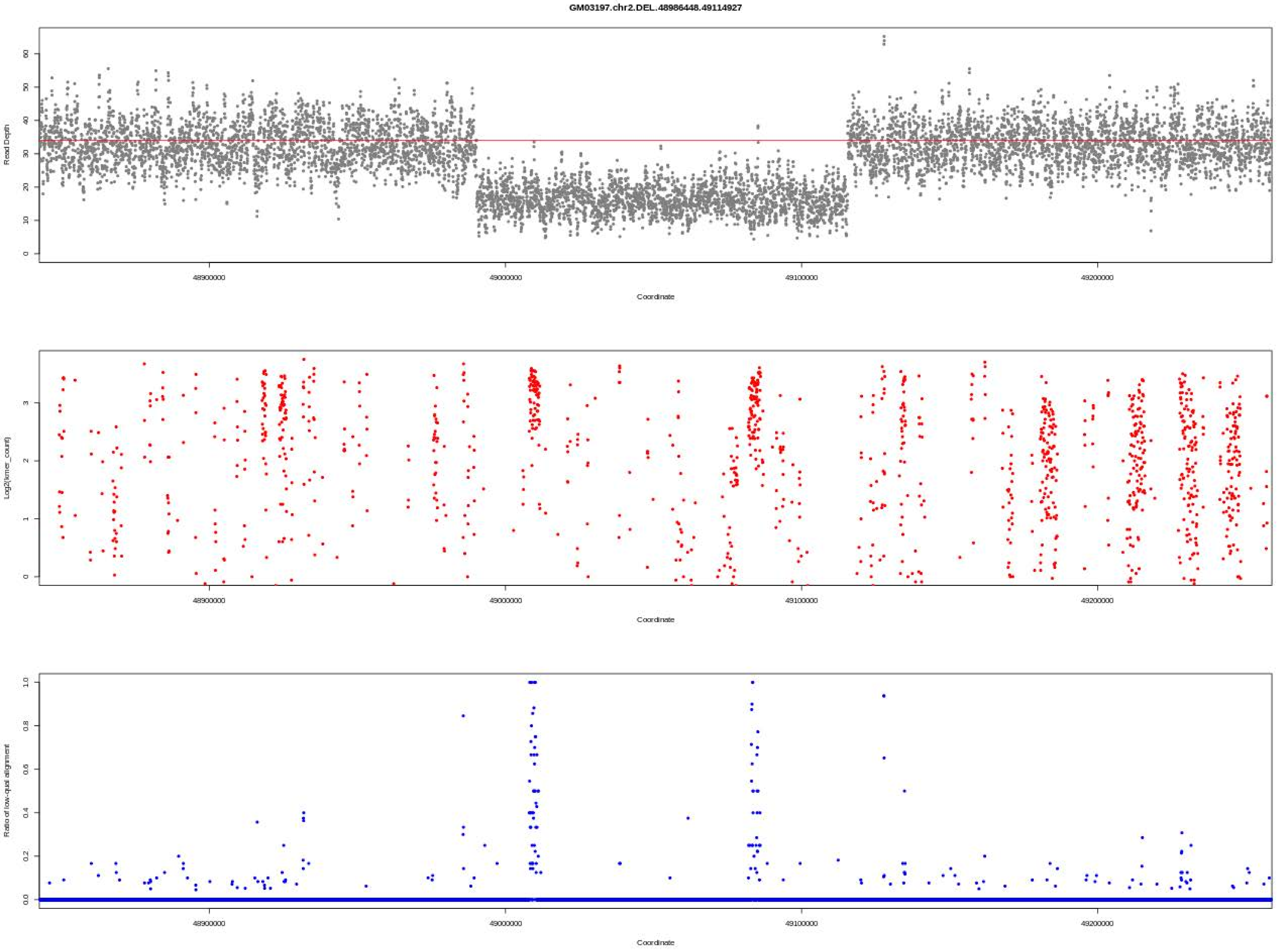
GM03197.chr2.DEL.48986448.49114927
Read Depth
Coordinate
Log2(kmer_count)
Coordinate
Ratio of low-qual alignment
Coordinate
48900000
49000000
49100000
49200000

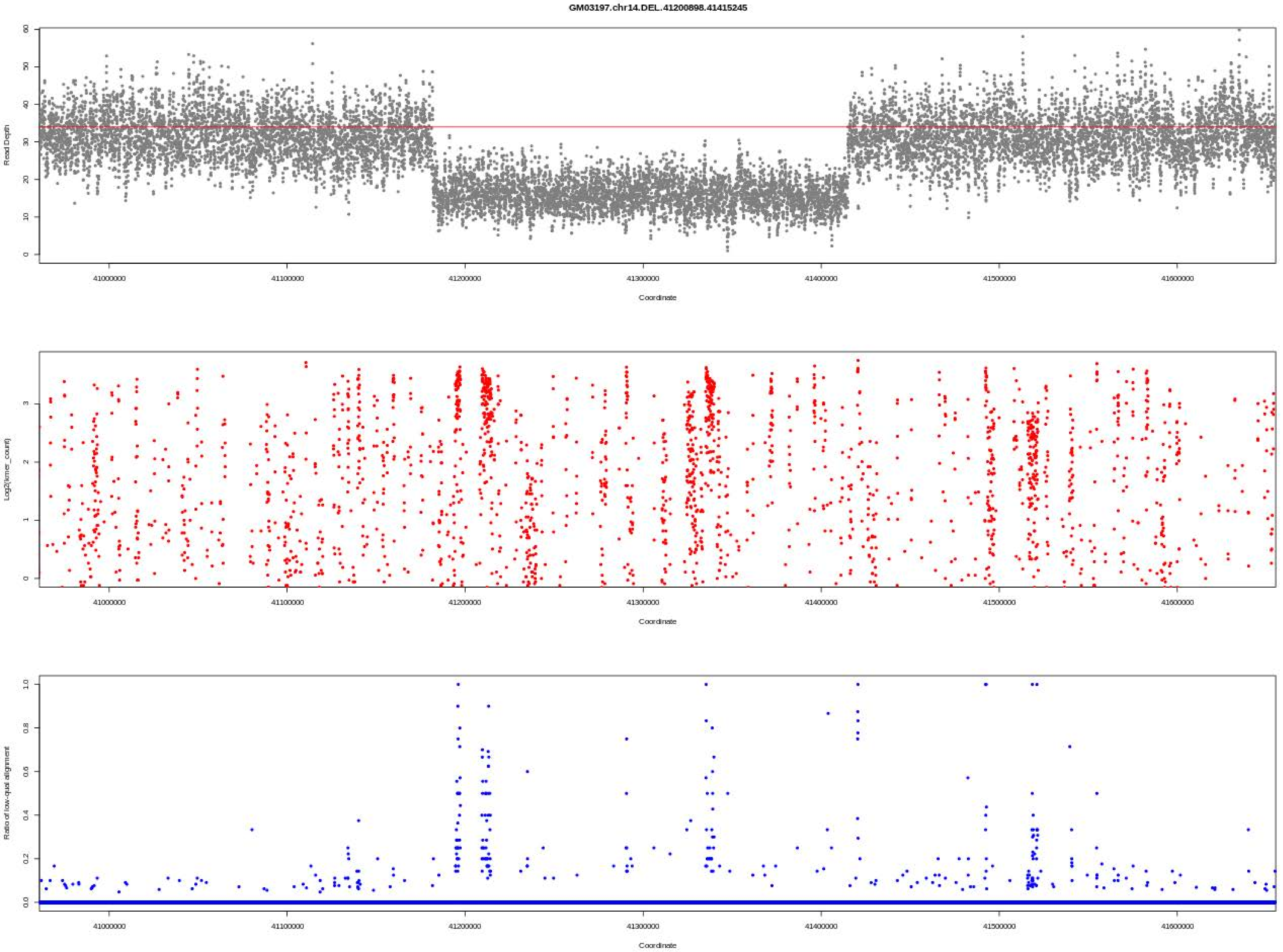
GM03197.chr14.DEL.41200898.41415245
Read Depth
Coordinate
Log2(kmer_count)
Coordinate
Ratio of low-qual alignment
Coordinate

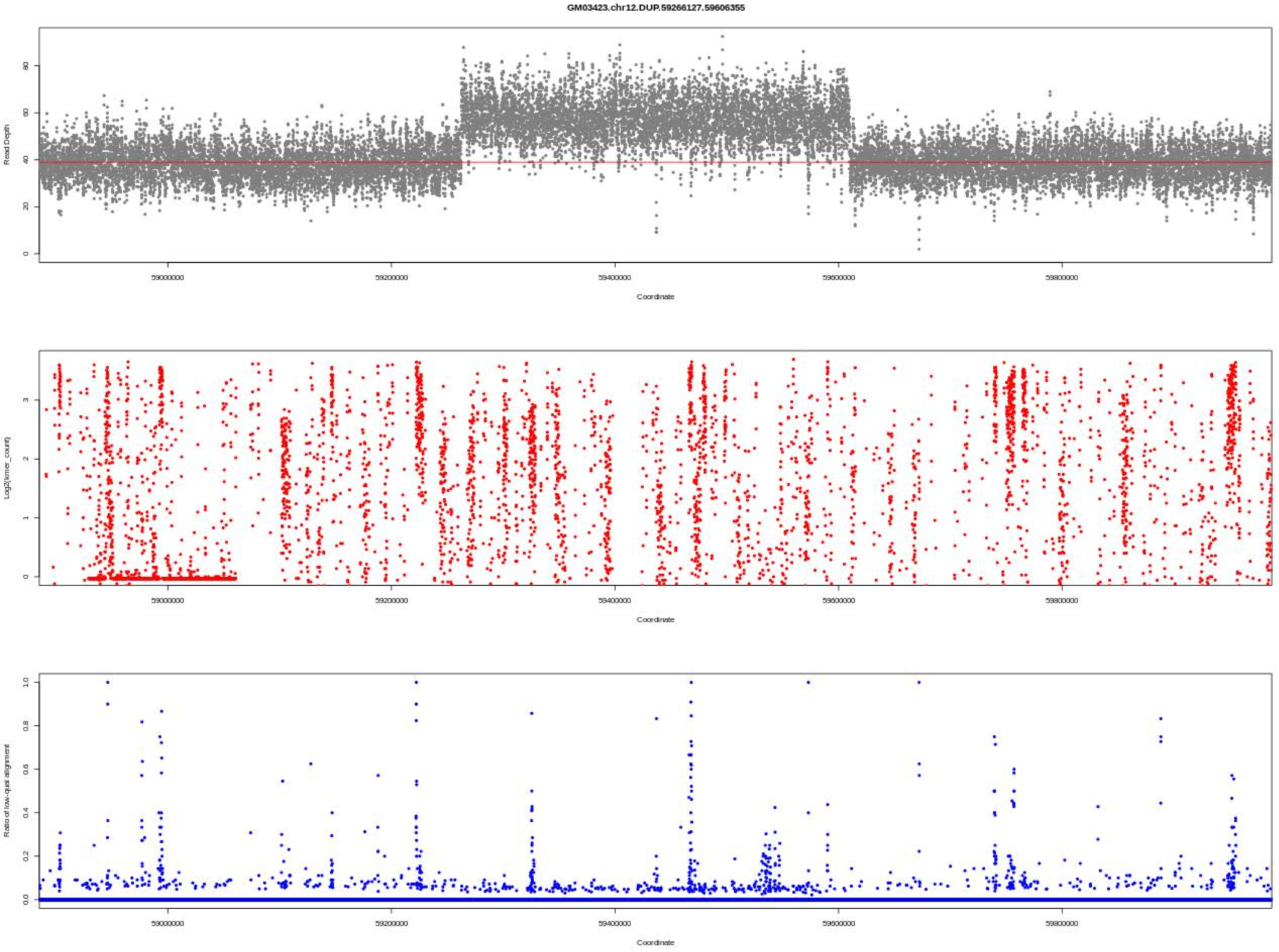
GM03423.chr12.DUP.59266127.59606355
Read Depth
0
20
40
60
80
59000000
59200000
59400000
59600000
59800000
Coordinate
Log2(kmer_count)
0
1
2
3
Coordinate
Ratio of low-qual alignment
0.0
0.2
0.4
0.6
0.8
1.0
Coordinate

GM03997.chr5.DUP.170295343.170425959

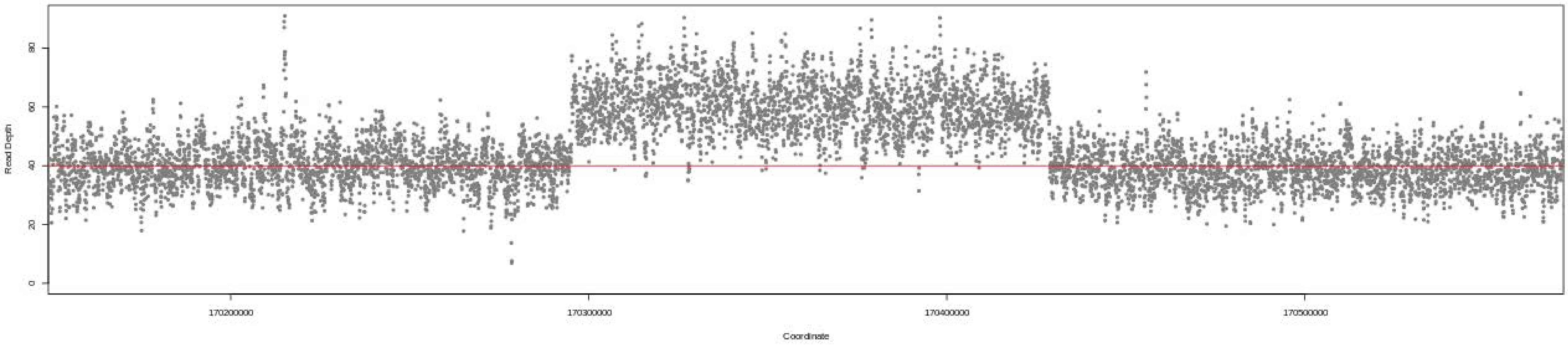

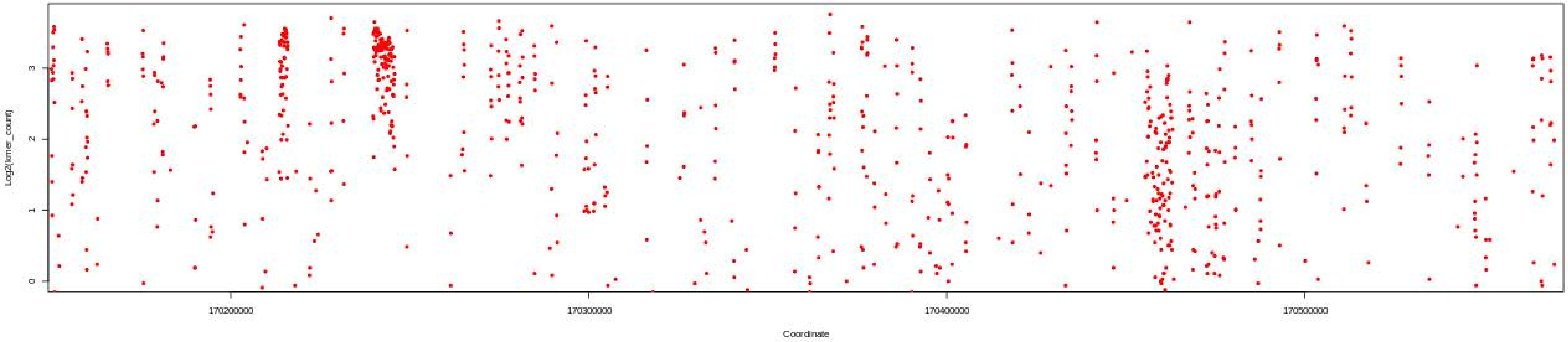

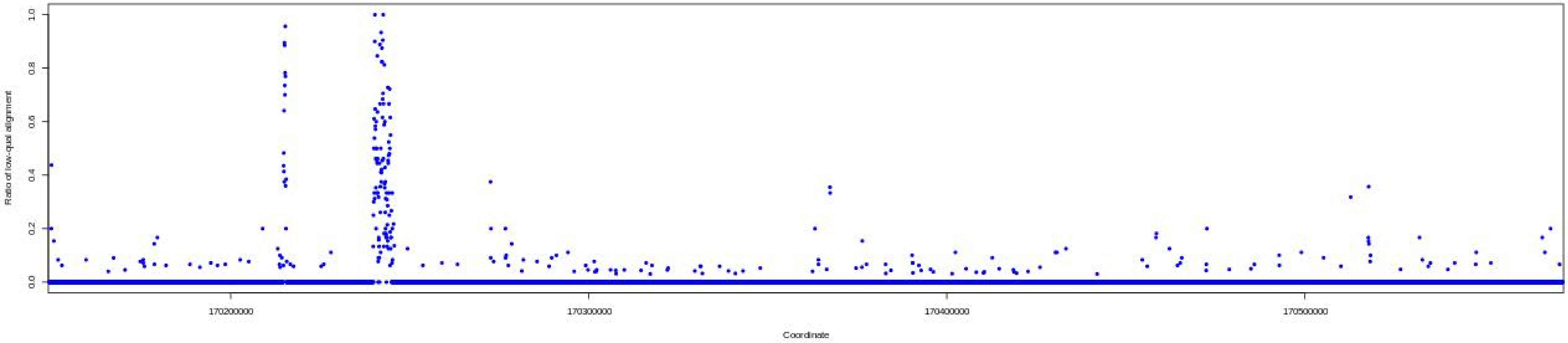

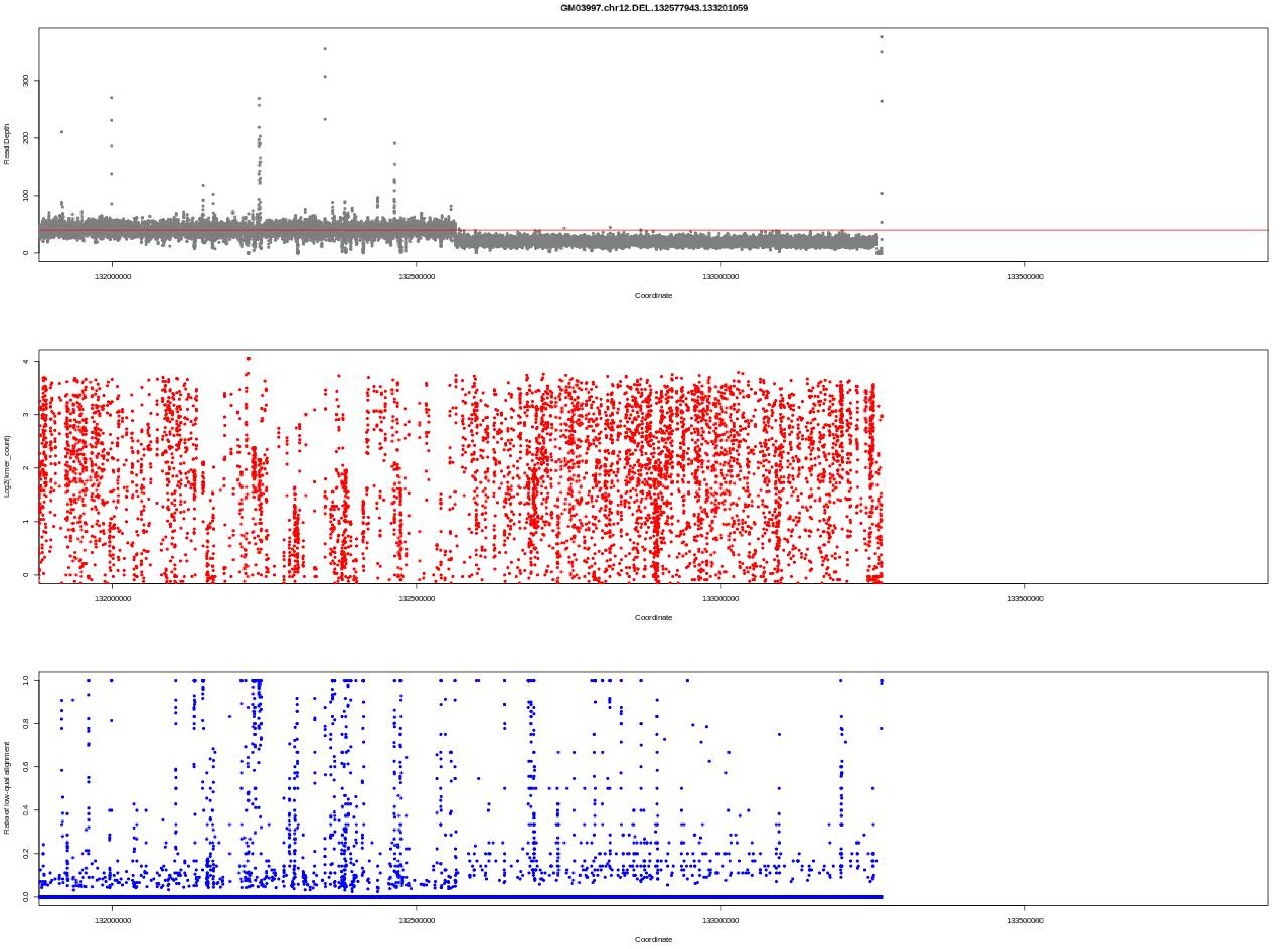
GM03997.chr12.DEL.132577943.133201059
Read Depth
Log2(kmer_count)
Ratio of low-qual alignment
Coordinate
132000000
132500000
133000000
133500000

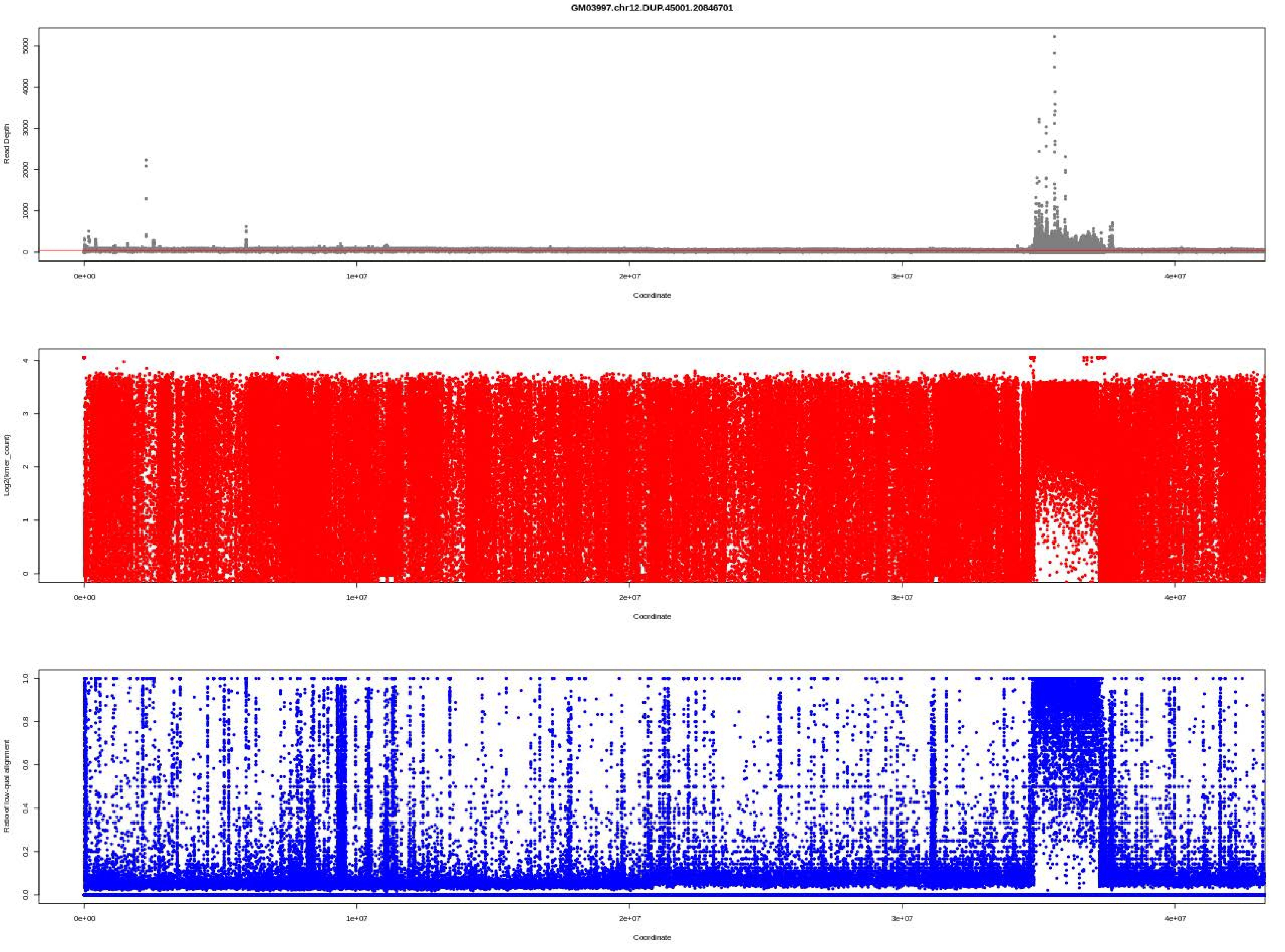
GM03997.chr12.DUP.45001.20846701
Read Depth
Coordinate
Log2(kmer_count)
Coordinate
Ratio of low-qual alignment
Coordinate

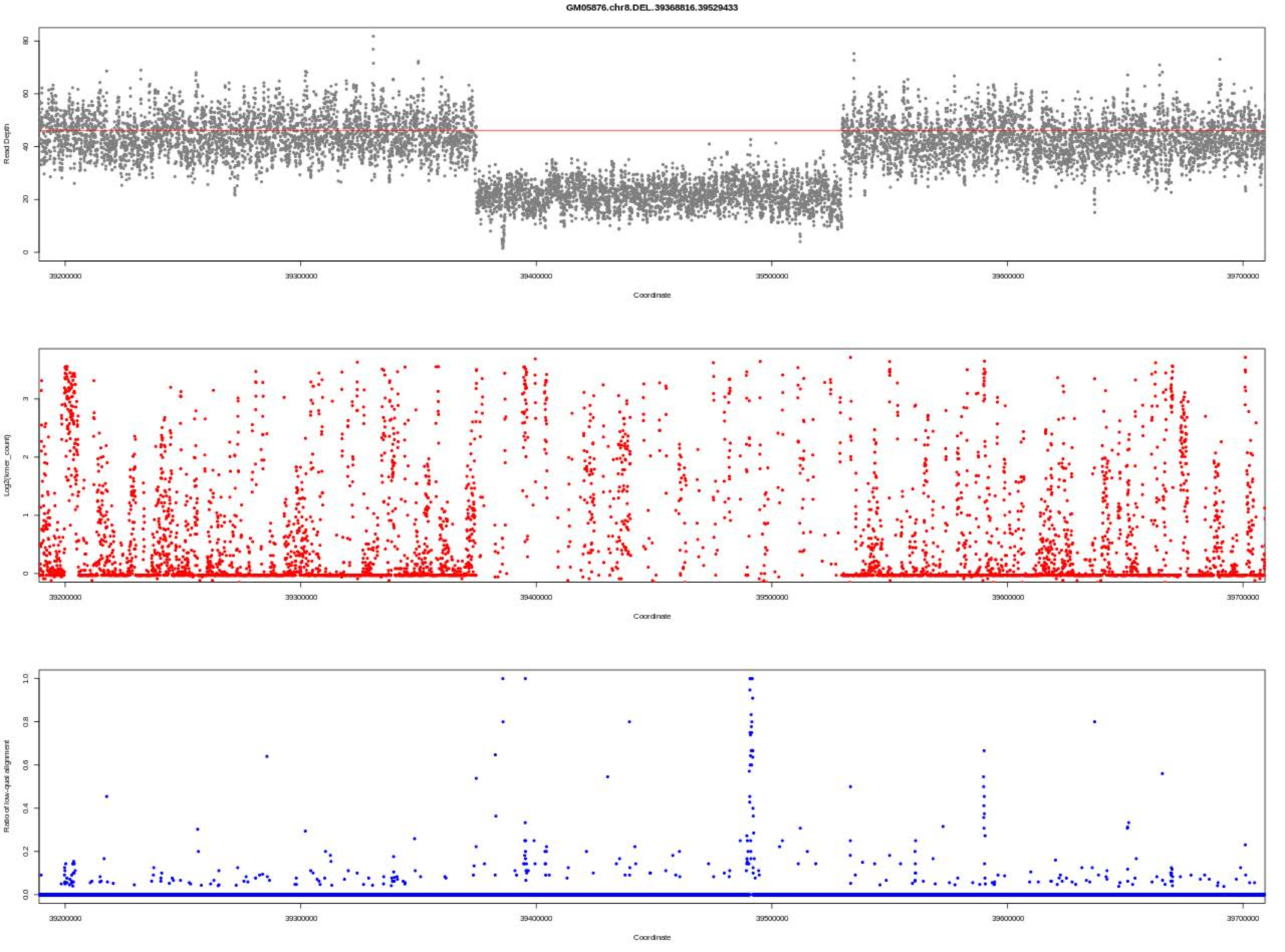

GM05876.chr8.DEL.39368816.39529433
Read Depth
Coordinate
Log2(kmer_count)
Coordinate
Ratio of low-qual alignment
Coordinate

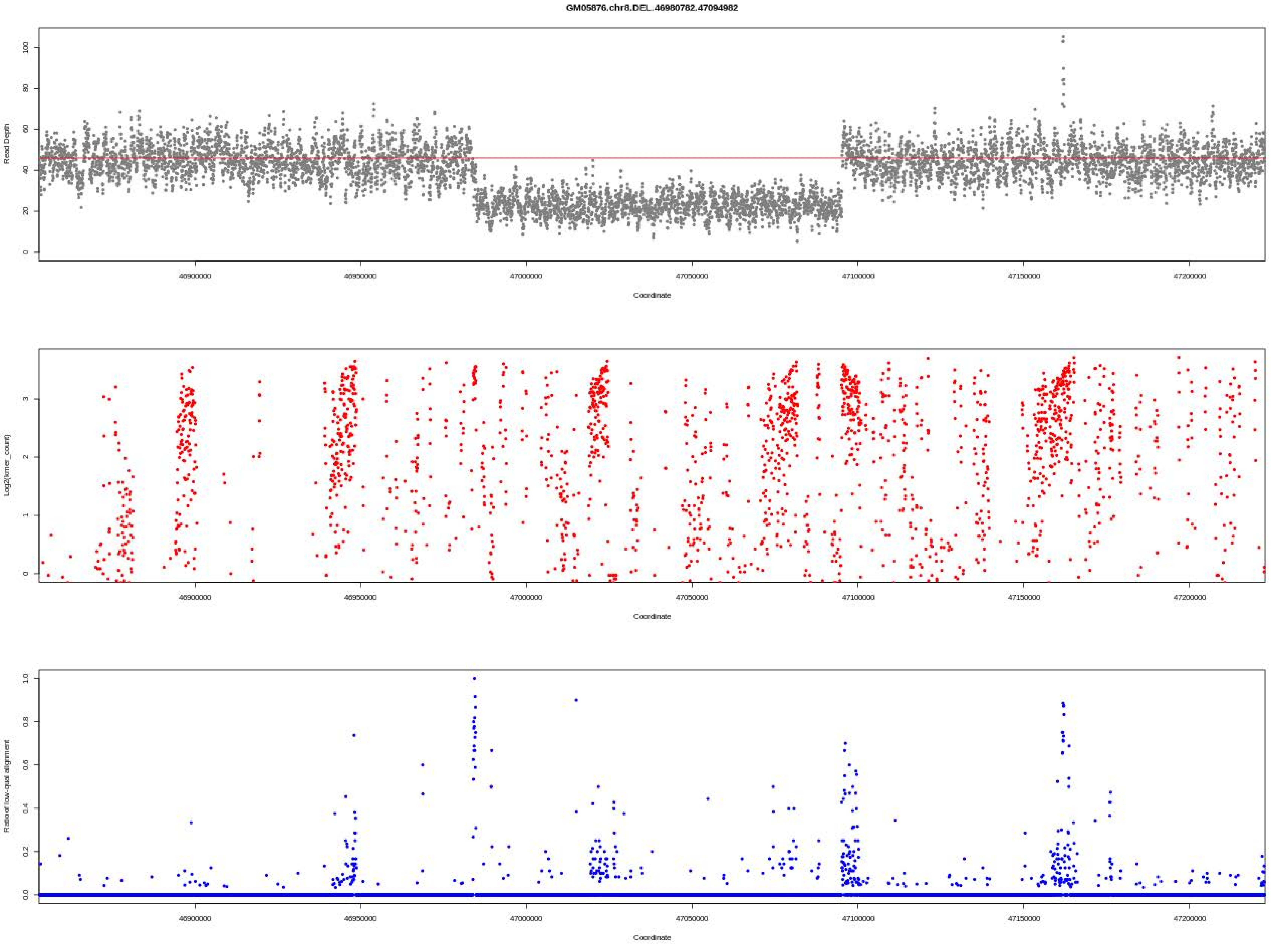

GM05876.chr8.DEL.46980782.47094982
Read Depth
Coordinate
Log2(kmer_count)
Coordinate
Ratio of low-qual alignment
Coordinate

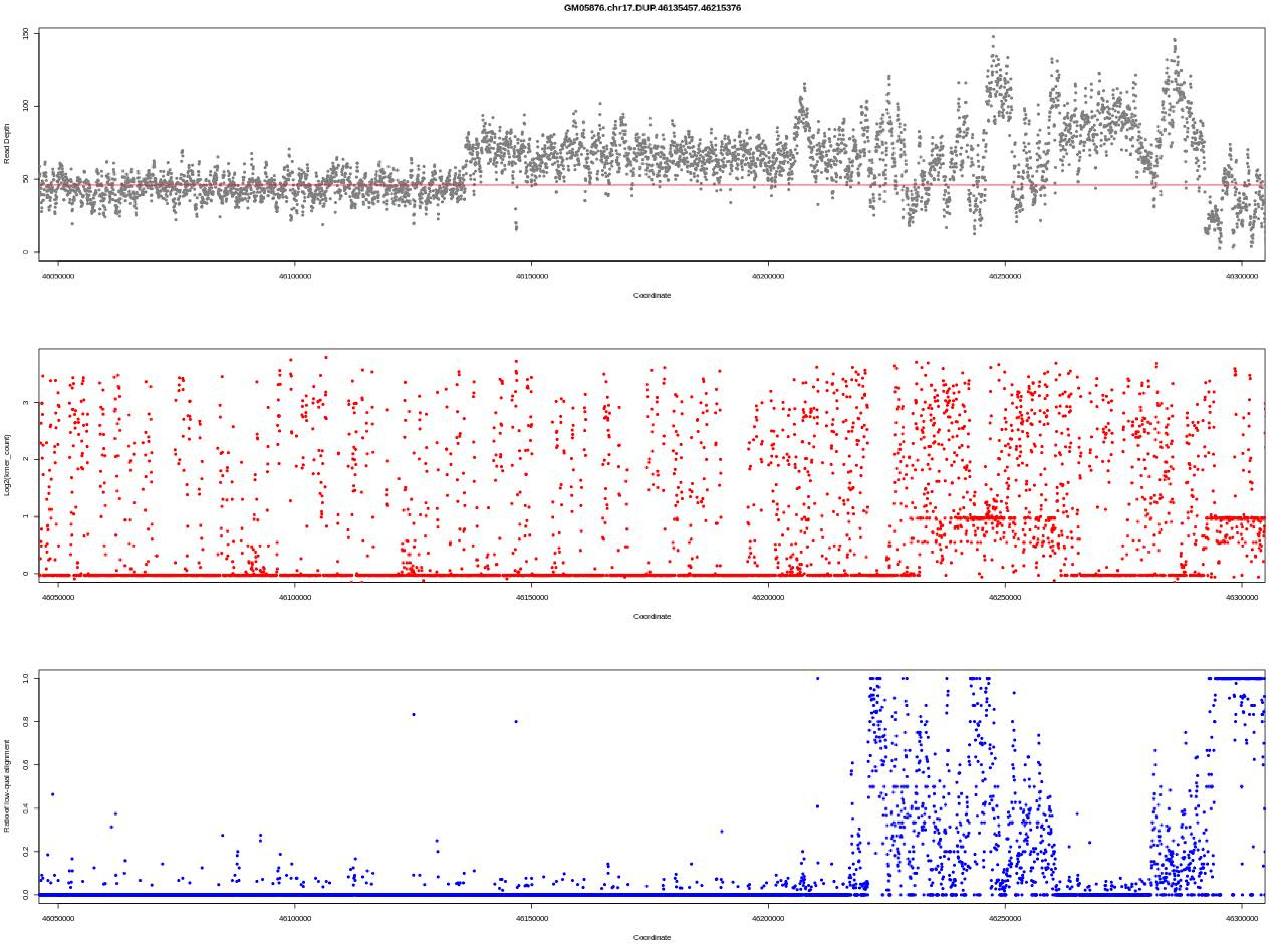
GM05876.chr17.DUP.46135457.46215376
Read Depth
Log2(kmer_count)
Ratio of low-qual alignment
Coordinate
46050000
46100000
46150000
46200000
46250000
46300000

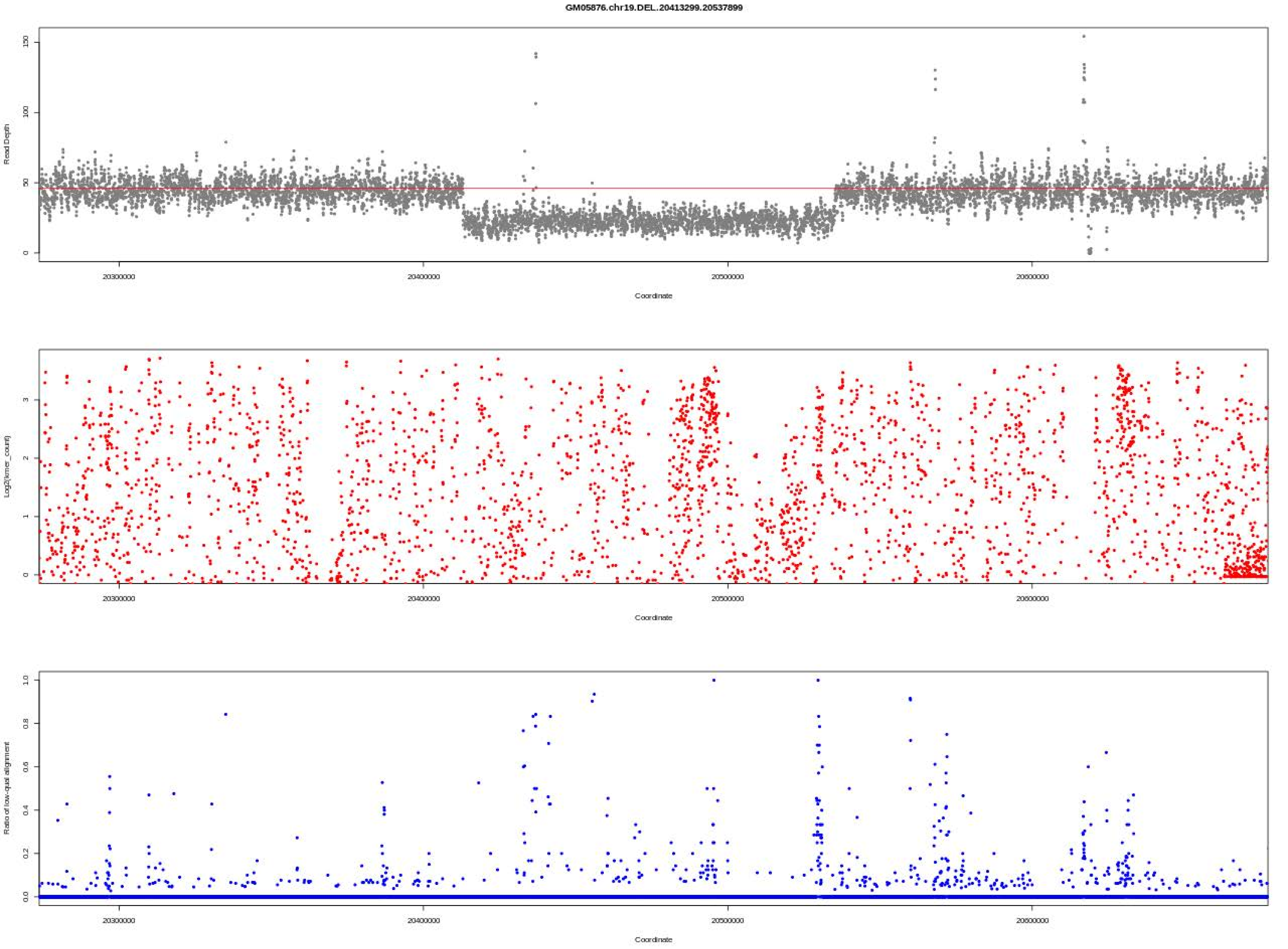

GM05876.chr19.DEL..20413299.20537899
Read Depth
0
50
100
150
20300000
20400000
20500000
20600000
Coordinate
Log2(kmer_count)
0
1
2
3
Ratio of low-qual alignment
0.0
0.2
0.4
0.6
0.8
1.0

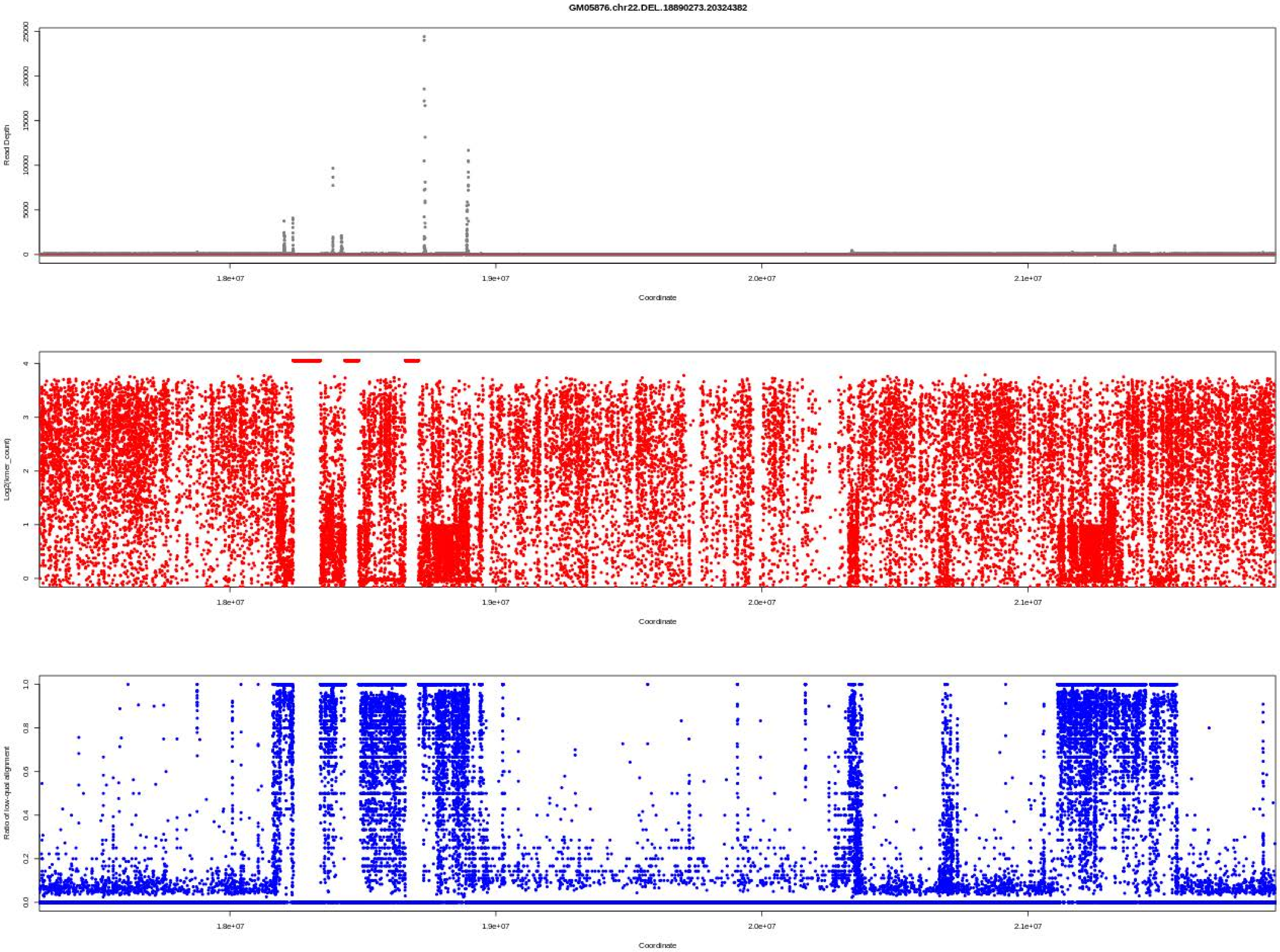

GM05876.chr22.DEL.18890273.20324382
Read Depth
Coordinate
Log2(kmer_count)
Coordinate
Ratio of low-qual alignment
Coordinate
1.8e+07
1.9e+07
2.0e+07
2.1e+07

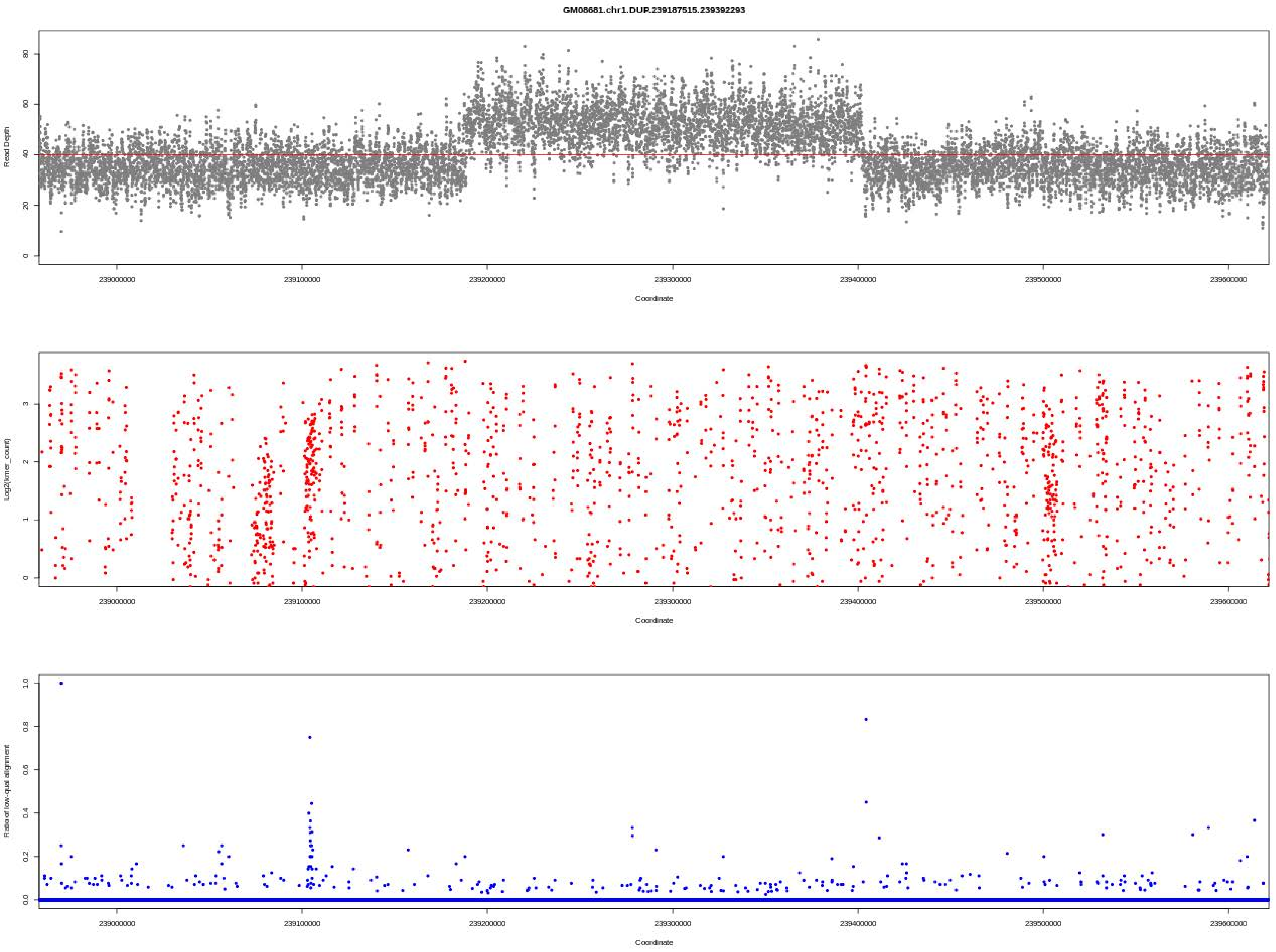
GM08681.chr1.DUP.239187515.239392293
Read Depth
Coordinate
Log2(kmer_count)
Coordinate
Ratio of low-qual alignment
Coordinate
239000000
239100000
239200000
239300000
239400000
239500000
239600000

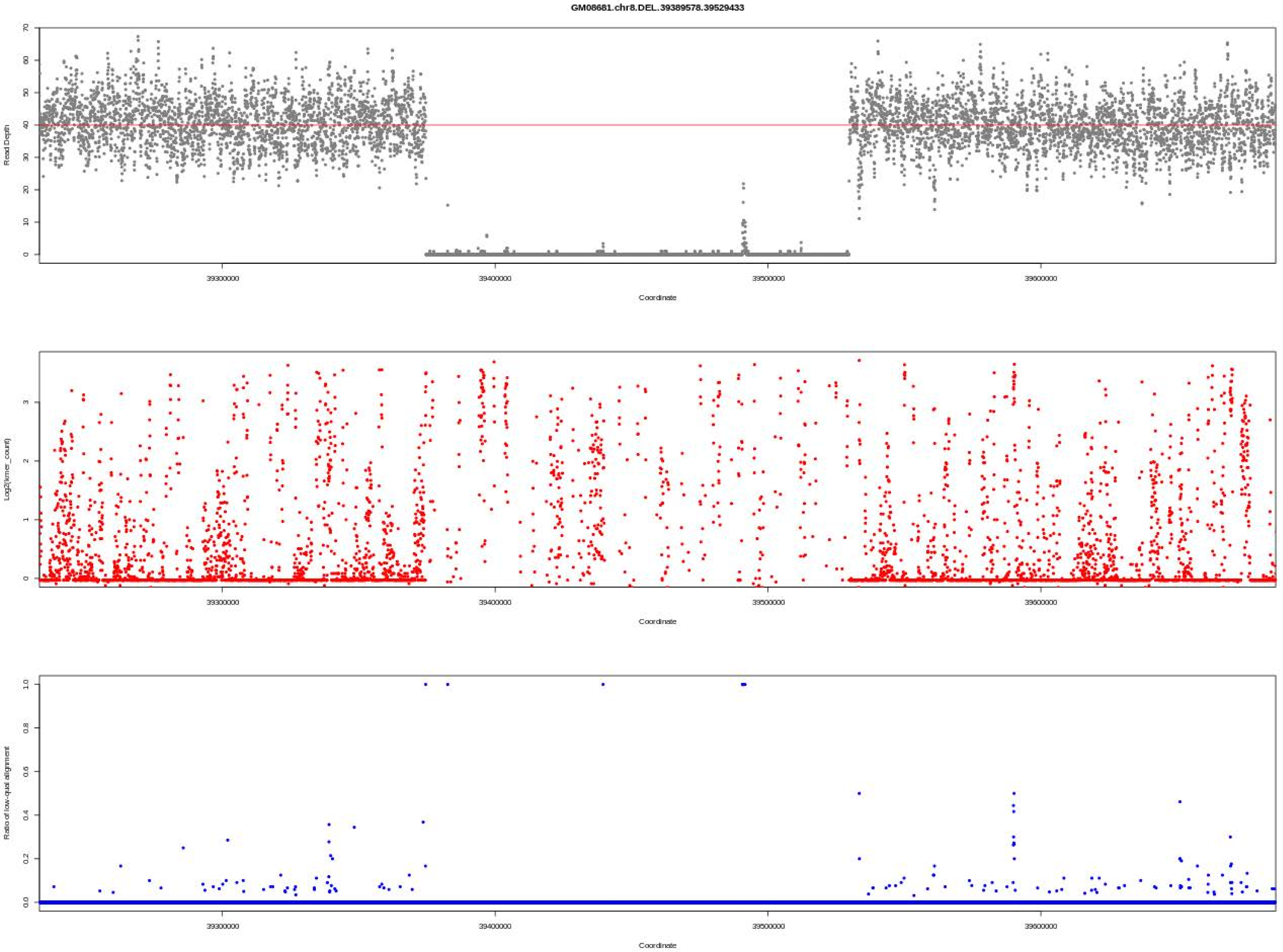
GM08681.chr8.DEL.39389578.39529433
Read Depth
Coordinate
Log2(kmer_count)
Coordinate
Ratio of low-qual alignment
Coordinate
39300000
39400000
39500000
39600000

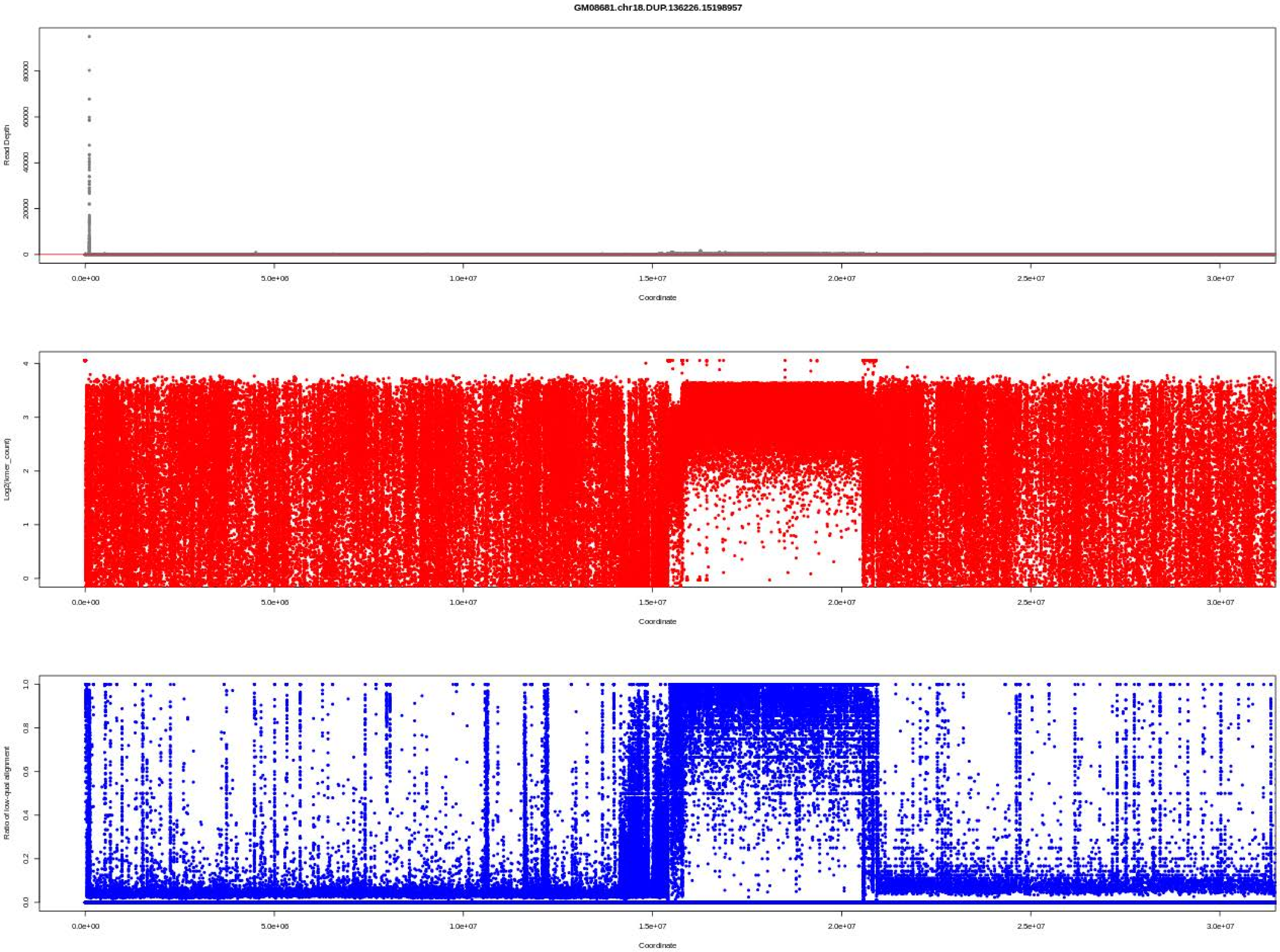

GM08681.chr18.DUP.136226.15198957
Read Depth
Log2(kmer_count)
Ratio of low-qual alignment
Coordinate
0.0e+00
5.0e+06
1.0e+07
1.5e+07
2.0e+07
2.5e+07
3.0e+07

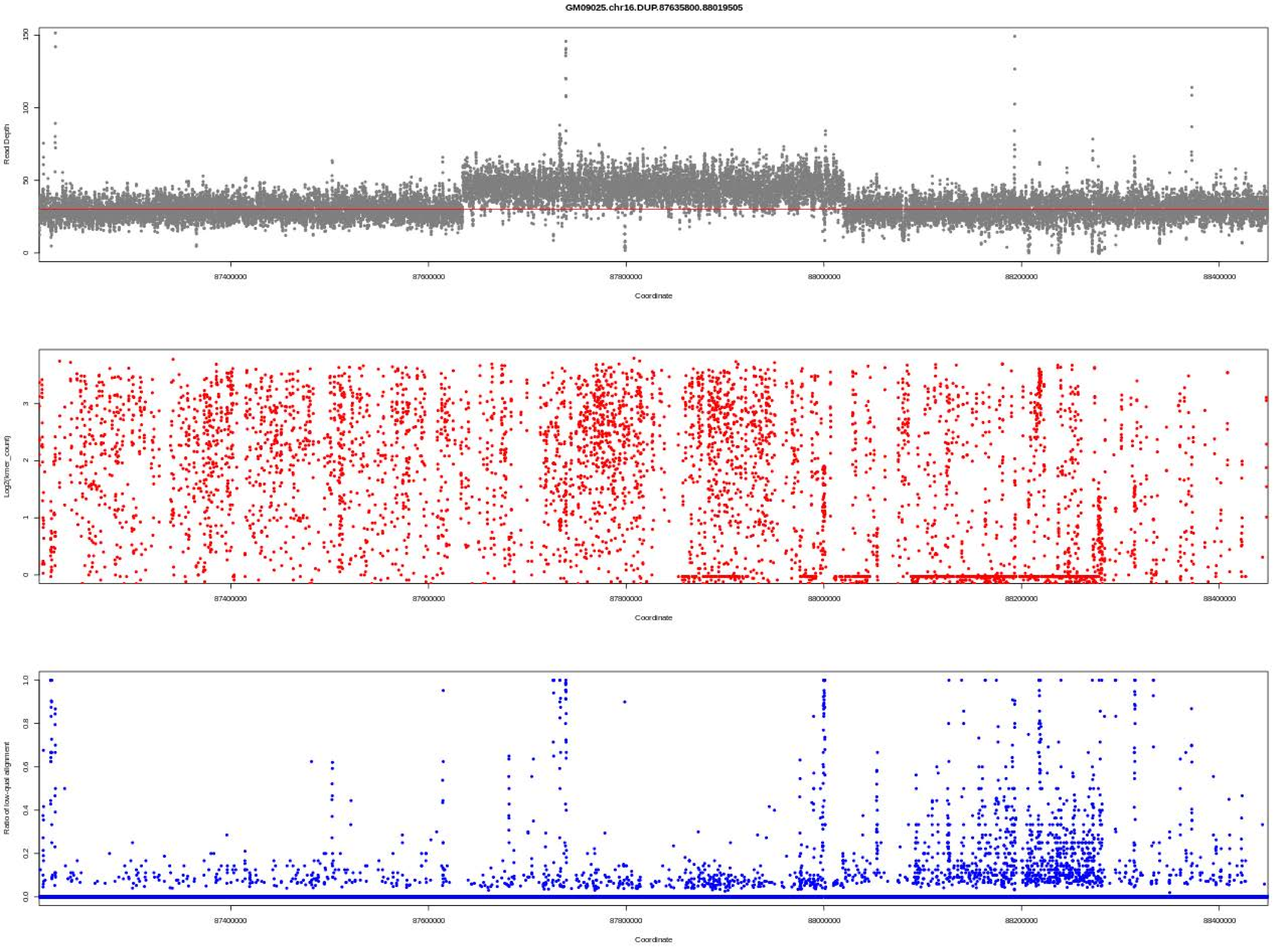

GM09025.chr16.DUP.87635800.88019505
Read Depth
Coordinate
Log2(kmer_count)
Coordinate
Ratio of low-qual alignment
Coordinate
87400000
87600000
87800000
88000000
88200000
88400000

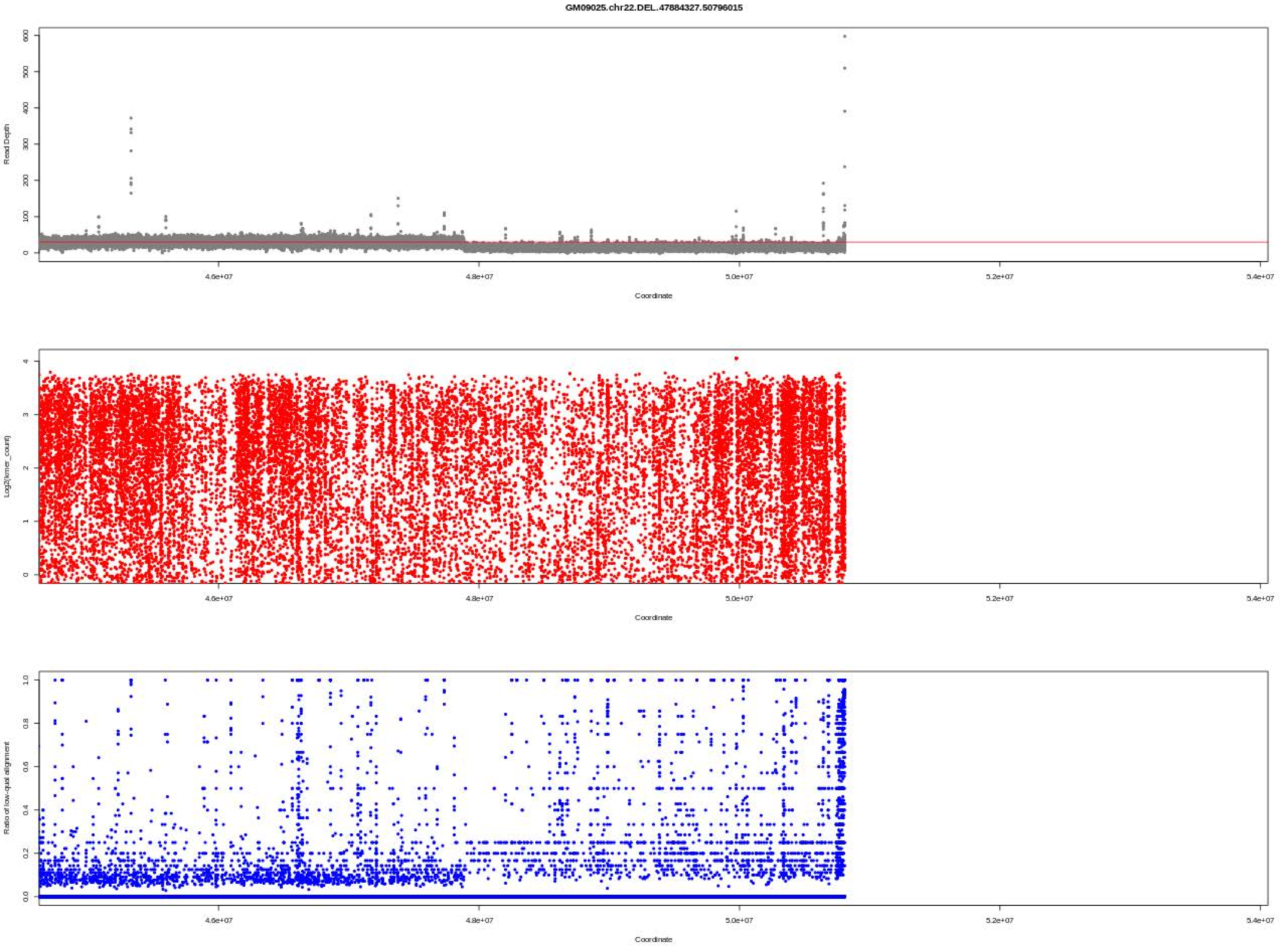

GM09025.chr22.DEL.47884327.50796015
Read Depth
Coordinate
Log2(kmer_count)
Coordinate
Ratio of low-qual alignment
Coordinate

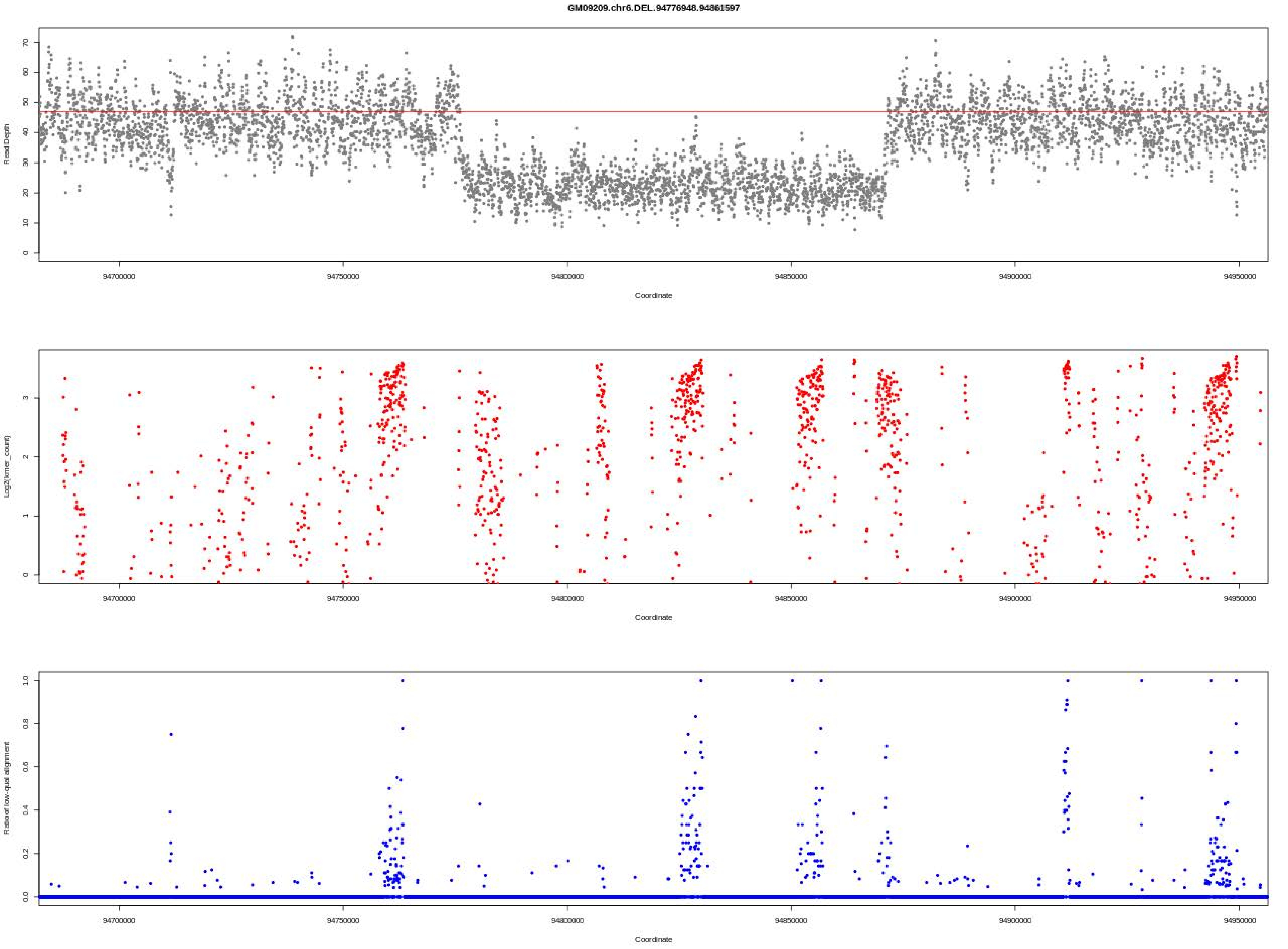
GM09209.chr6.DEL.94776948.94861597
Read Depth
Coordinate
Log2(kmer_count)
Coordinate
Ratio of low-qual alignment
Coordinate

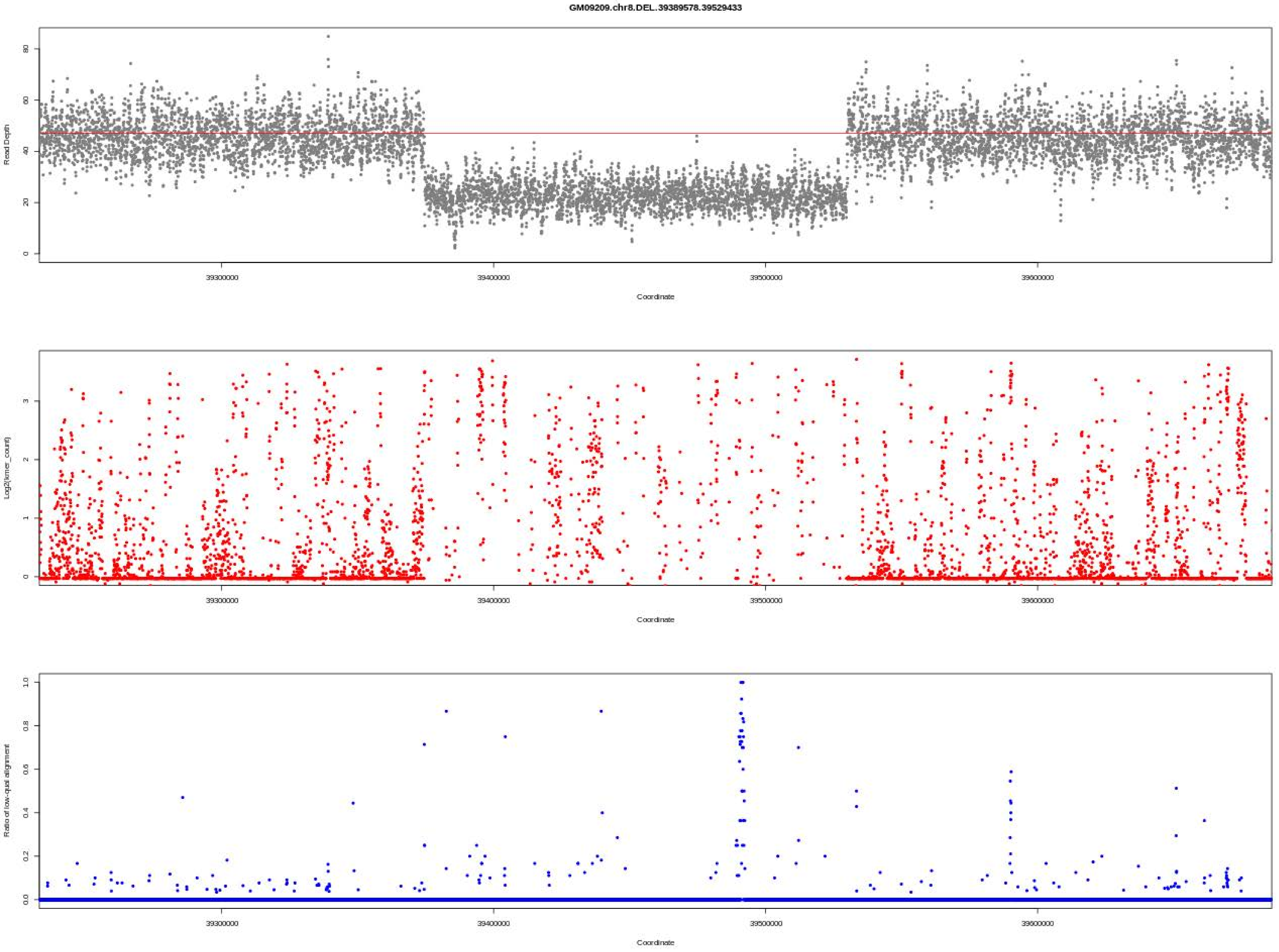
GM09209.chr8.DEL.39389578.39529433
Read Depth
Coordinate
Log2(kmer_count)
Coordinate
Ratio of low-qual alignment
Coordinate

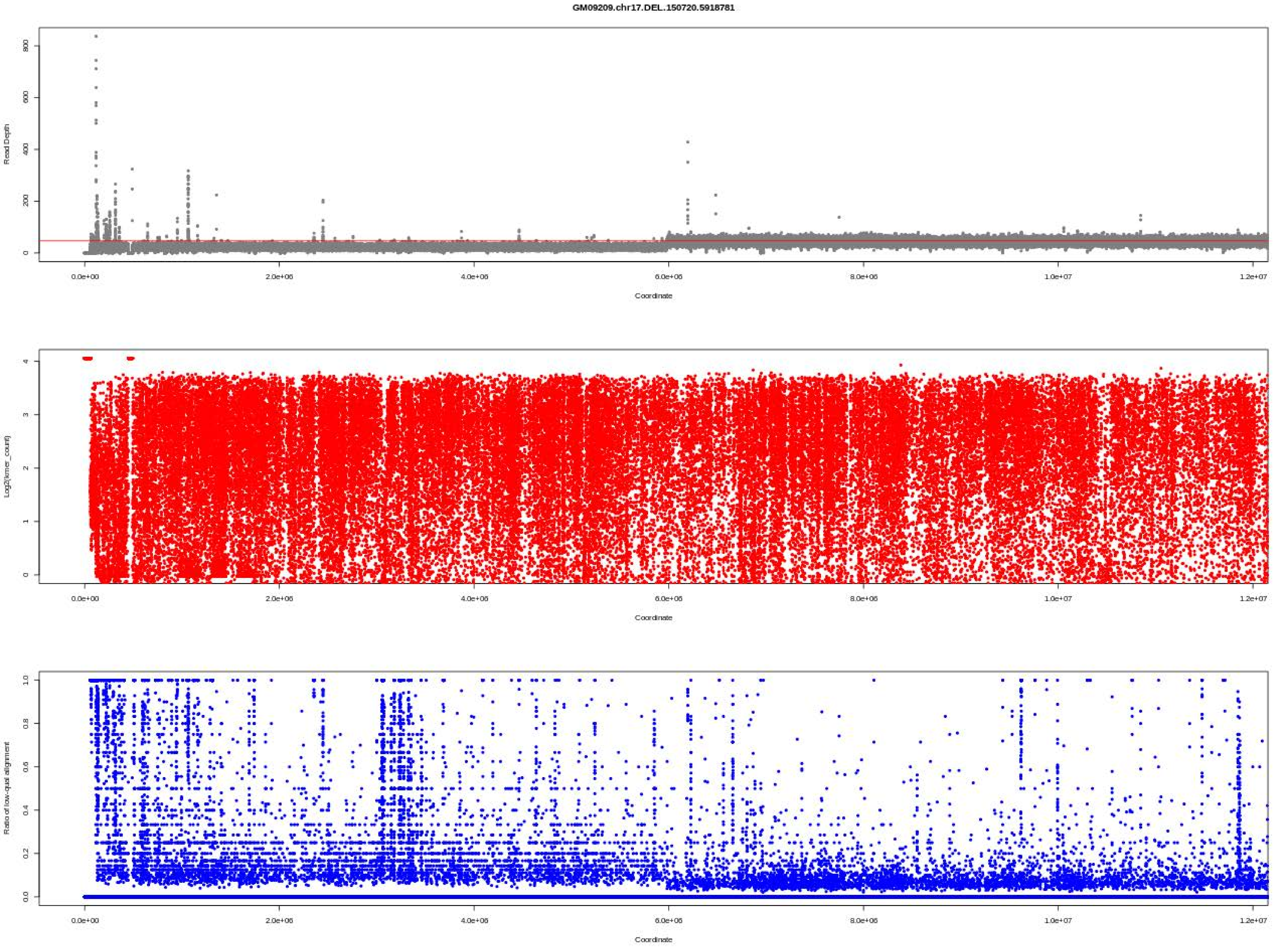

GM09209.chr17.DEL.150720.5918781
Read Depth
Coordinate
Log2(kmer_count)
Coordinate
Ratio of low-qual alignment
Coordinate

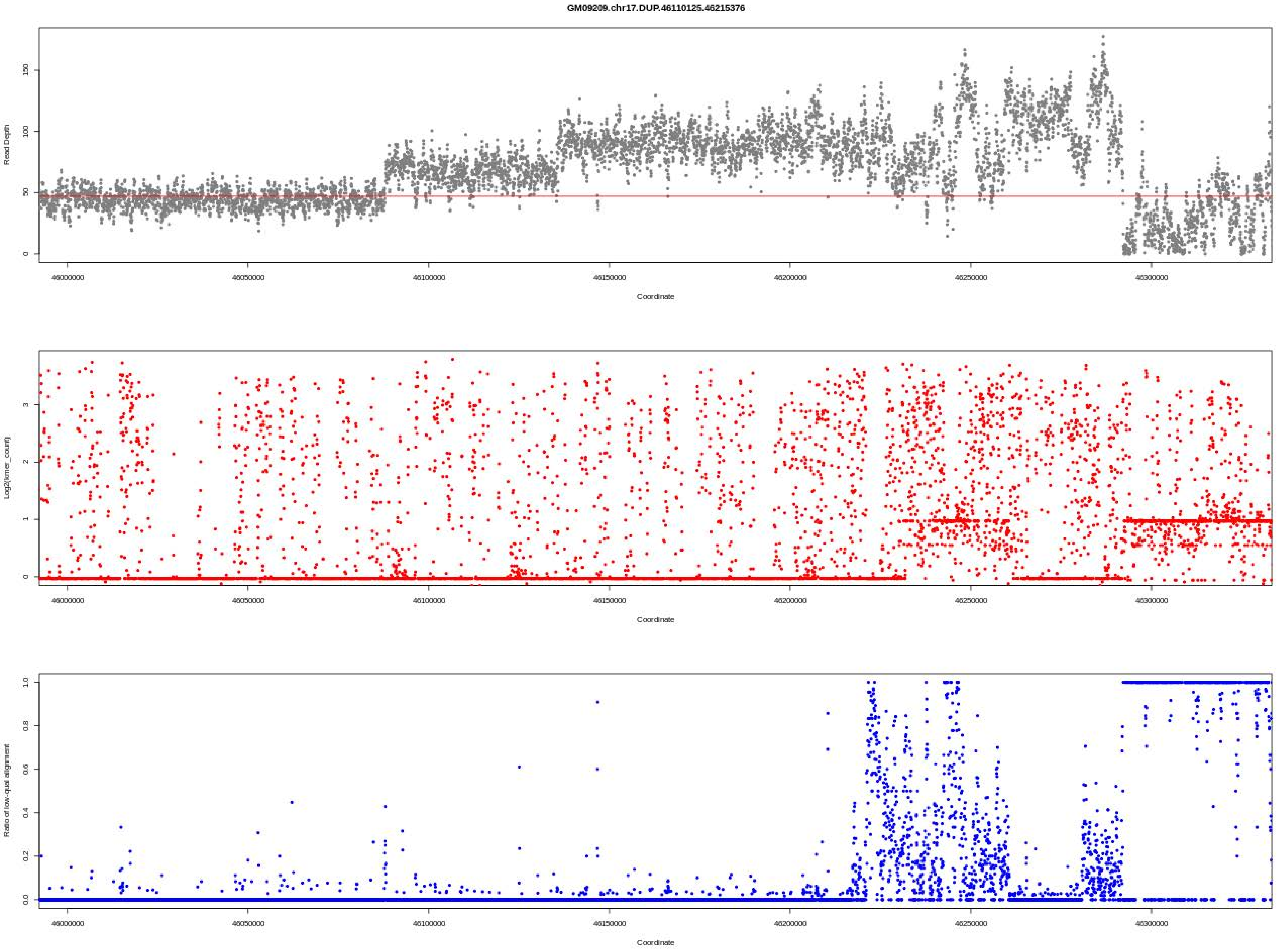
GM09209.chr17.DUP.46110125.46215376
Read Depth
Coordinate
Log2(kmer_count)
Coordinate
Ratio of low-qual alignment
Coordinate

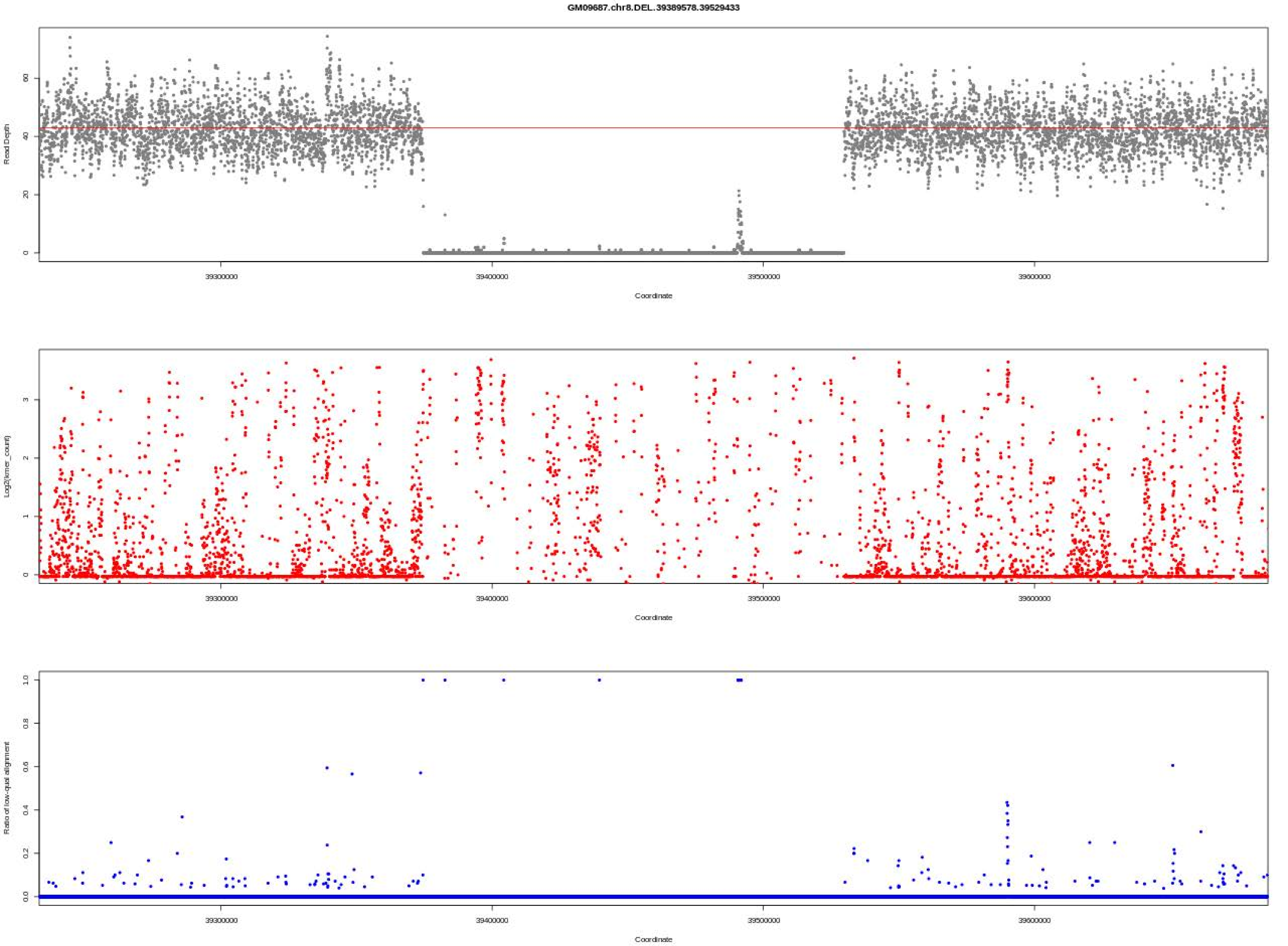

GM09687.chr8.DEL.39389578.39529433
Read Depth
Coordinate
Log2(kmer_count)
Coordinate
Ratio of low-qual alignment
Coordinate
39300000
39400000
39500000
39600000

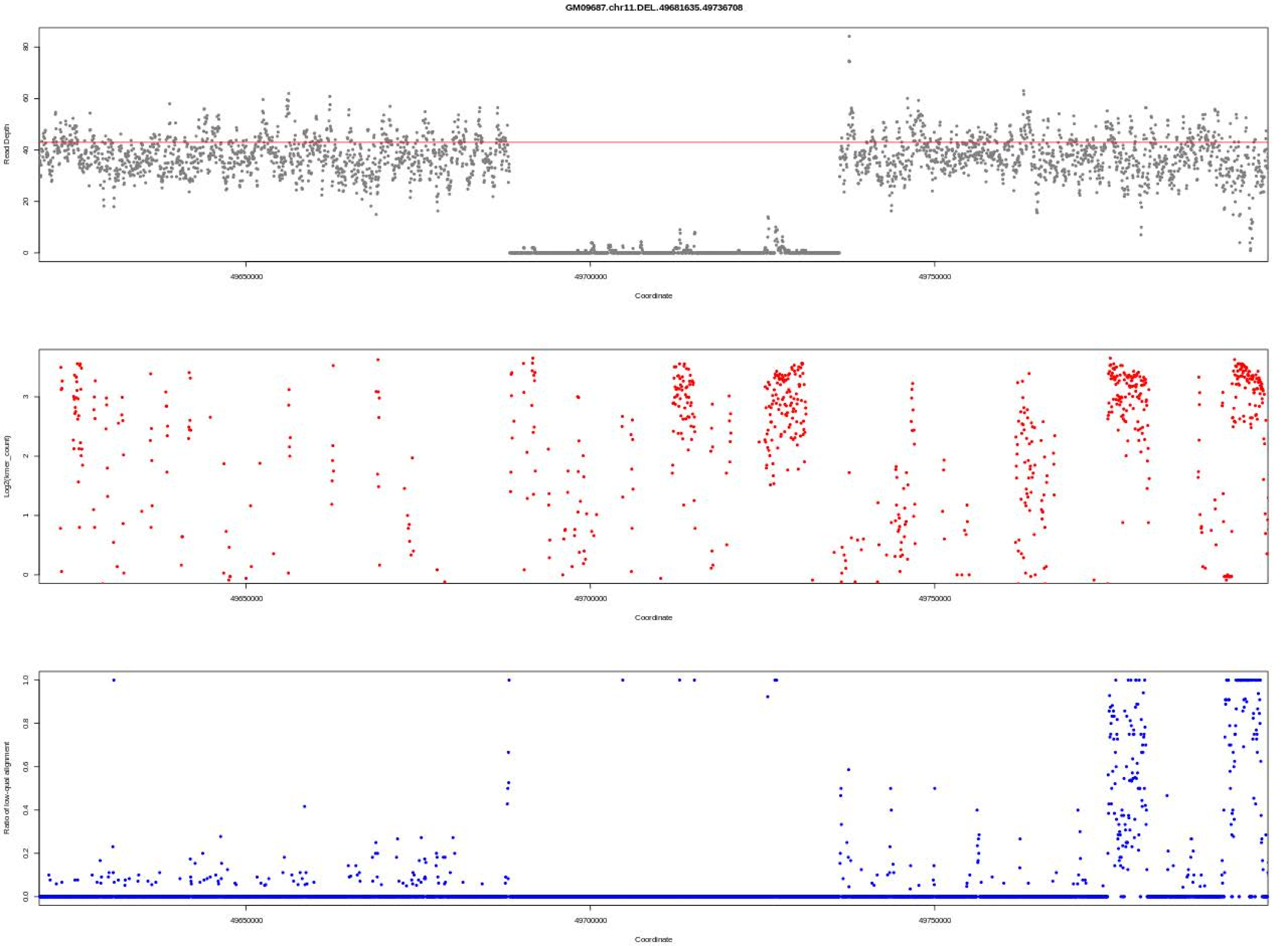

GM09687.chr11.DEL.49681635.49736708
Read Depth
Coordinate
Log2(kmer_count)
Coordinate
Ratio of low-qual alignment
Coordinate
49650000
49700000
49750000

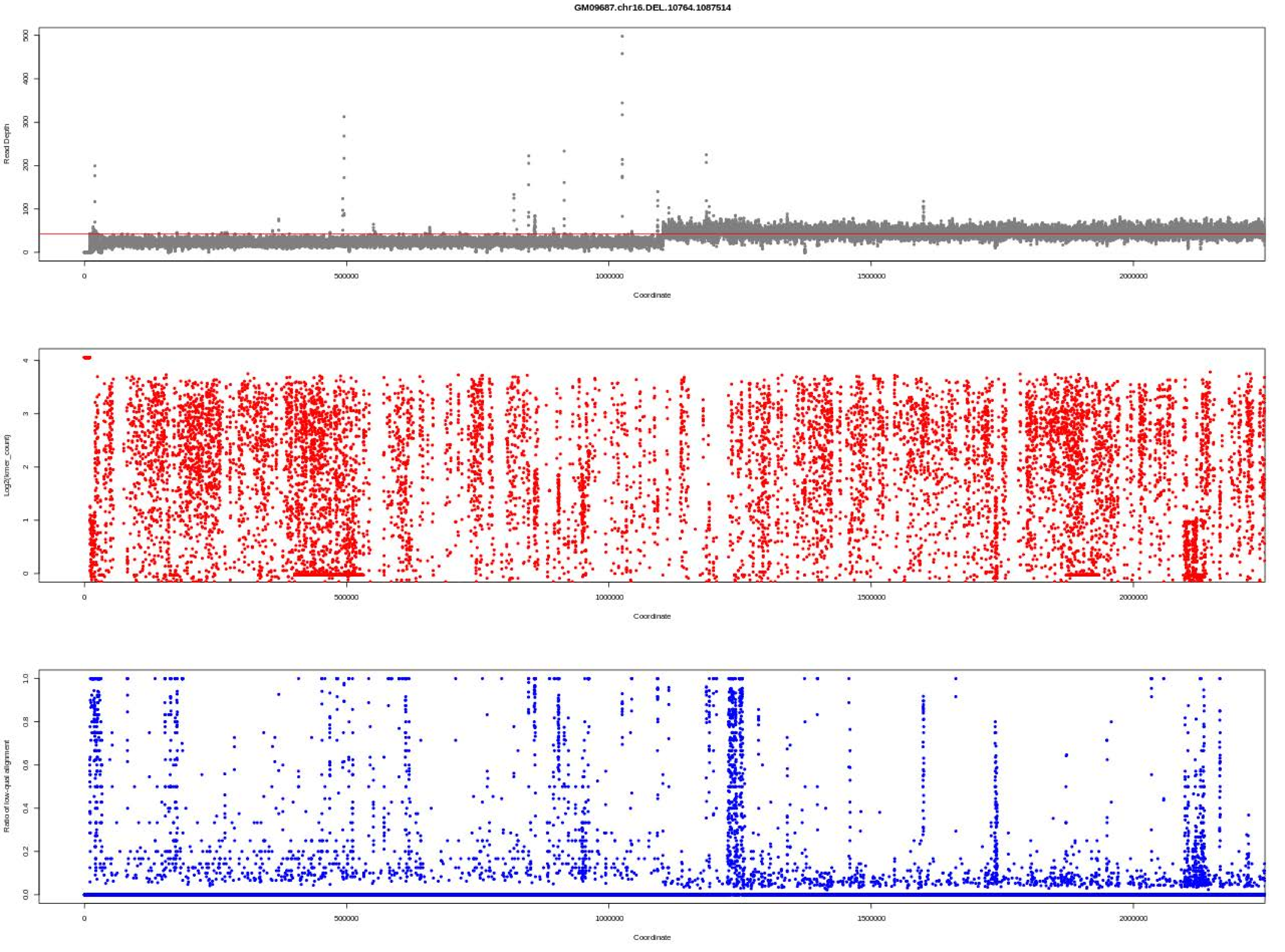

GM09687.chr16.DEL.10764.1087514
Read Depth
Coordinate
Log2(kmer_count)
Coordinate
Ratio of low-qual alignment
Coordinate

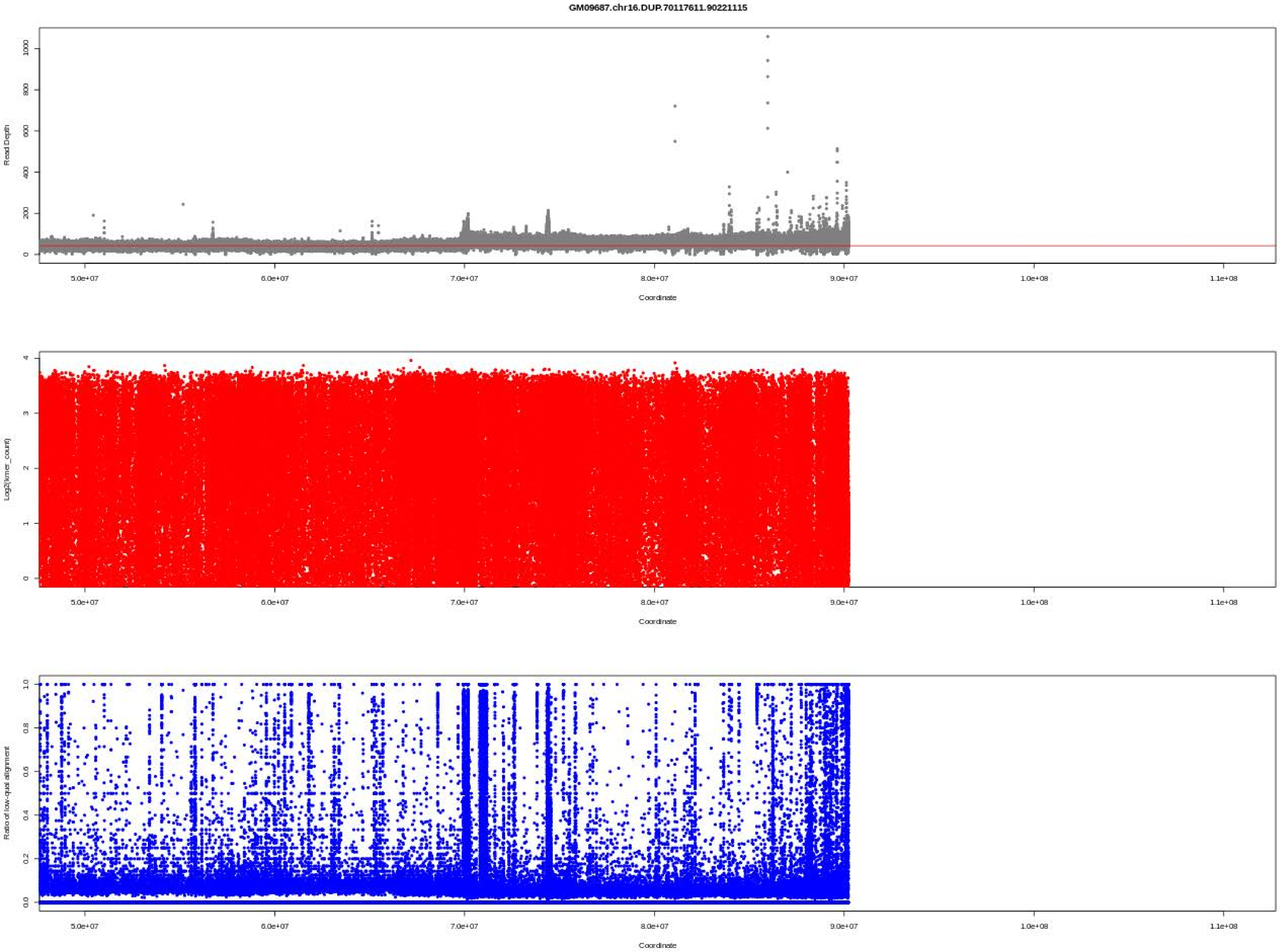
GM09687.chr16.DUP.70117611.90221115
Read Depth
0
200
400
600
800
1000
5.0e+07
6.0e+07
7.0e+07
8.0e+07
9.0e+07
1.0e+08
1.1e+08
Coordinate
Log2(kmer_count)
0
1
2
3
4
Coordinate
Ratio of low-qual alignment
0.0
0.2
0.4
0.6
0.8
1.0
Coordinate

GM09687.chr17.DUP.46110125.46211076

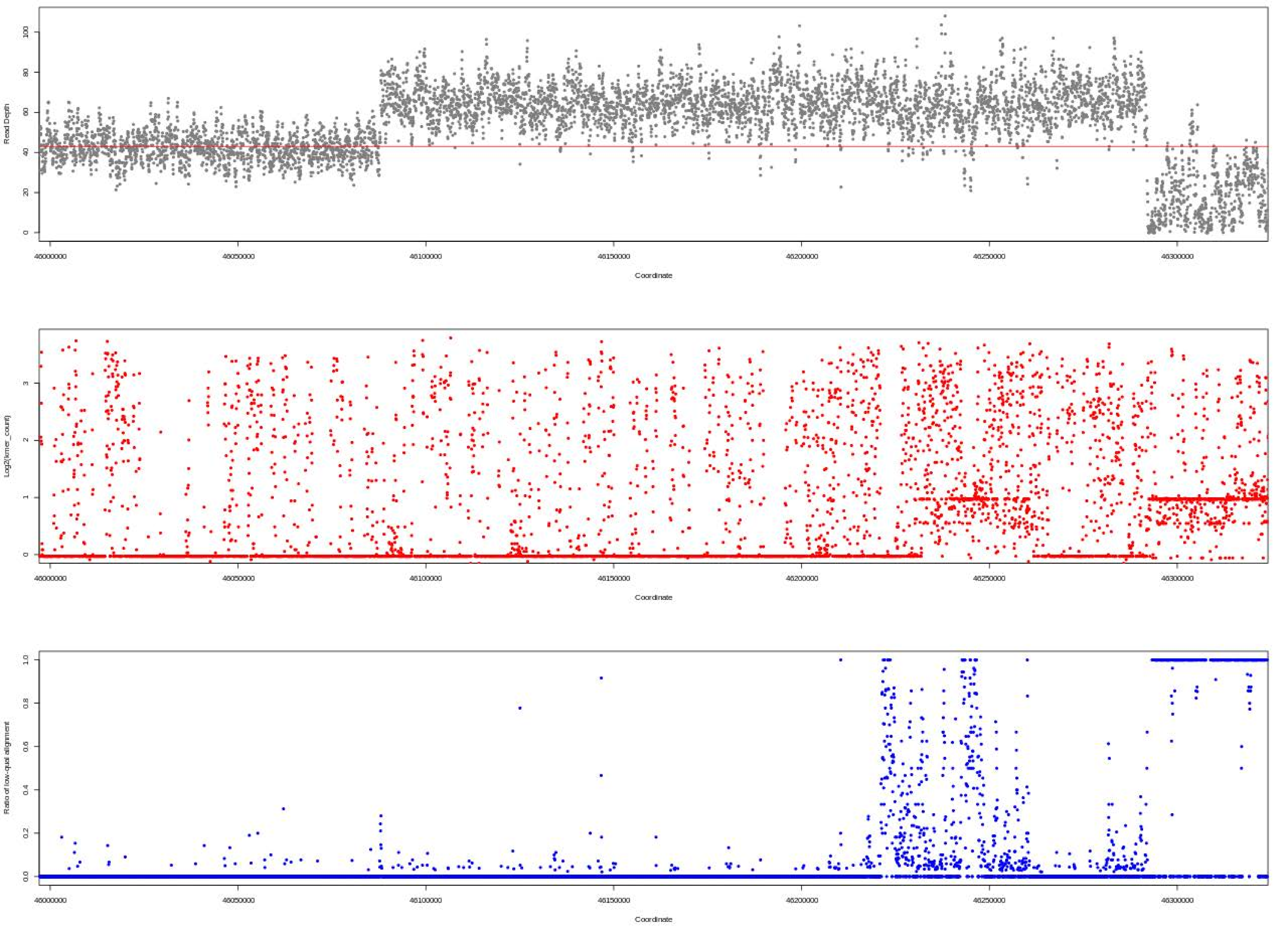

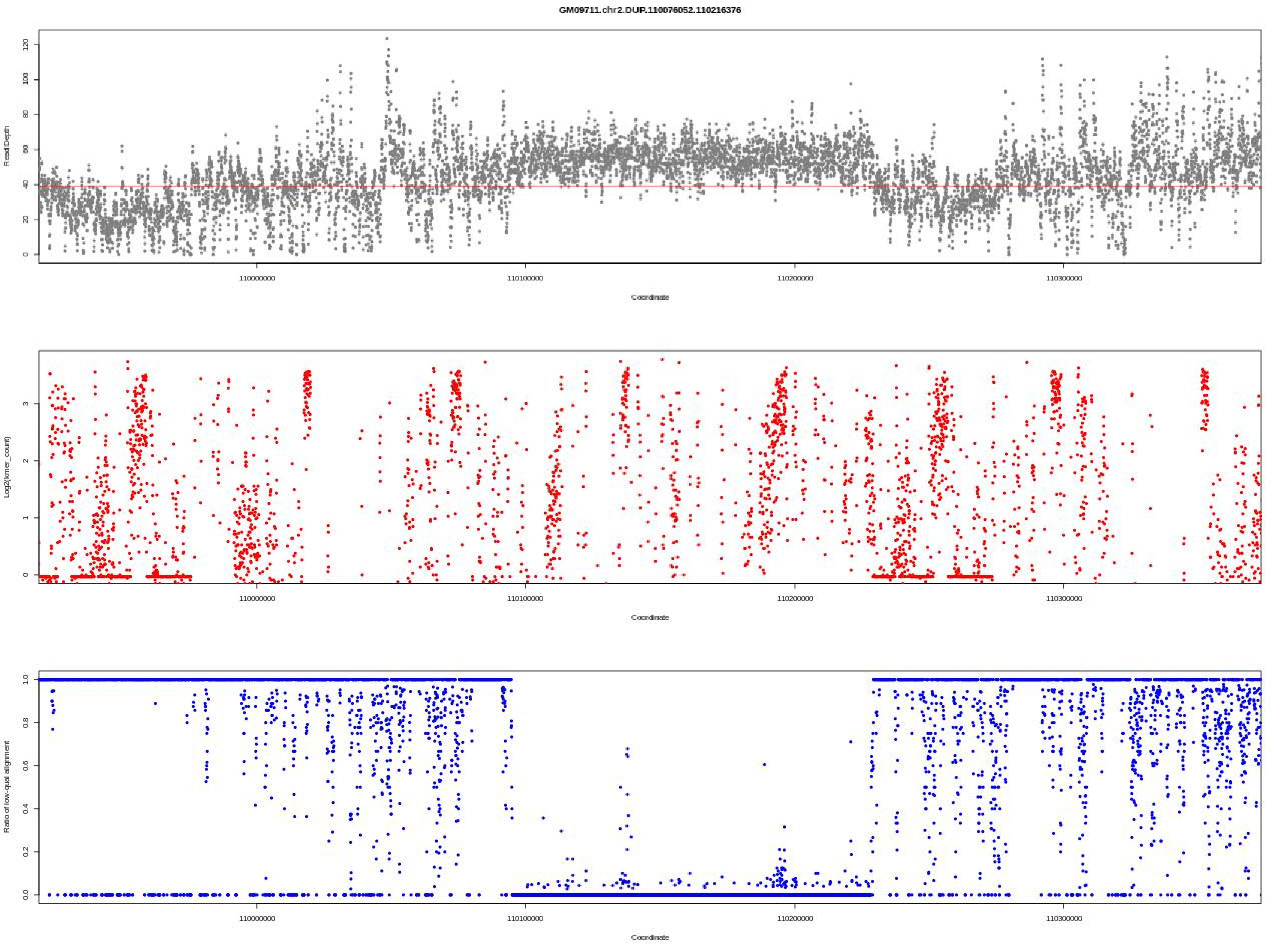
GM09711.chr2.DUP.110076052.110216376
Read Depth
Coordinate
Log2(kmer_count)
Coordinate
Ratio of low-qual alignment
Coordinate
110000000
110100000
110200000
110300000

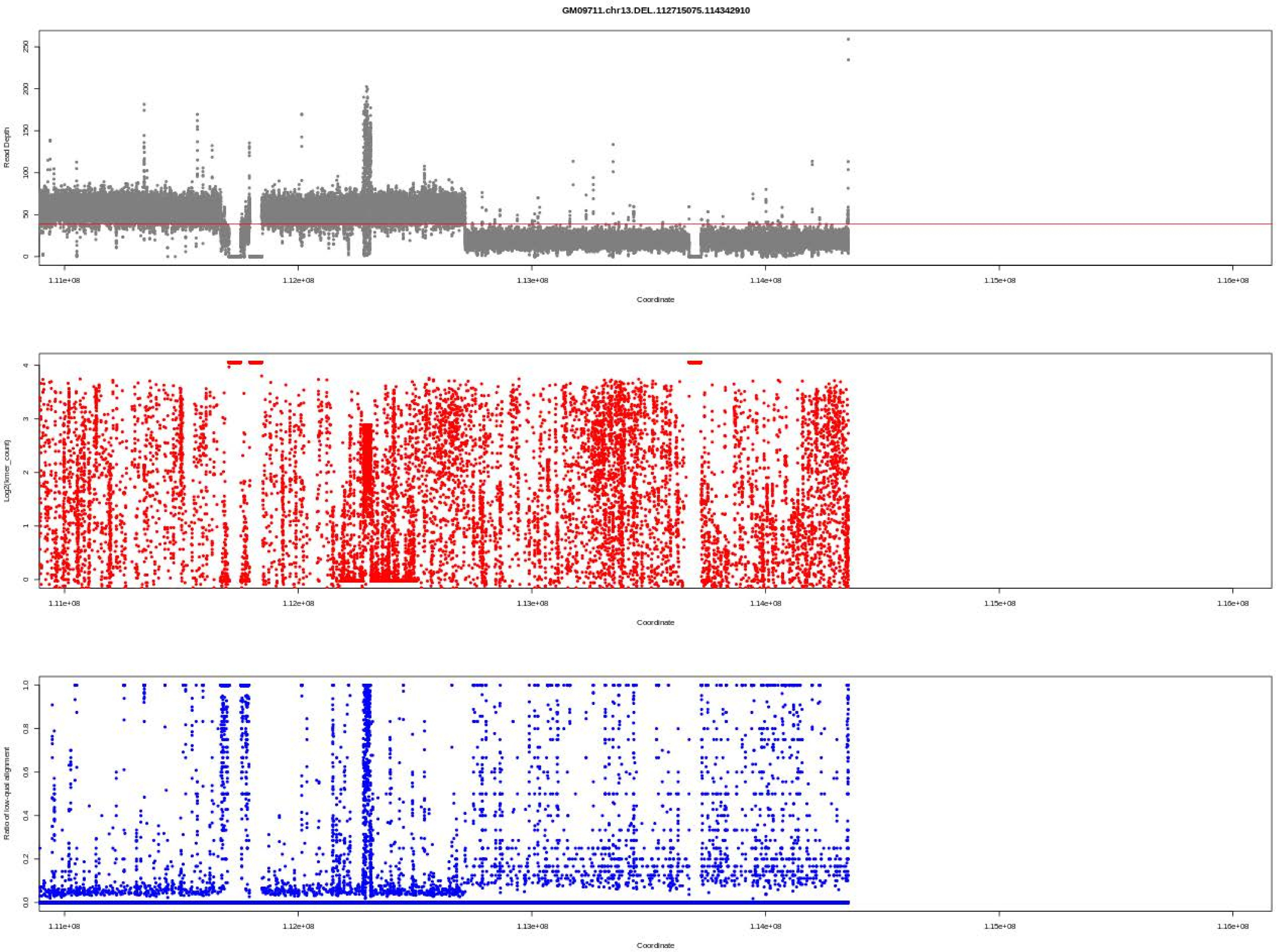
GM09711.chr13.DEL.112715075.114342910
Read Depth
Coordinate
Log2(kmer_count)
Coordinate
Ratio of low-qual alignment
Coordinate
1.11e+08
1.12e+08
1.13e+08
1.14e+08
1.15e+08
1.16e+08

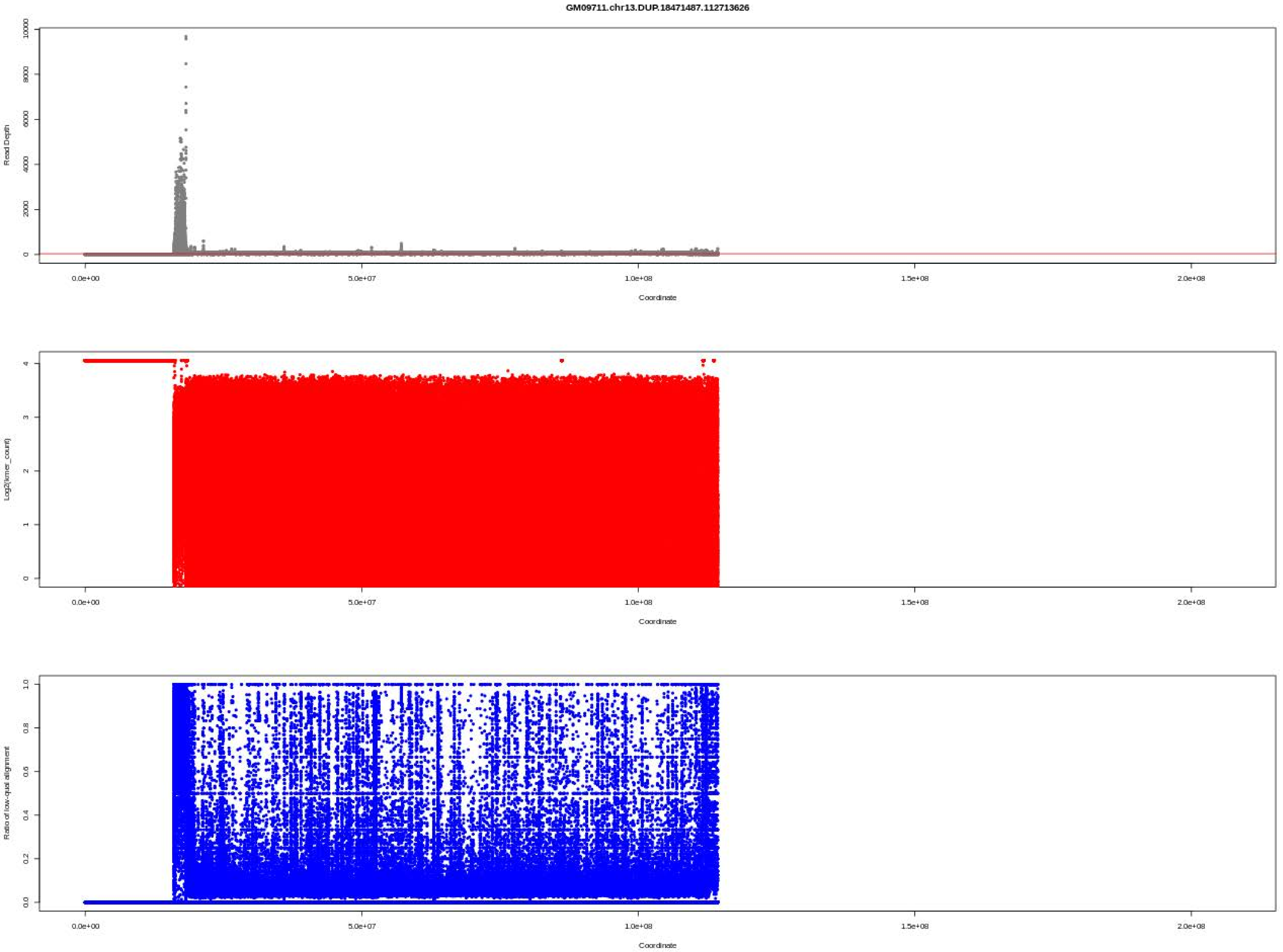
GM09711.chr13.DUP.18471487.112713626
Read Depth
Coordinate
Log2(kmer_count)
Coordinate
Ratio of low-qual alignment
Coordinate

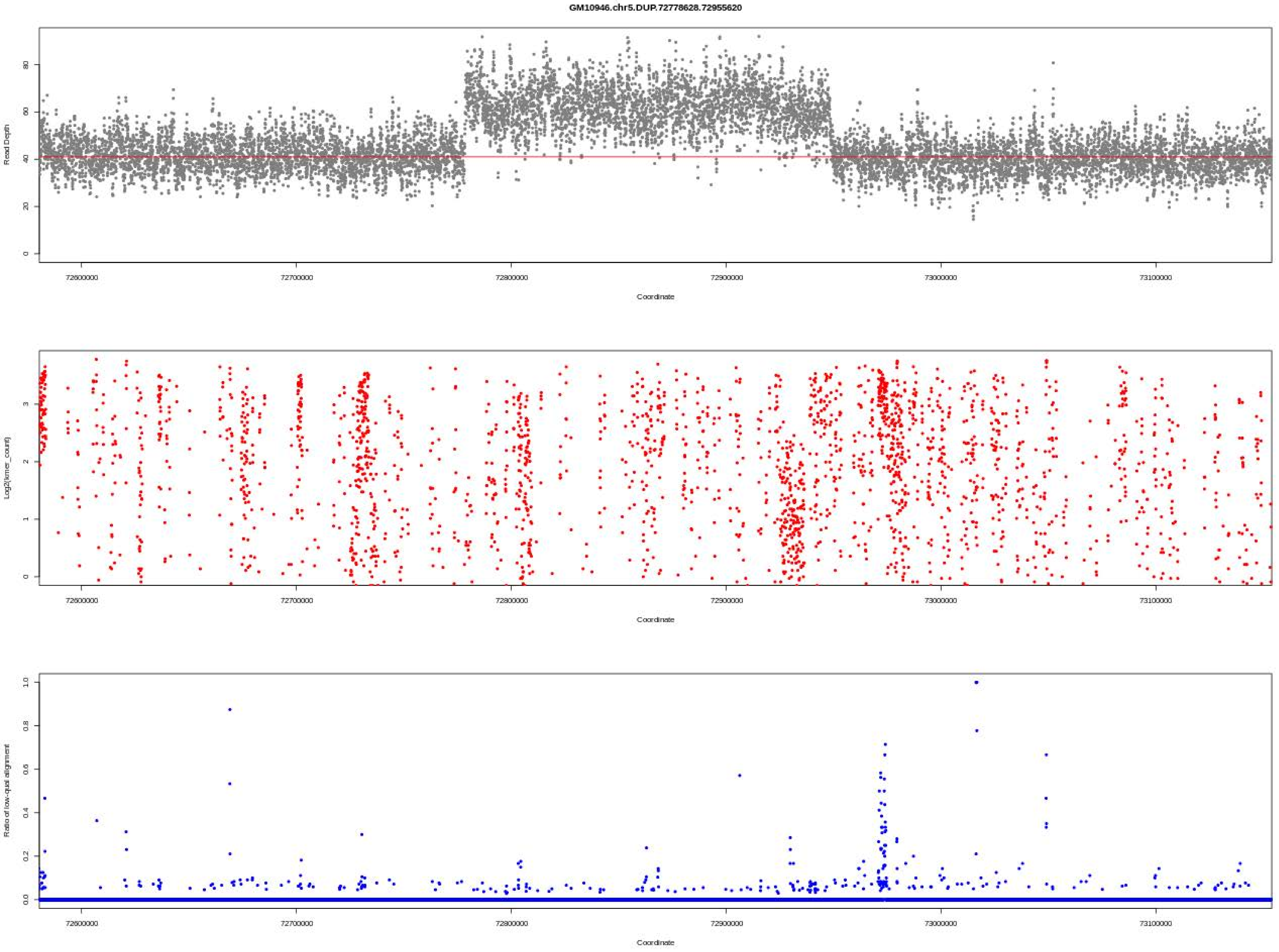
GM10946.chr5.DUP.72778628.72955620
Read Depth
0
20
40
60
80
72600000
72700000
72800000
72900000
73000000
73100000
Coordinate
Log2(kmer_count)
0
1
2
3
72600000
72700000
72800000
72900000
73000000
73100000
Coordinate
Ratio of low-qual alignment
0.0
0.2
0.4
0.6
0.8
1.0
72600000
72700000
72800000
72900000
73000000
73100000
Coordinate

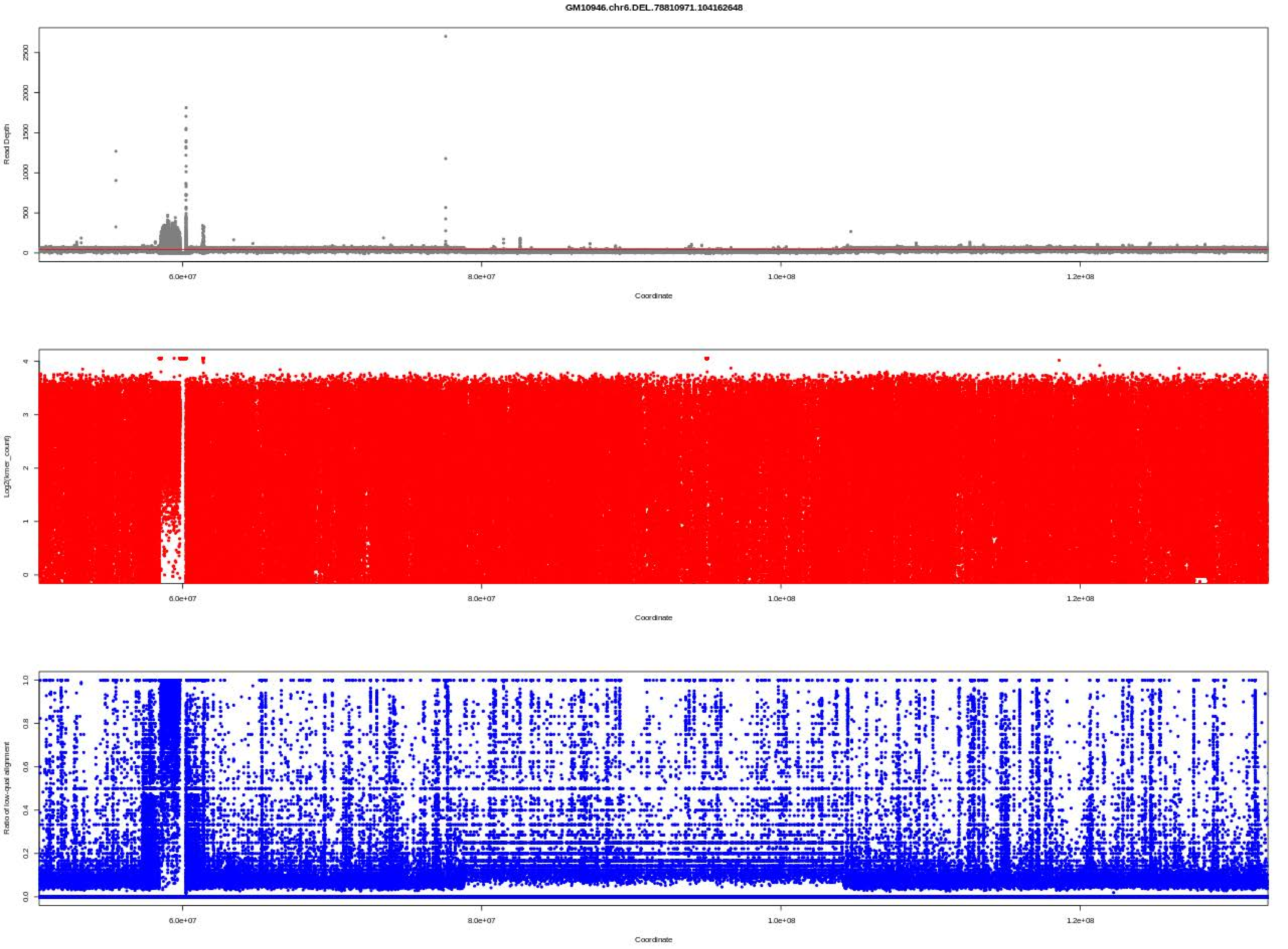
GM10946.chr6.DEL.78810971.104162648
Read Depth
Coordinate
Log2(kmer_count)
Coordinate
Ratio of low-qual alignment
Coordinate

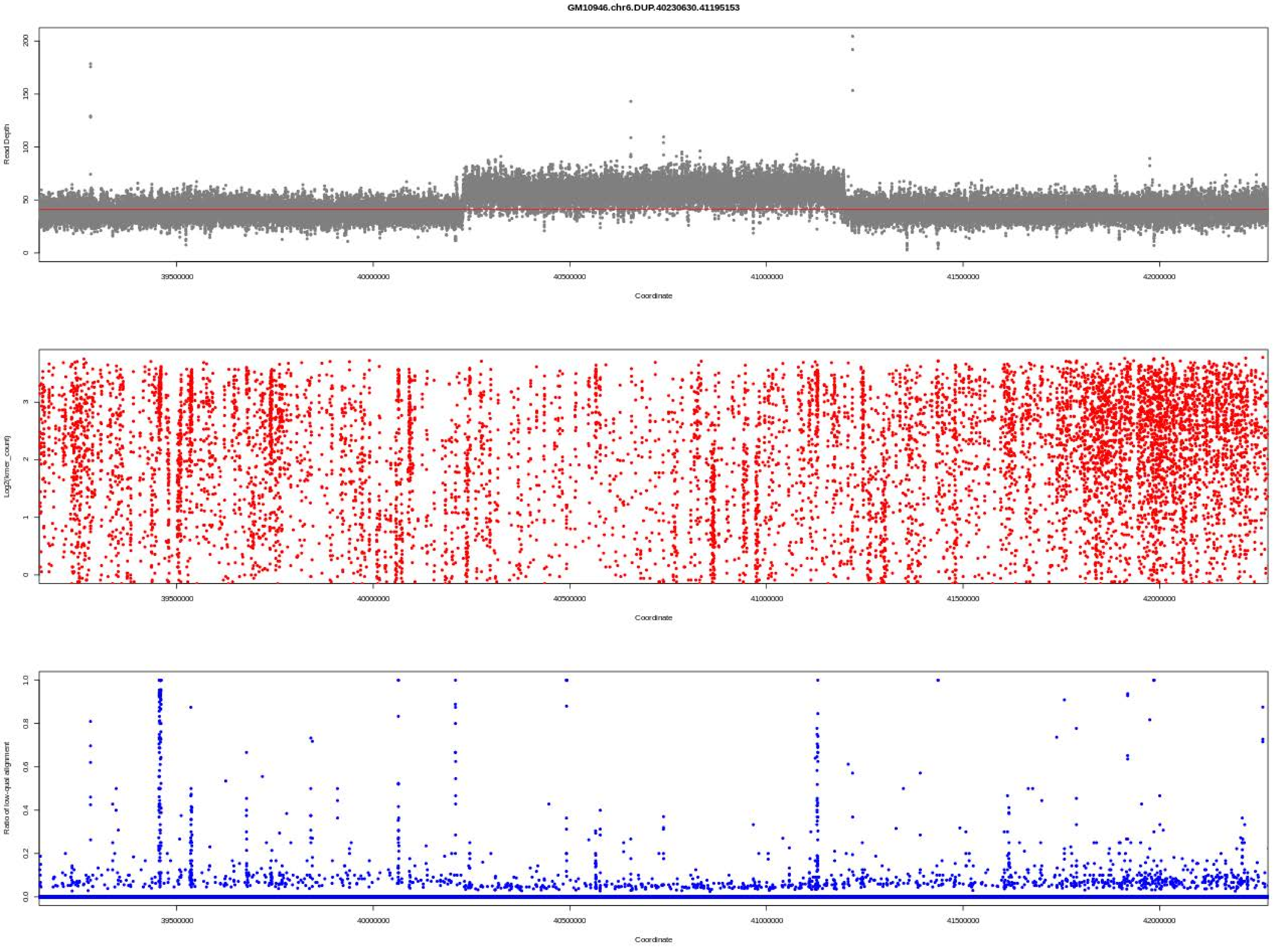
GM10946.chr6.DUP.40230630.41195153
Read Depth
Coordinate
Log2(kmer_count)
Coordinate
Ratio of low-qual alignment
Coordinate
39500000
40000000
40500000
41000000
41500000
42000000

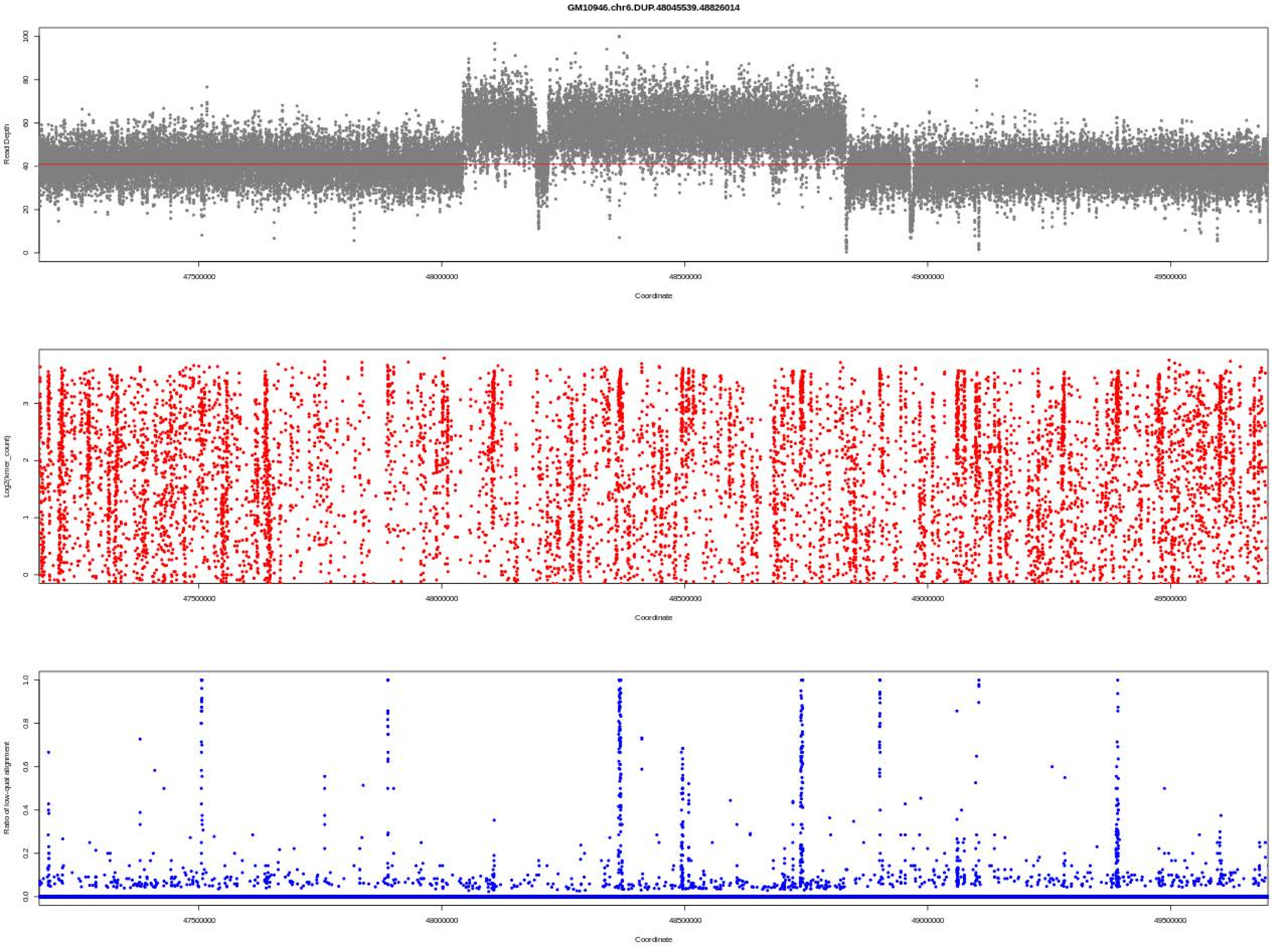
GM10946.chr6.DUP.48045539.48826014
Read Depth
0
20
40
60
80
100
47500000
48000000
48500000
49000000
49500000
Coordinate
Log2(kmer_count)
0
1
2
3
47500000
48000000
48500000
49000000
49500000
Coordinate
Ratio of low-qual alignment
0.0
0.2
0.4
0.6
0.8
1.0
47500000
48000000
48500000
49000000
49500000
Coordinate

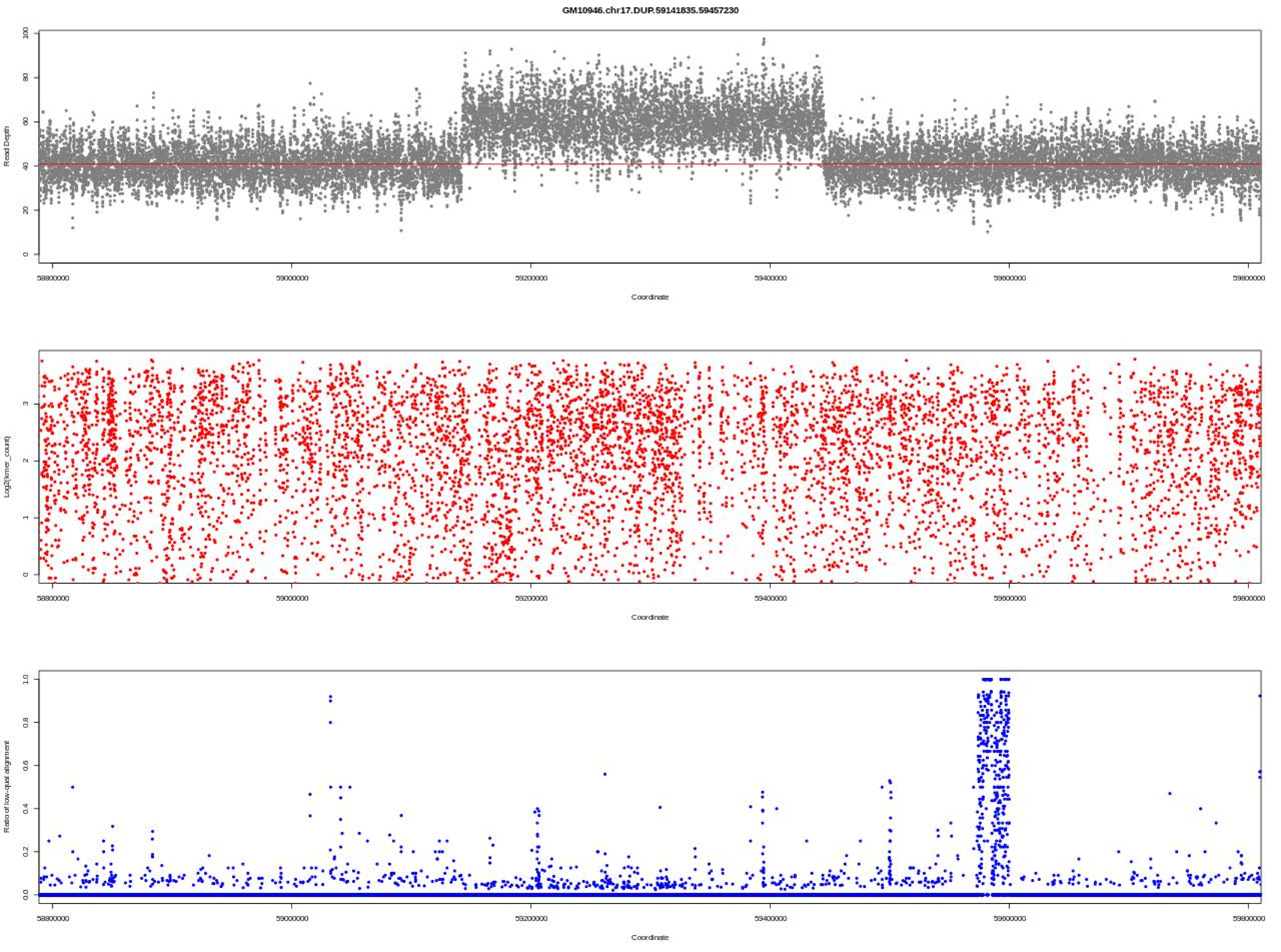
GM10946.chr17.DUP.59141835.59457230
Read Depth
Coordinate
Log2(kmer_count)
Coordinate
Ratio of low-qual alignment
Coordinate

GM11428.chr3.DEL.18654.5327217

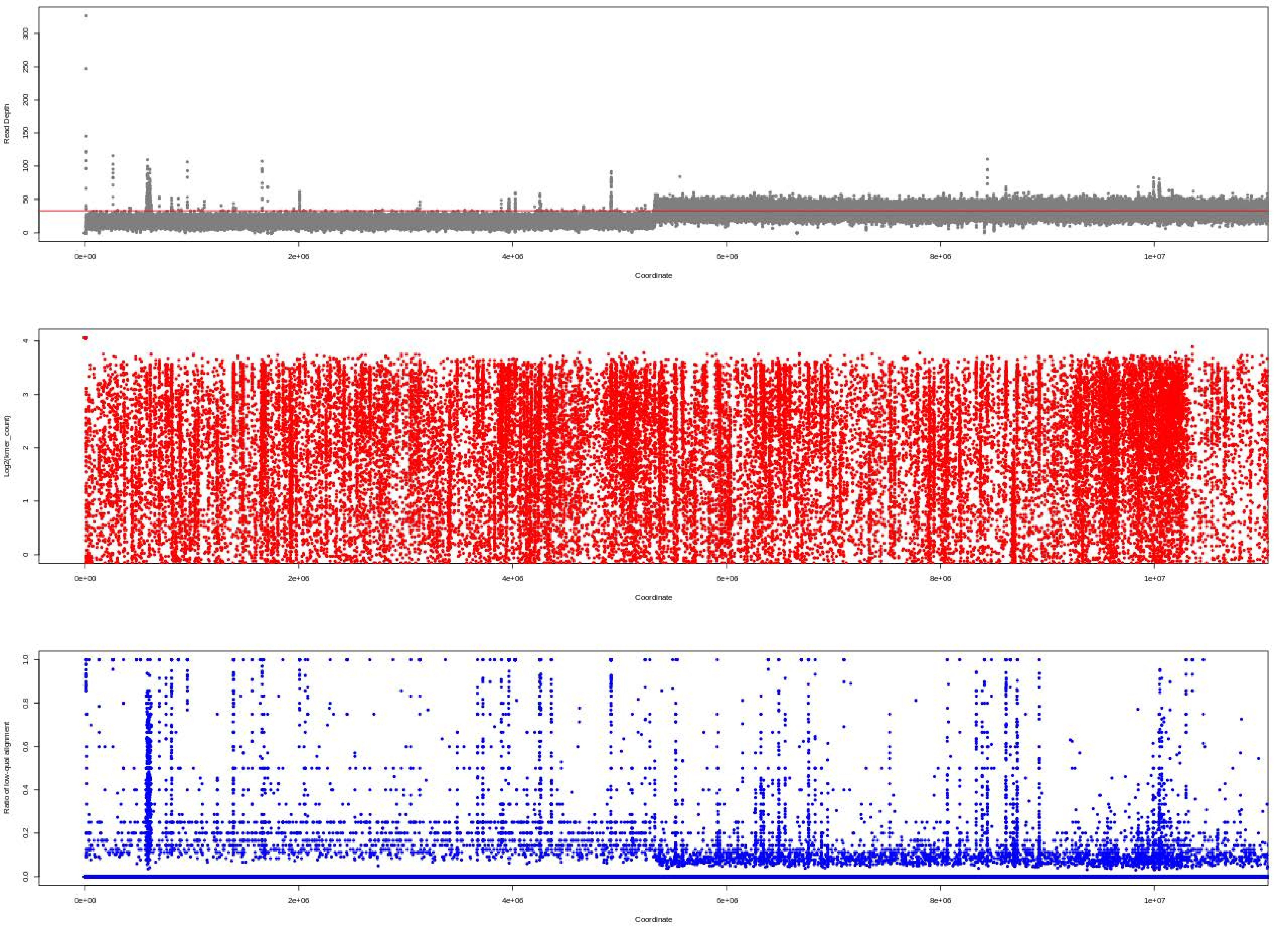

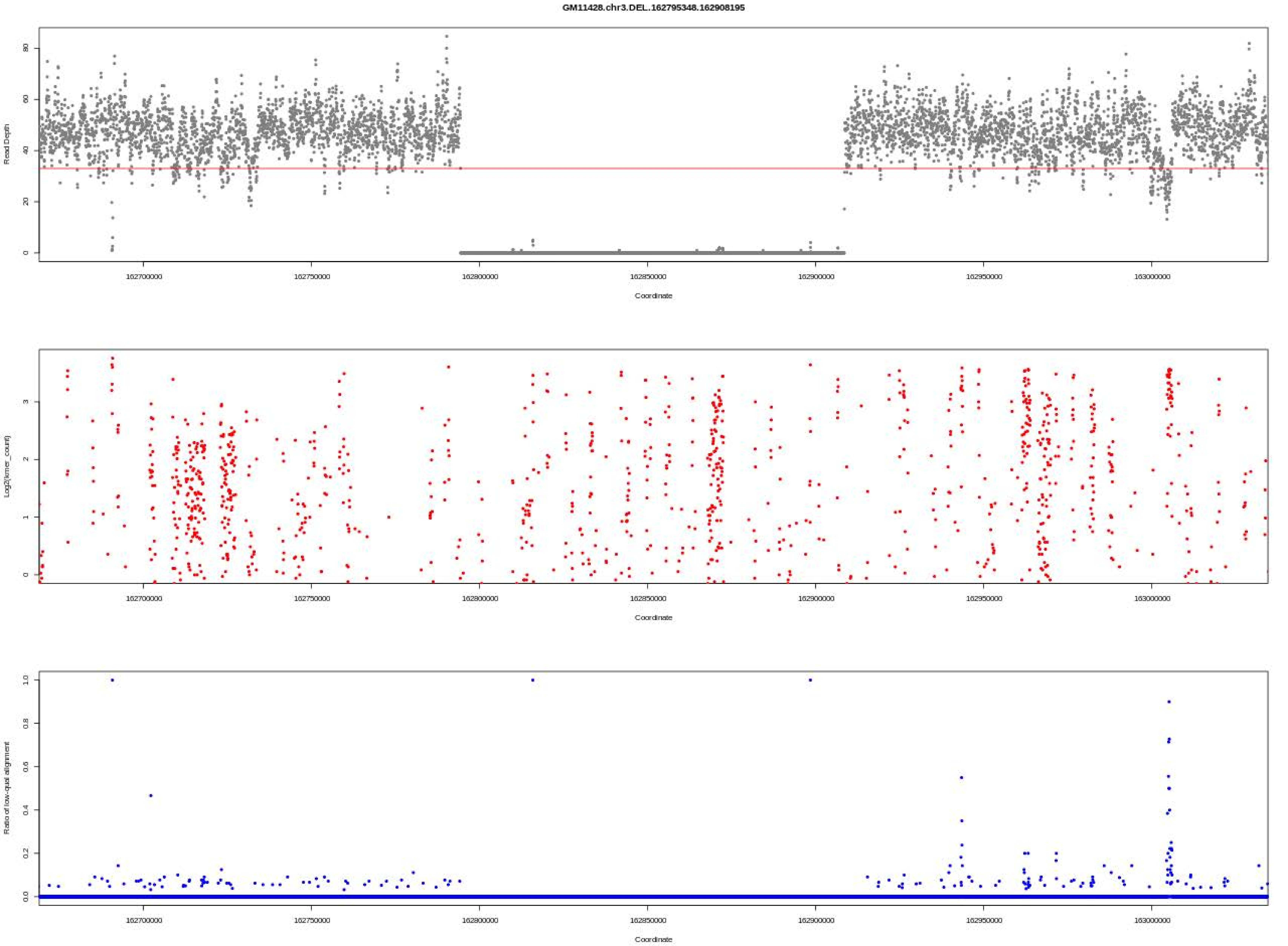
GM11428.chr3.DEL.162795348.162908195
Read Depth
Log2(kmer_count)
Ratio of low-qual alignment
Coordinate
162700000
162750000
162800000
162850000
162900000
162950000
163000000

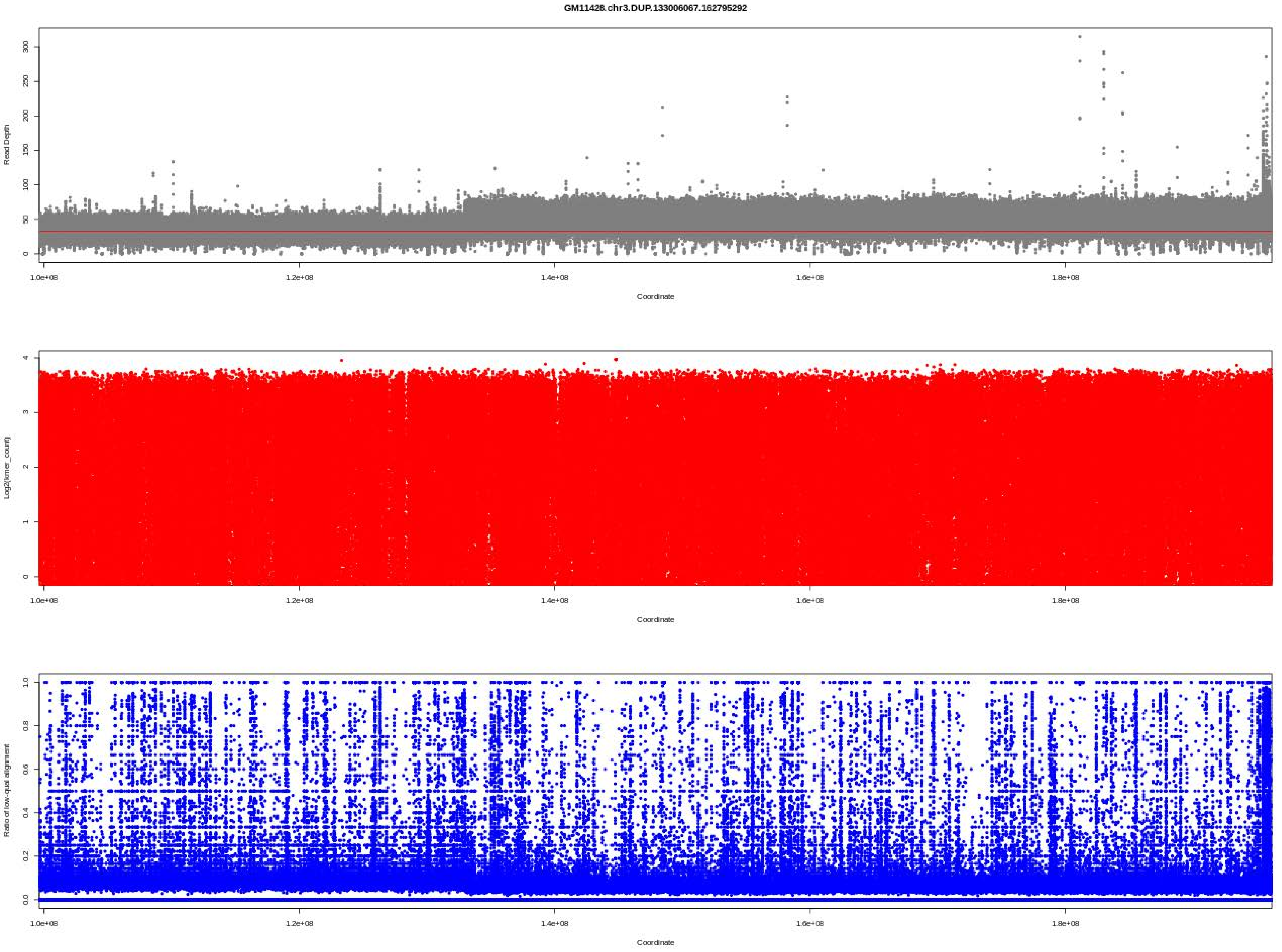
GM11428.chr3.DUP.133006067.162795292
Read Depth
0
50
100
150
200
250
300
1.0e+08
1.2e+08
1.4e+08
1.6e+08
1.8e+08
Coordinate
Log2(kmer_count)
0
1
2
3
4
1.0e+08
1.2e+08
1.4e+08
1.6e+08
1.8e+08
Coordinate
Ratio of low-qual alignment
0.0
0.2
0.4
0.6
0.8
1.0
1.0e+08
1.2e+08
1.4e+08
1.6e+08
1.8e+08
Coordinate

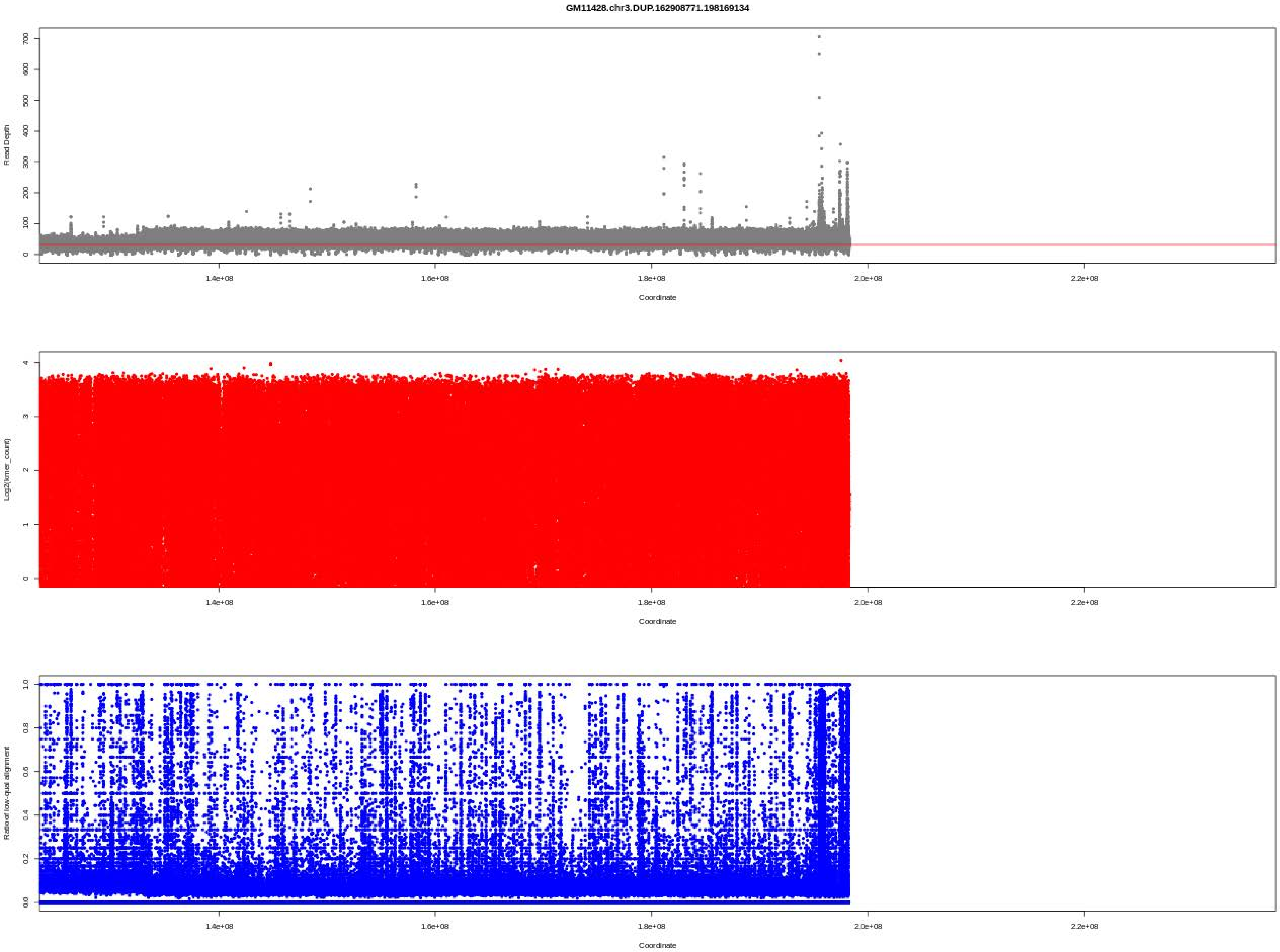
GM11428.chr3.DUP.162908771.198169134
Read Depth
Coordinate
Log2(kmer_count)
Coordinate
Ratio of low-qual alignment
Coordinate
1.4e+08
1.6e+08
1.8e+08
2.0e+08
2.2e+08

GM11516.chr14.DEL.105780376.106771352

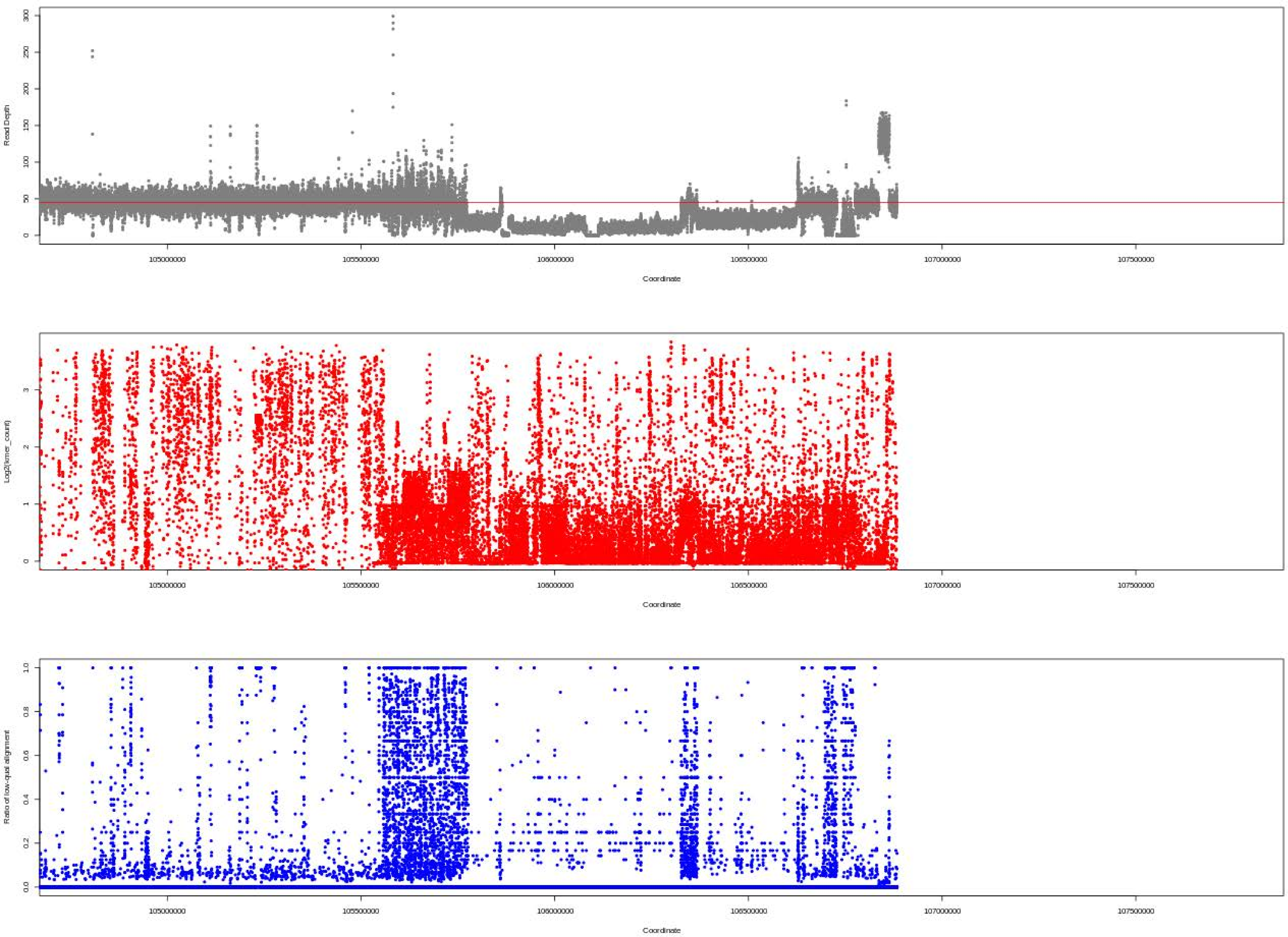

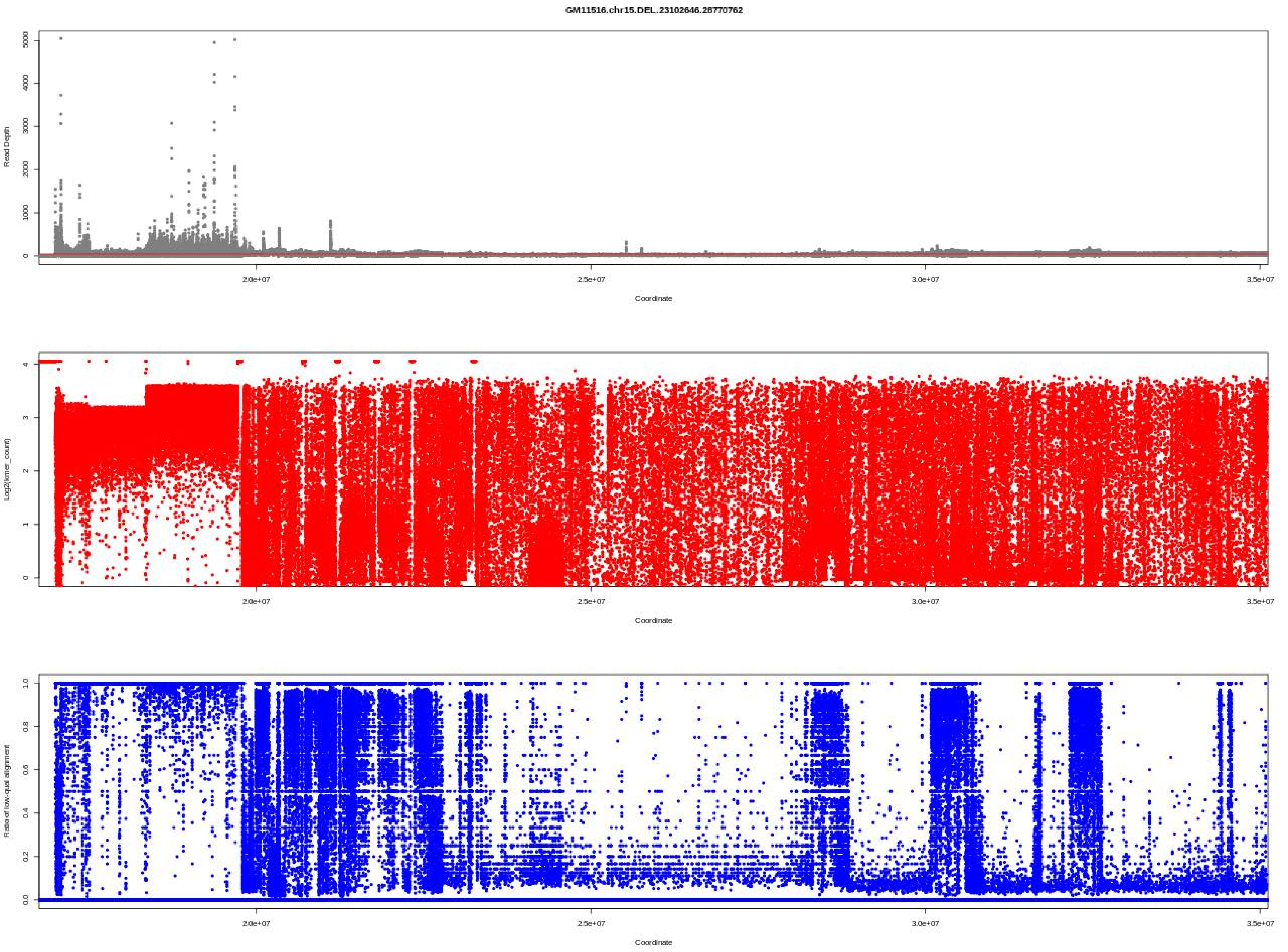
GM11516.chr15.DEL..23102646.28770762
Read Depth
Coordinate
Log2(kmer_count)
Coordinate
Ratio of low-qual alignment
Coordinate

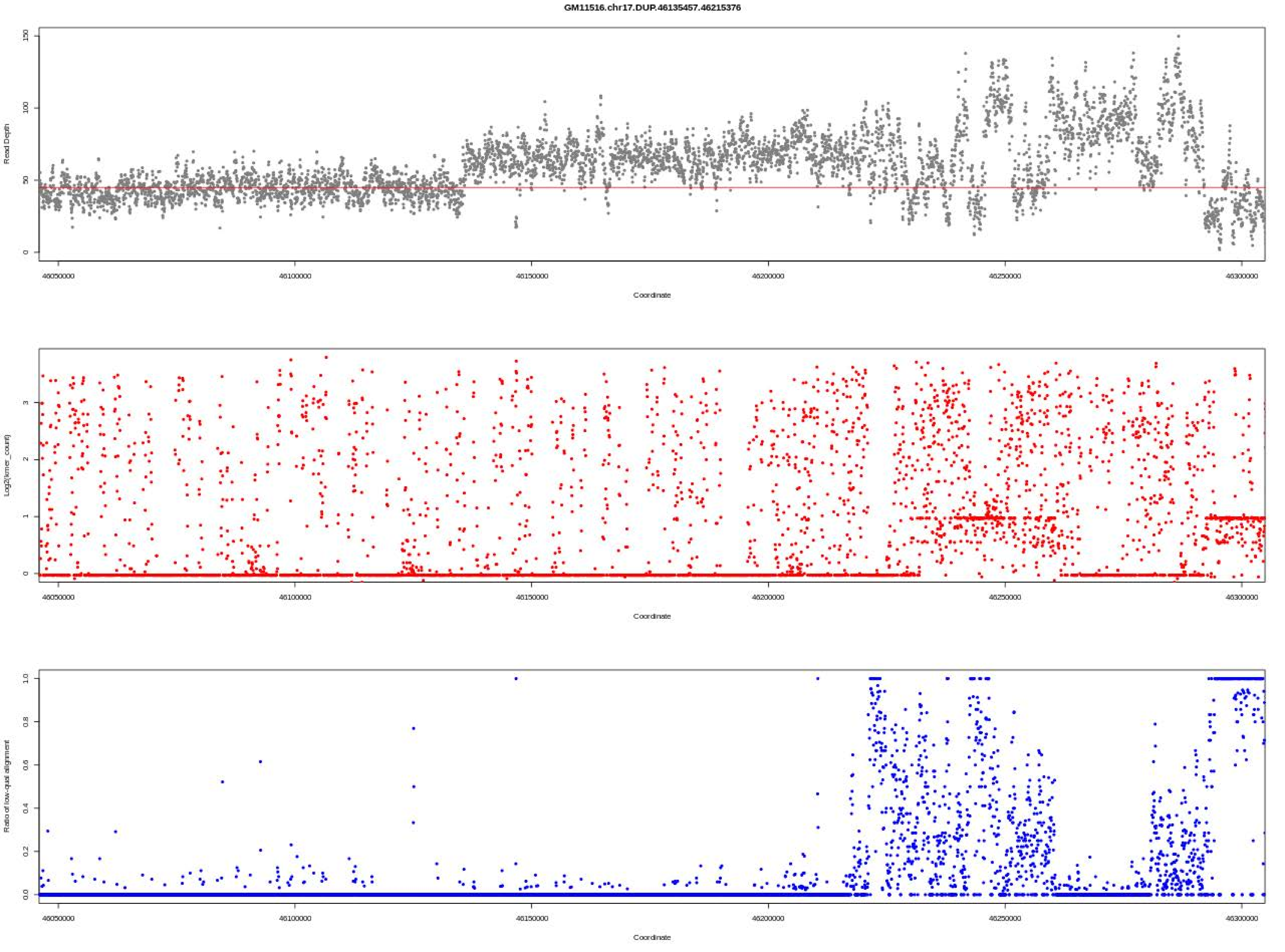

GM11516.chr17.DUP.46135457.46215376
Read Depth
0
50
100
150
Coordinate
46050000
46100000
46150000
46200000
46250000
46300000
Log2(kmer_count)
0
1
2
3
Ratio of low-qual alignment
0.0
0.2
0.4
0.6
0.8
1.0

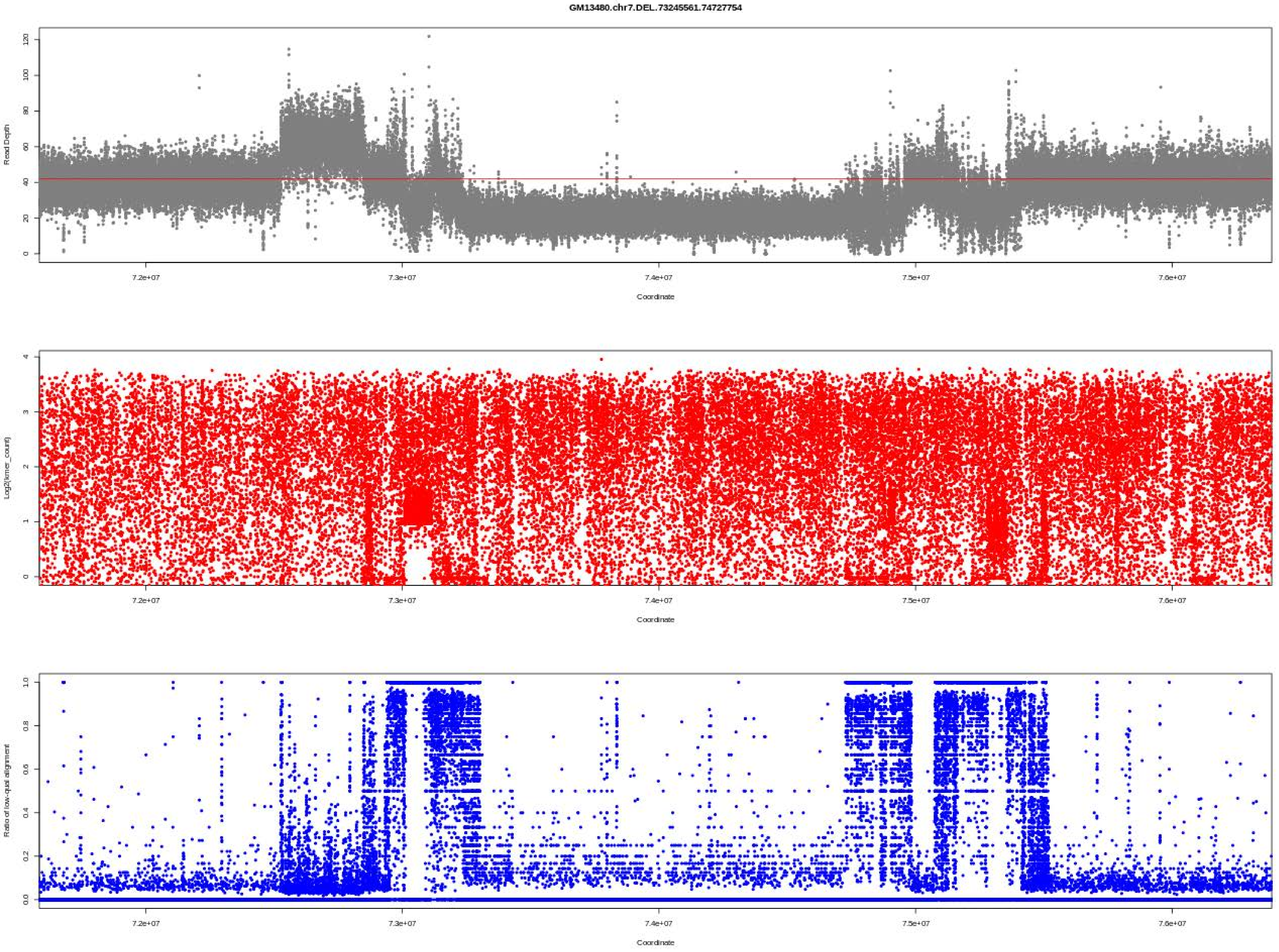

GM13480.chr7.DEL.73245561.74727754
Read Depth
Coordinate
Log2(kmer_count)
Coordinate
Ratio of low-qual alignment
Coordinate
7.2e+07
7.3e+07
7.4e+07
7.5e+07
7.6e+07

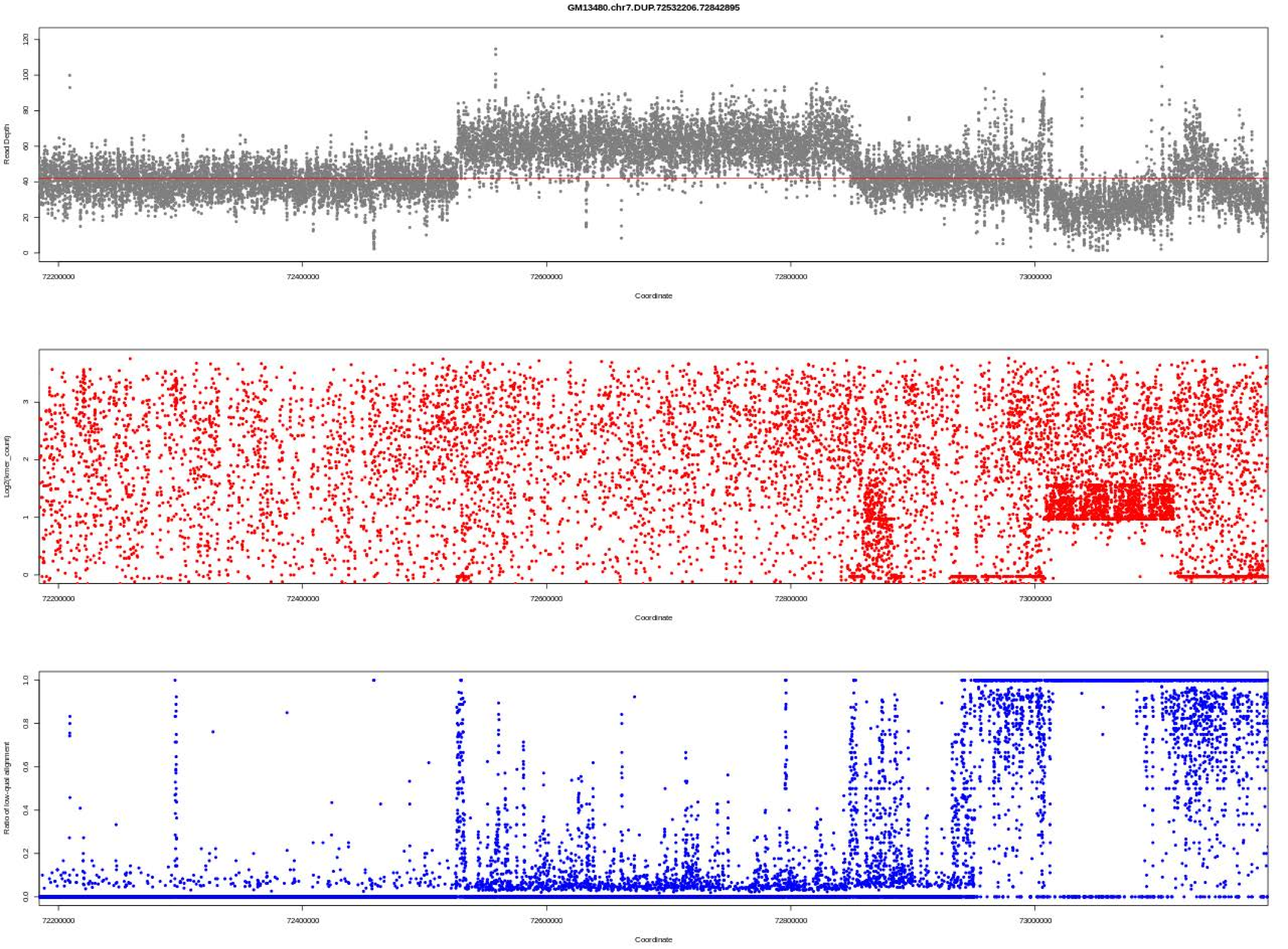

GM13480.chr7.DUP.72532206.72842895
Read Depth
Coordinate
Log2(kmer_count)
Coordinate
Ratio of low-qual alignment
Coordinate

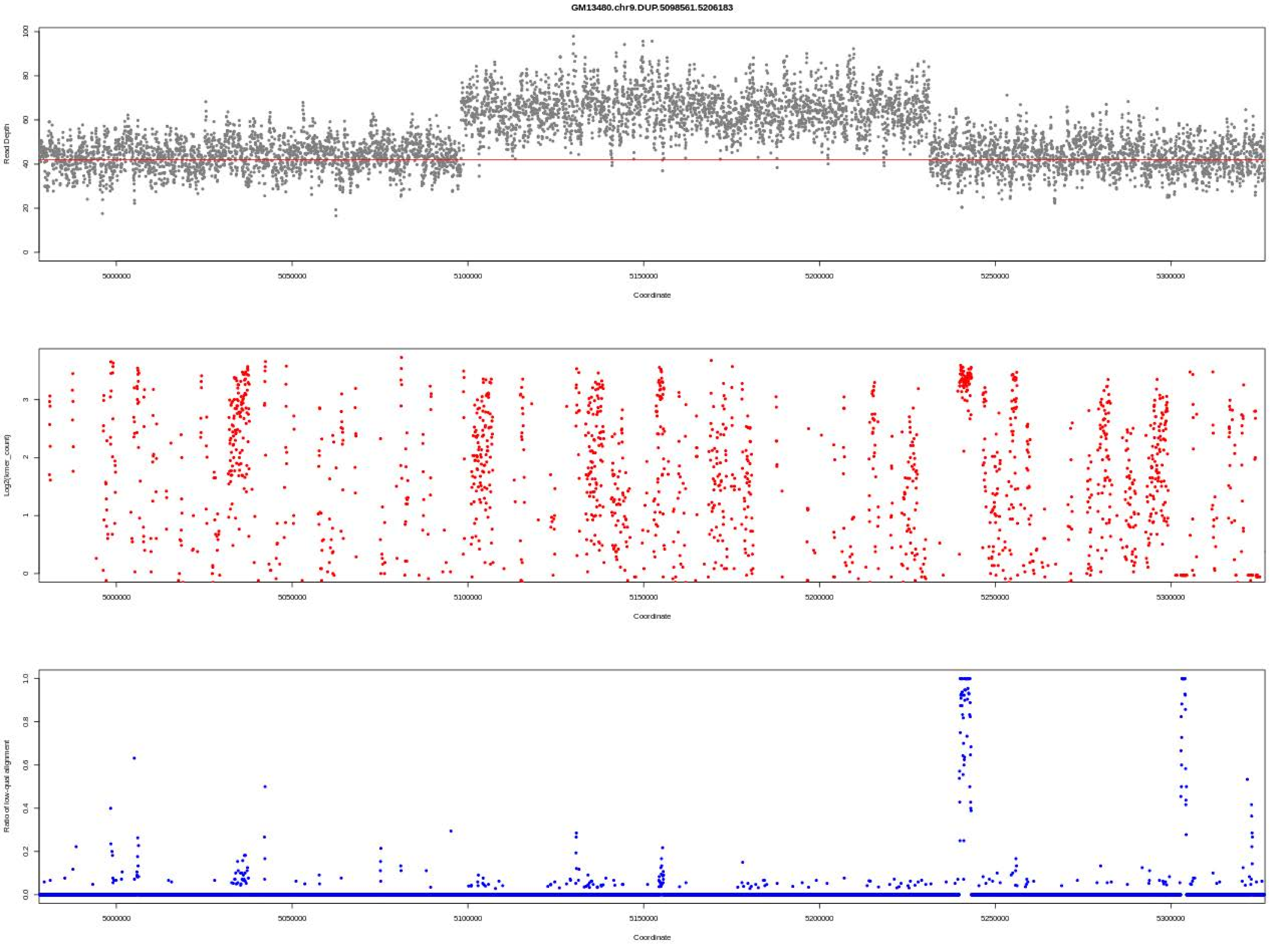
GM13480.chr9.DUP.5098561.5206183
Read Depth
Coordinate
Log2(kmer_count)
Coordinate
Ratio of low-qual alignment
Coordinate

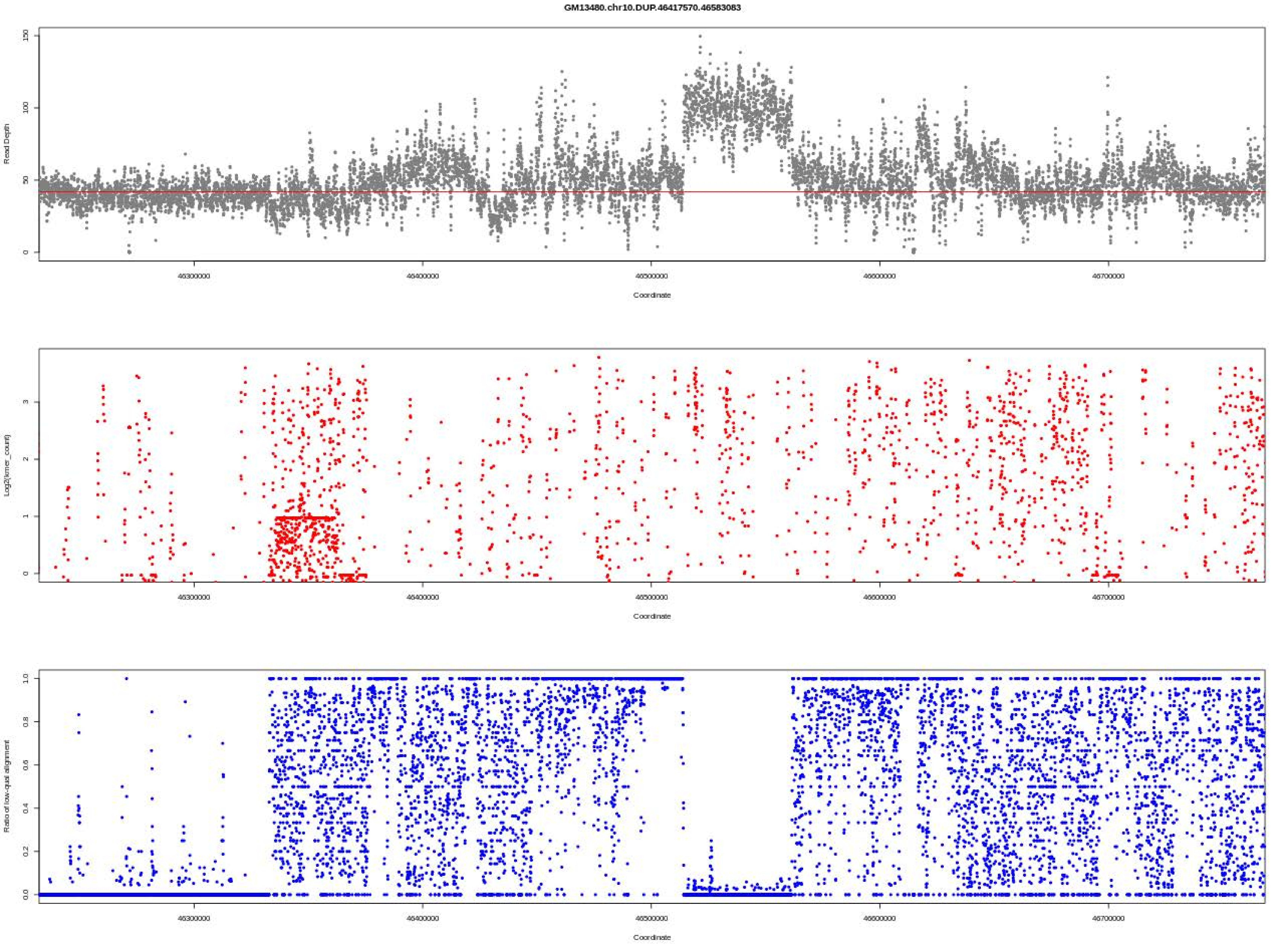
GM13480.chr10.DUP.46417570.46583083
Read Depth
Coordinate
Log2(kmer_count)
Coordinate
Ratio of low-qual alignment
Coordinate

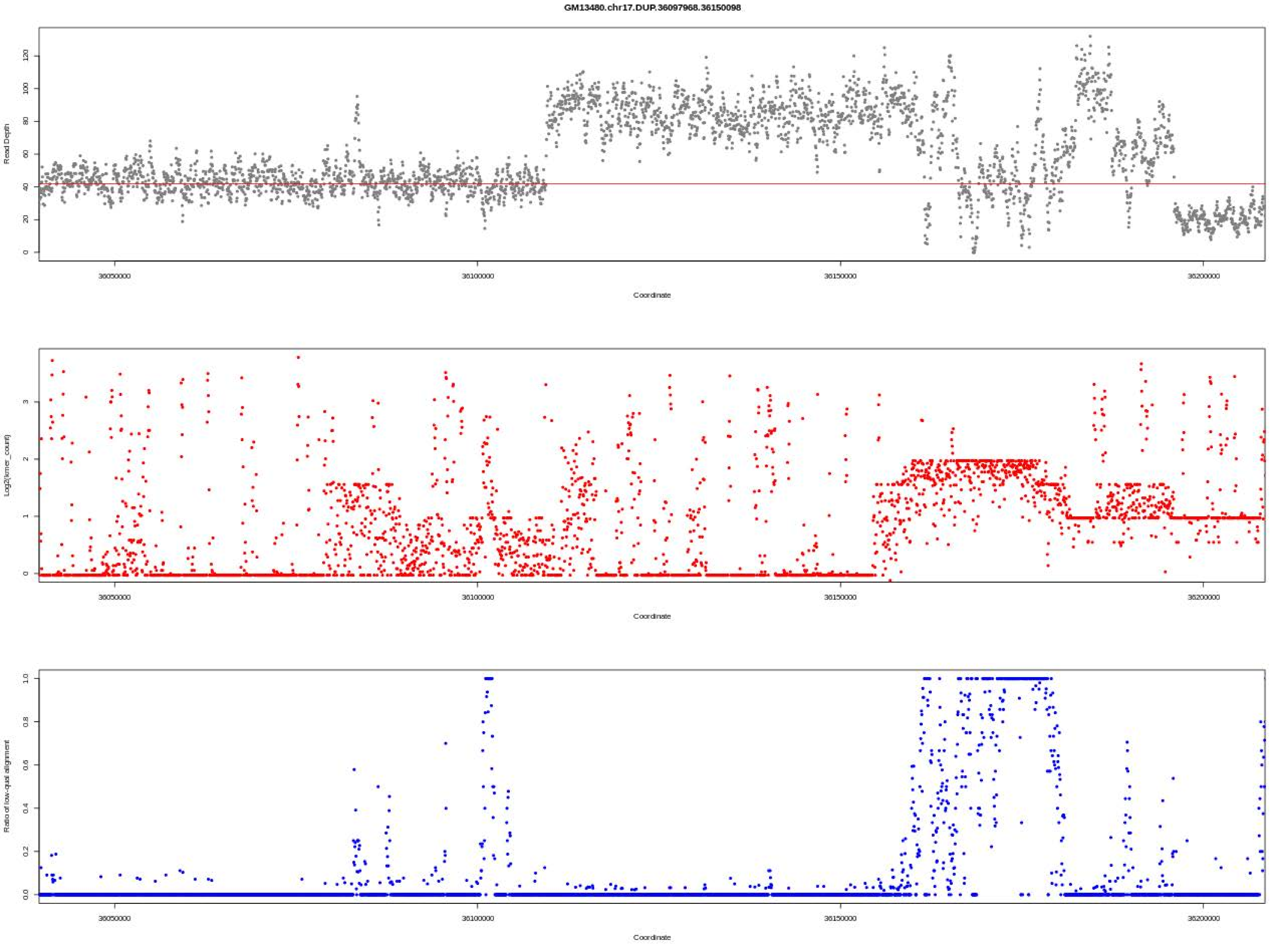

GM13480.chr17.DUP.36097968.36150098
Read Depth
Coordinate
Log2(kmer_count)
Coordinate
Ratio of low-qual alignment
Coordinate

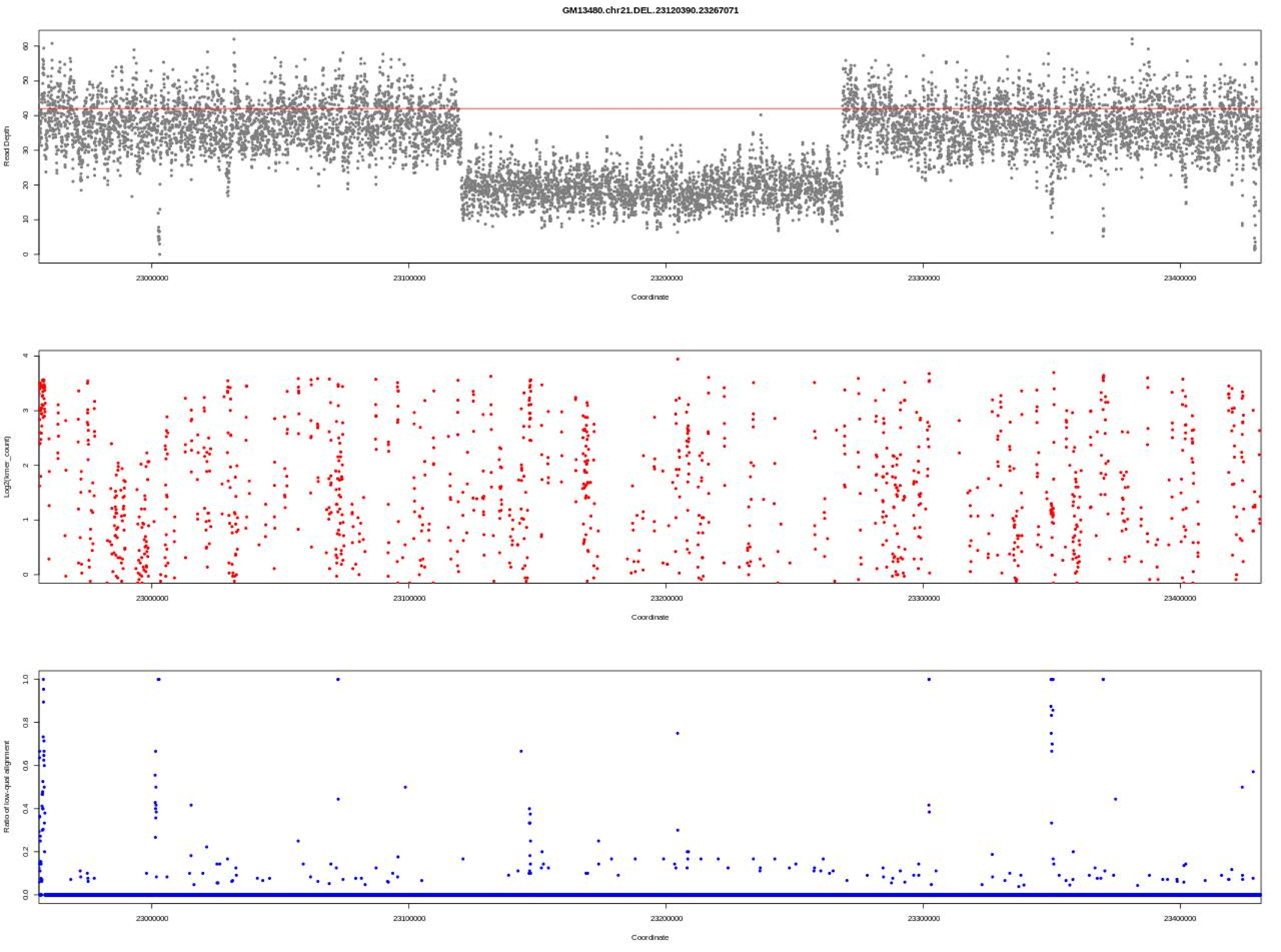
GM13480.chr21.DEL.23120390.23267071
Read Depth
Coordinate
Log2(kmer_count)
Coordinate
Ratio of low-qual alignment
Coordinate

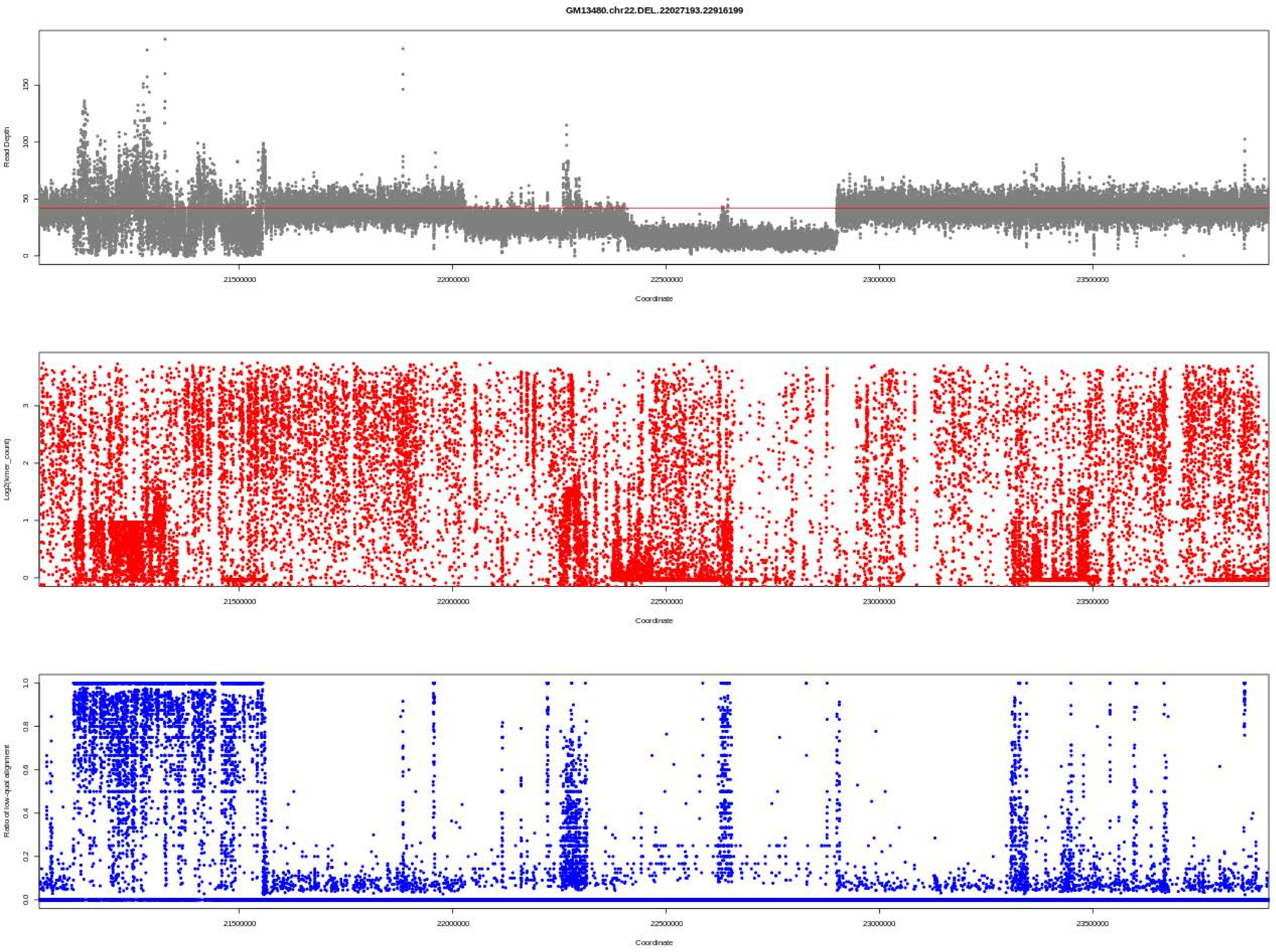
GM13480.chr22.DEL..22027193.22916199
Read Depth
0
50
100
150
21500000
22000000
22500000
23000000
23500000
Coordinate
Log2(kmer_count)
0
1
2
3
21500000
22000000
22500000
23000000
23500000
Coordinate
Ratio of low-qual alignment
0.0
0.2
0.4
0.6
0.8
1.0
21500000
22000000
22500000
23000000
23500000
Coordinate

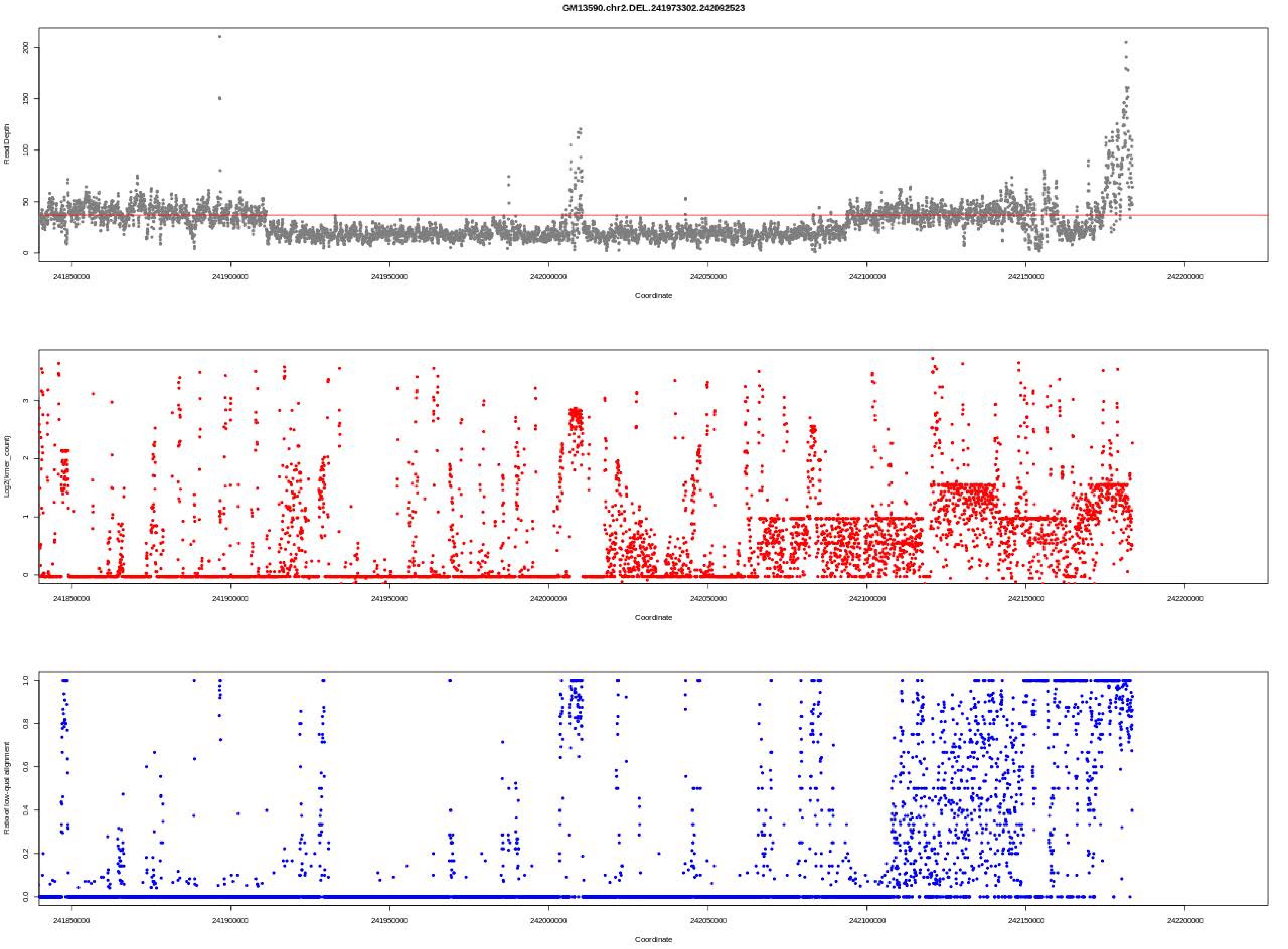
GM13590.chr2.DEL.241973302.242092523
Read Depth
Coordinate
Log2(kmer_count)
Coordinate
Ratio of low-qual alignment
Coordinate
241850000
241900000
241950000
242000000
242050000
242100000
242150000
242200000

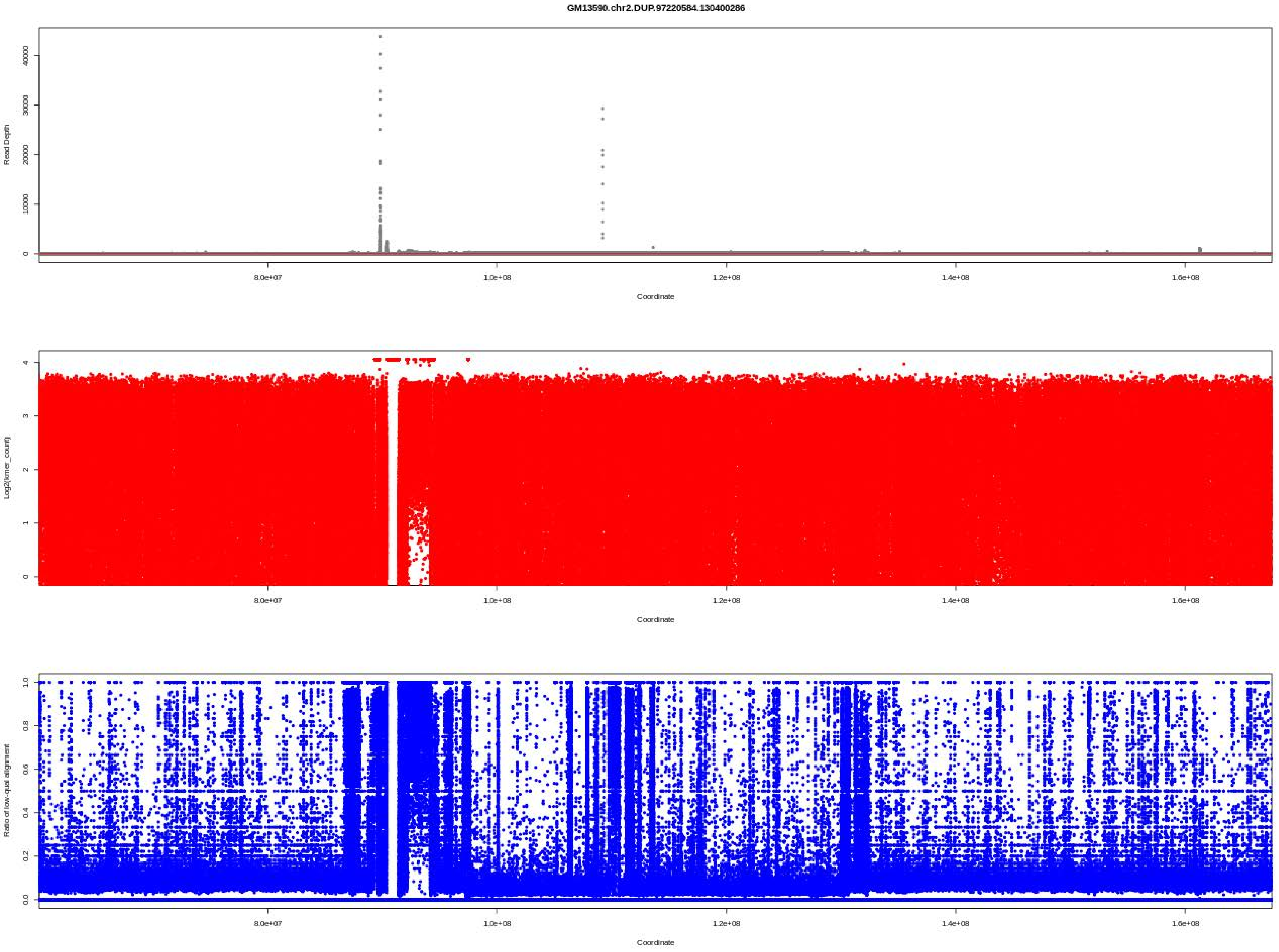
GM13590.chr2.DUP.97220584.130400286
Read Depth
0
10000
20000
30000
40000
8.0e+07
1.0e+08
1.2e+08
1.4e+08
1.6e+08
Coordinate
Log2(kmer_count)
0
1
2
3
4
8.0e+07
1.0e+08
1.2e+08
1.4e+08
1.6e+08
Coordinate
Ratio of low-qual alignment
0.0
0.2
0.4
0.6
0.8
1.0
8.0e+07
1.0e+08
1.2e+08
1.4e+08
1.6e+08
Coordinate

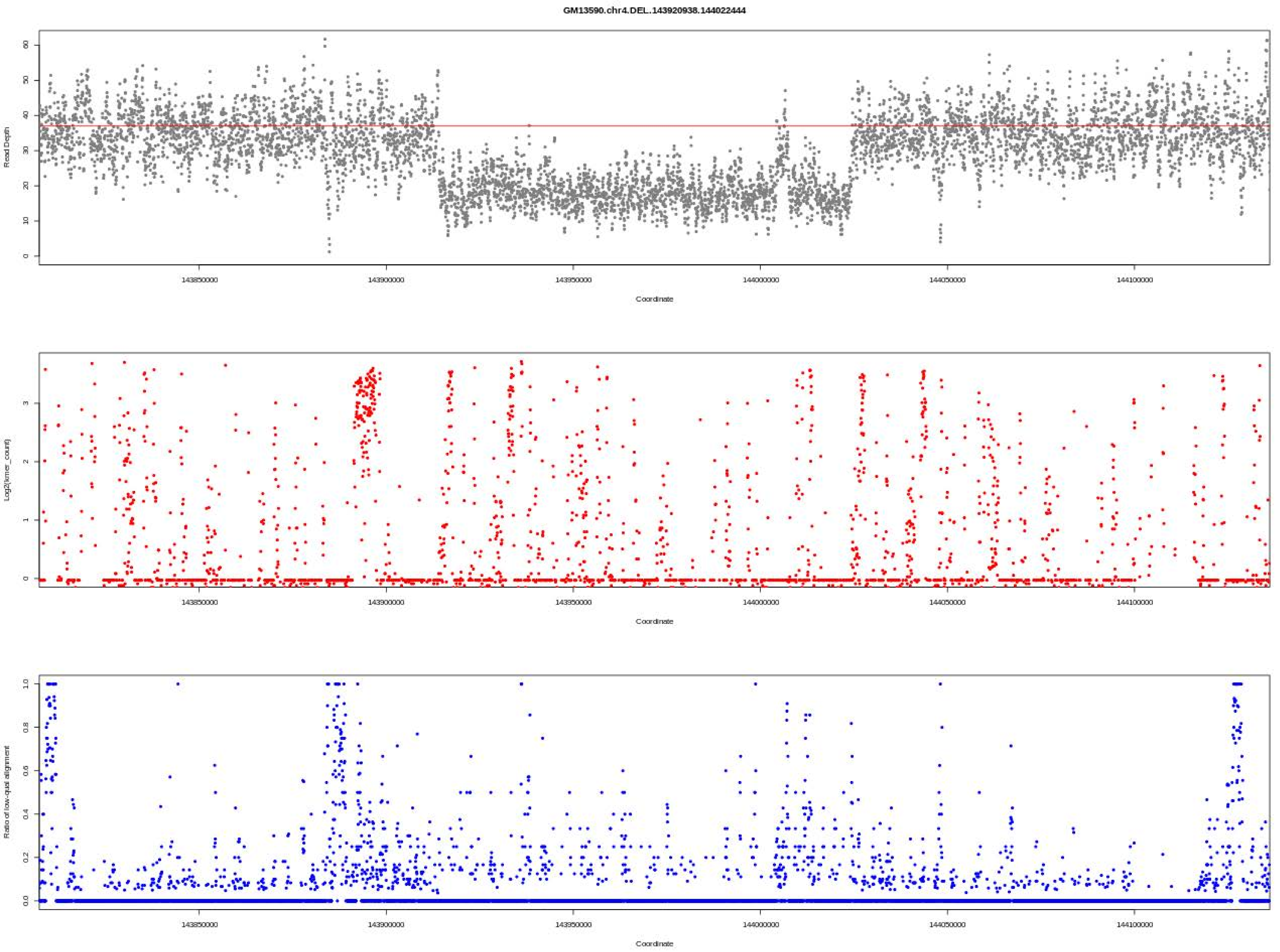
GM13590.chr4.DEL.143920938.144022444
Read Depth
Coordinate
143850000
143900000
143950000
144000000
144050000
144100000
Log2(kmer_count)
Ratio of low-qual alignment

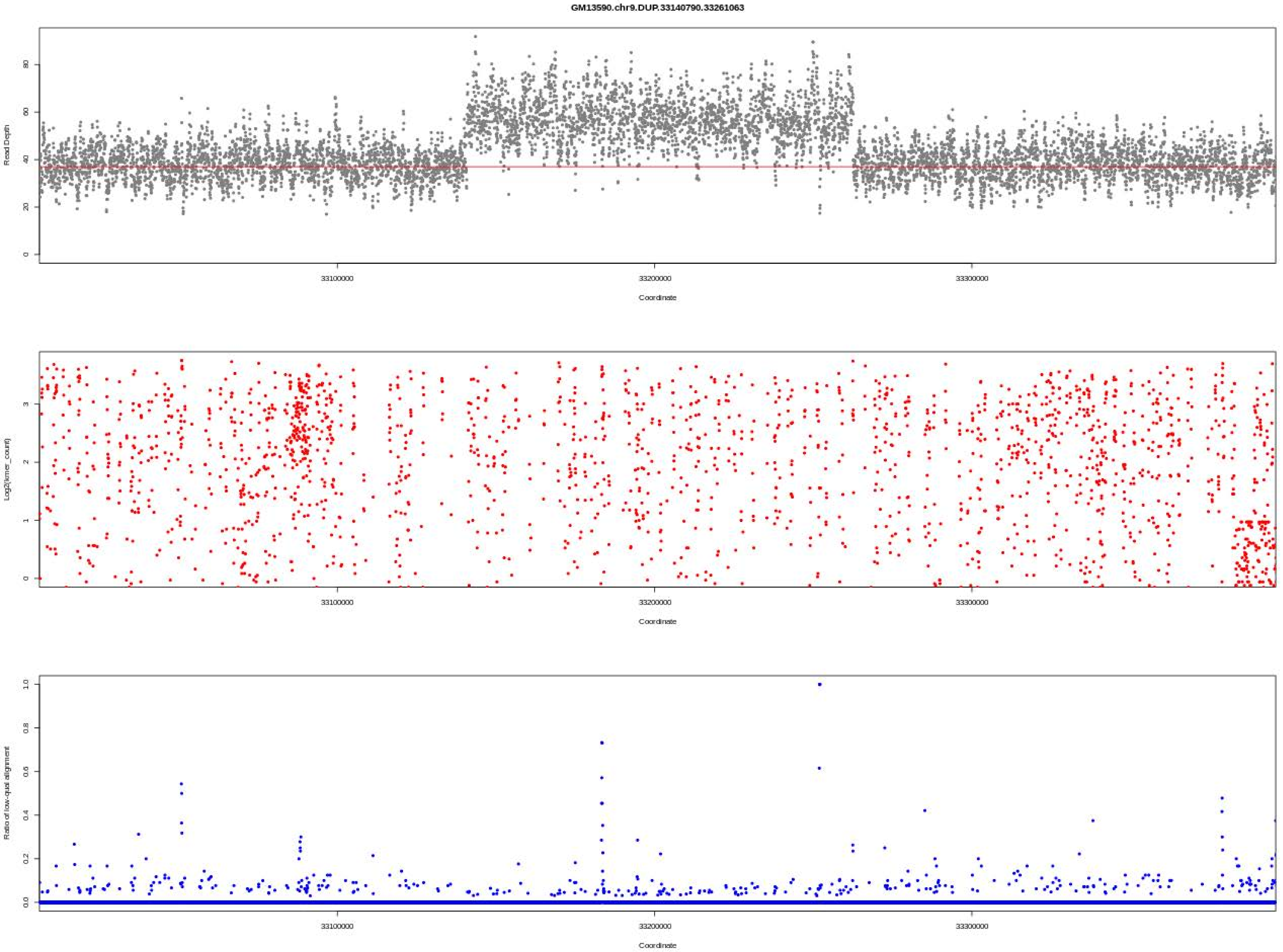

GM13590.chr9.DUP.33140790.33261063
Read Depth
Coordinate
Log2(kmer_count)
Coordinate
Ratio of low-qual alignment
Coordinate

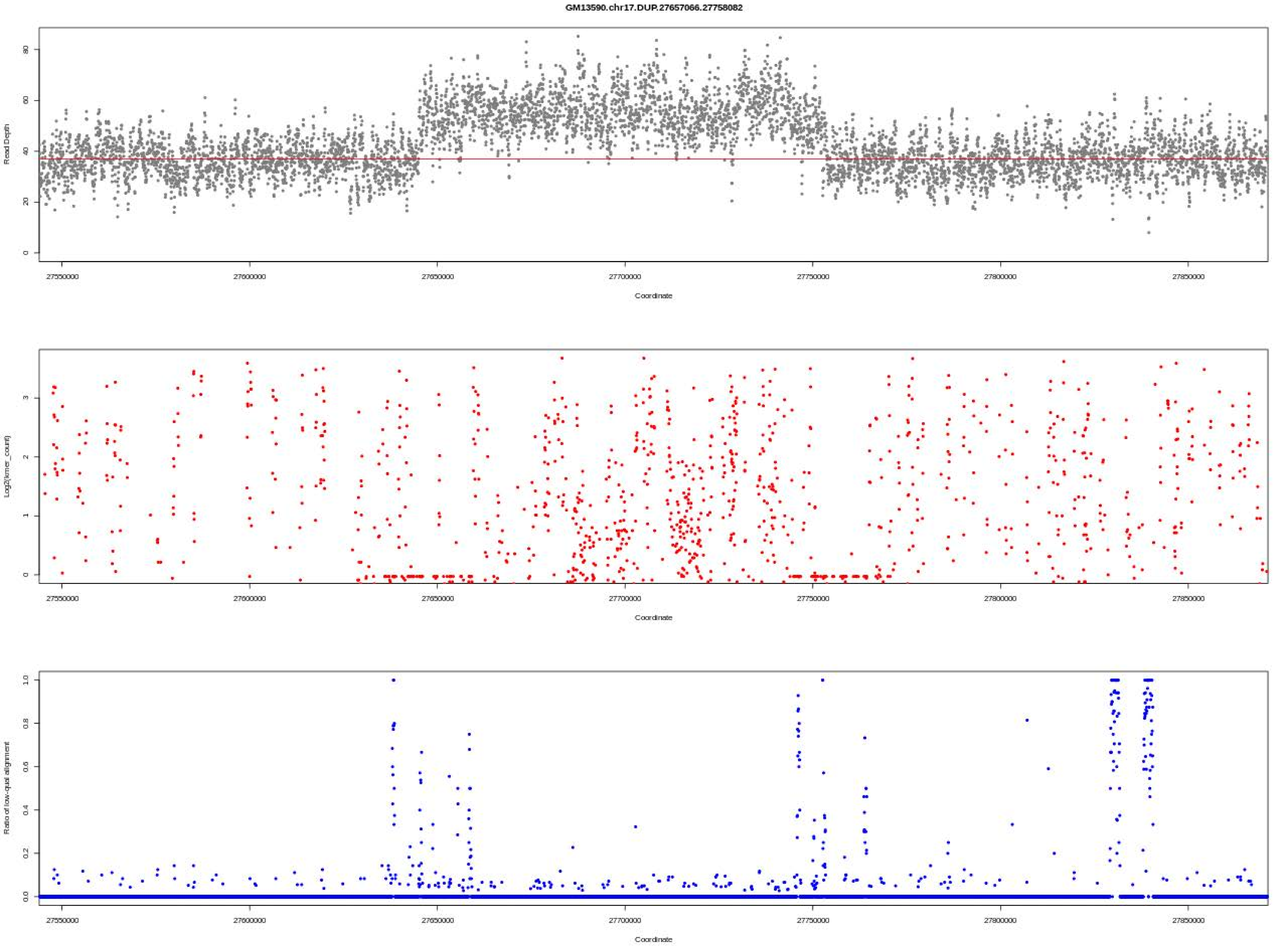
GM13590.chr17.DUP.27657066.27758082
Read Depth
0
20
40
60
80
27550000
27600000
27650000
27700000
27750000
27800000
27850000
Coordinate
Log2(kmer_count)
0
1
2
3
Coordinate
Ratio of low-qual alignment
0.0
0.2
0.4
0.6
0.8
1.0
Coordinate

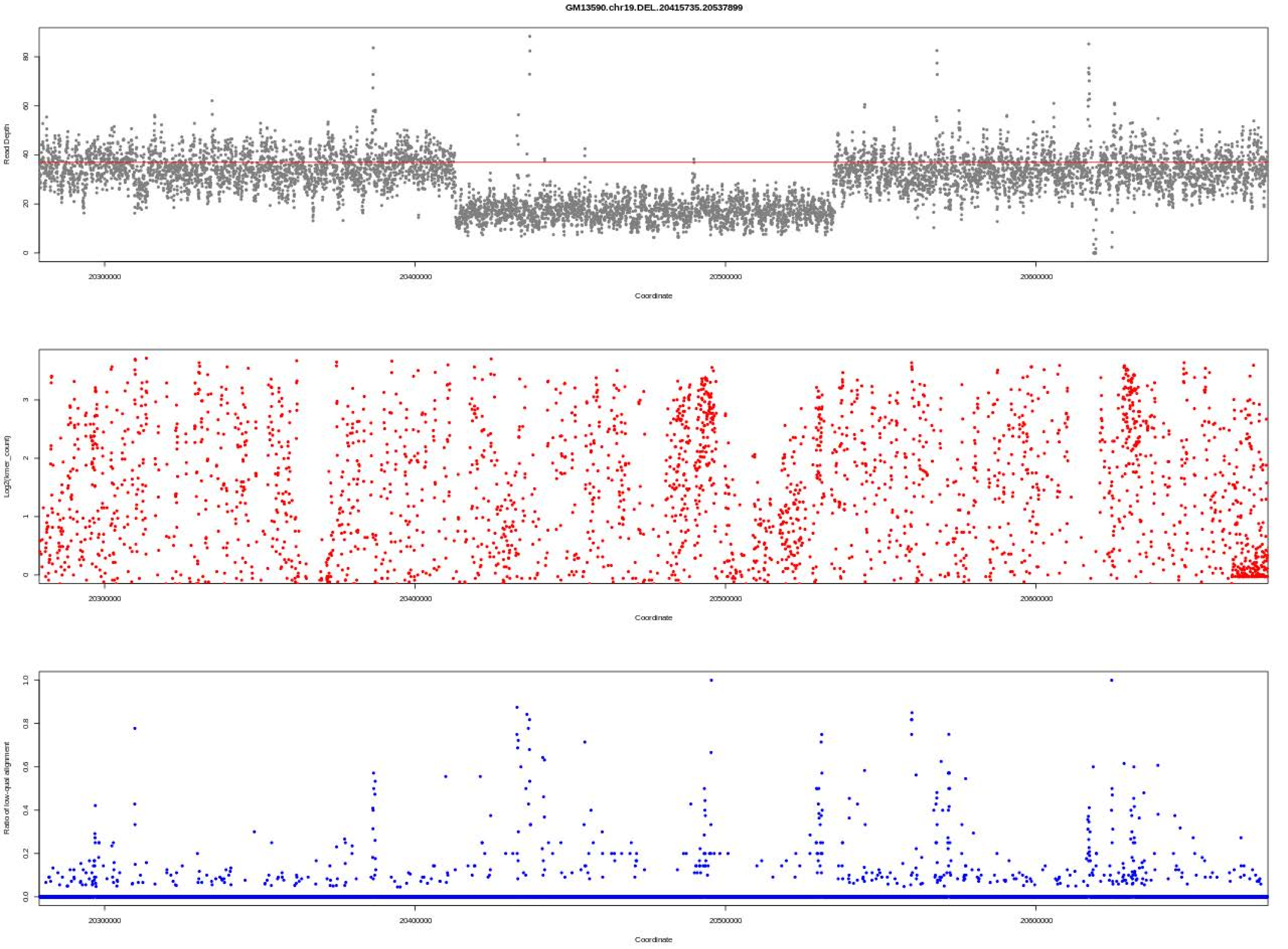

GM13590.chr19.DEL..20415735.20537899
Read Depth
Coordinate
Log2(kmer_count)
Coordinate
Ratio of low-qual alignment
Coordinate
20300000
20400000
20500000
20600000

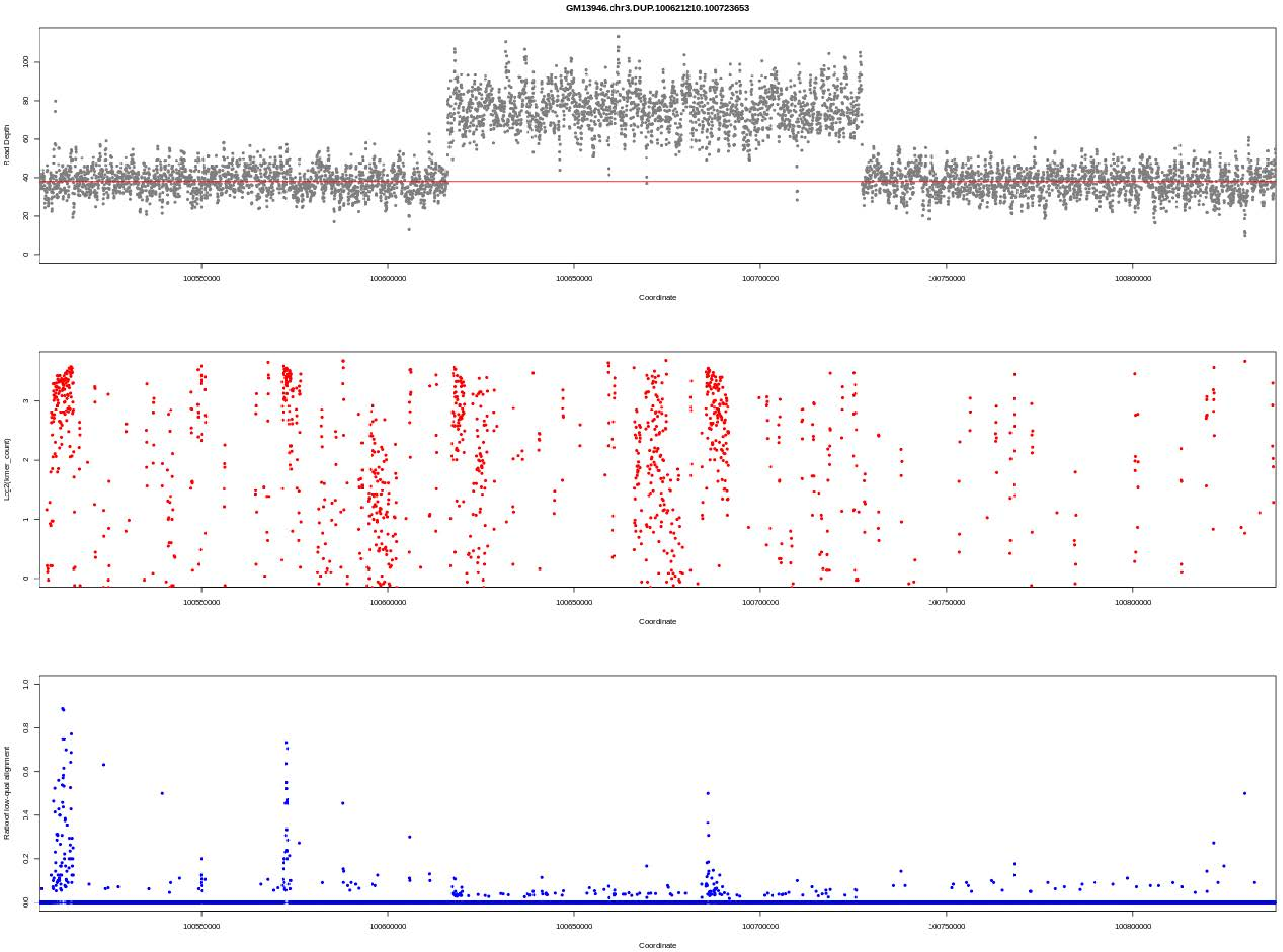

GM13946.chr3.DUP.100621210.100723653
Read Depth
Coordinate
Log2(kmer_count)
Coordinate
Ratio of low-qual alignment
Coordinate

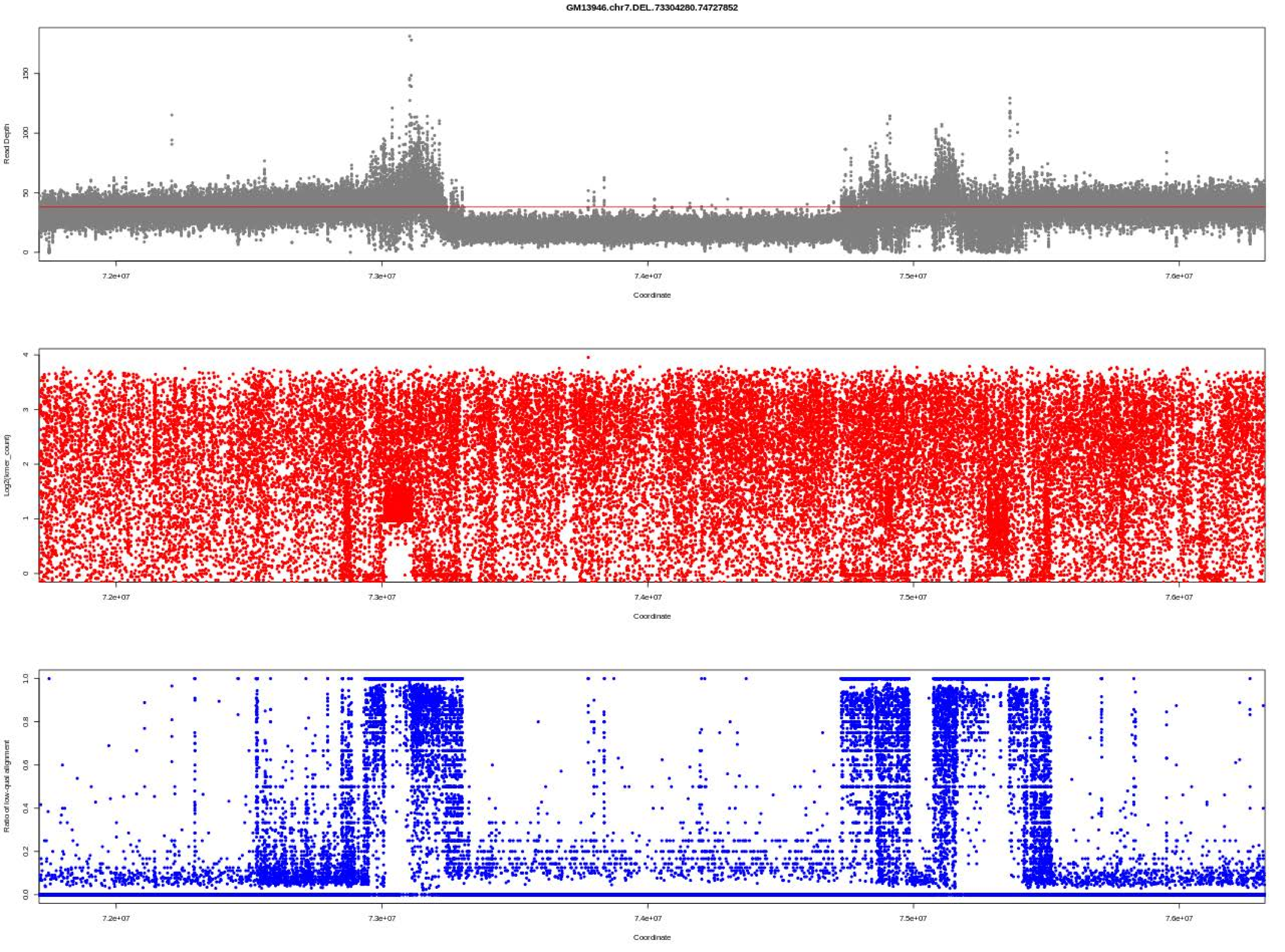

GM13946.chr7.DEL.73304280.74727852
Read Depth
Coordinate
Log2(kmer_count)
Coordinate
Ratio of low-qual alignment
Coordinate

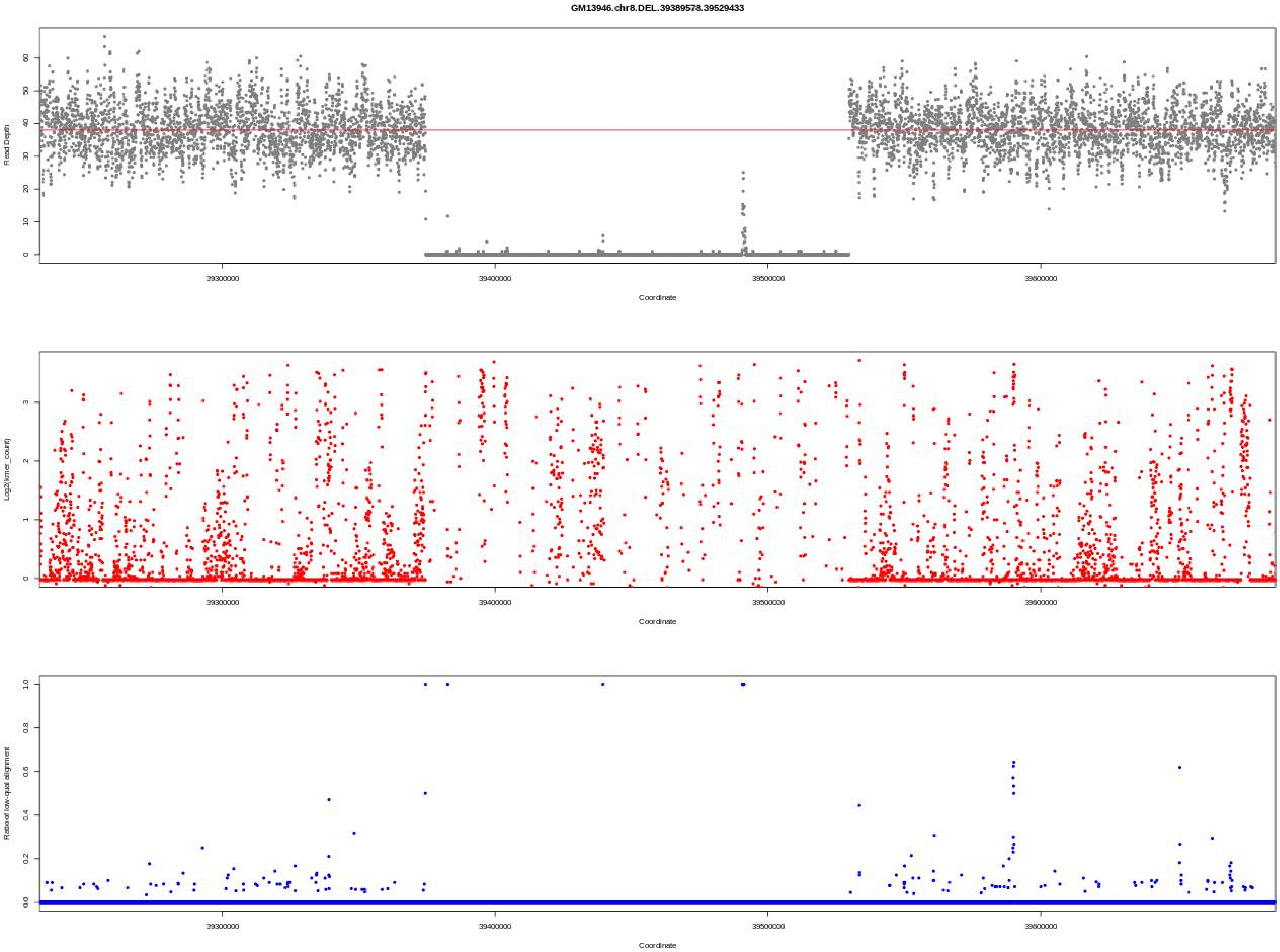

GM13946.chr8.DEL.39389578.39529433
Read Depth
Coordinate
Log2(kmer_count)
Coordinate
Ratio of low-qual alignment
Coordinate
39300000
39400000
39500000
39600000

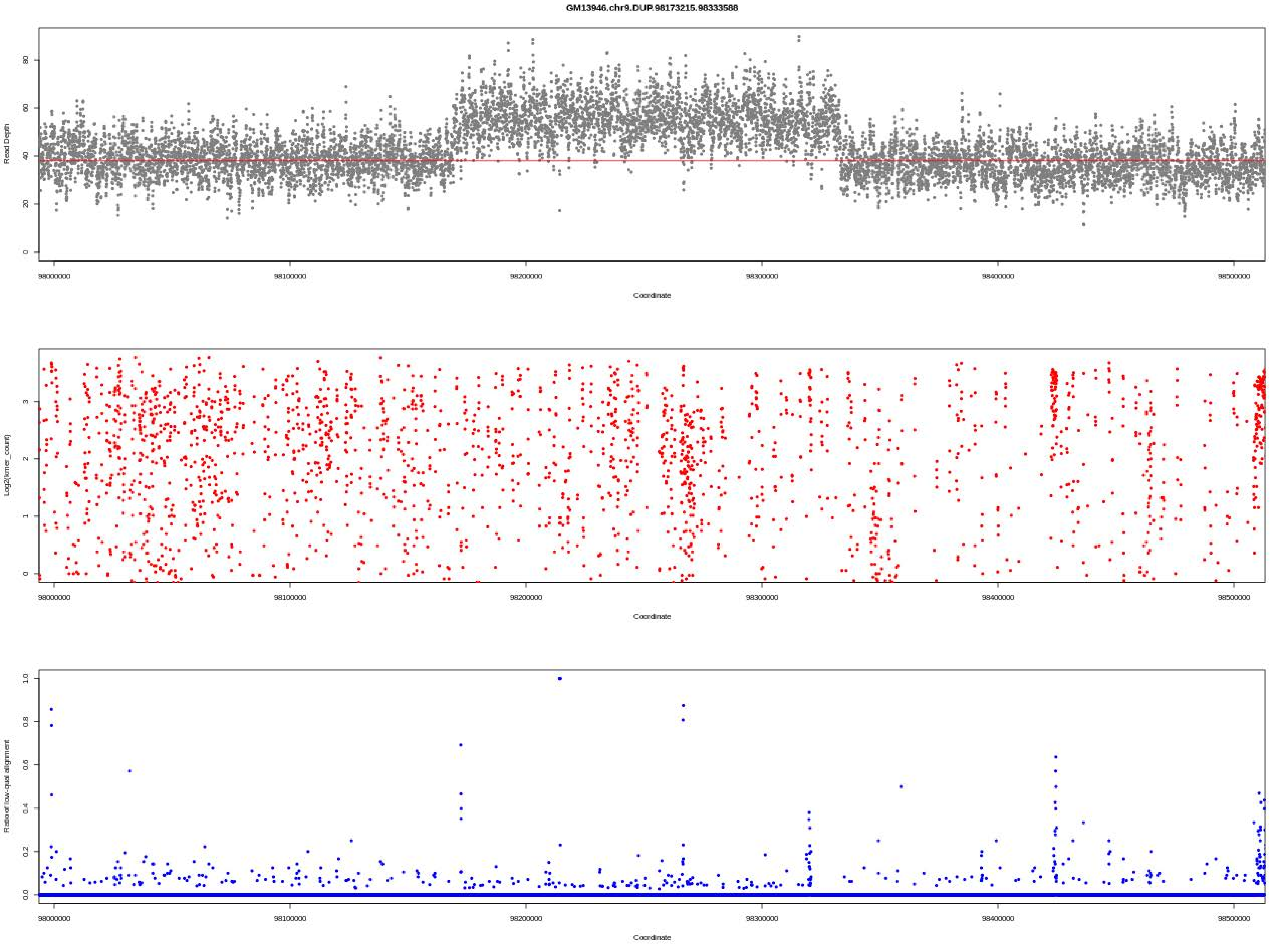
GM13946.chr9.DUP.98173215.98333588
Read Depth
Coordinate
Log2(kmer_count)
Coordinate
Ratio of low-qual alignment
Coordinate
98000000
98100000
98200000
98300000
98400000
98500000

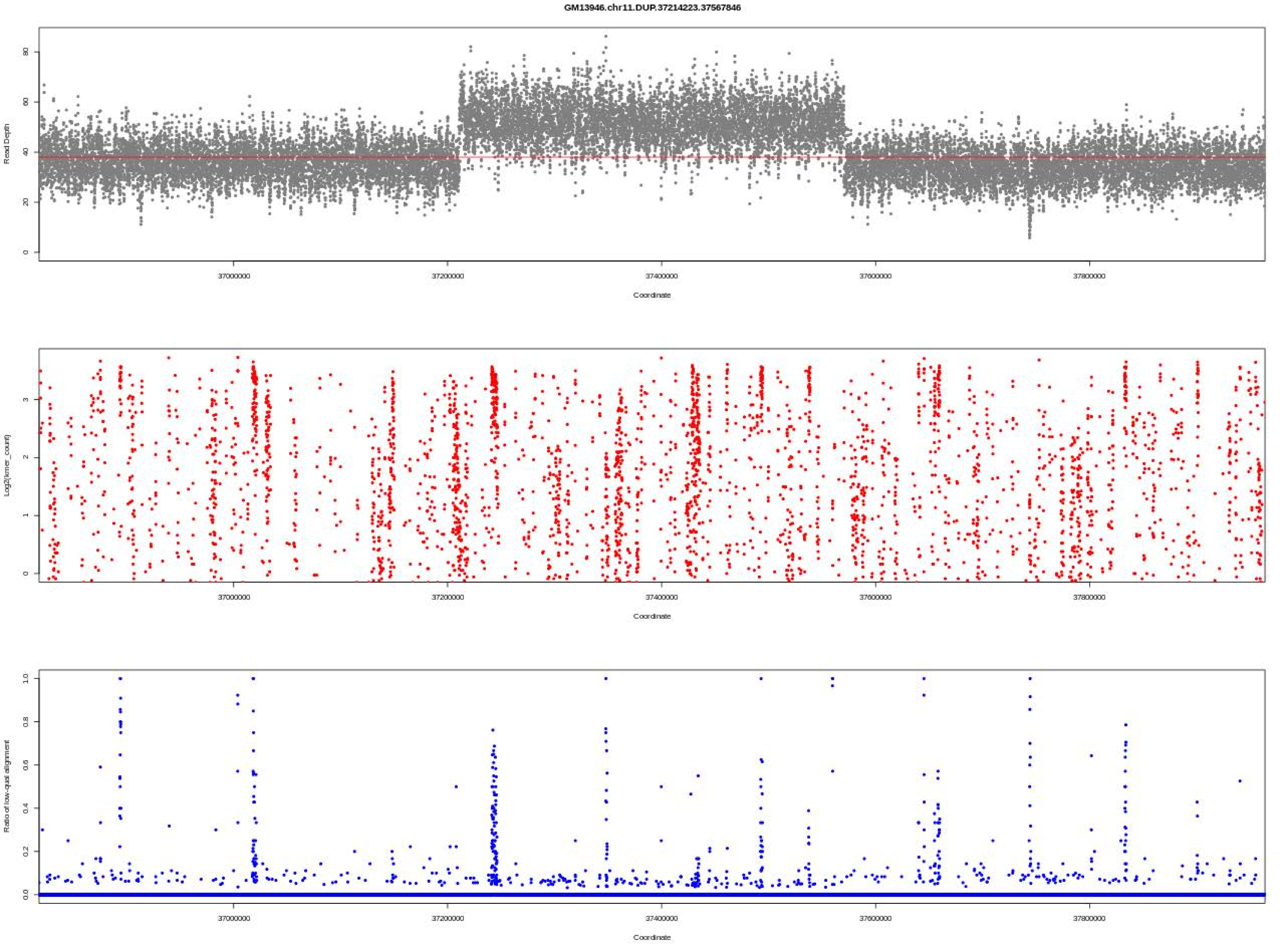
GM13946.chr11.DUP.37214223.37567846
Read Depth
Log2(kmer_count)
Ratio of low-qual alignment
Coordinate
37000000
37200000
37400000
37600000
37800000

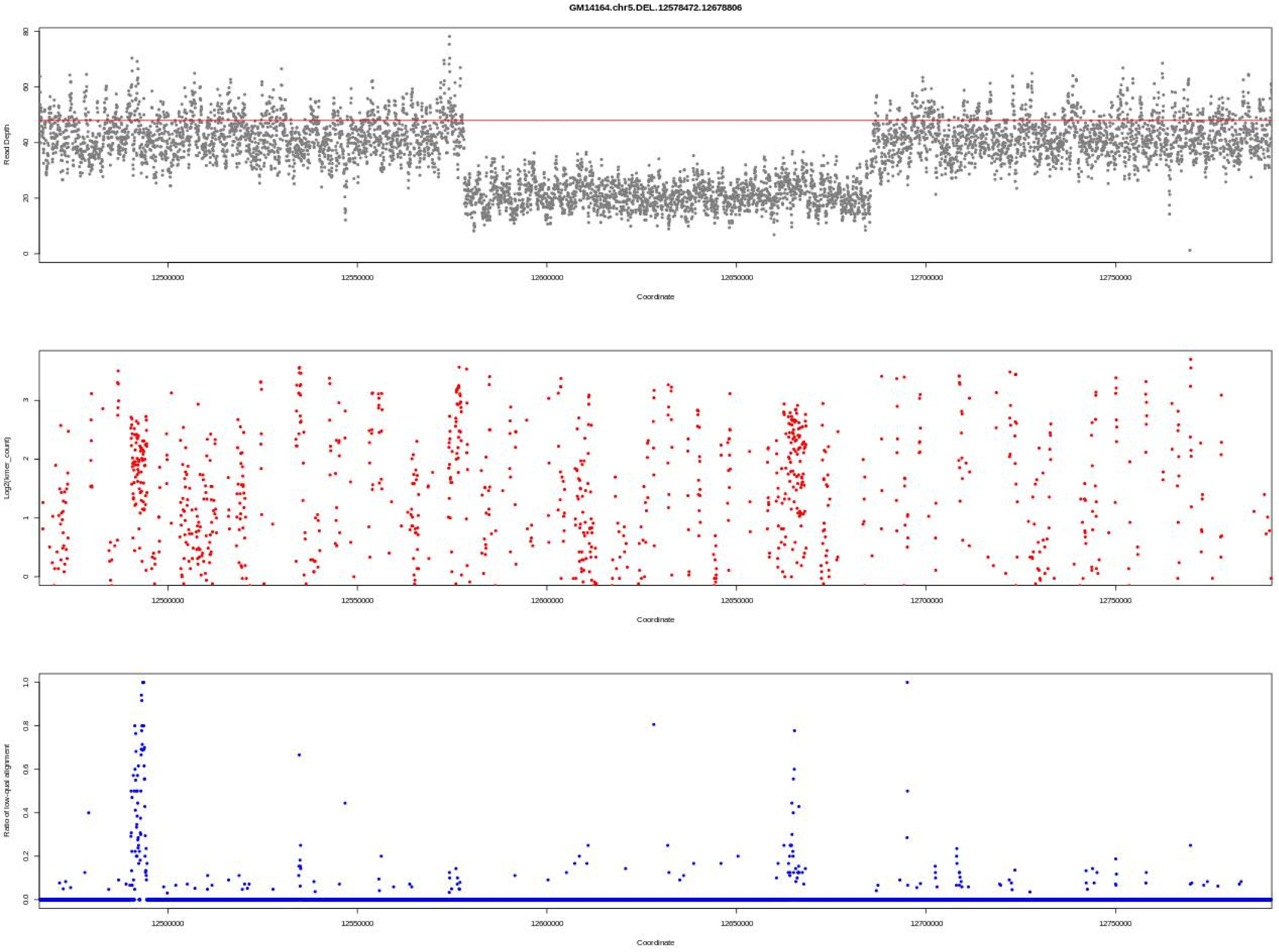
GM14164.chr5.DEL.12578472.12678806
Read Depth
Coordinate
Log2(kmer_count)
Coordinate
Ratio of low-qual alignment
Coordinate

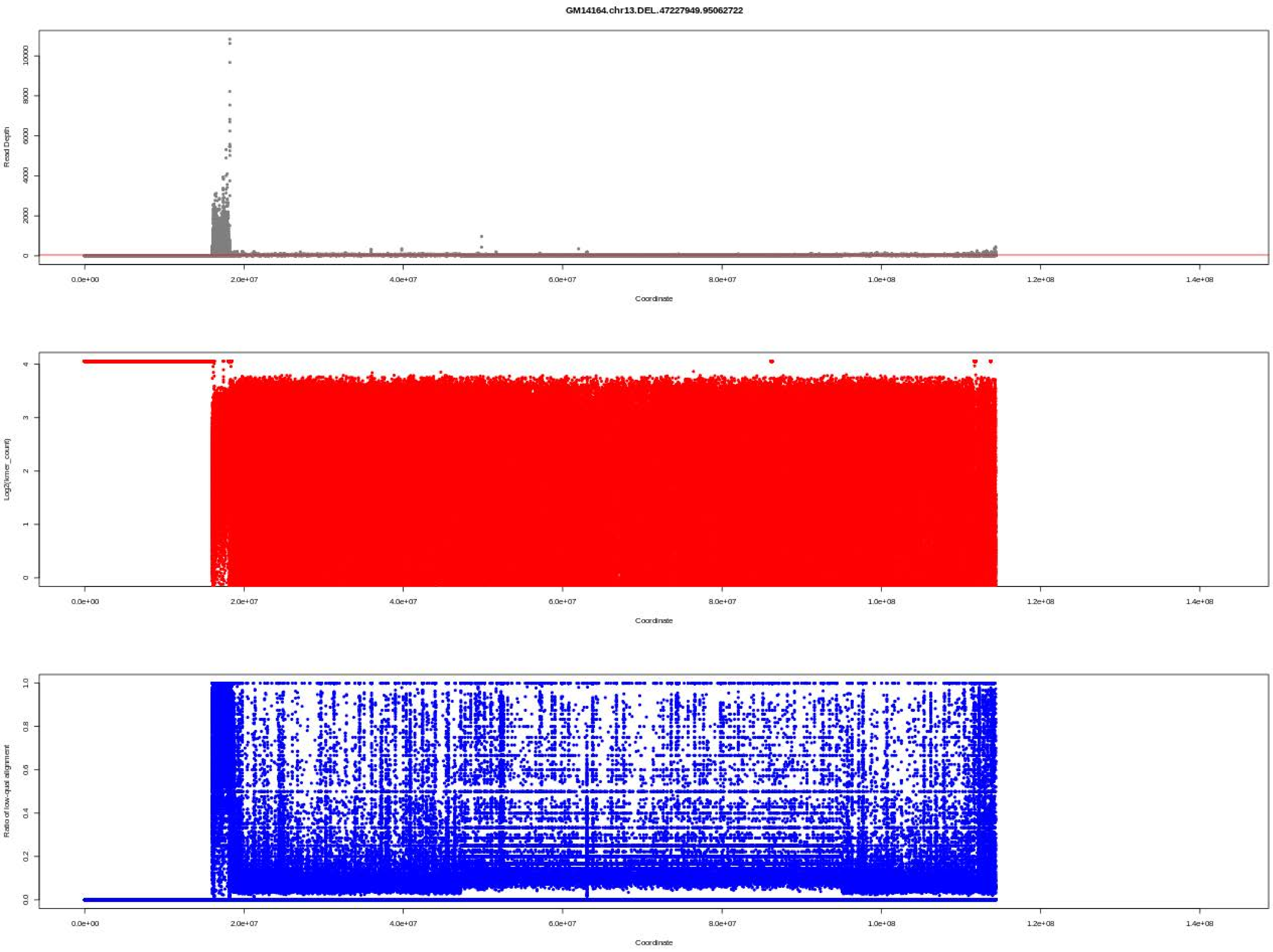

GM14164.chr13.DEL.47227949.95062722
Read Depth
Coordinate
Log2(kmer_count)
Coordinate
Ratio of low-qual alignment
Coordinate

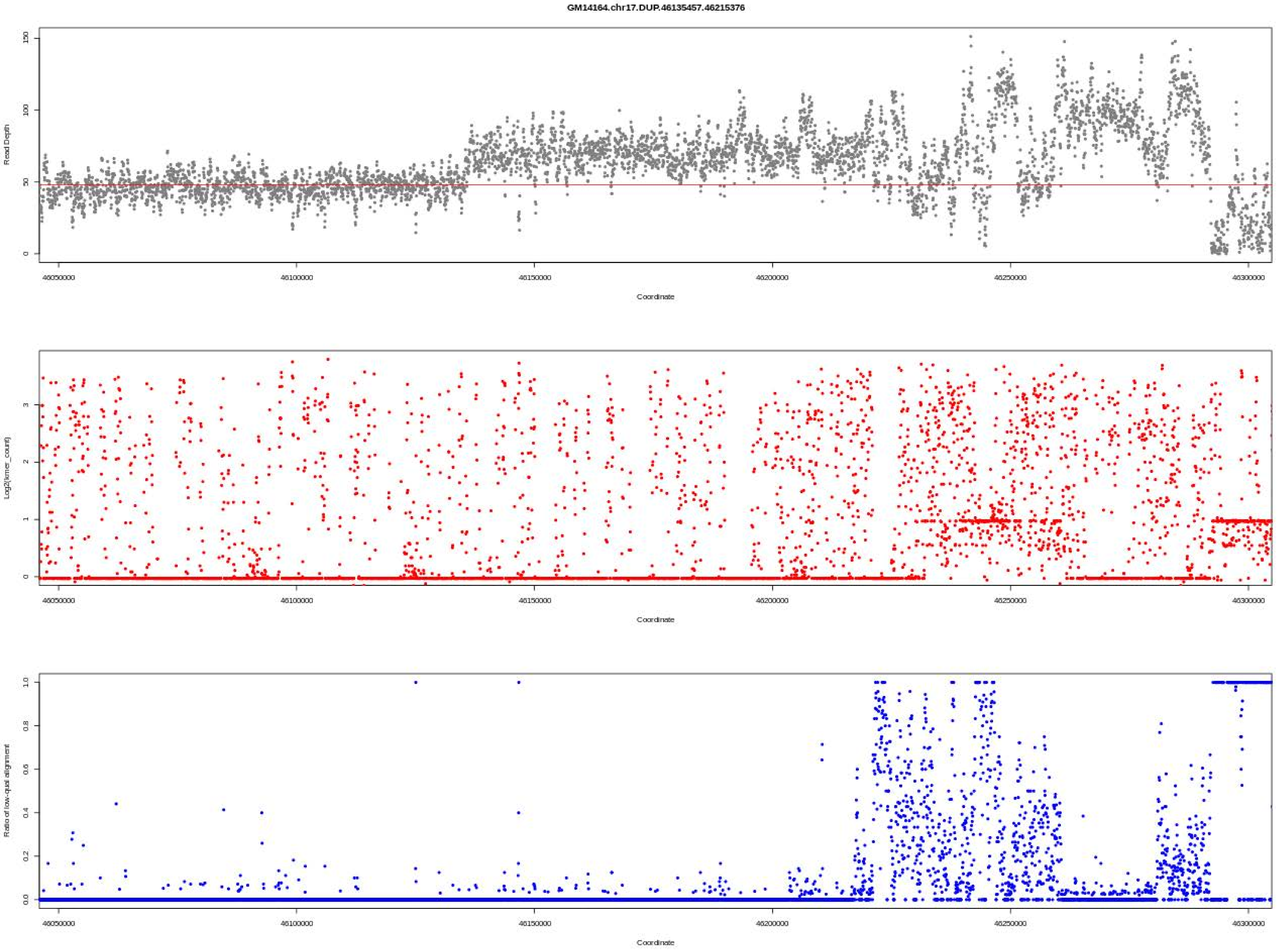

GM14164.chr17.DUP.46135457.46215376
Read Depth
Coordinate
Log2(kmer_count)
Coordinate
Ratio of low-qual alignment
Coordinate

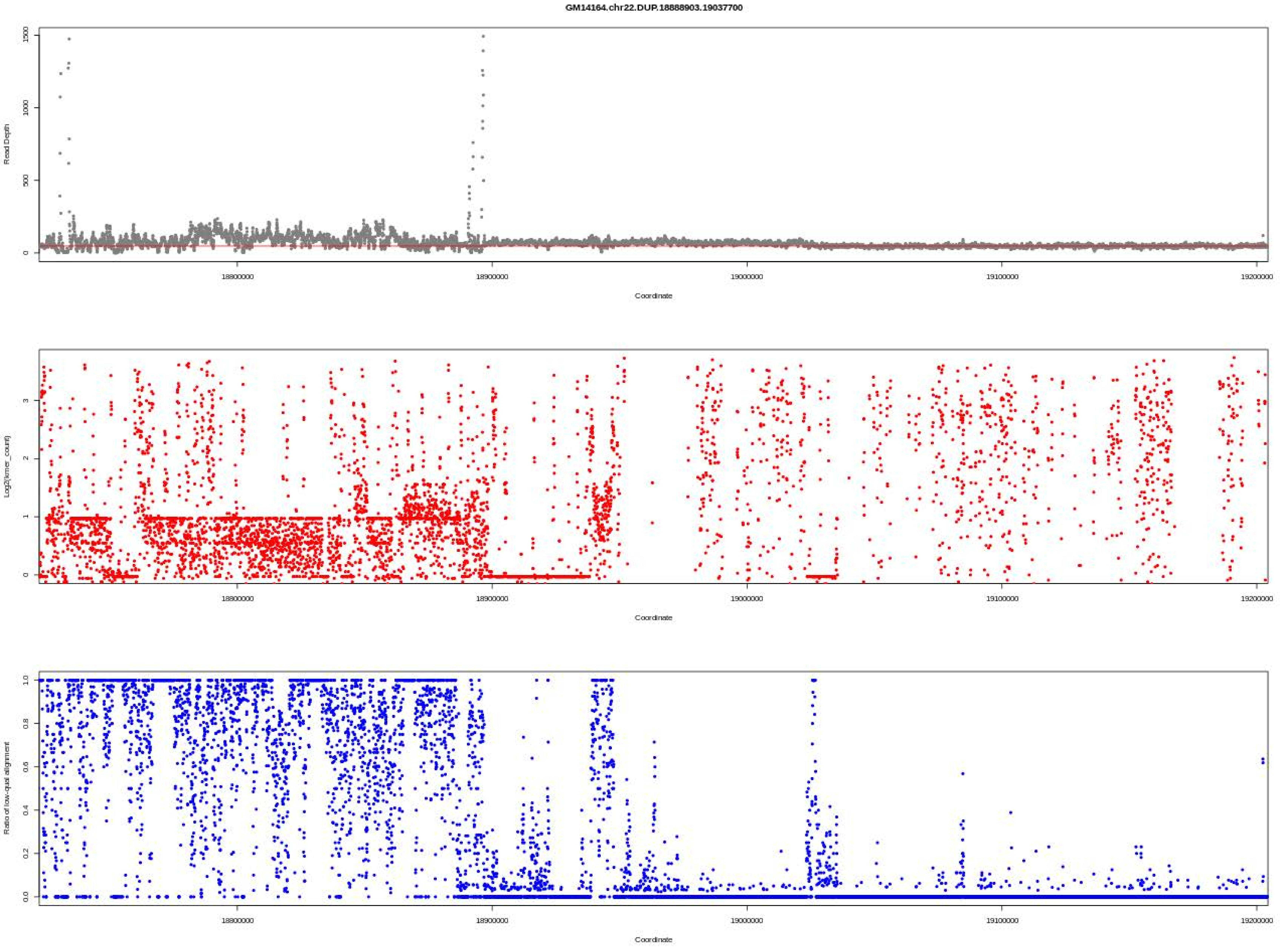
GM14164.chr22.DUP.18888903.19037700
Read Depth
0
500
1000
1500
18800000
18900000
19000000
19100000
19200000
Coordinate
Log2(kmer_count)
0
1
2
3
Coordinate
Ratio of low-qual alignment
0.0
0.2
0.4
0.6
0.8
1.0
Coordinate

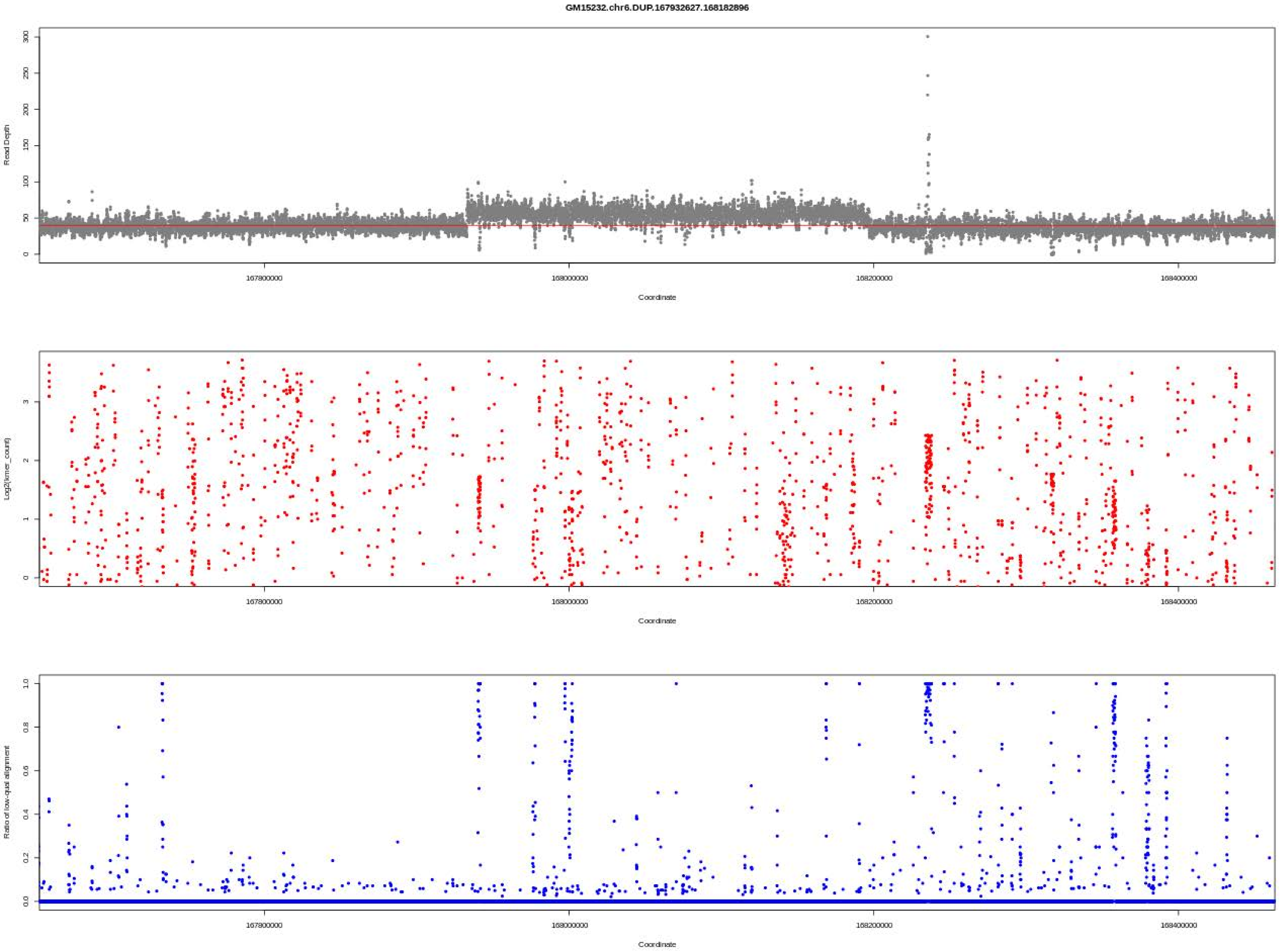

GM15232.chr6.DUP.167932627.168182896
Read Depth
Coordinate
Log2(kmer_count)
Coordinate
Ratio of low-qual alignment
Coordinate

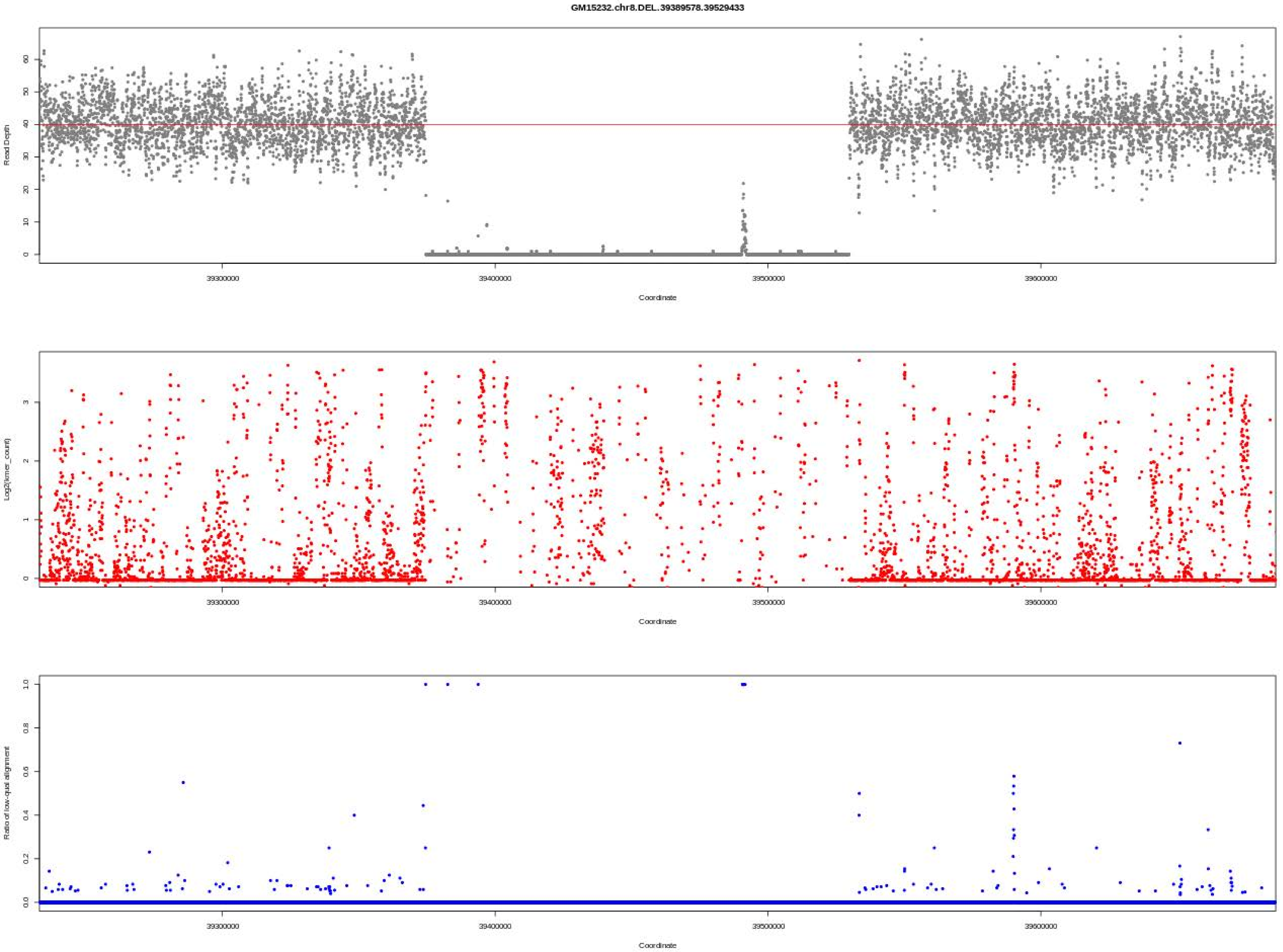
GM15232.chr8.DEL.39389578.39529433
Read Depth
Coordinate
Log2(kmer_count)
Coordinate
Ratio of low-qual alignment
Coordinate

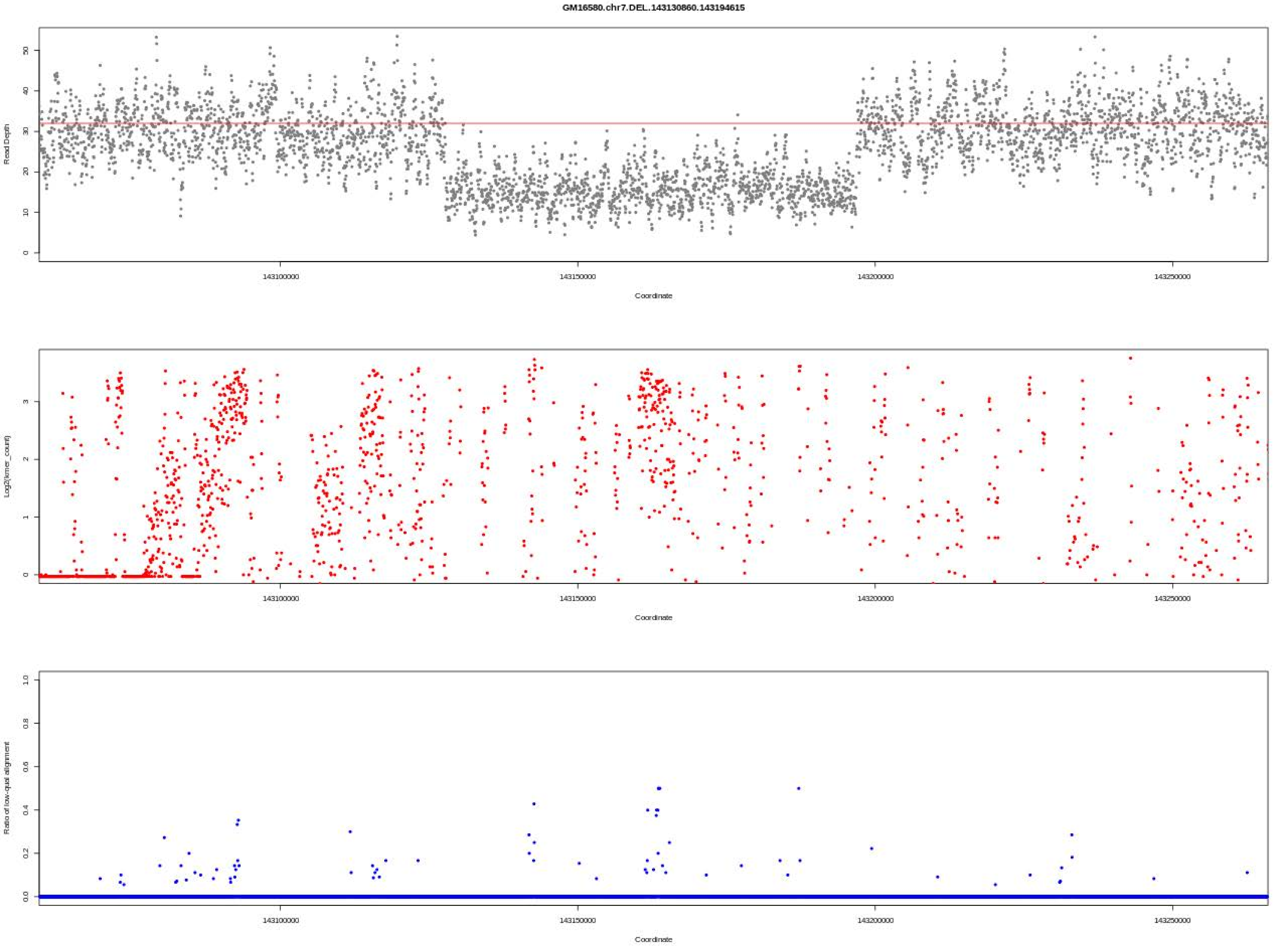

GM16580.chr7.DEL.143130860.143194615
Read Depth
Coordinate
Log2(kmer_count)
Coordinate
Ratio of low-qual alignment
Coordinate
143100000
143150000
143200000
143250000

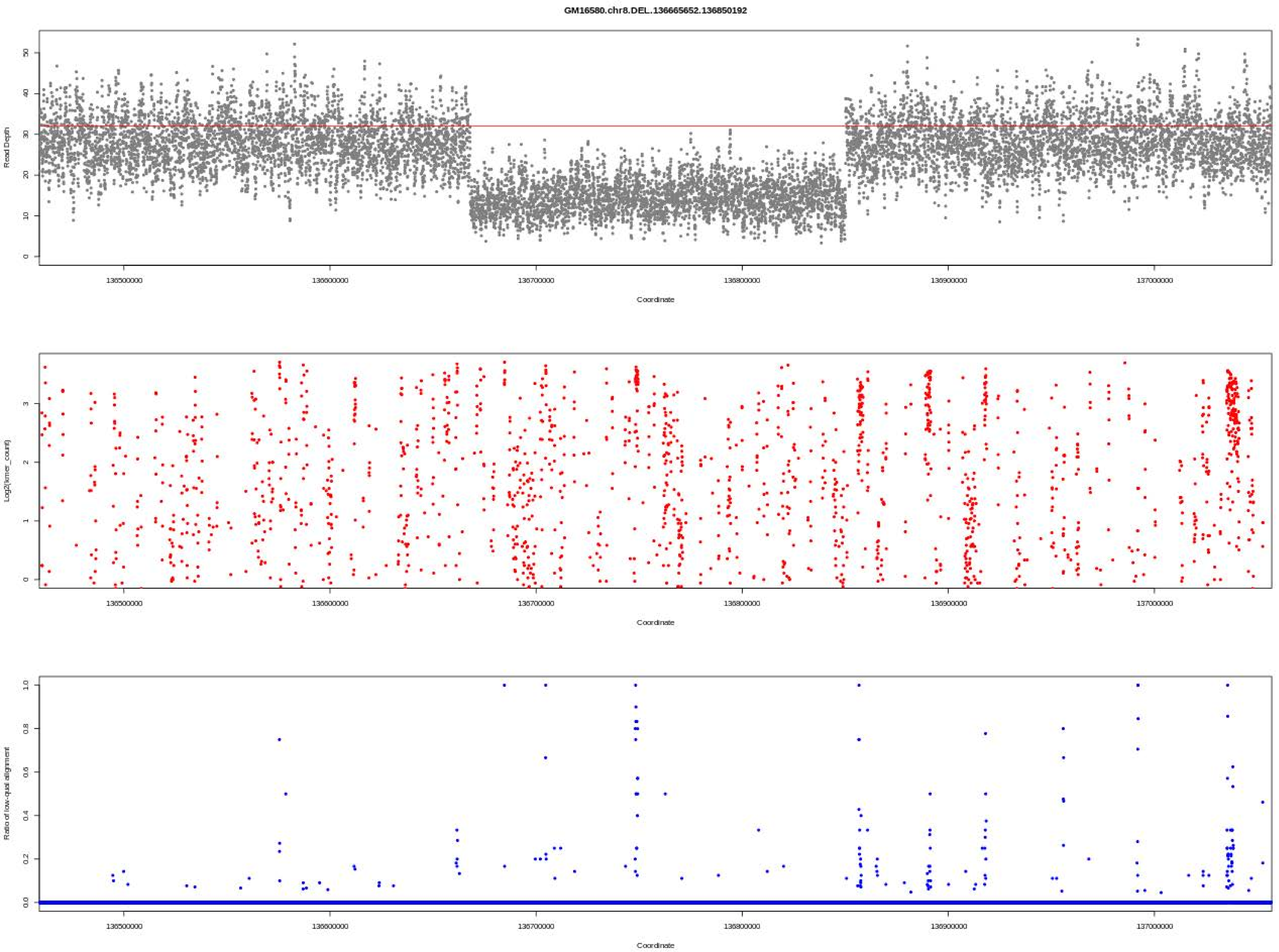
GM16580.chr8.DEL..136665652.136850192
Read Depth
Coordinate
Log2(kmer_count)
Coordinate
Ratio of low-qual alignment
Coordinate

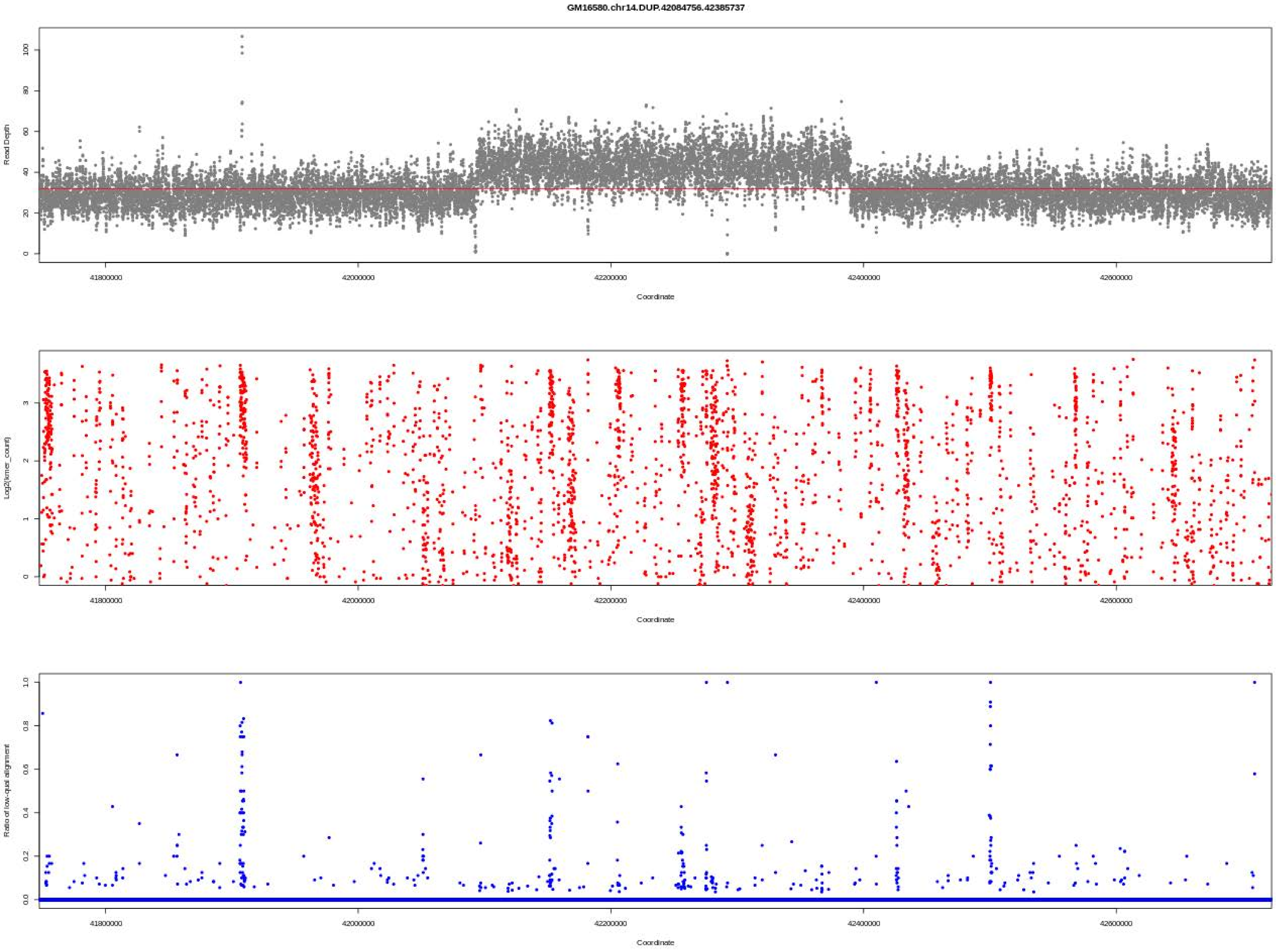
GM16580.chr14.DUP.42084756.42385737
Read Depth
Coordinate
Log2(kmer_count)
Coordinate
Ratio of low-qual alignment
Coordinate

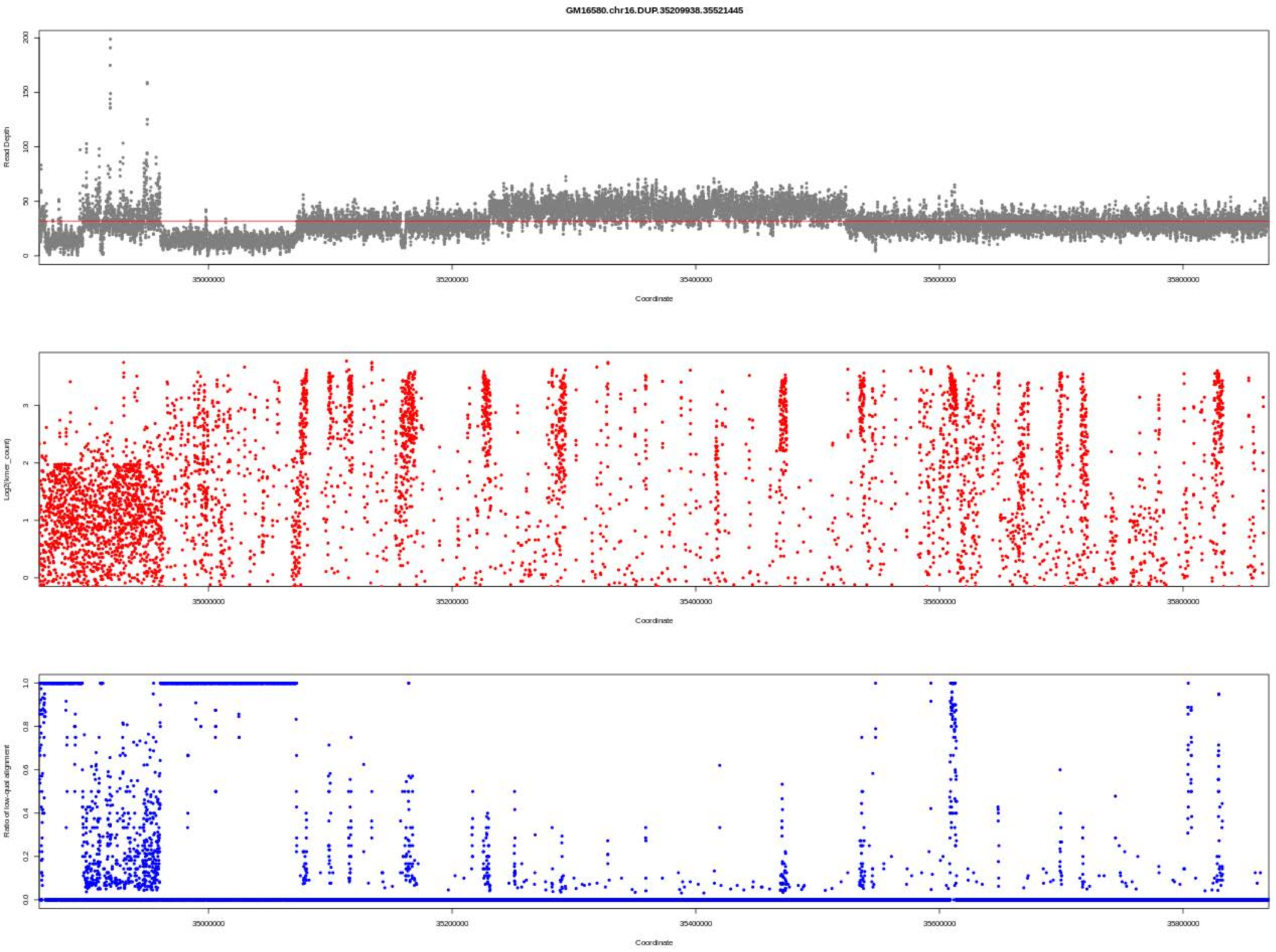

GM16580.chr16.DUP.35209938.35521445
Read Depth
Coordinate
Log2(kmer_count)
Coordinate
Ratio of low-qual alignment
Coordinate

GM16580.chr18.DEL.11542.1636725

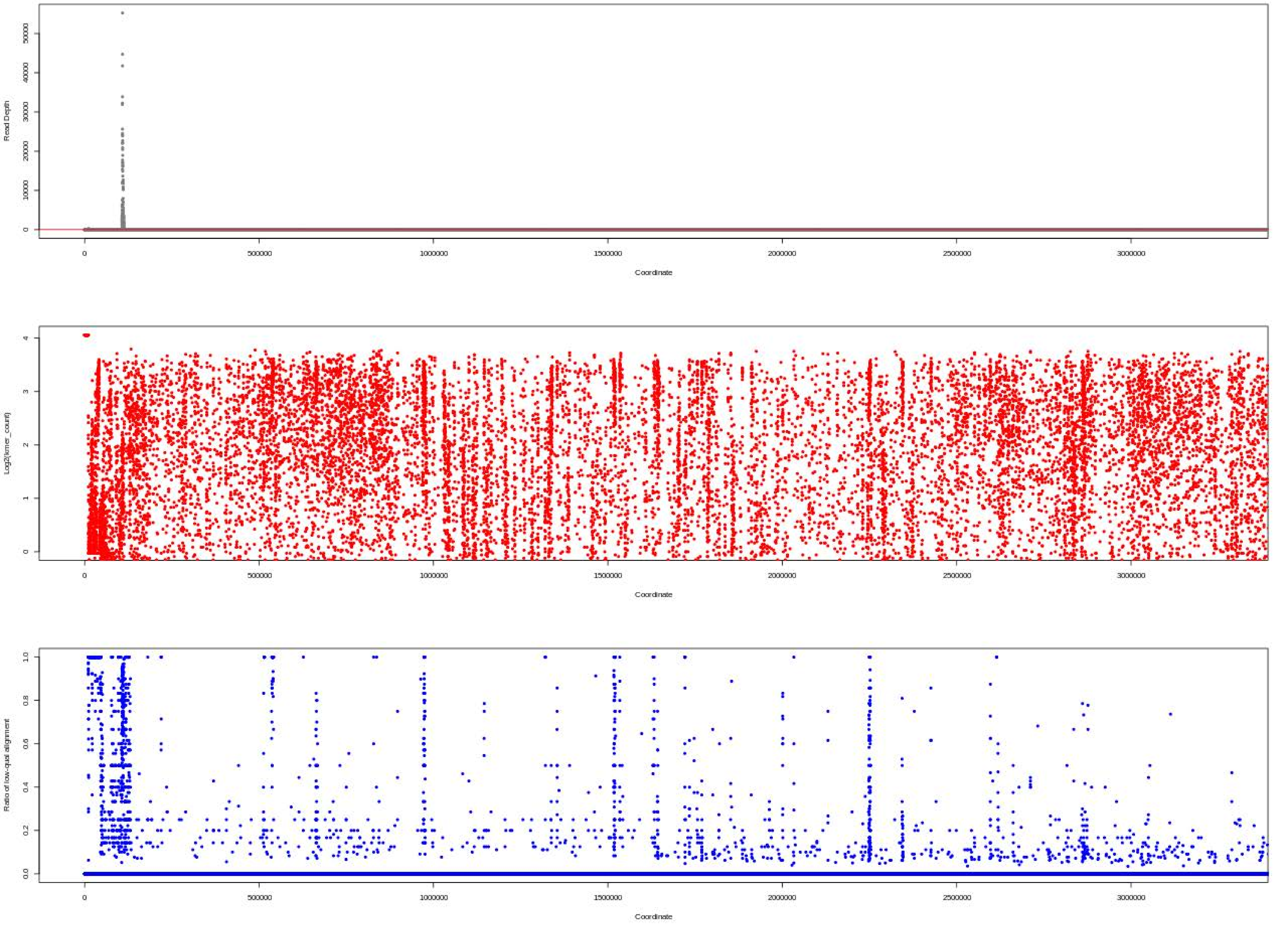

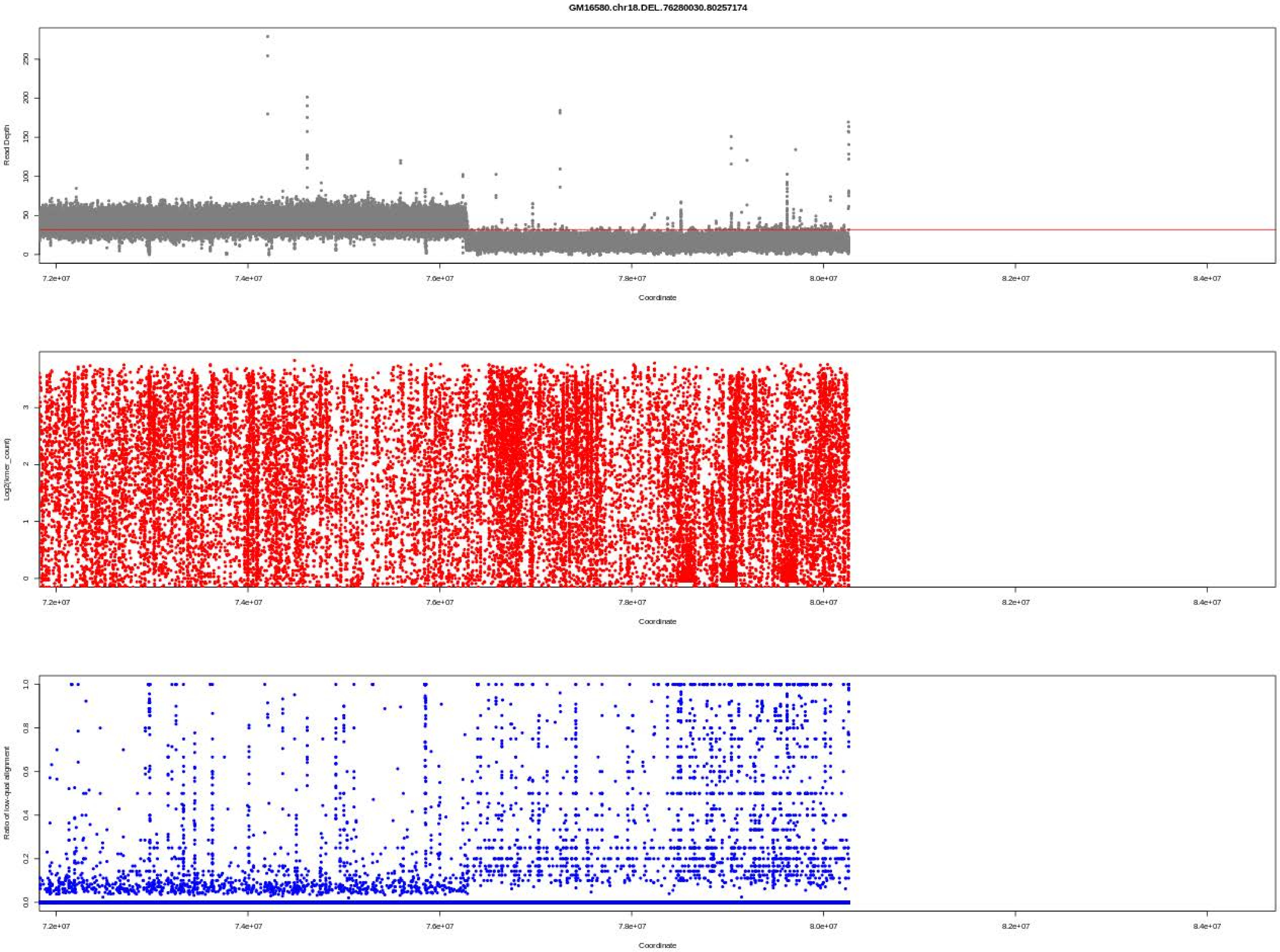

GM16580.chr18.DEL.76280030.80257174
Read Depth
0
50
100
150
200
250
7.2e+07
7.4e+07
7.6e+07
7.8e+07
8.0e+07
8.2e+07
8.4e+07
Coordinate
Log2(kmer_count)
0
1
2
3
Coordinate
Ratio of low-qual alignment
0.0
0.2
0.4
0.6
0.8
1.0
Coordinate

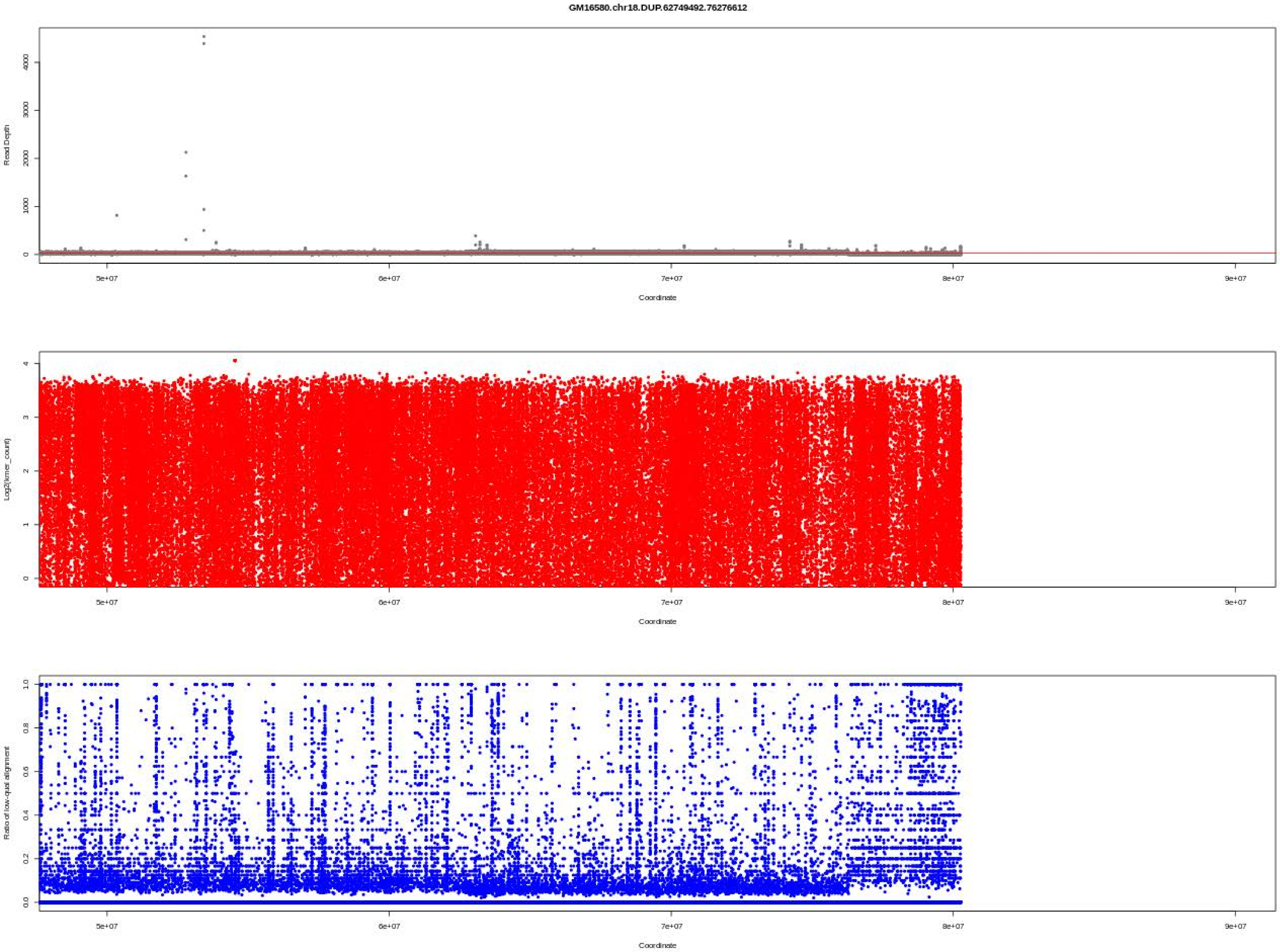

GM16580.chr18.DUP.62749492.76276612
Read Depth
0
1000
2000
3000
4000
5e+07
6e+07
7e+07
8e+07
9e+07
Coordinate
Log2(kmer_count)
0
1
2
3
4
5e+07
6e+07
7e+07
8e+07
9e+07
Coordinate
Ratio of low-qual alignment
0.0
0.2
0.4
0.6
0.8
1.0
5e+07
6e+07
7e+07
8e+07
9e+07
Coordinate

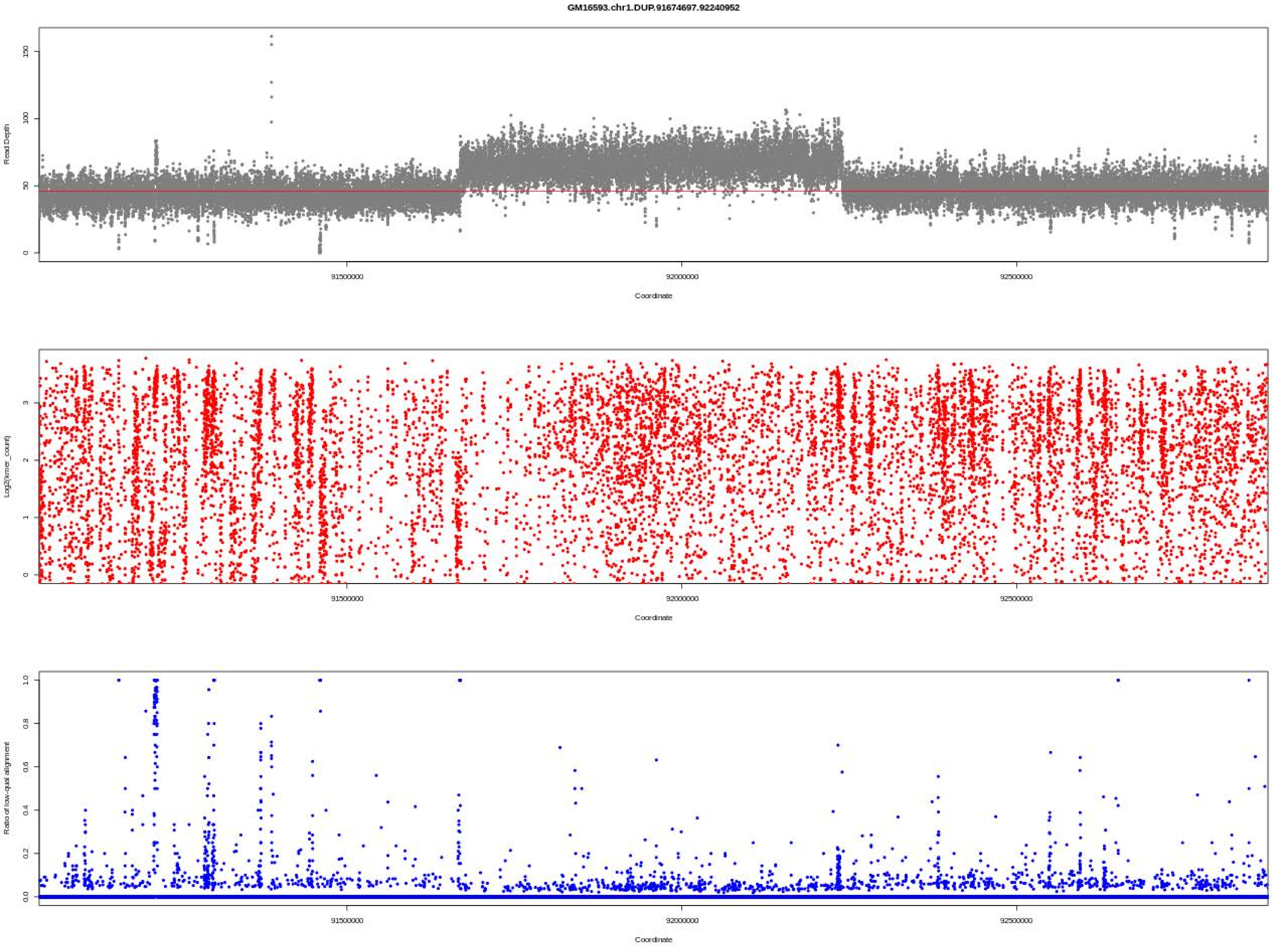
GM16593.chr1.DUP.91674697.92240952
Read Depth
Coordinate
Log2(kmer_count)
Coordinate
Ratio of low-qual alignment
Coordinate

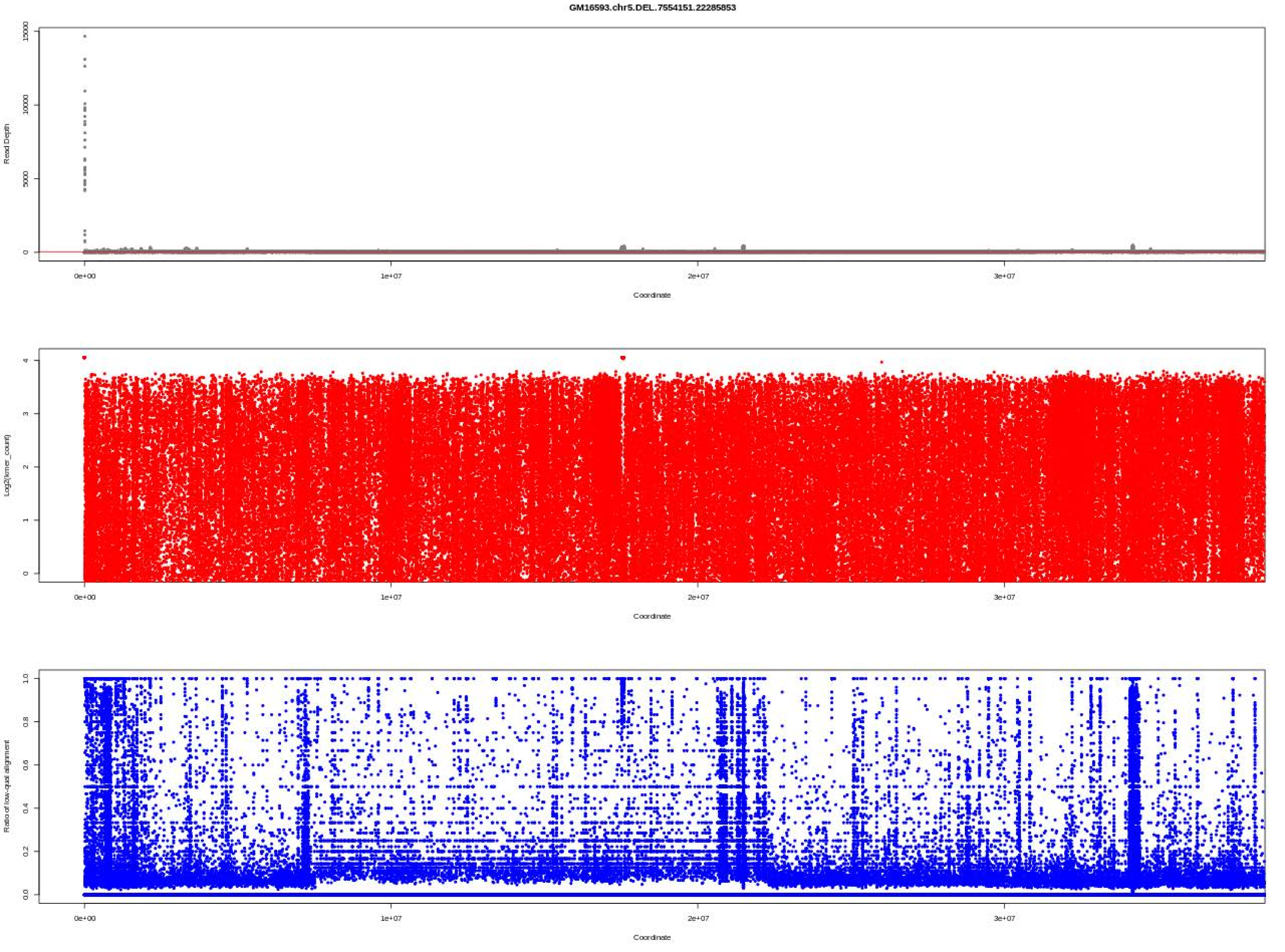
GM16593.chr5.DEL.7554151.22285853
Read Depth
Coordinate
Log2(kmer_count)
Coordinate
Ratio of low-qual alignment
Coordinate

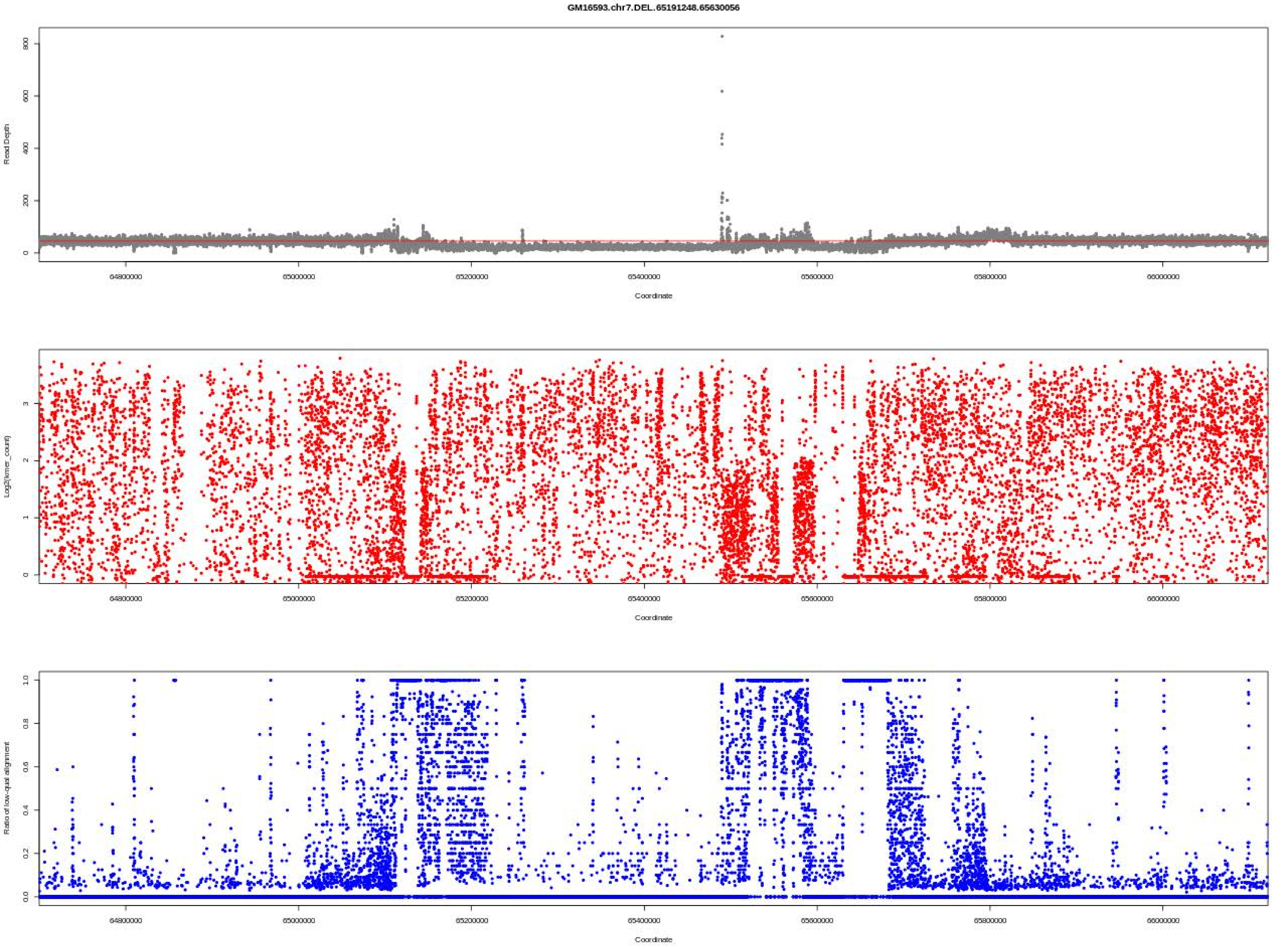
GM16593.chr7.DEL.65191248.65630056
Read Depth
Coordinate
Log2(kmer_count)
Coordinate
Ratio of low-qual alignment
Coordinate

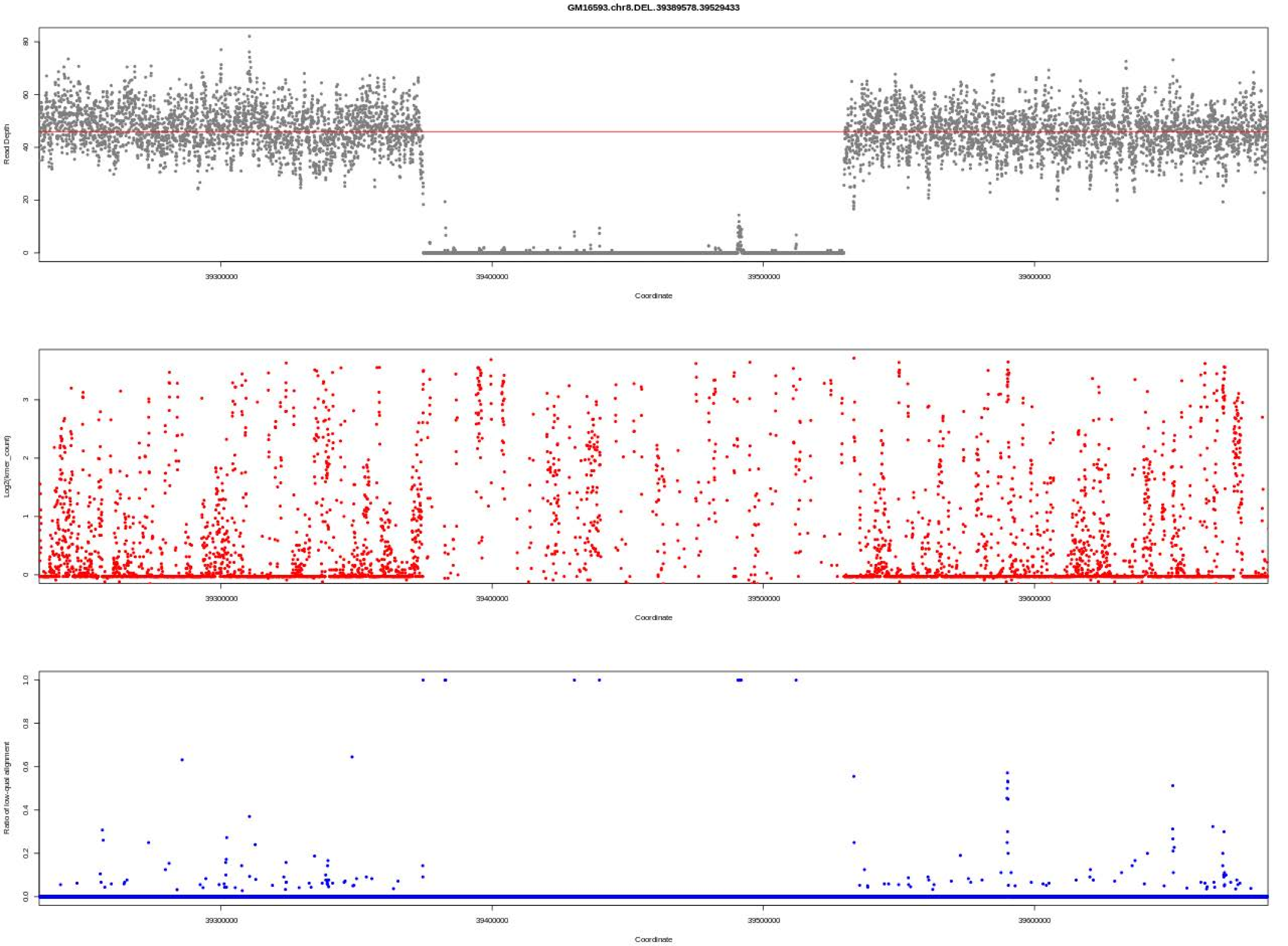
GM16593.chr8.DEL.39389578.39529433
Read Depth
Coordinate
Log2(kmer_count)
Coordinate
Ratio of low-qual alignment
Coordinate
39300000
39400000
39500000
39600000

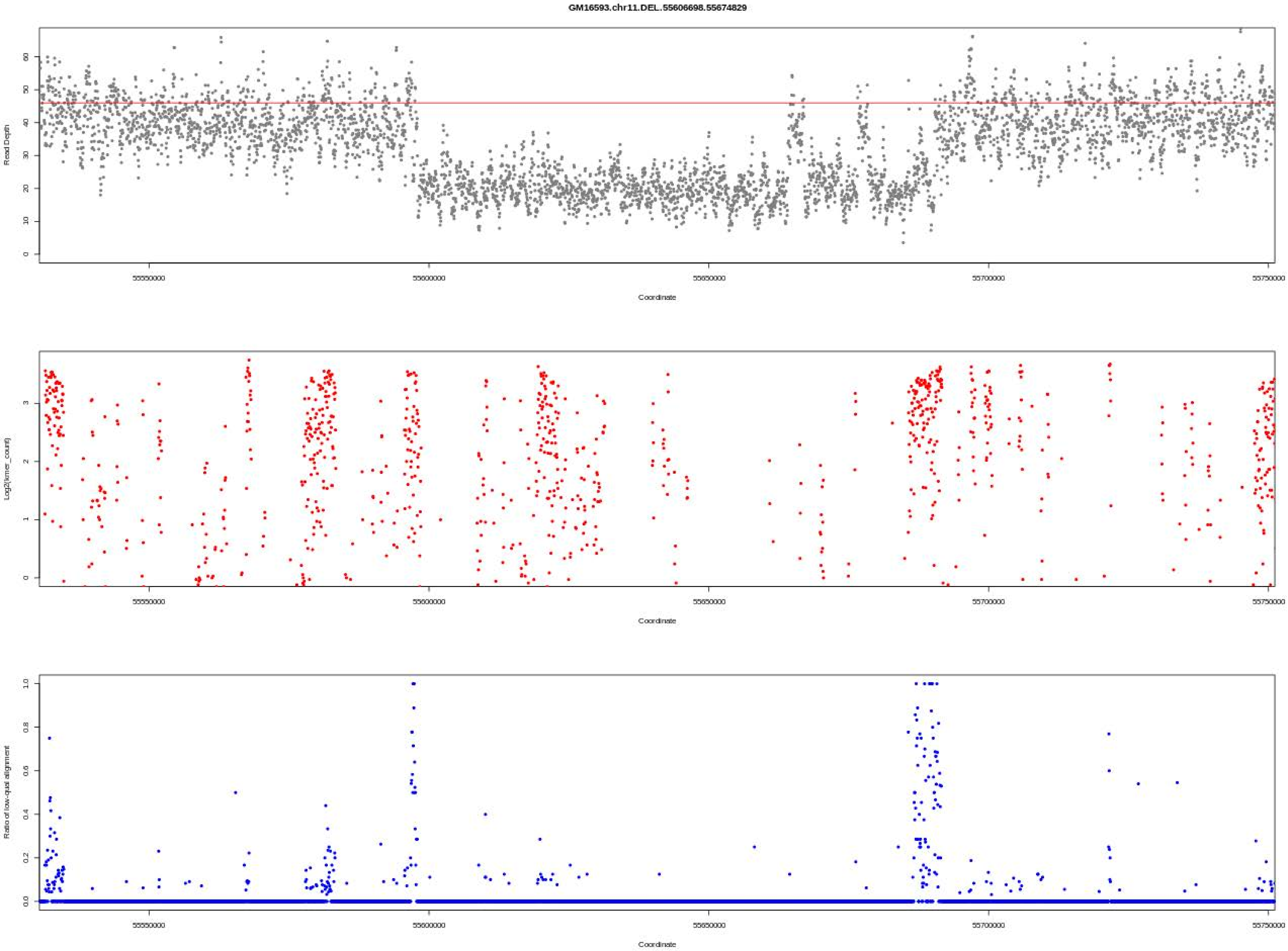

GM16593.chr11.DEL.55606698.55674829
Read Depth
Log2(kmer_count)
Ratio of low-qual alignment
Coordinate
55550000
55600000
55650000
55700000
55750000

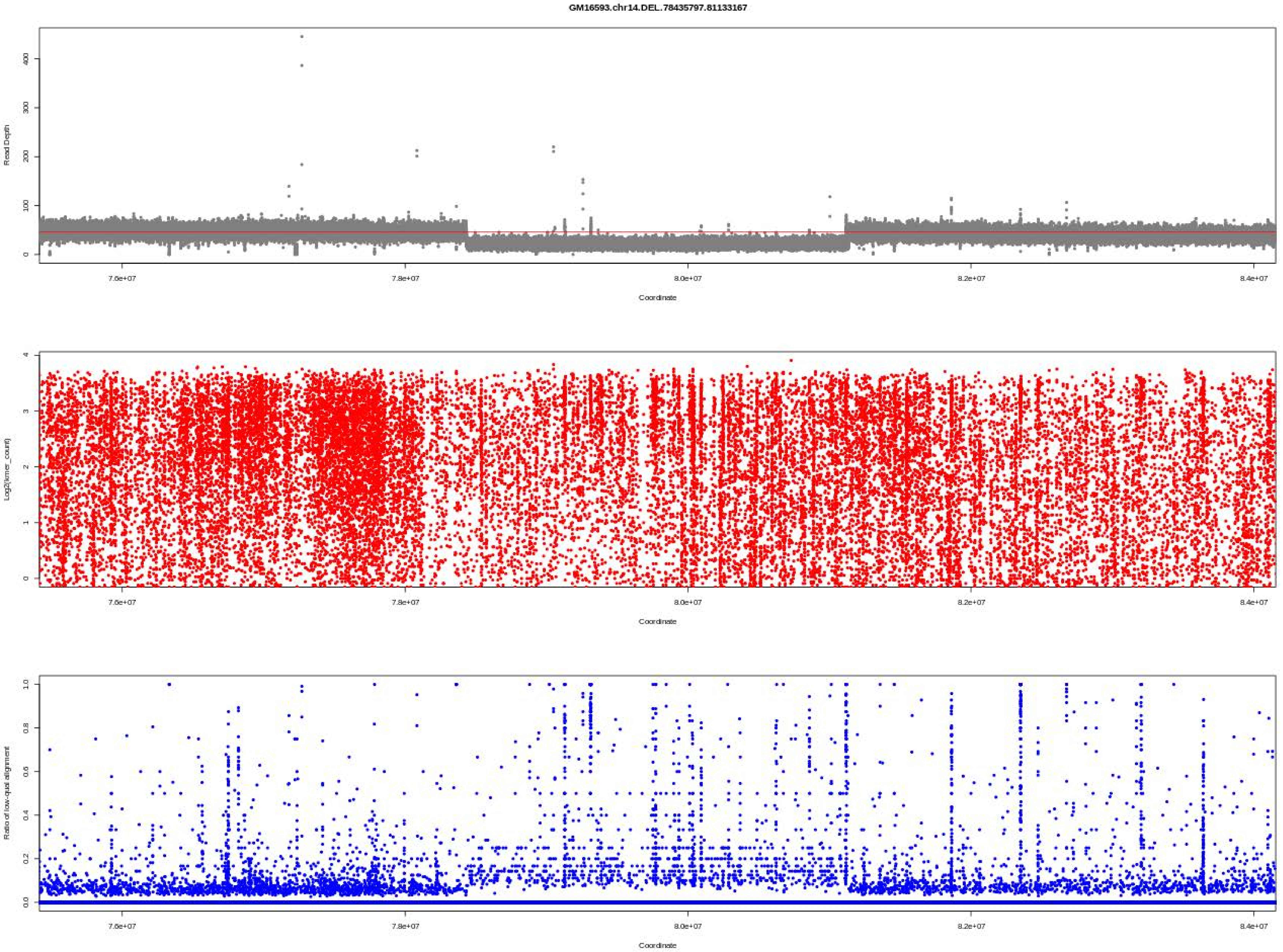
GM16593.chr14.DEL..78435797.81133167
Read Depth
Coordinate
Log2(kmer_count)
Coordinate
Ratio of low-qual alignment
Coordinate
7.6e+07
7.8e+07
8.0e+07
8.2e+07
8.4e+07

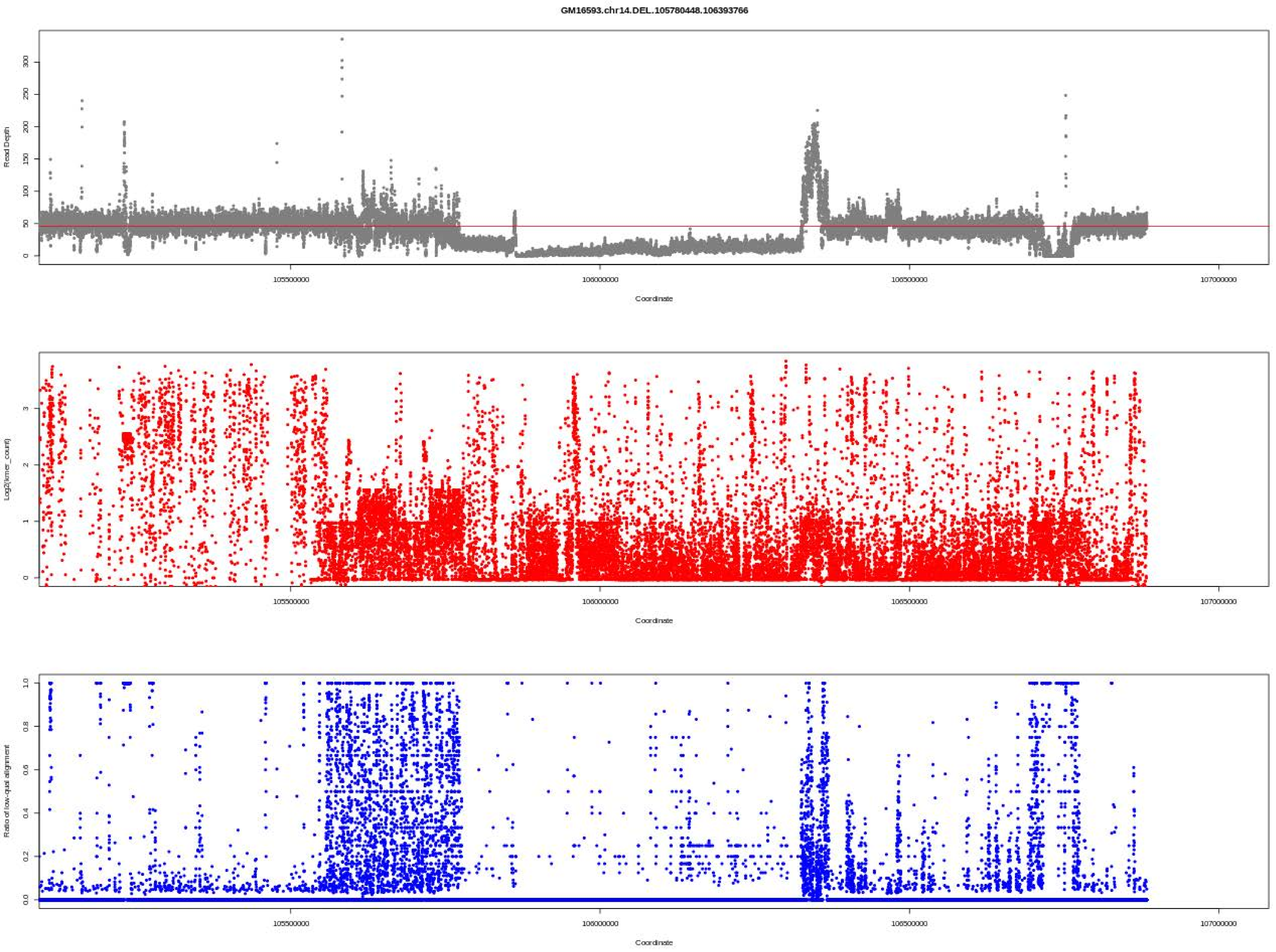
GM16593.chr14.DEL.105780448.106393766
Read Depth
Coordinate
Log2(kmer_count)
Coordinate
Ratio of low-qual alignment
Coordinate
105500000
106000000
106500000
107000000

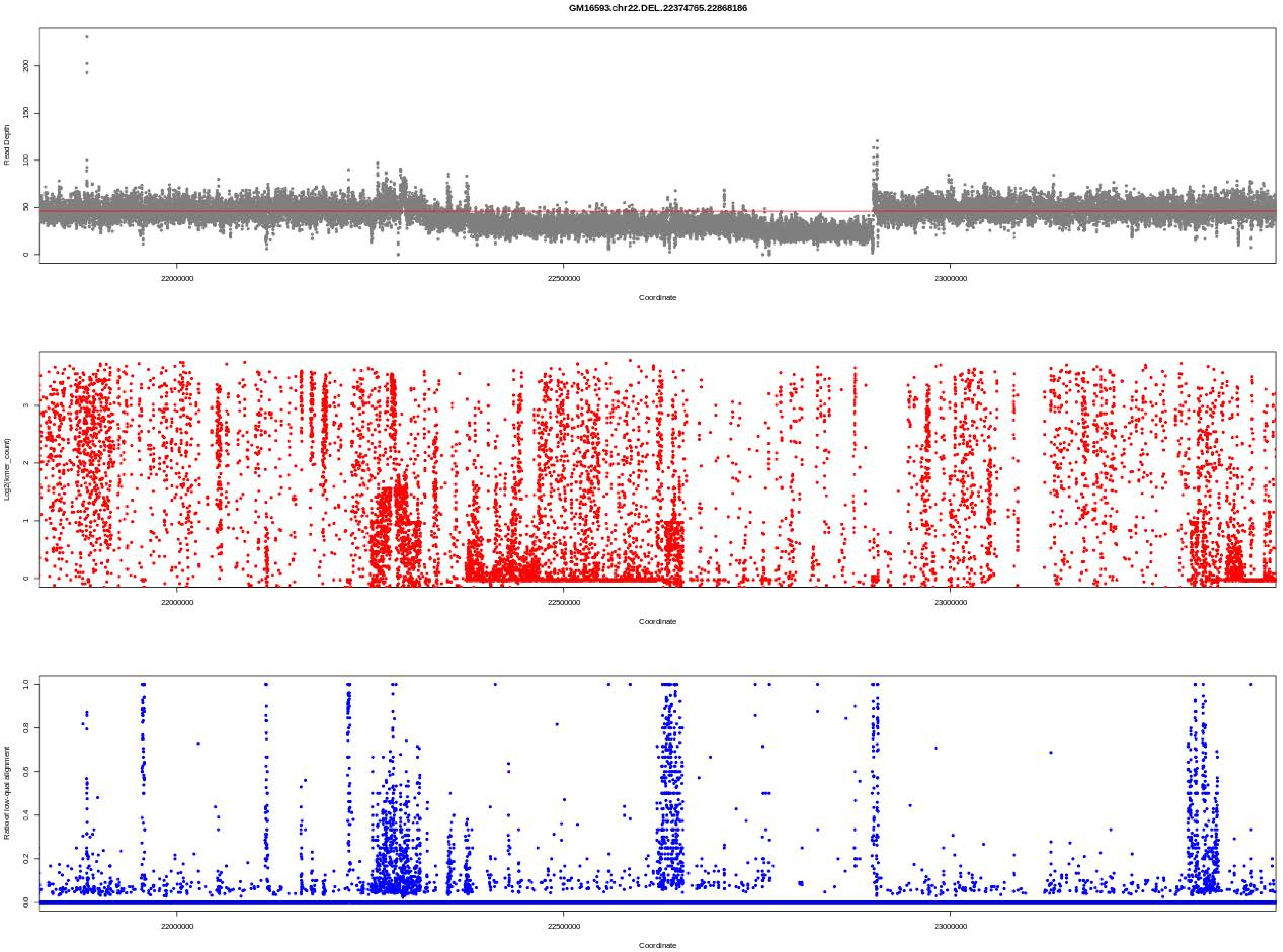

GM16593.chr22.DEL.22374765.22868186
Read Depth
0
50
100
150
200
22000000
22500000
23000000
Coordinate
Log2(kmer_count)
0
1
2
3
22000000
22500000
23000000
Coordinate
Ratio of low-qual alignment
0.0
0.2
0.4
0.6
0.8
1.0
22000000
22500000
23000000
Coordinate

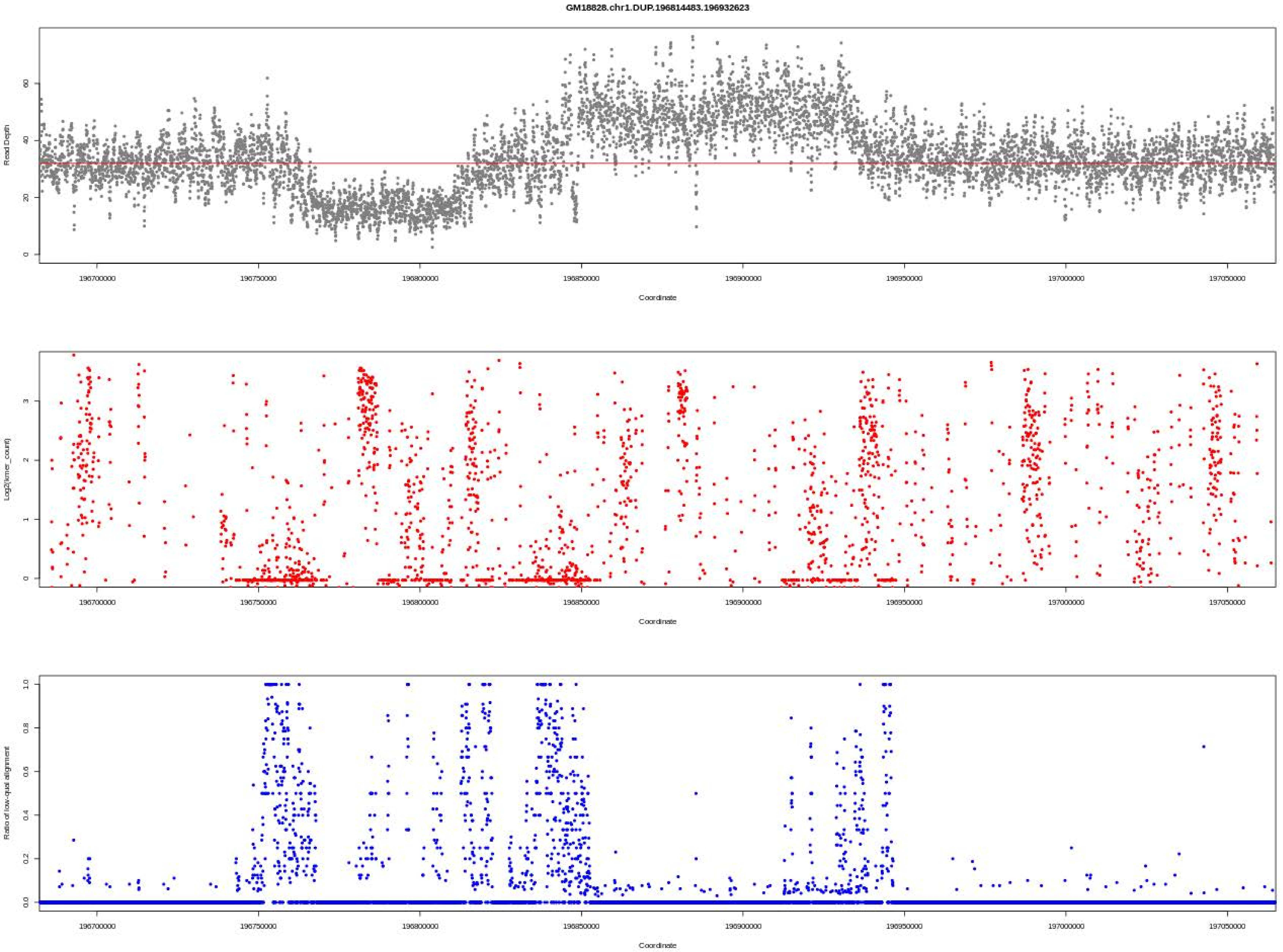

GM18828.chr1.DUP.196814483.196932623
Read Depth
Log2(kmer_count)
Ratio of low-qual alignment
Coordinate
196700000
196750000
196800000
196850000
196900000
196950000
197000000
197050000

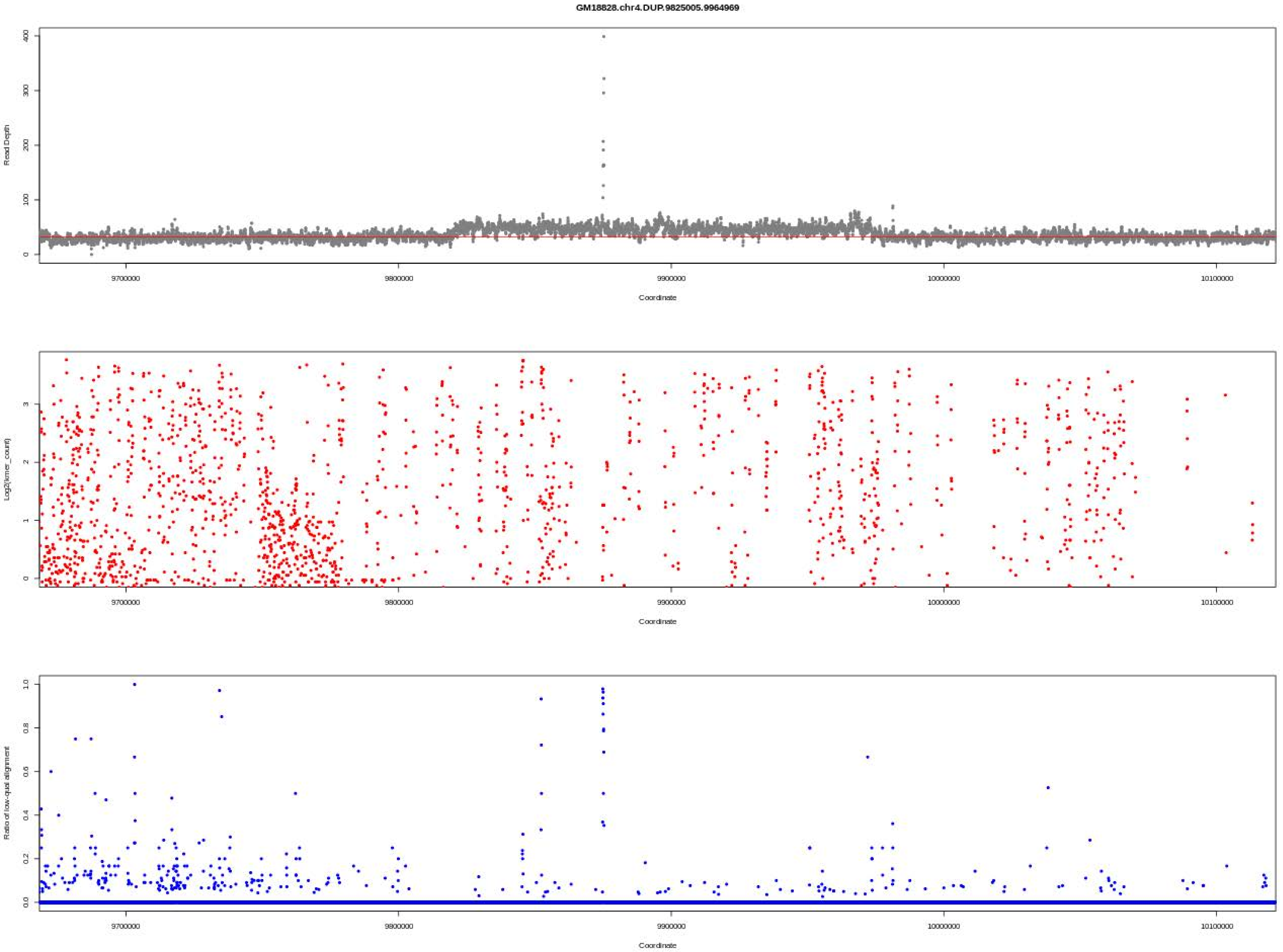

GM18828.chr4.DUP.9825005.9964969
Read Depth
0
100
200
300
400
9700000
9800000
9900000
10000000
10100000
Coordinate
Log2(kmer_count)
0
1
2
3
9700000
9800000
9900000
10000000
10100000
Coordinate
Ratio of low-qual alignment
0.0
0.2
0.4
0.6
0.8
1.0
9700000
9800000
9900000
10000000
10100000
Coordinate

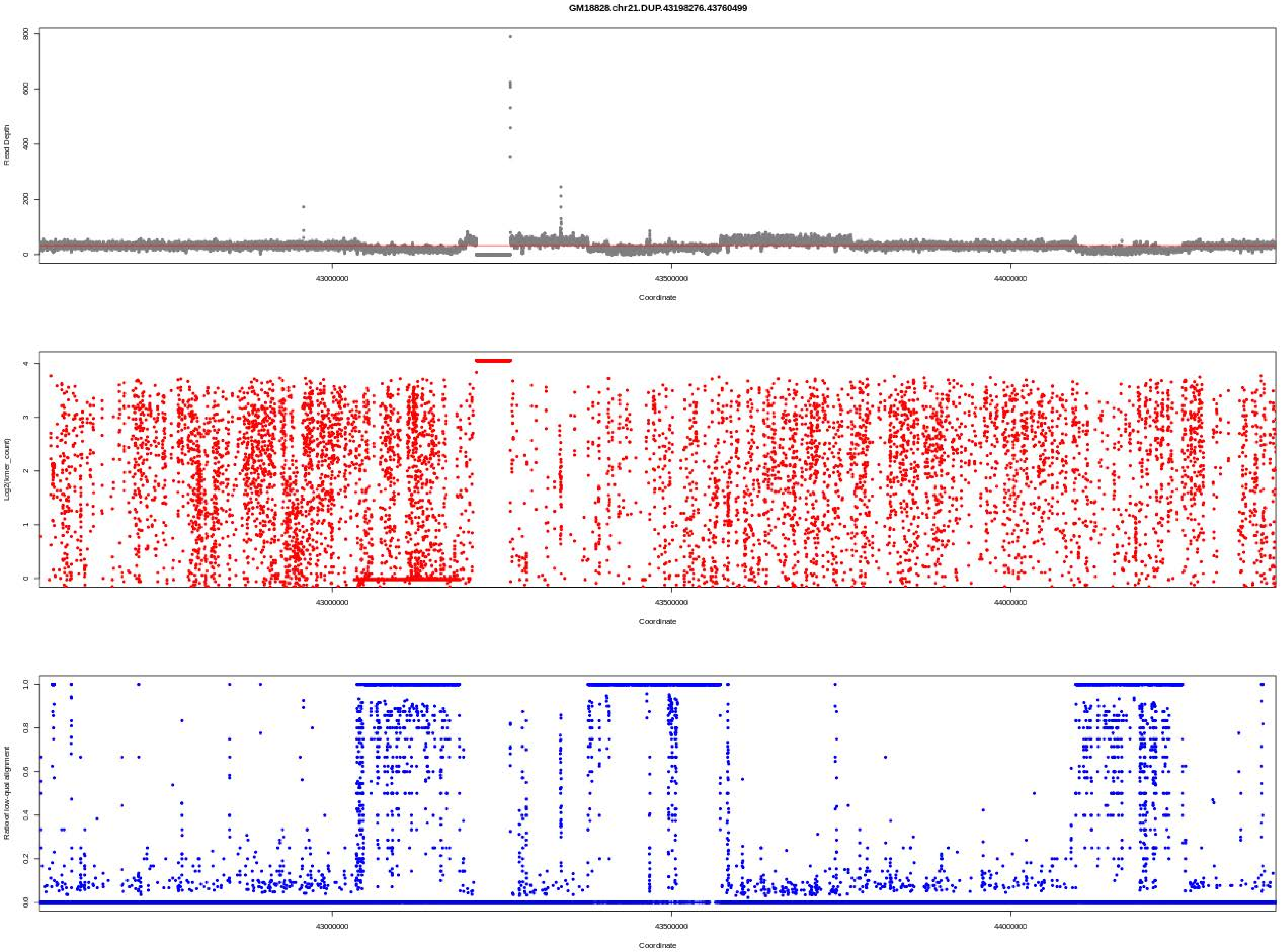

GM18828.chr21.DUP.43198276.43760499
Read Depth
Log2(kmer_count)
Ratio of low-qual alignment
Coordinate
43000000
43500000
44000000

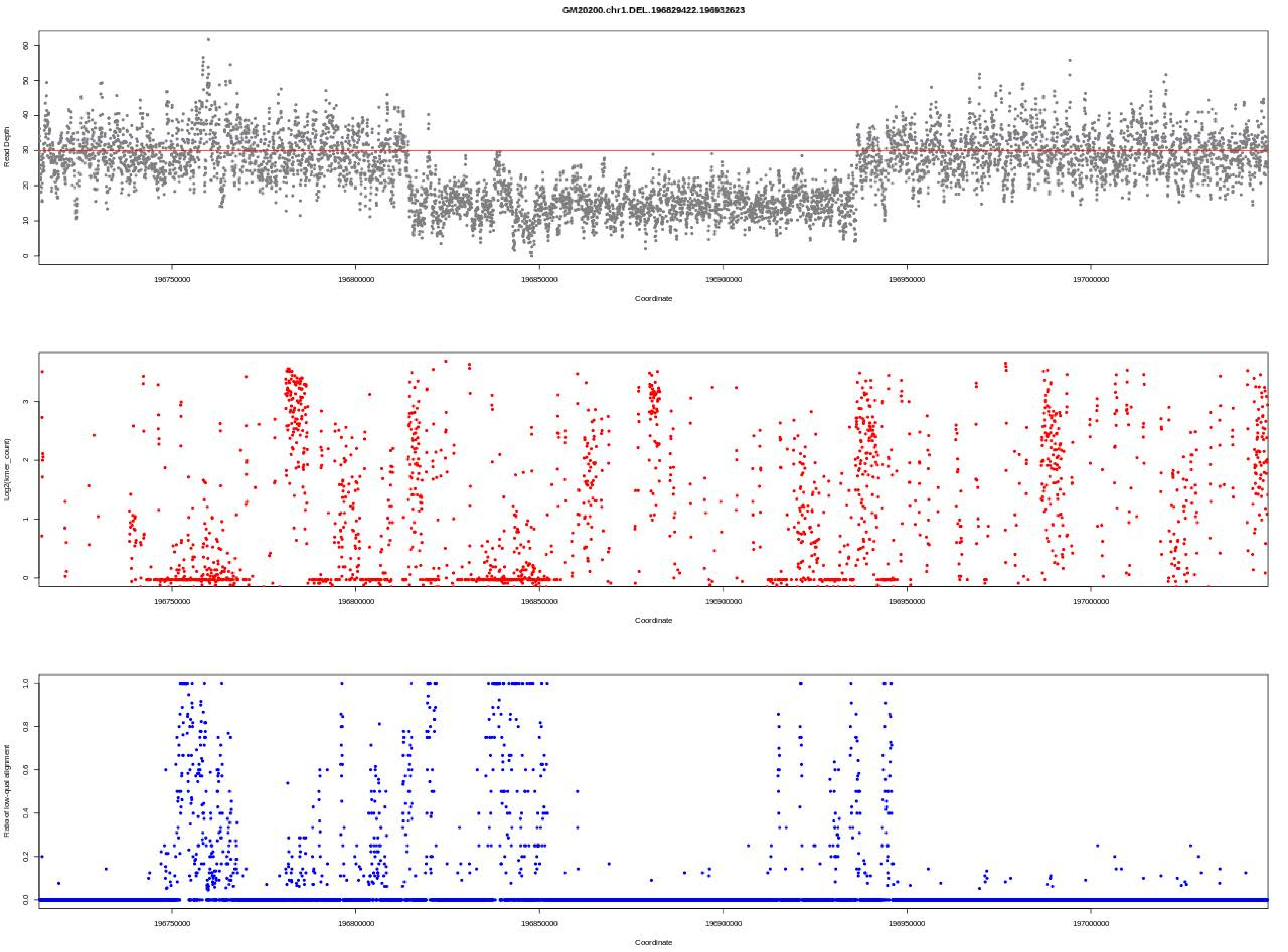
GM20200.chr1.DEL..196829422.196932623
Read Depth
Coordinate
Log2(kmer_count)
Coordinate
Ratio of low-qual alignment
Coordinate
196750000
196800000
196850000
196900000
196950000
197000000

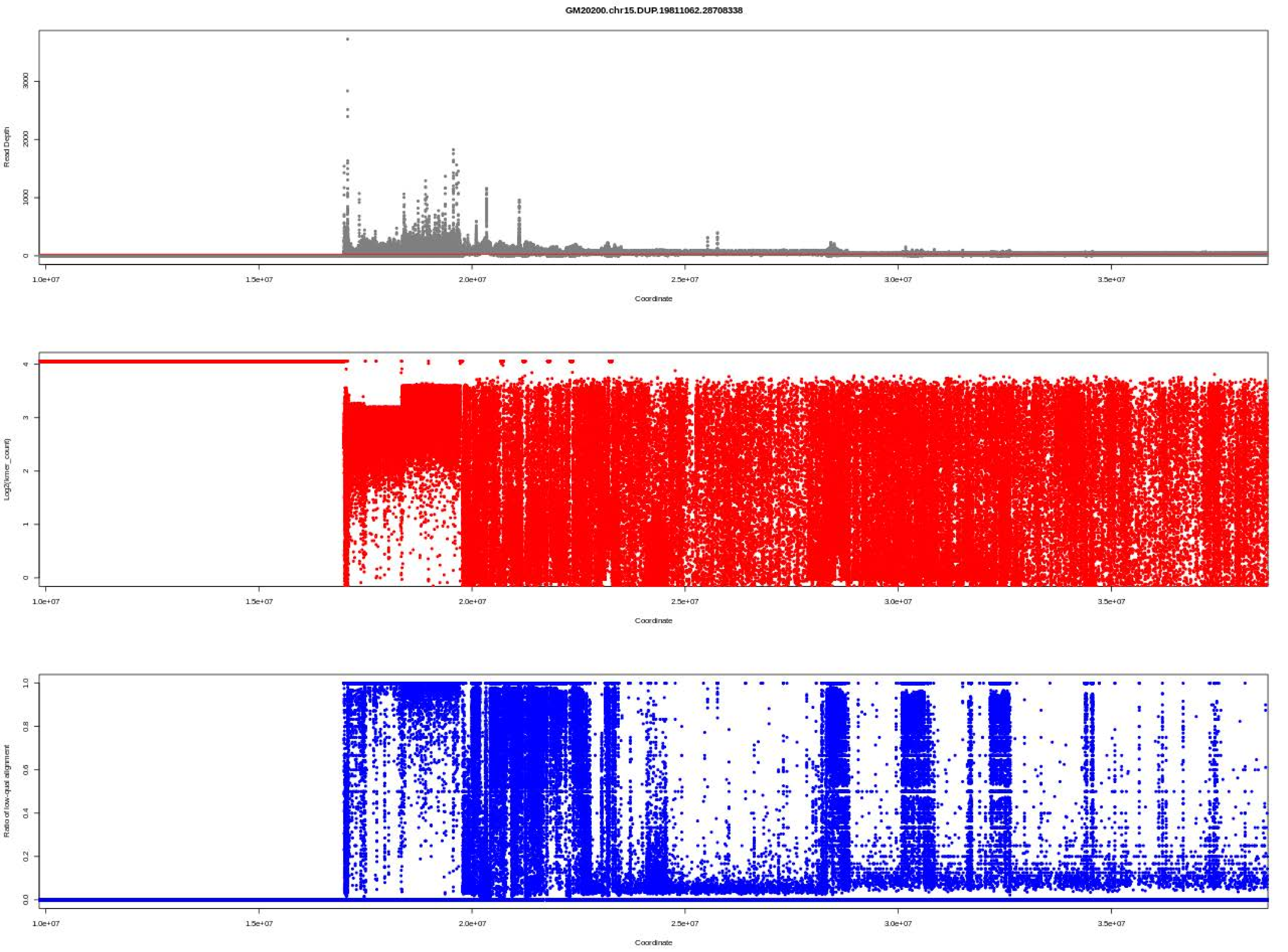
GM20200.chr15.DUP.19811062.28708338
Read Depth
Coordinate
Log2(kmer_count)
Coordinate
Ratio of low-qual alignment
Coordinate

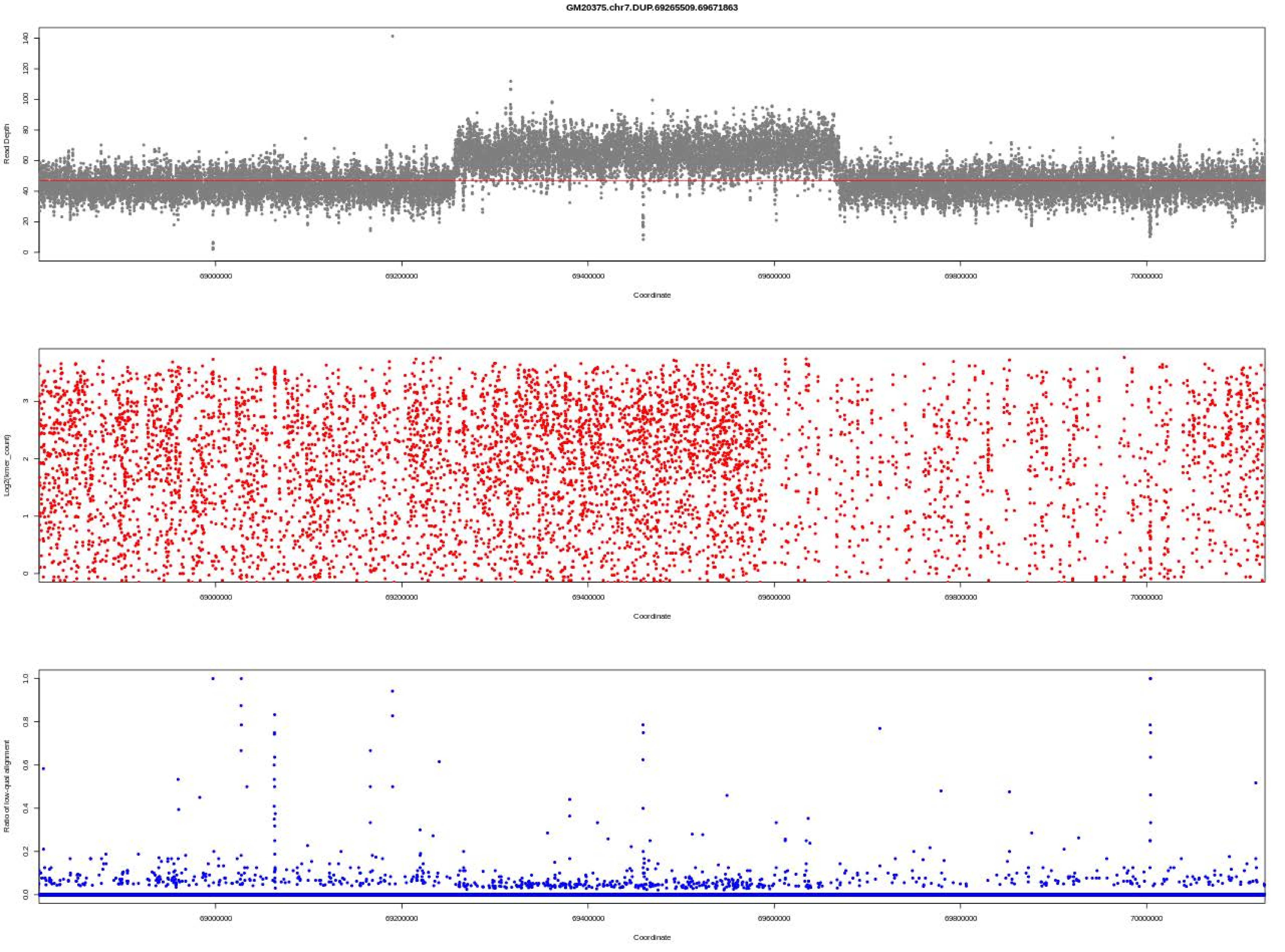
GM20375.chr7.DUP.69265509.69671863
Read Depth
Coordinate
Log2(kmer_count)
Ratio of low-qual alignment
69000000
69200000
69400000
69600000
69800000
70000000

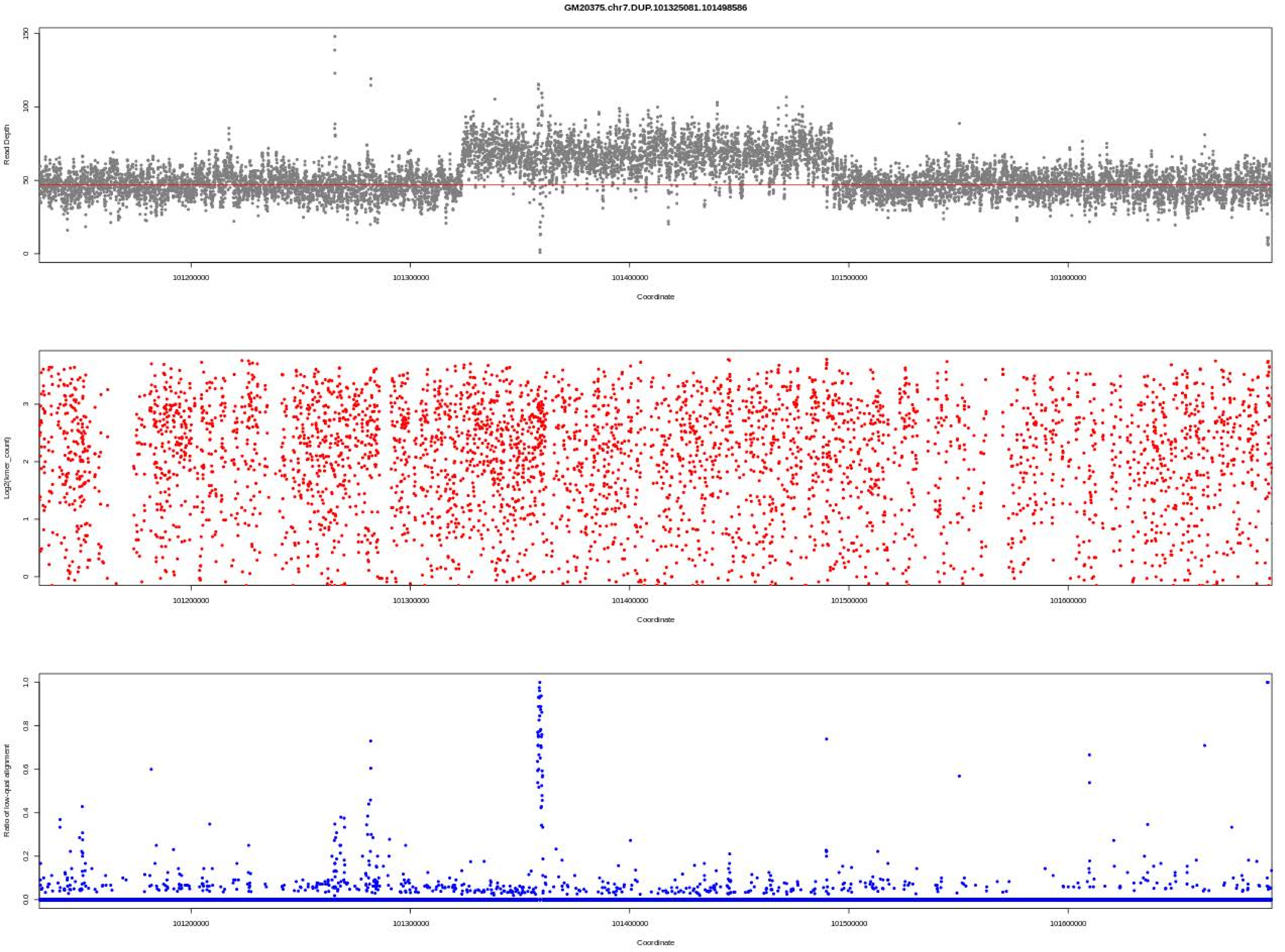

GM20375.chr7.DUP.101325081.101498586
Read Depth
Coordinate
Log2(kmer_count)
Coordinate
Ratio of low-qual alignment
Coordinate
101200000
101300000
101400000
101500000
101600000

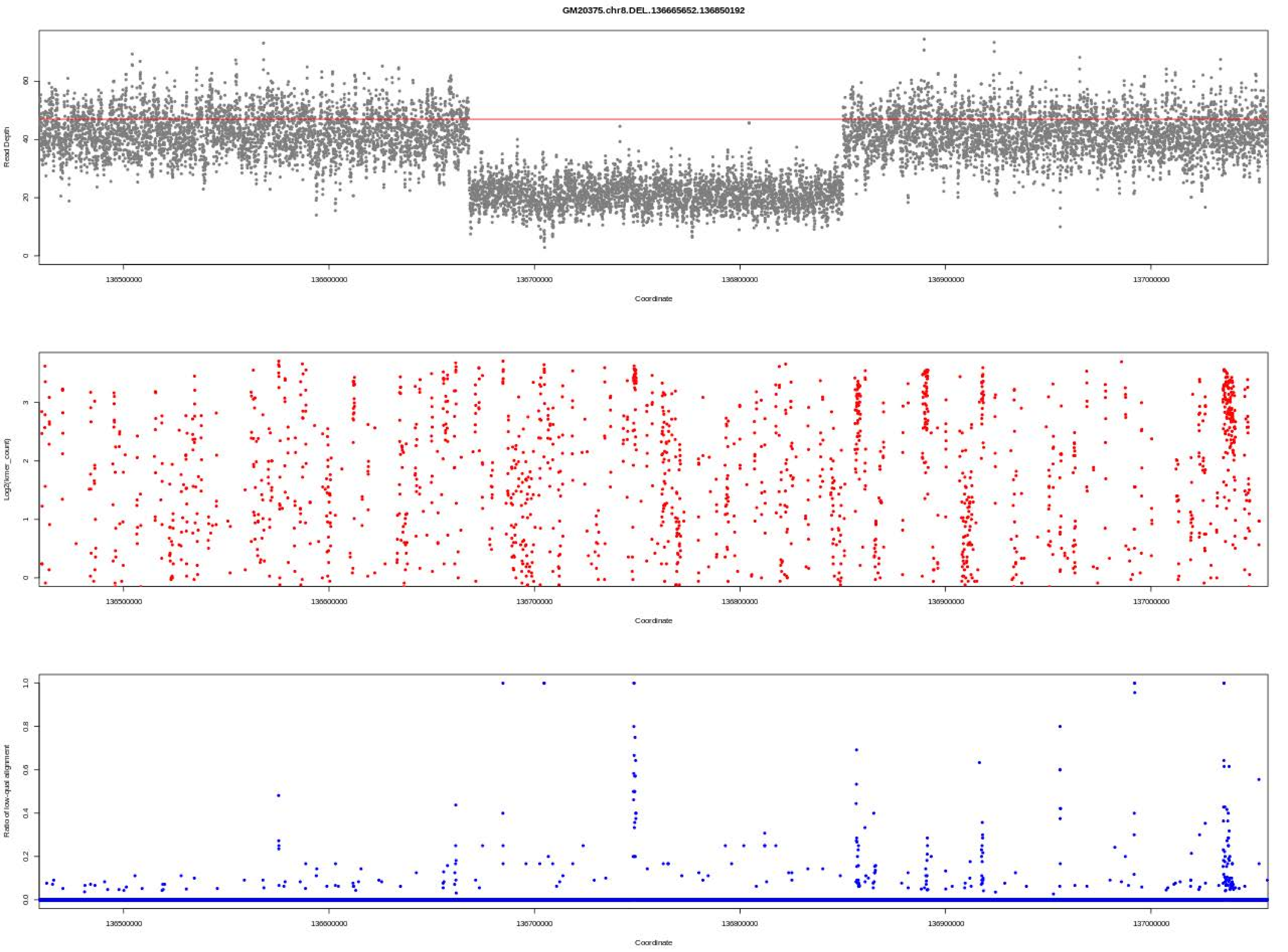

GM20375.chr8.DEL..136665652.136850192
Read Depth
Coordinate
Log2(kmer_count)
Coordinate
Ratio of low-qual alignment
Coordinate

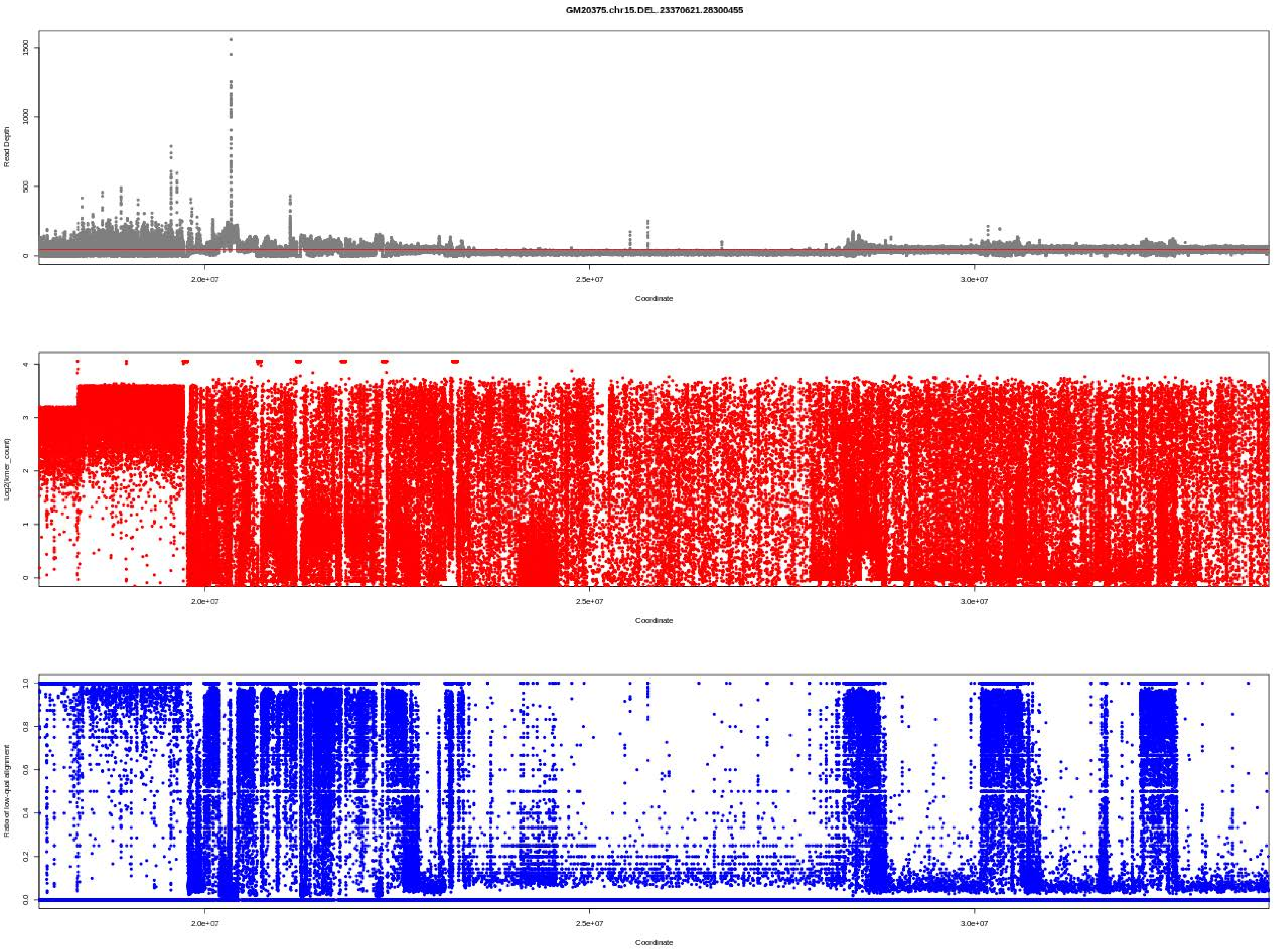

GM20375.chr15.DEL..23370621.28300455
Read Depth
0
500
1000
1500
2.0e+07
2.5e+07
3.0e+07
Coordinate
Log2(kmer_count)
0
1
2
3
4
2.0e+07
2.5e+07
3.0e+07
Coordinate
Ratio of low-qual alignment
0.0
0.2
0.4
0.6
0.8
1.0
2.0e+07
2.5e+07
3.0e+07
Coordinate

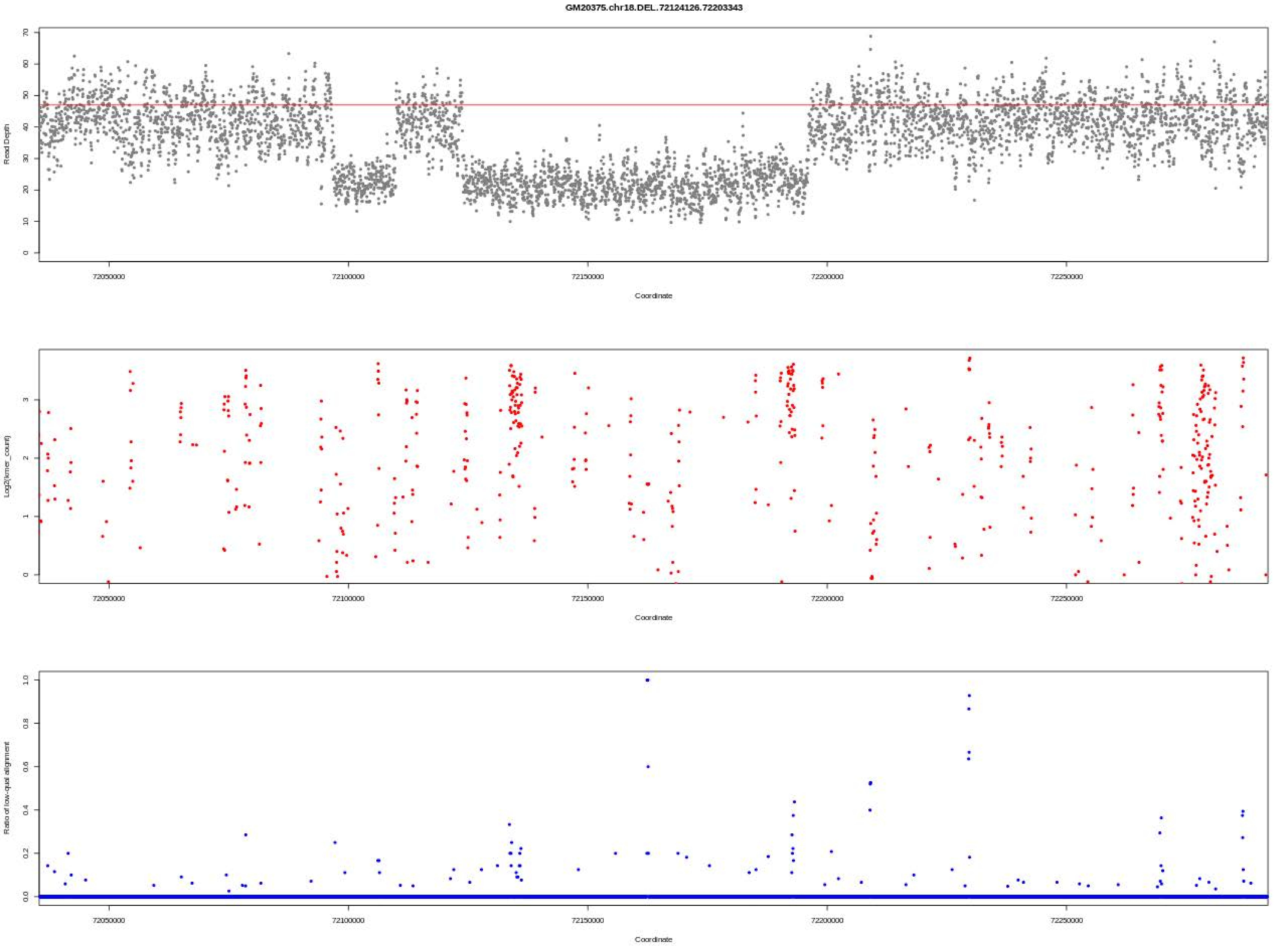
GM20375.chr18.DEL.72124126.72203343
Read Depth
Coordinate
Log2(kmer_count)
Coordinate
Ratio of low-qual alignment
Coordinate
72050000
72100000
72150000
72200000
72250000

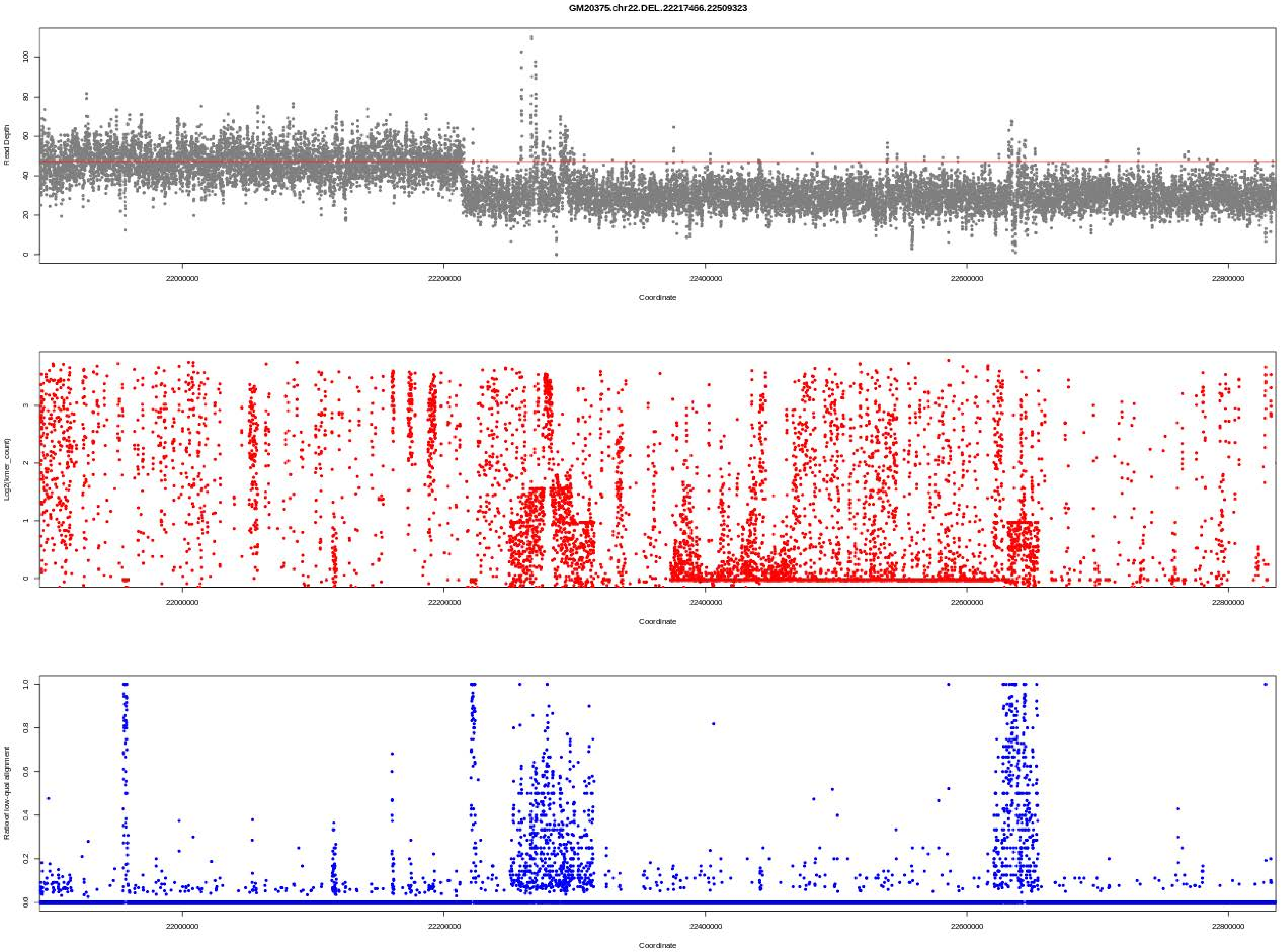
GM20375.chr22.DEL.22217466.22509323
Read Depth
Coordinate
Log2(kmer_count)
Coordinate
Ratio of low-qual alignment
Coordinate
22000000
22200000
22400000
22600000
22800000

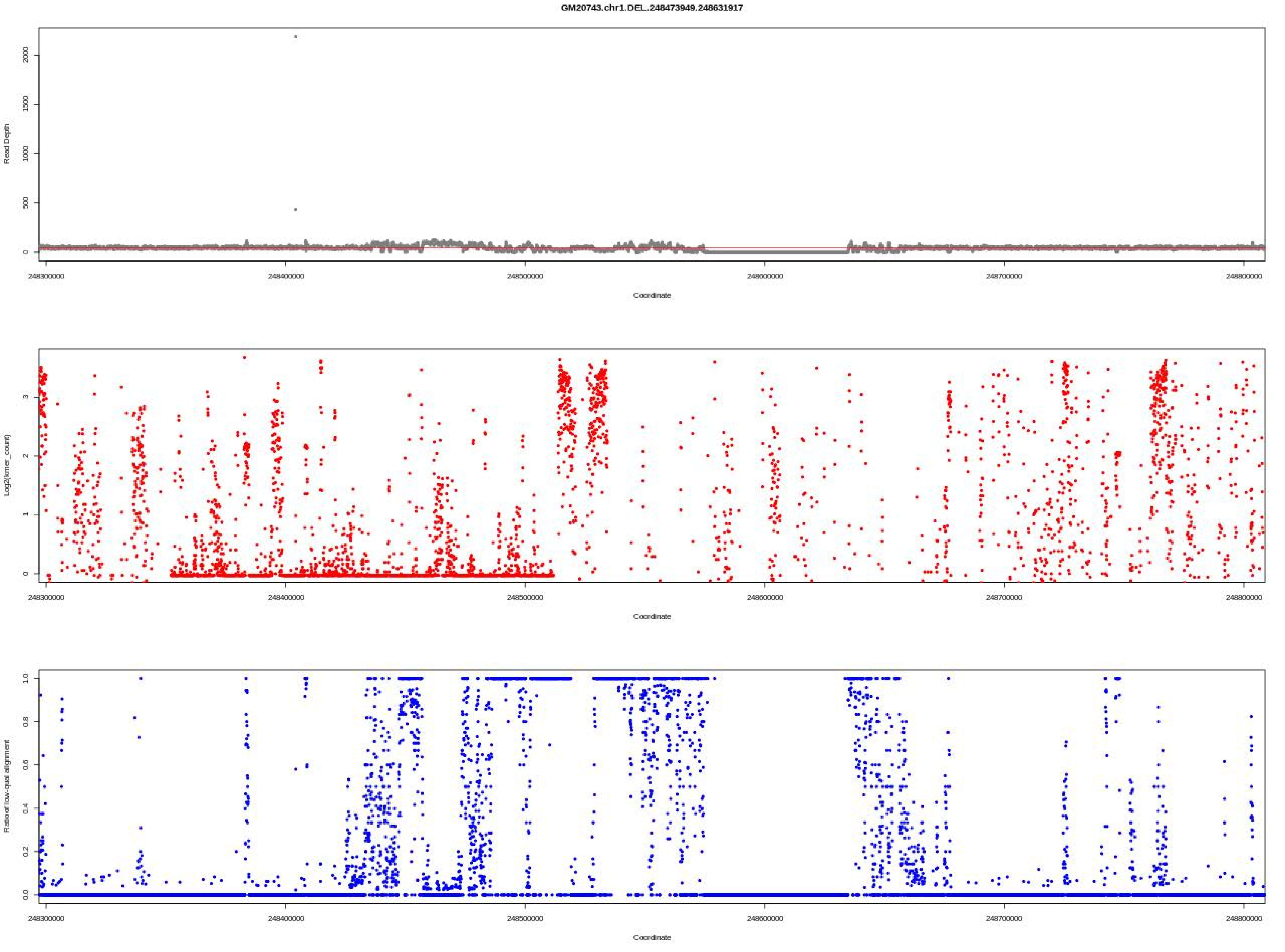

GM20743.chr1.DEL.248473949.248631917
Read Depth
0
500
1000
1500
2000
248300000
248400000
248500000
248600000
248700000
248800000
Coordinate
Log2(kmer_count)
0
1
2
3
Coordinate
Ratio of low-qual alignment
0.0
0.2
0.4
0.6
0.8
1.0
Coordinate

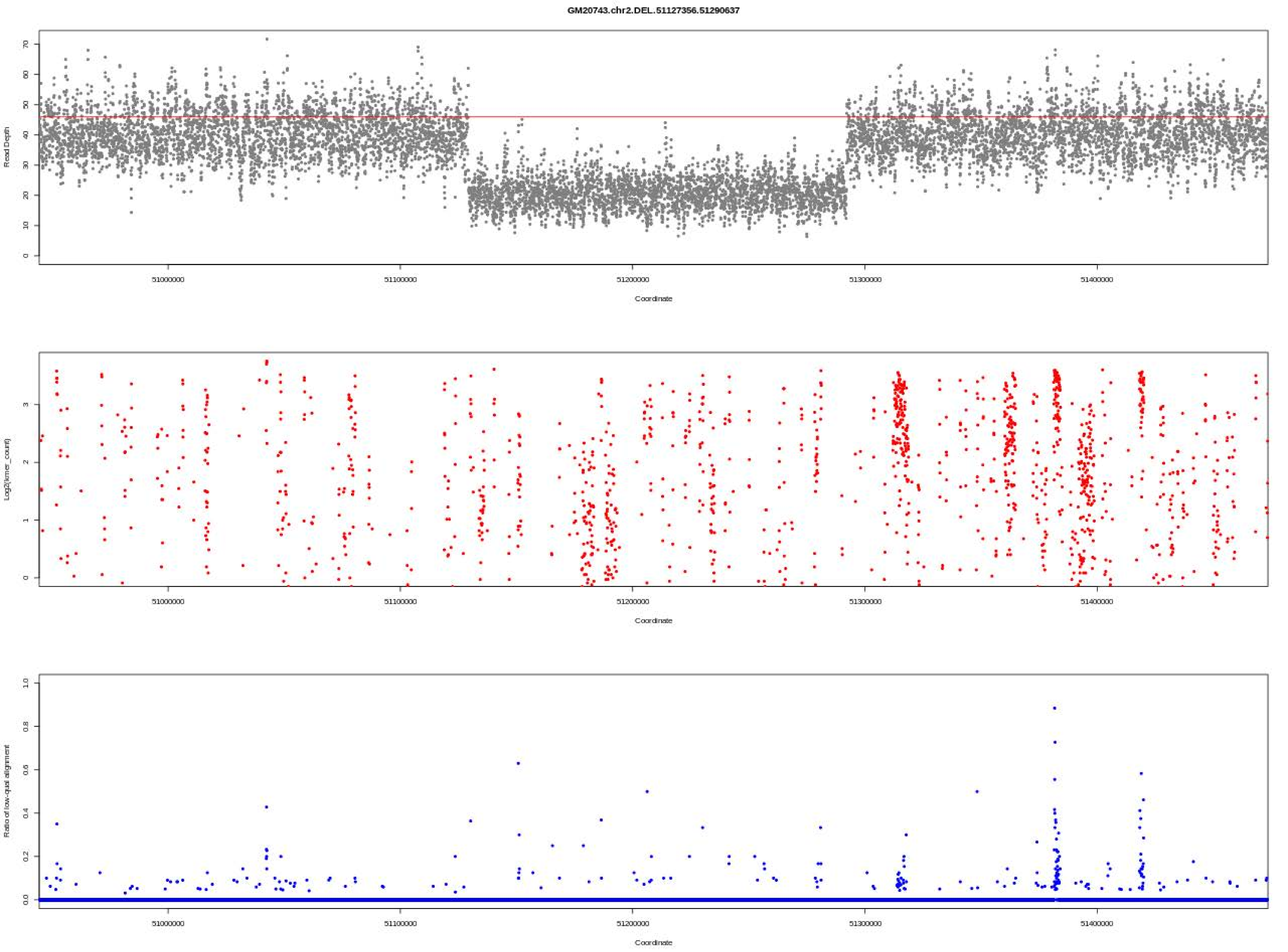
GM20743.chr2.DEL.51127356.51290637
Read Depth
0
10
20
30
40
50
60
70
51000000
51100000
51200000
51300000
51400000
Coordinate
Log2(kmer_count)
0
1
2
3
51000000
51100000
51200000
51300000
51400000
Coordinate
Ratio of low-qual alignment
0.0
0.2
0.4
0.6
0.8
1.0
51000000
51100000
51200000
51300000
51400000
Coordinate

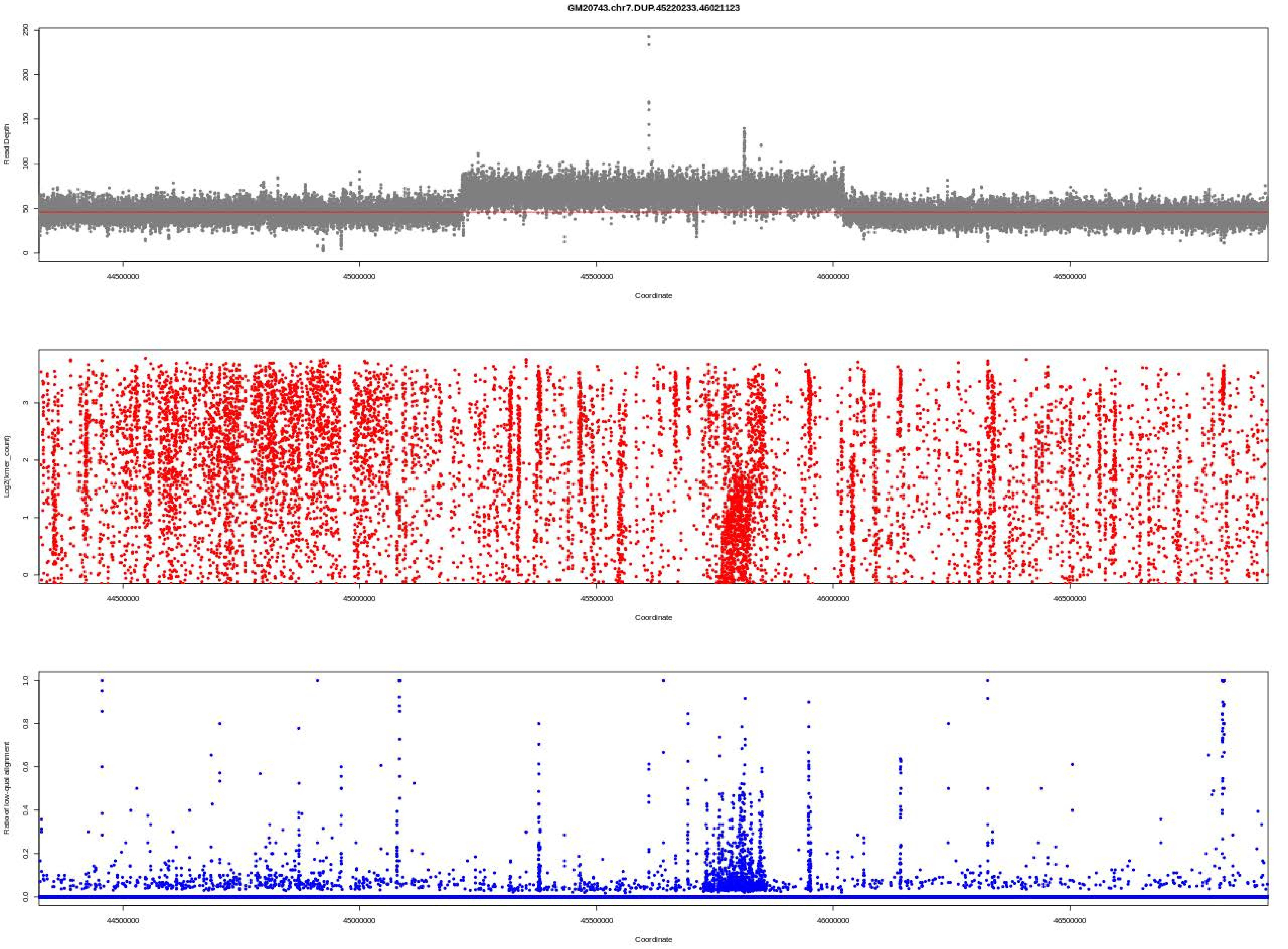
GM20743.chr7.DUP.45220233.46021123
Read Depth
Coordinate
Log2(kmer_count)
Coordinate
Ratio of low-qual alignment
Coordinate

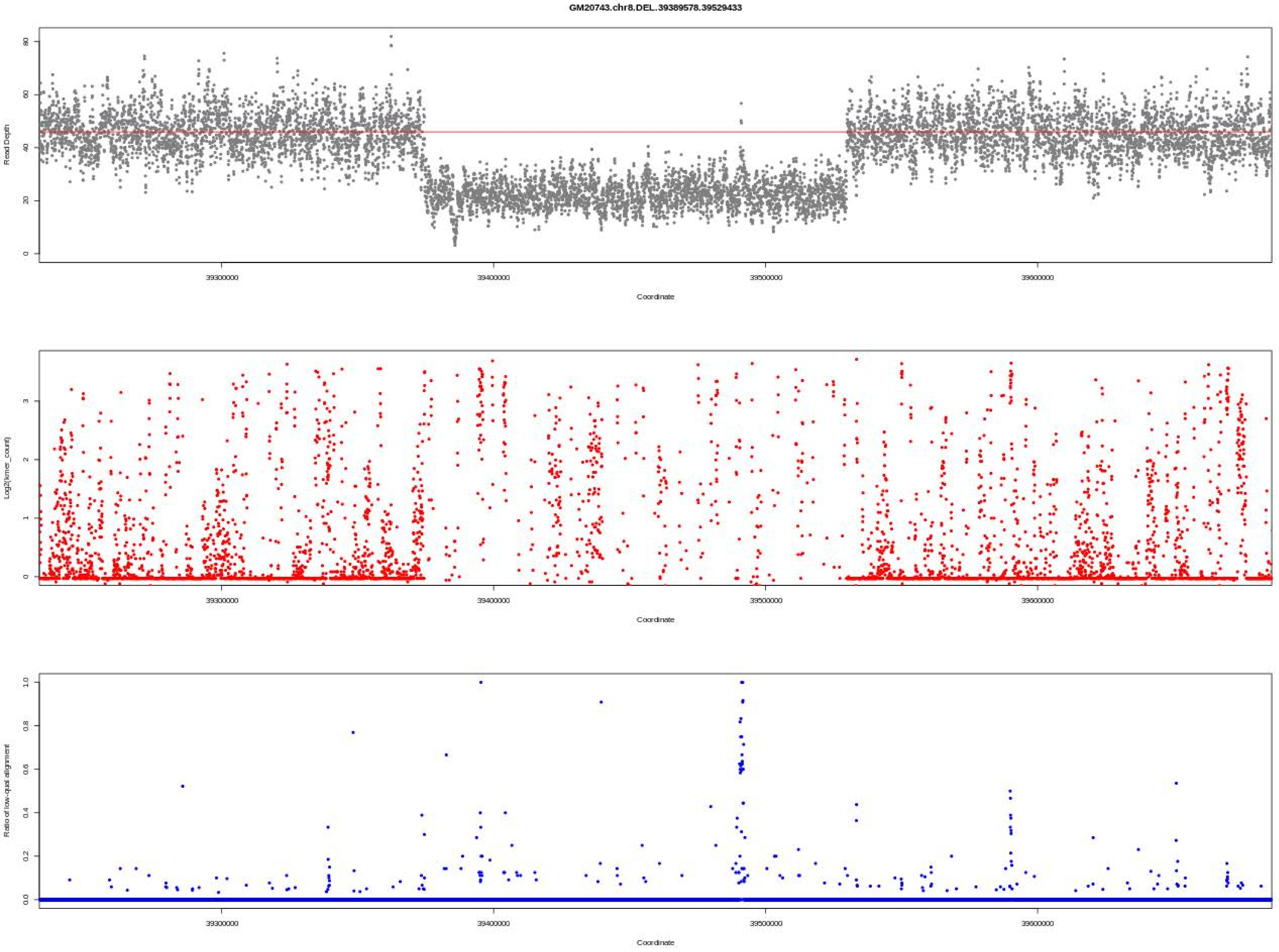
GM20743.chr8.DEL.39389578.39529433
Read Depth
Coordinate
Log2(kmer_count)
Coordinate
Ratio of low-qual alignment
Coordinate
39300000
39400000
39500000
39600000

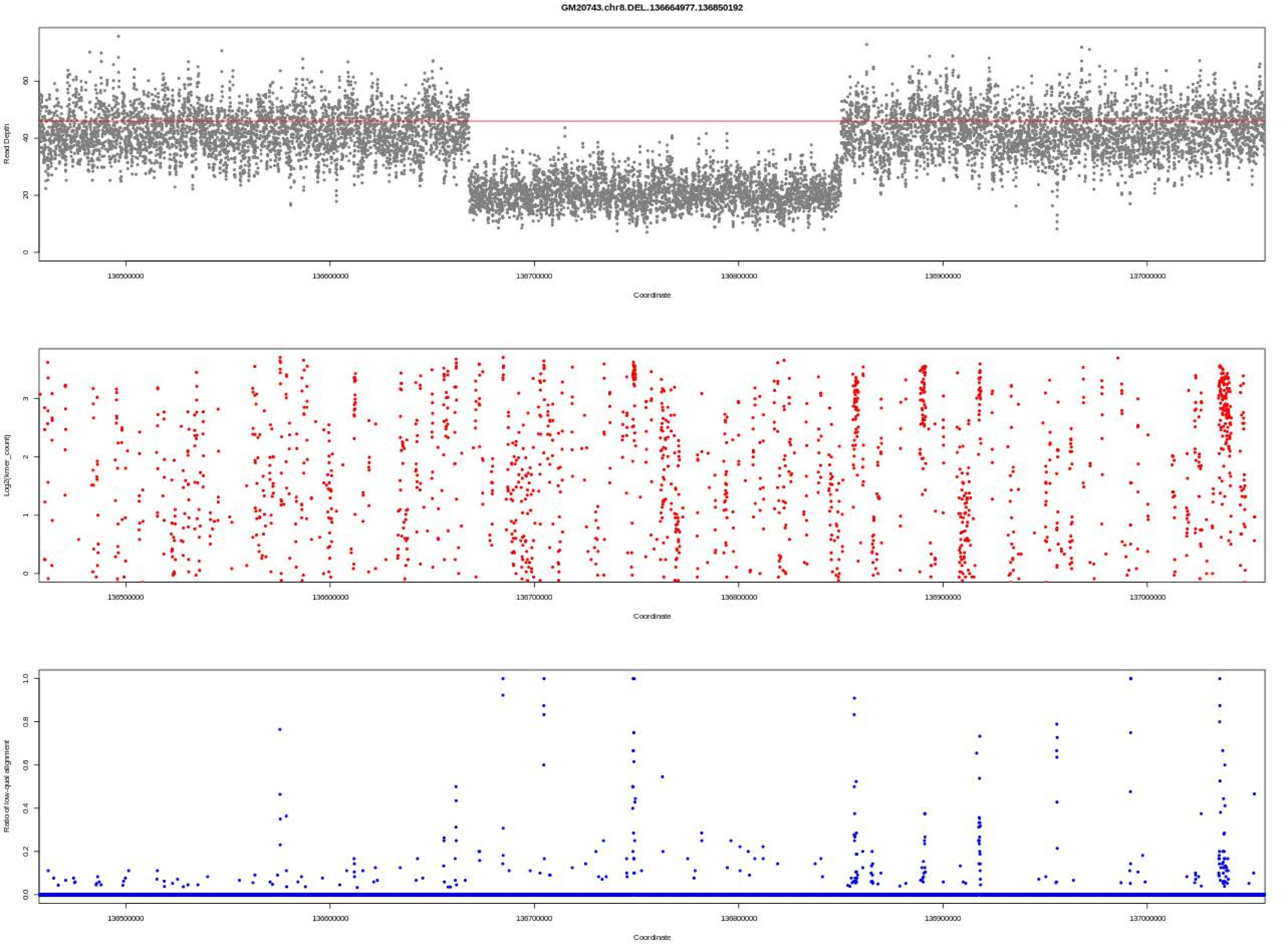
GM20743.chr8.DEL.136664977.136850192
Read Depth
Coordinate
Log2(kmer_count)
Coordinate
Ratio of low-qual alignment
Coordinate

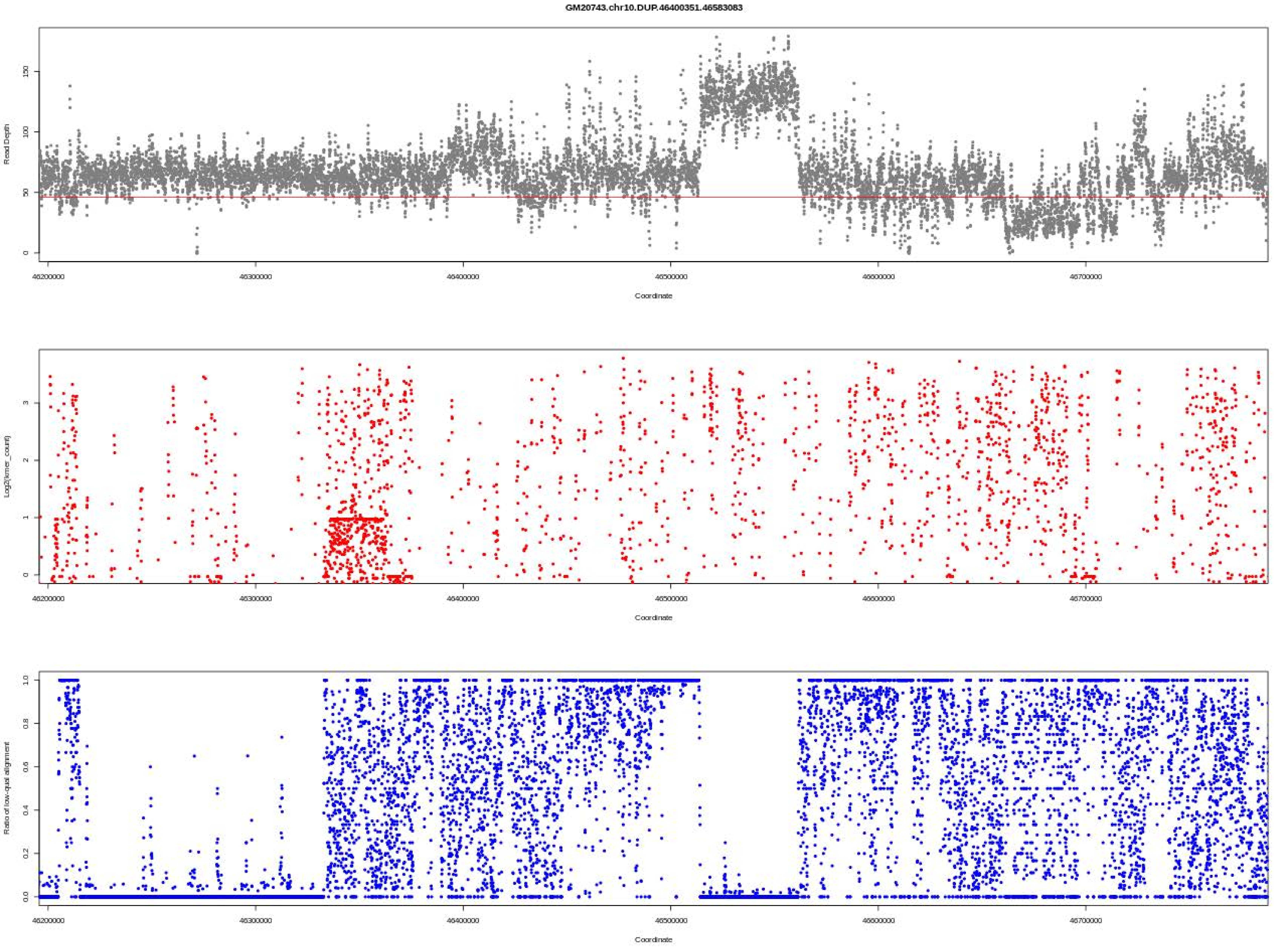
GM20743.chr10.DUP.46400351.46583083
Read Depth
Coordinate
Log2(kmer_count)
Coordinate
Ratio of low-qual alignment
Coordinate

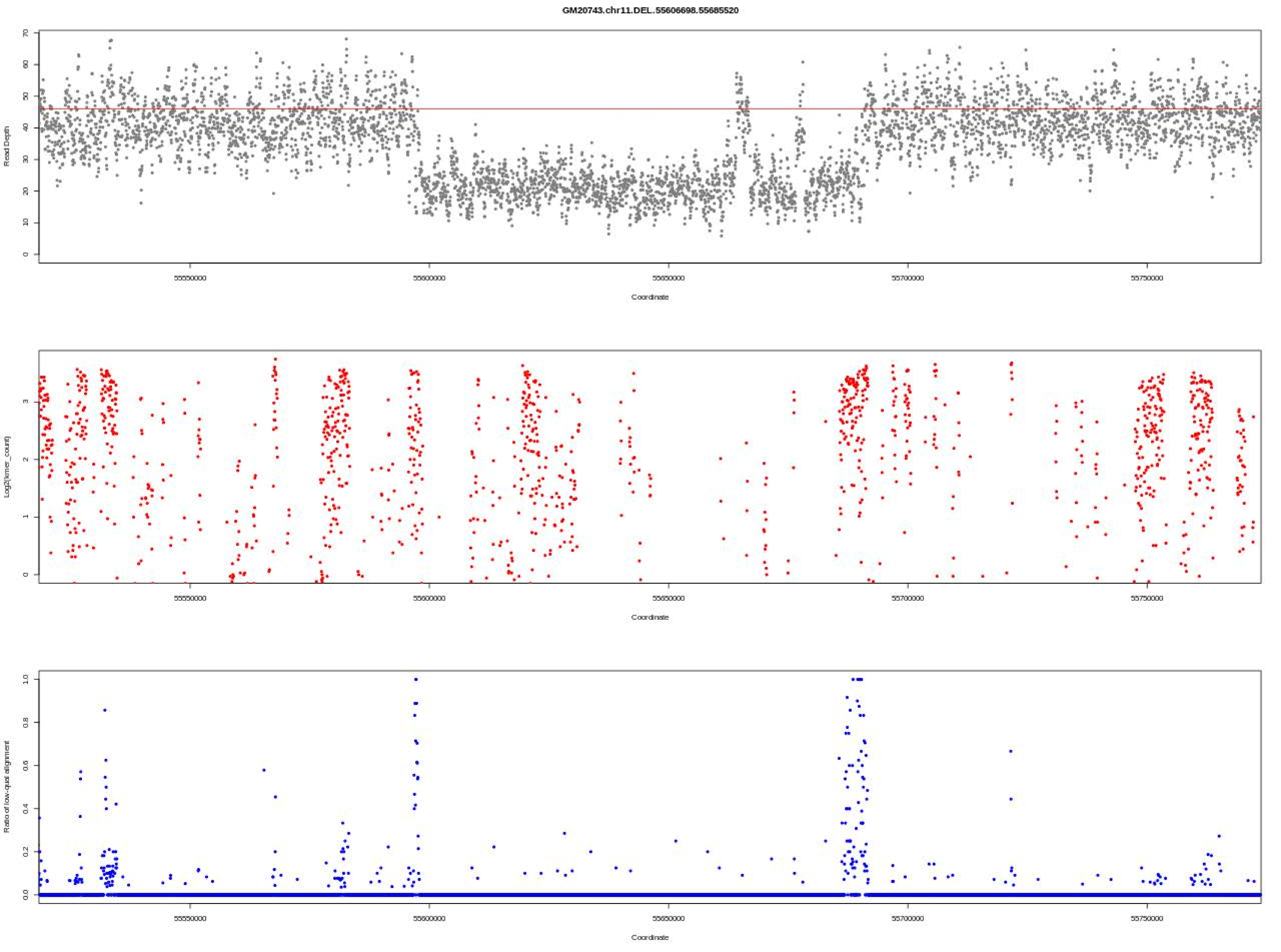
GM20743.chr11.DEL.55606698.55685520
Read Depth
Coordinate
Log2(kmer_count)
Coordinate
Ratio of low-qual alignment
Coordinate

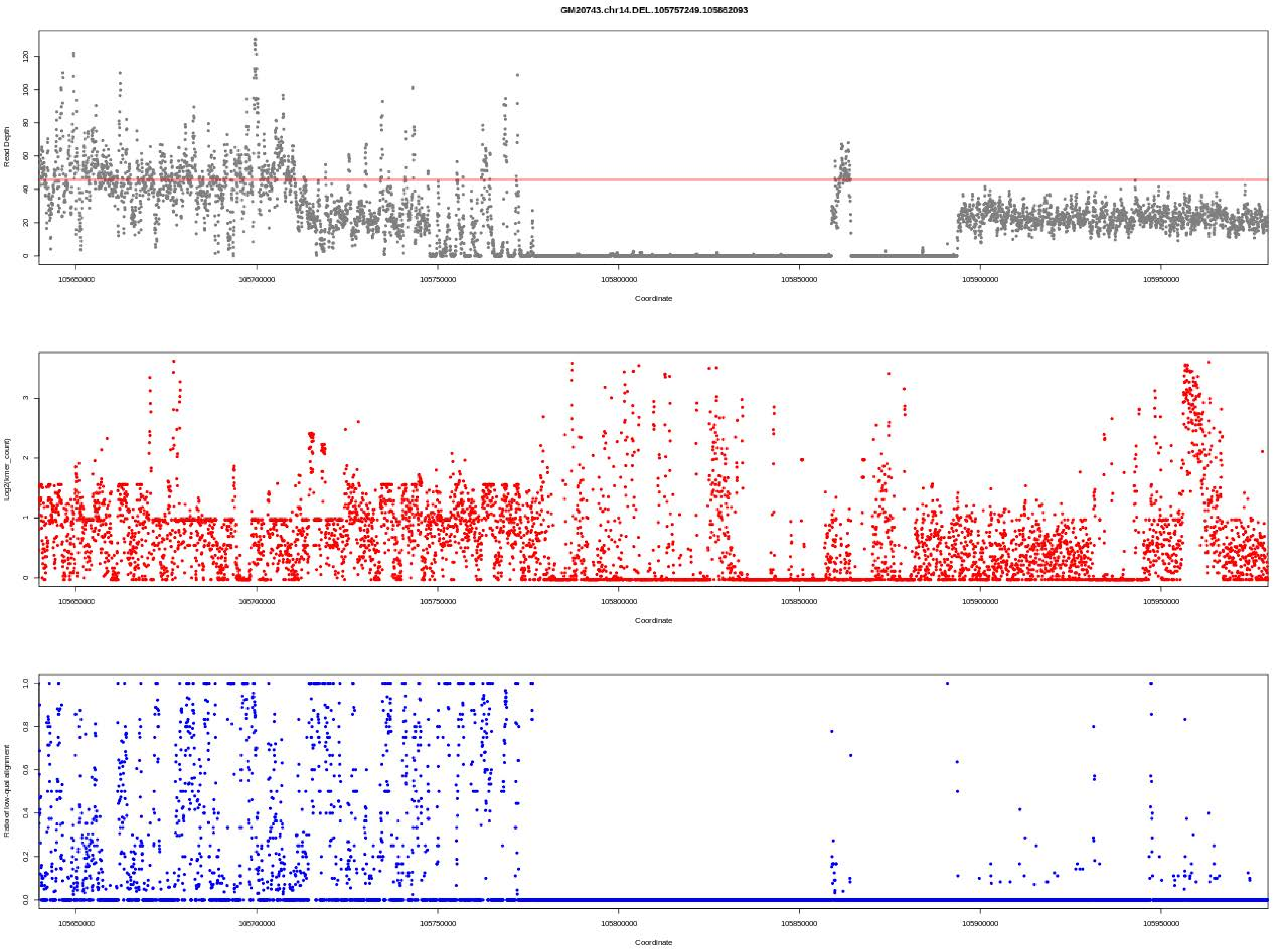
GM20743.chr14.DEL.105757249.105862093
Read Depth
Coordinate
Log2(kmer_count)
Coordinate
Ratio of low-qual alignment
Coordinate
105650000
105700000
105750000
105800000
105850000
105900000
105950000

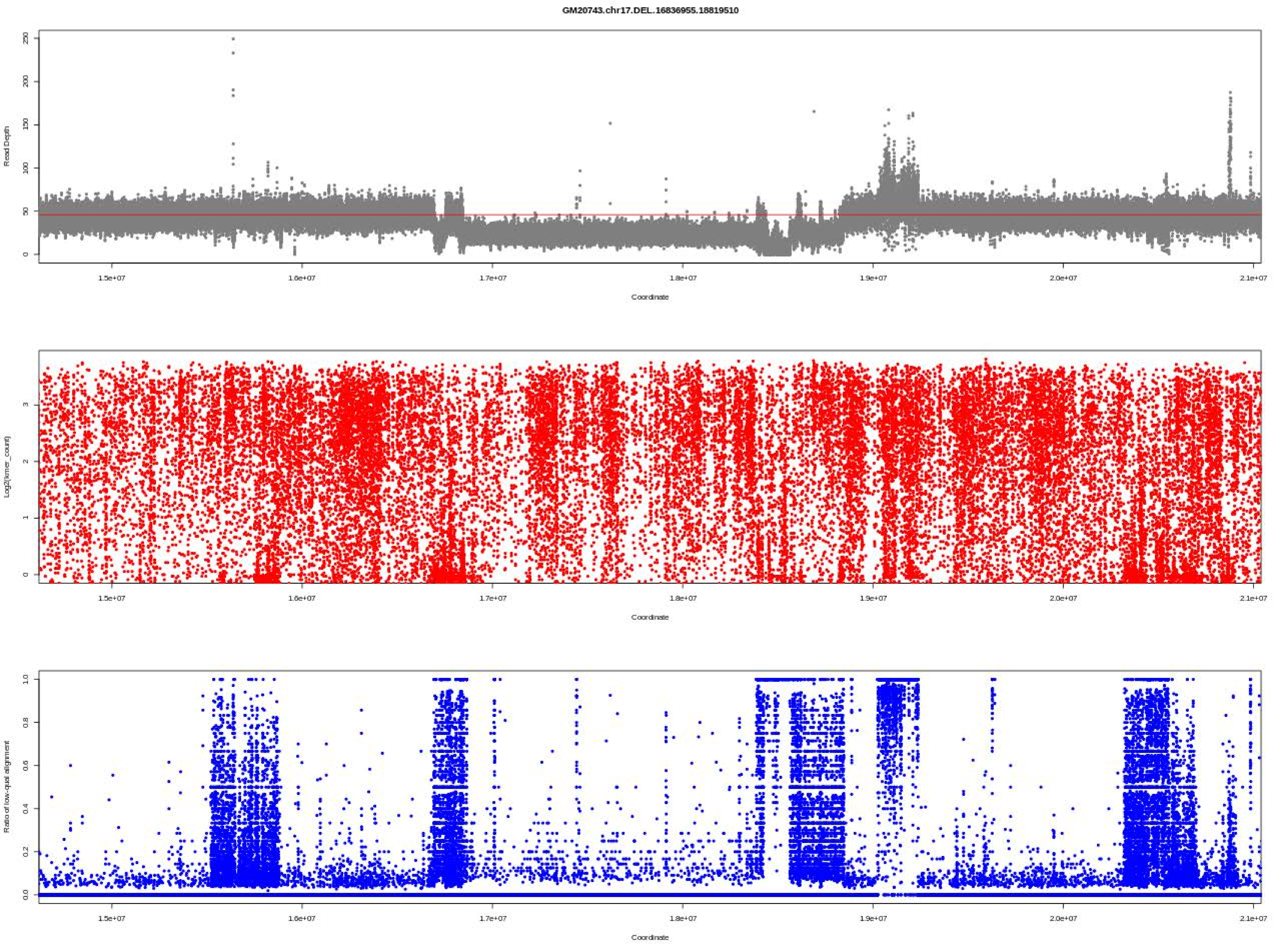

GM20743.chr17.DEL.16836955.18819510
Read Depth
Coordinate
Log2(kmer_count)
Coordinate
Ratio of low-qual alignment
Coordinate

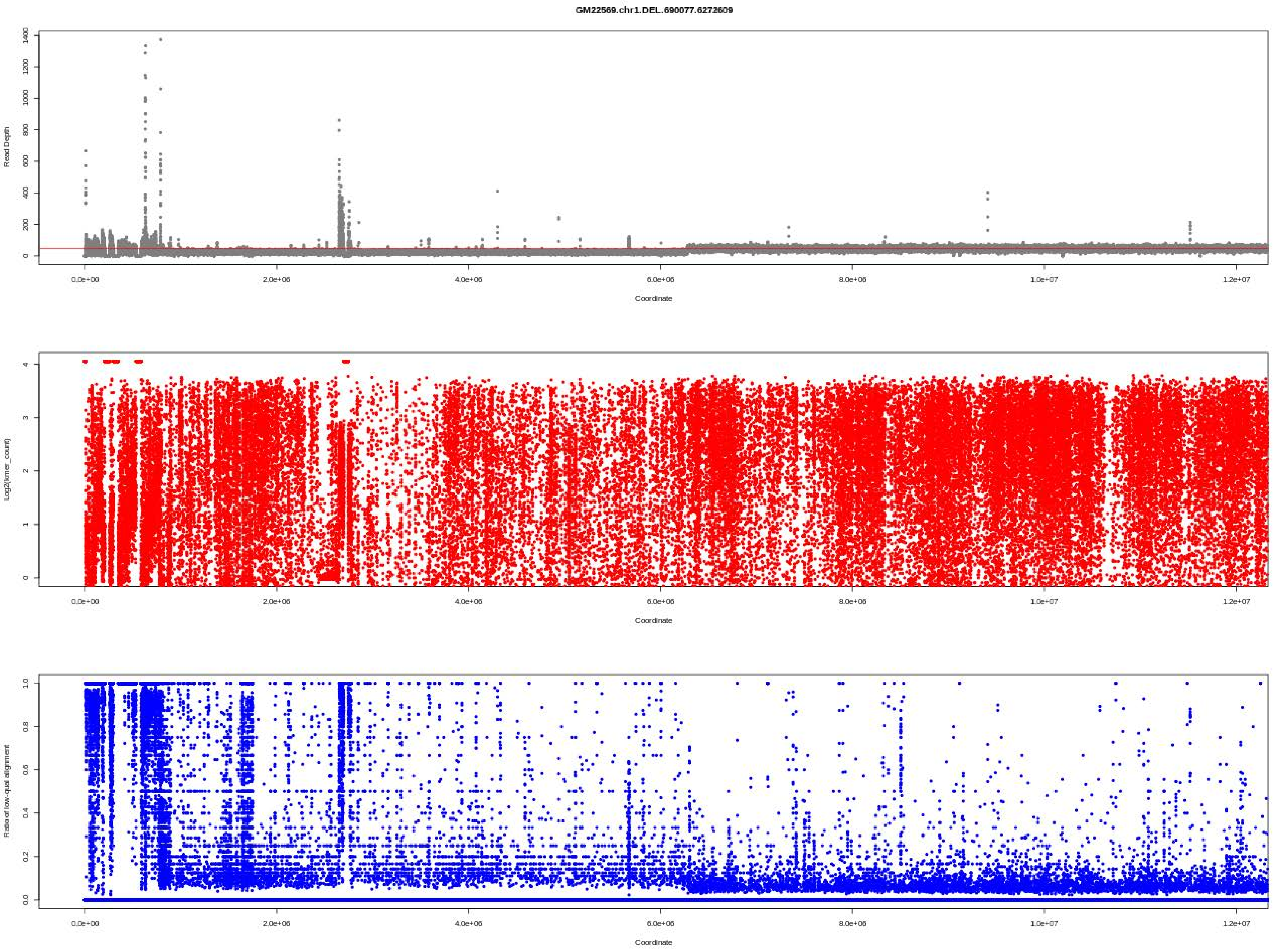
GM22569.chr1.DEL.690077.6272609
Read Depth
Coordinate
Log2(kmer_count)
Coordinate
Ratio of low-qual alignment
Coordinate

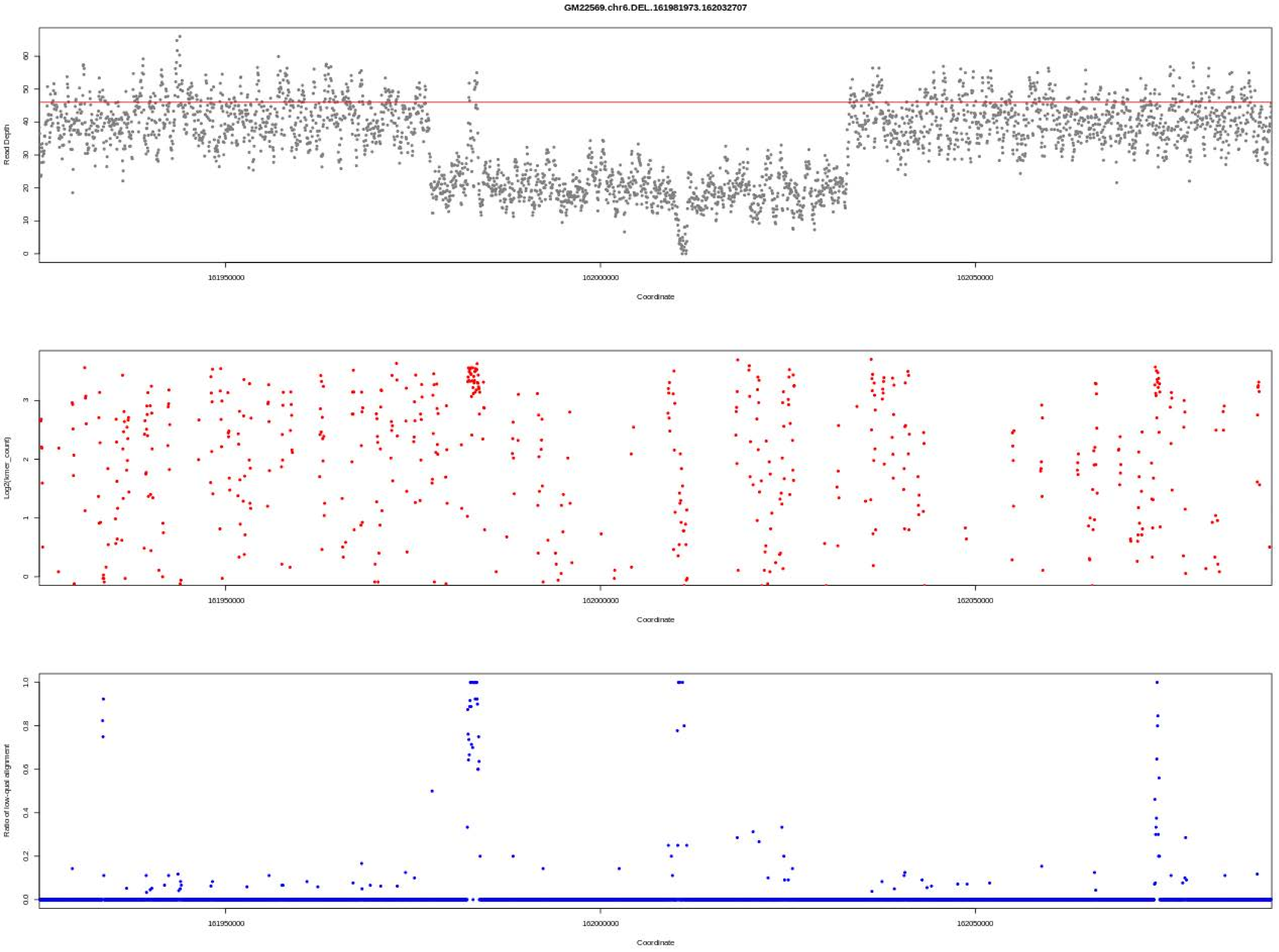
GM22569.chr6.DEL..161981973.162032707
Read Depth
Coordinate
Log2(kmer_count)
Coordinate
Ratio of low-qual alignment
Coordinate
161950000
162000000
162050000

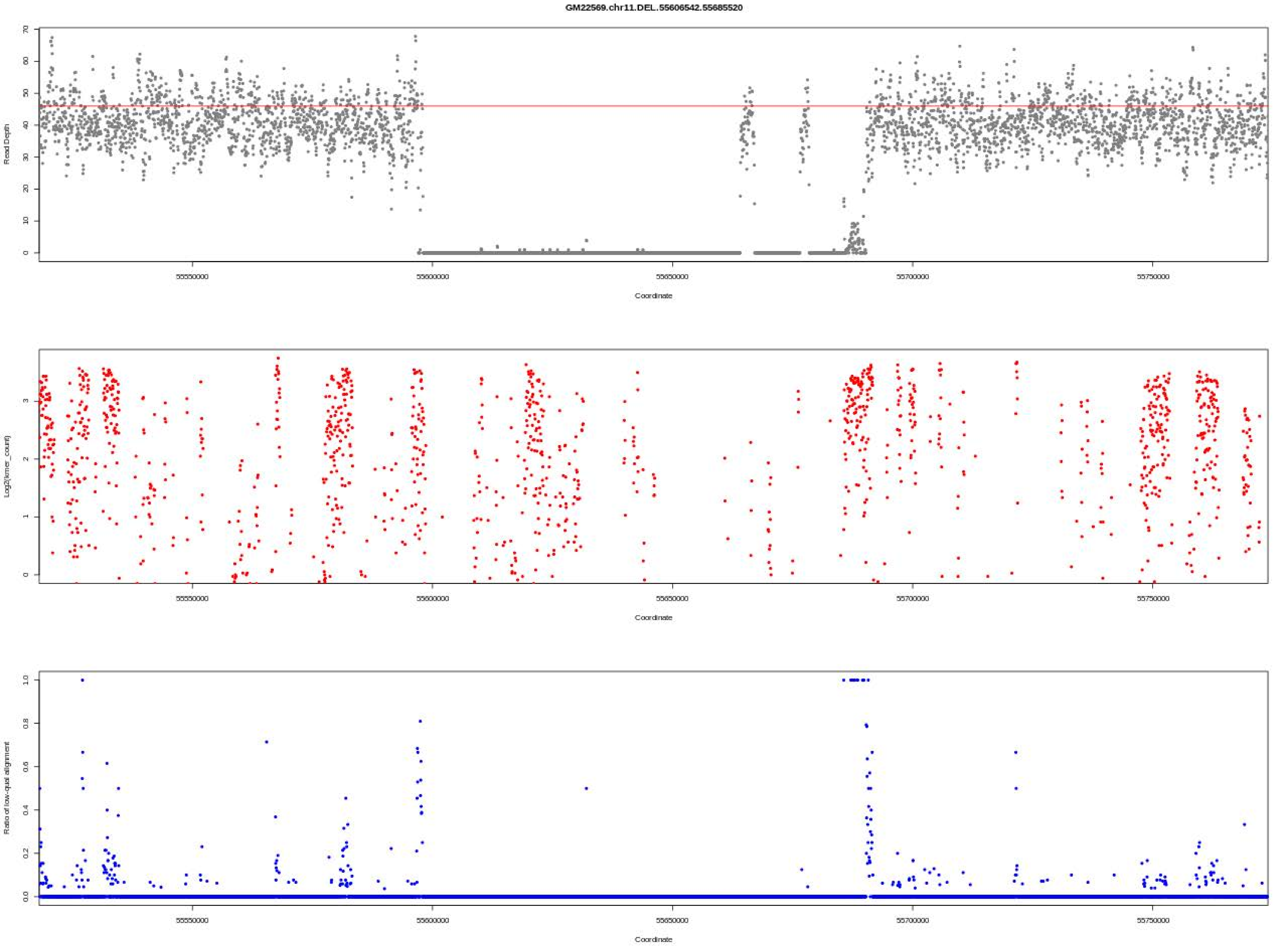
GM22569.chr11.DEL.55606542.55685520
Read Depth
Coordinate
Log2(kmer_count)
Coordinate
Ratio of low-qual alignment
Coordinate
55550000
55600000
55650000
55700000
55750000

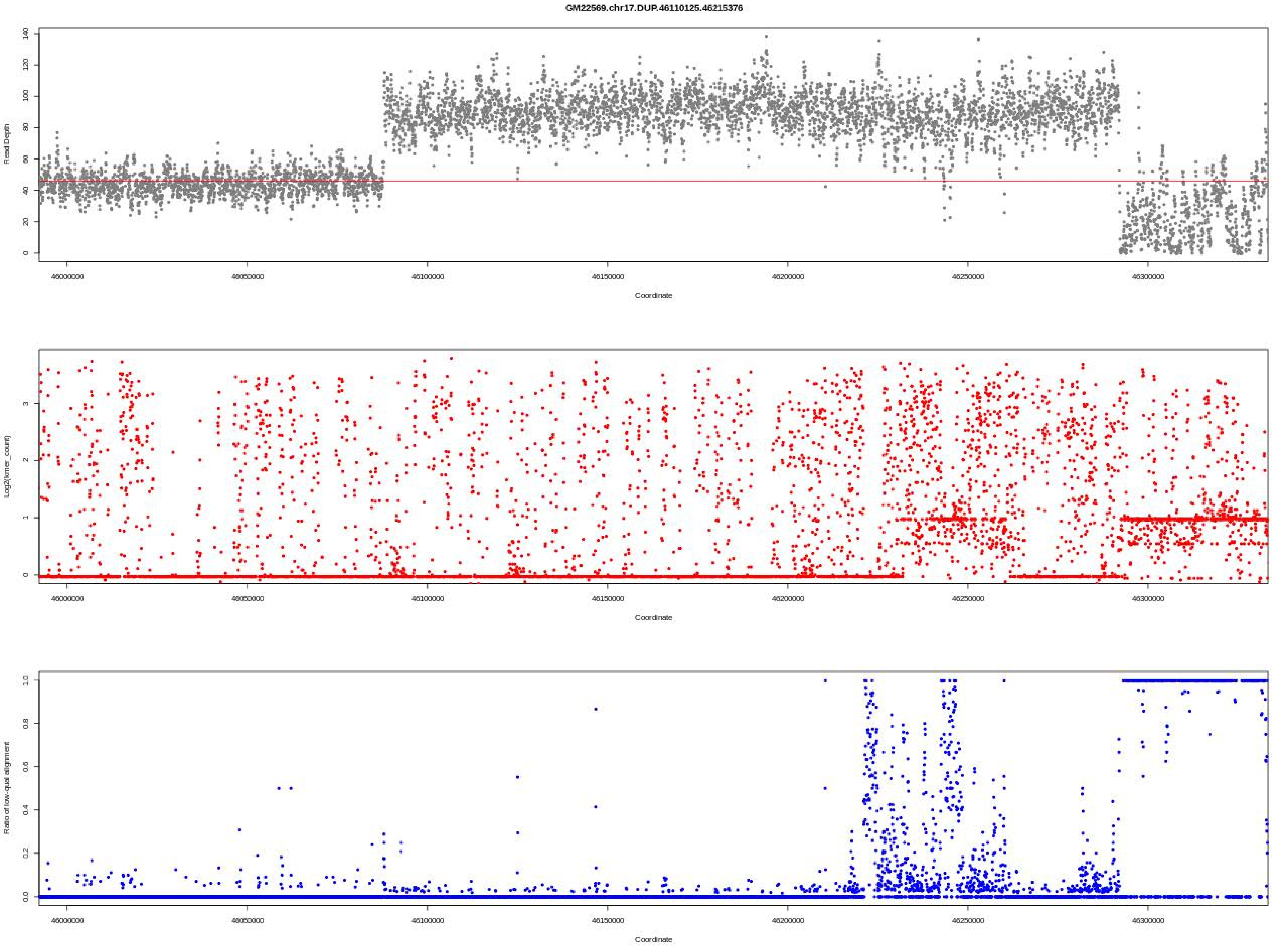

GM22569.chr17.DUP.46110125.46215376
Read Depth
Coordinate
Log2(kmer_count)
Coordinate
Ratio of low-qual alignment
Coordinate

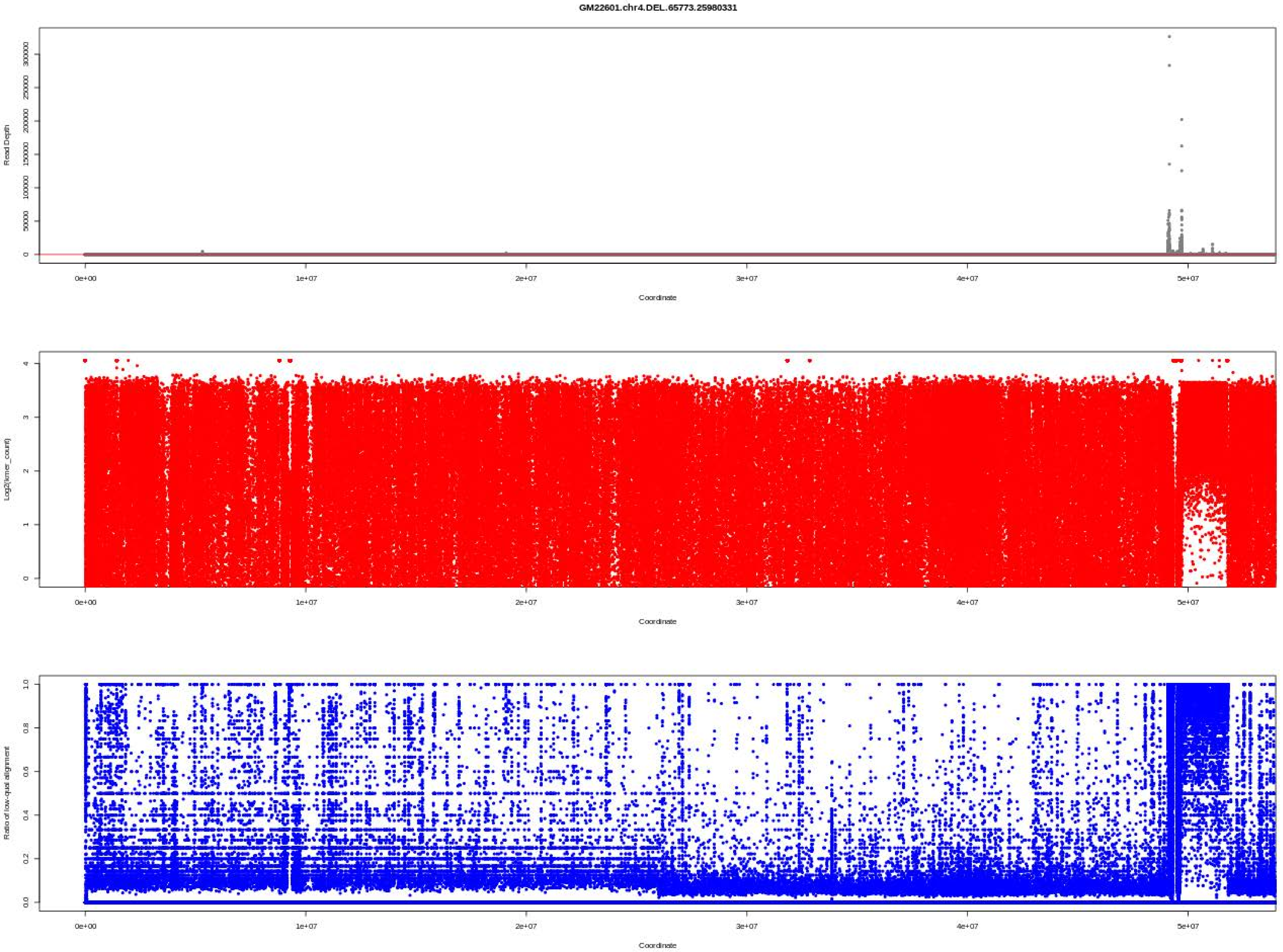
GM22601.chr4.DEL.65773.25980331
Read Depth
Coordinate
Log2(kmer_count)
Coordinate
Ratio of low-qual alignment
Coordinate

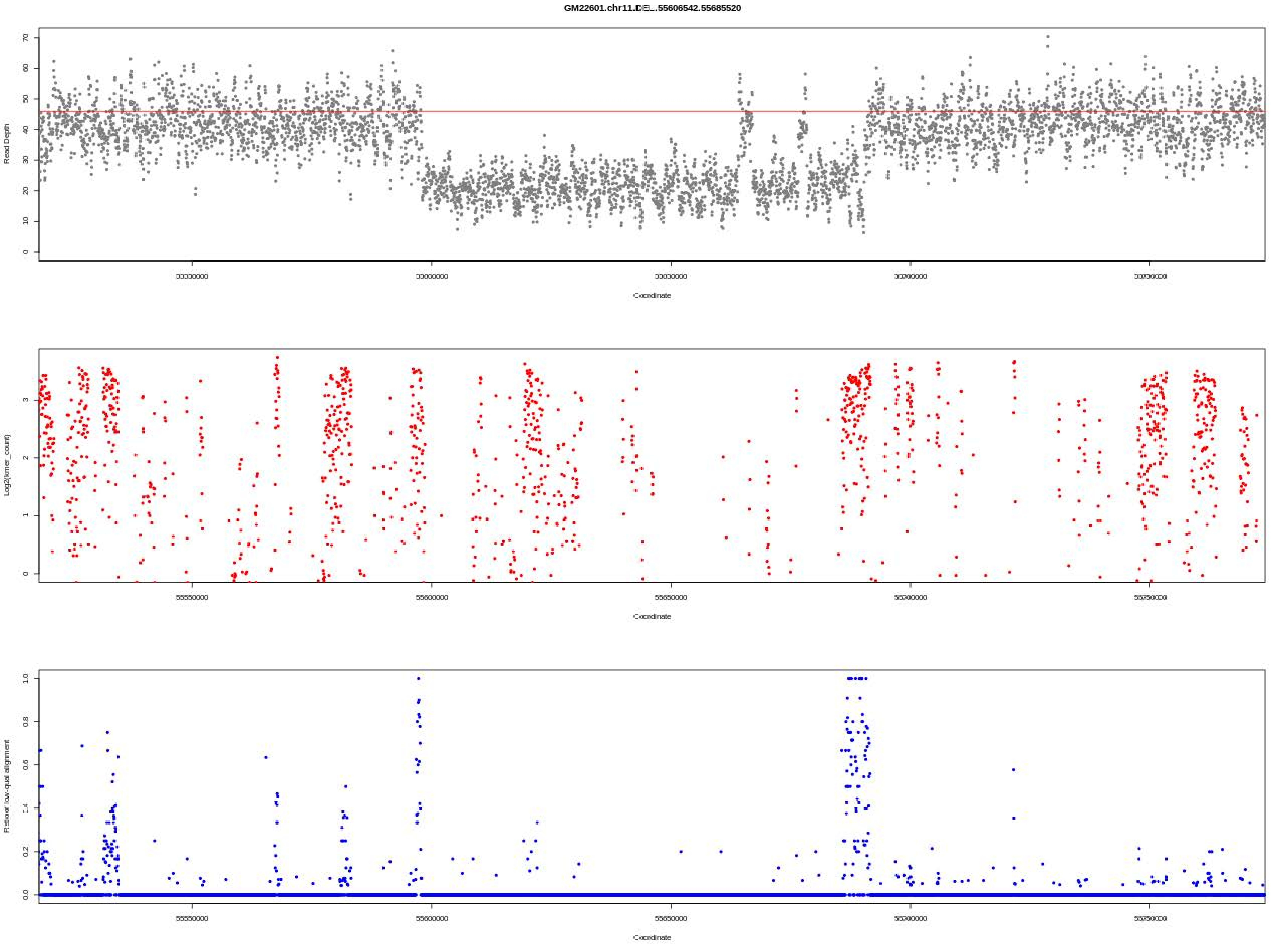

GM22601.chr11.DEL.55606542.55685520
Read Depth
Log2(kmer_count)
Ratio of low-qual alignment
Coordinate
55550000
55600000
55650000
55700000
55750000

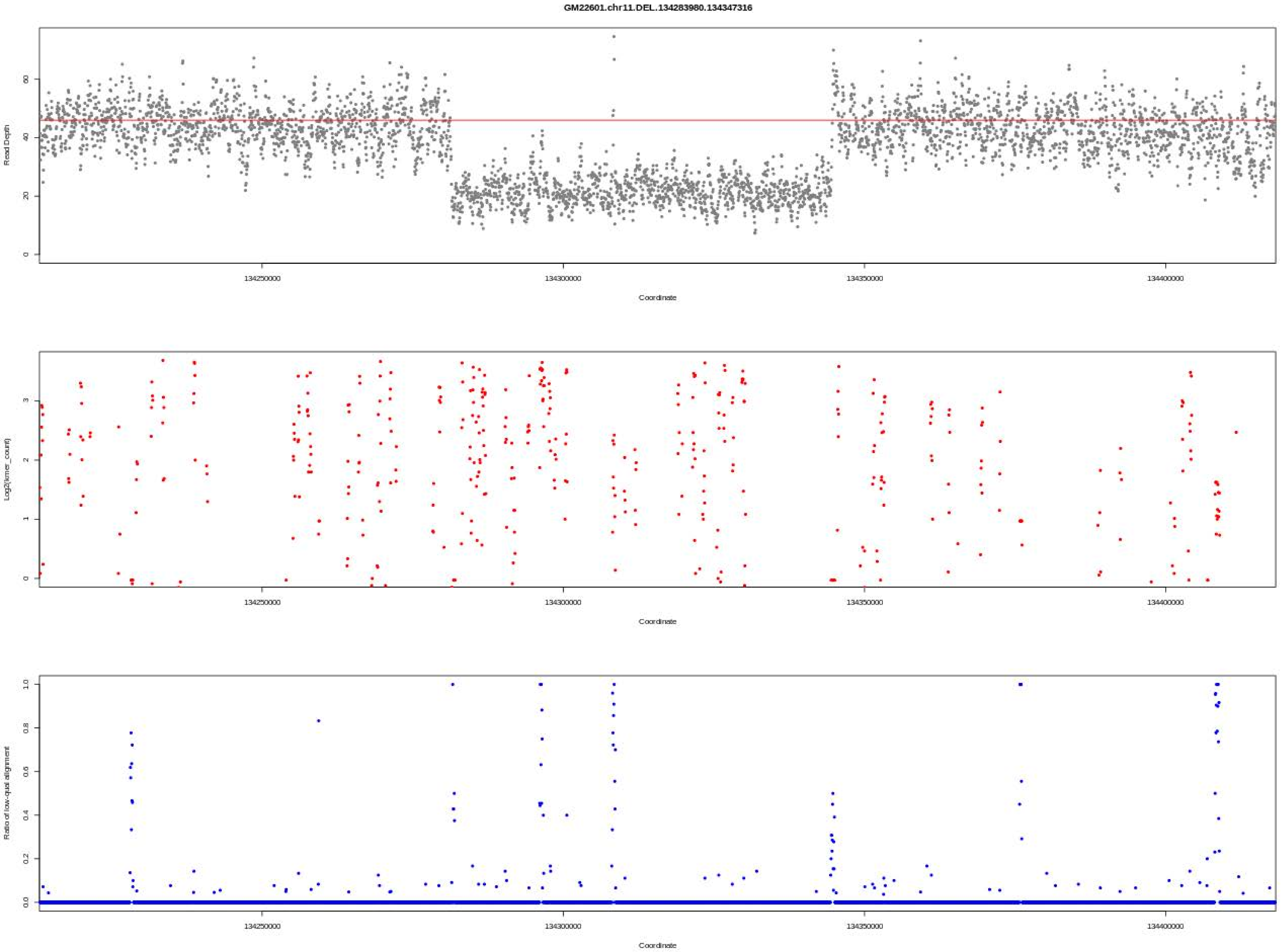
GM22601.chr11.DEL.134283980.134347316
Read Depth
Coordinate
Log2(kmer_count)
Coordinate
Ratio of low-qual alignment
Coordinate
134250000
134300000
134350000
134400000

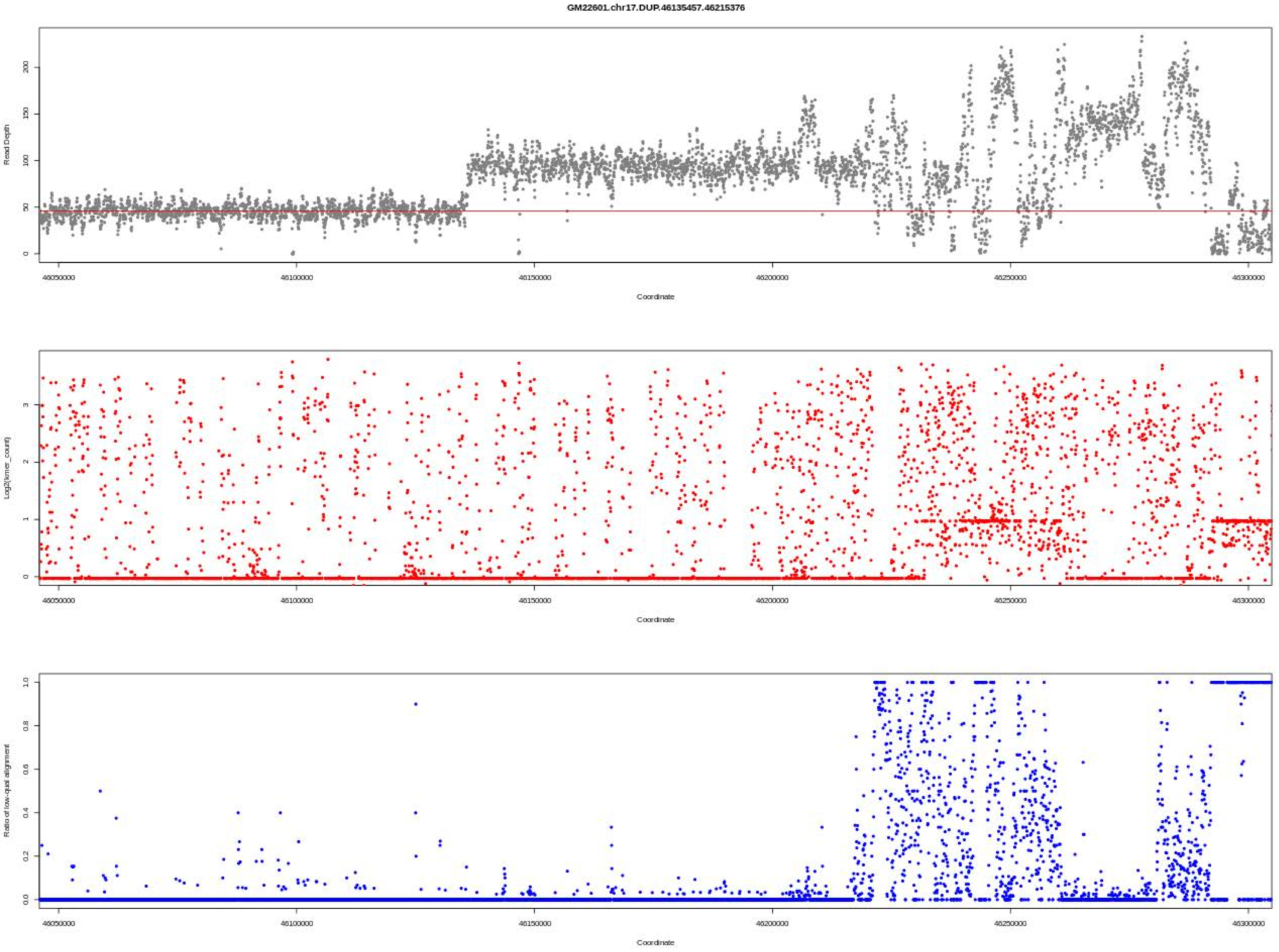
GM22601.chr17.DUP.46135457.46215376
Read Depth
Coordinate
Log2(kmer_count)
Coordinate
Ratio of low-qual alignment
Coordinate

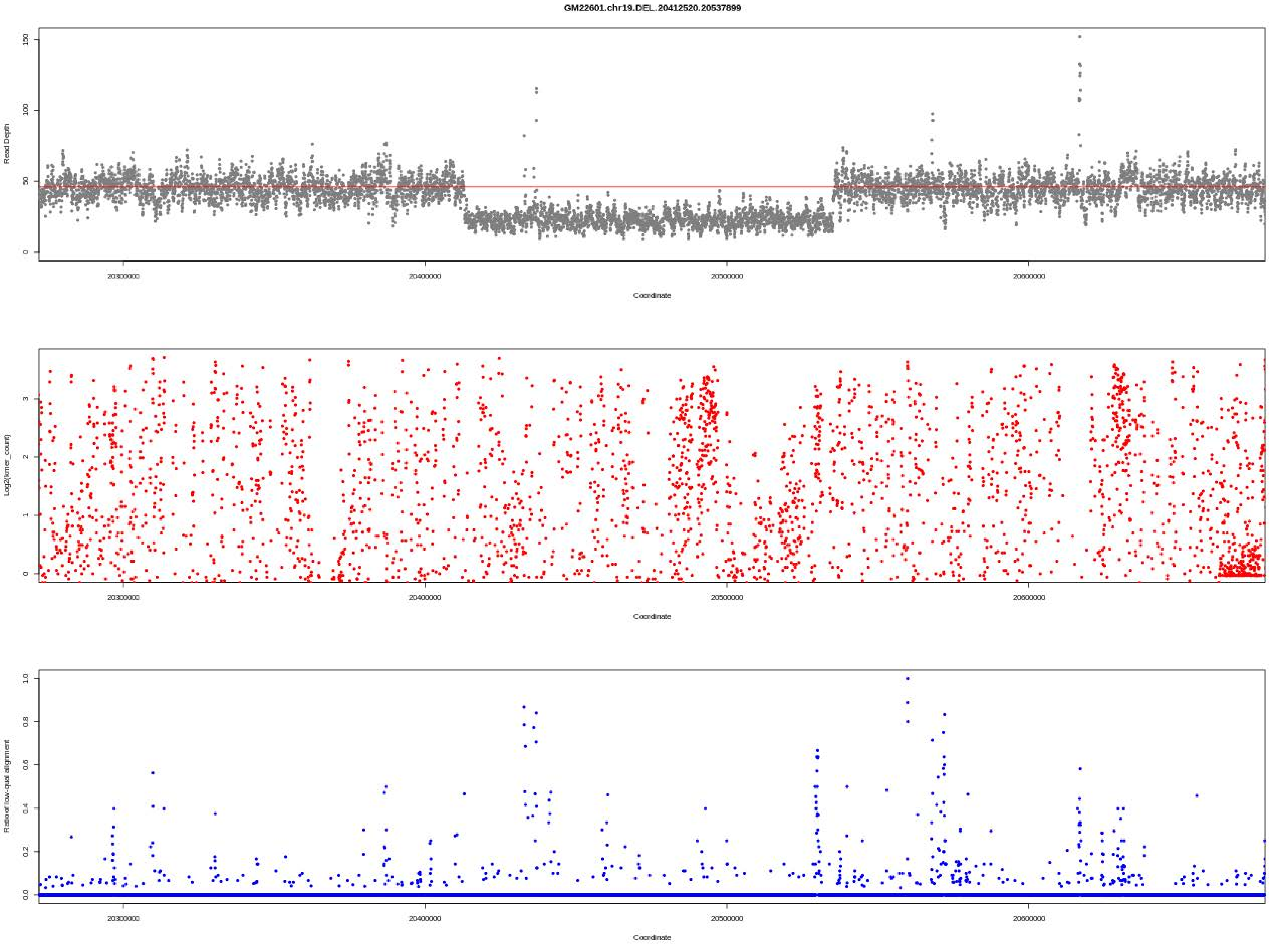
GM22601.chr19.DEL.20412520.20537899
Read Depth
150
100
50
0
20300000
20400000
20500000
20600000
Coordinate
Log2(kmer_count)
3
2
1
0
20300000
20400000
20500000
20600000
Coordinate
Ratio of low-qual alignment
1.0
0.8
0.6
0.4
0.2
0.0
20300000
20400000
20500000
20600000
Coordinate

### Figure S2

31 sub figures of Figure S2 for the 31 samples.

Each sub figure shows reported CNVs, from the inner circle to the outer circle, are from (1) Coriell Institute, (2) JAX-GM CMA, and (3) JAX-CNV on WGS.

Deletions and duplications are colored red and blue, respectively.

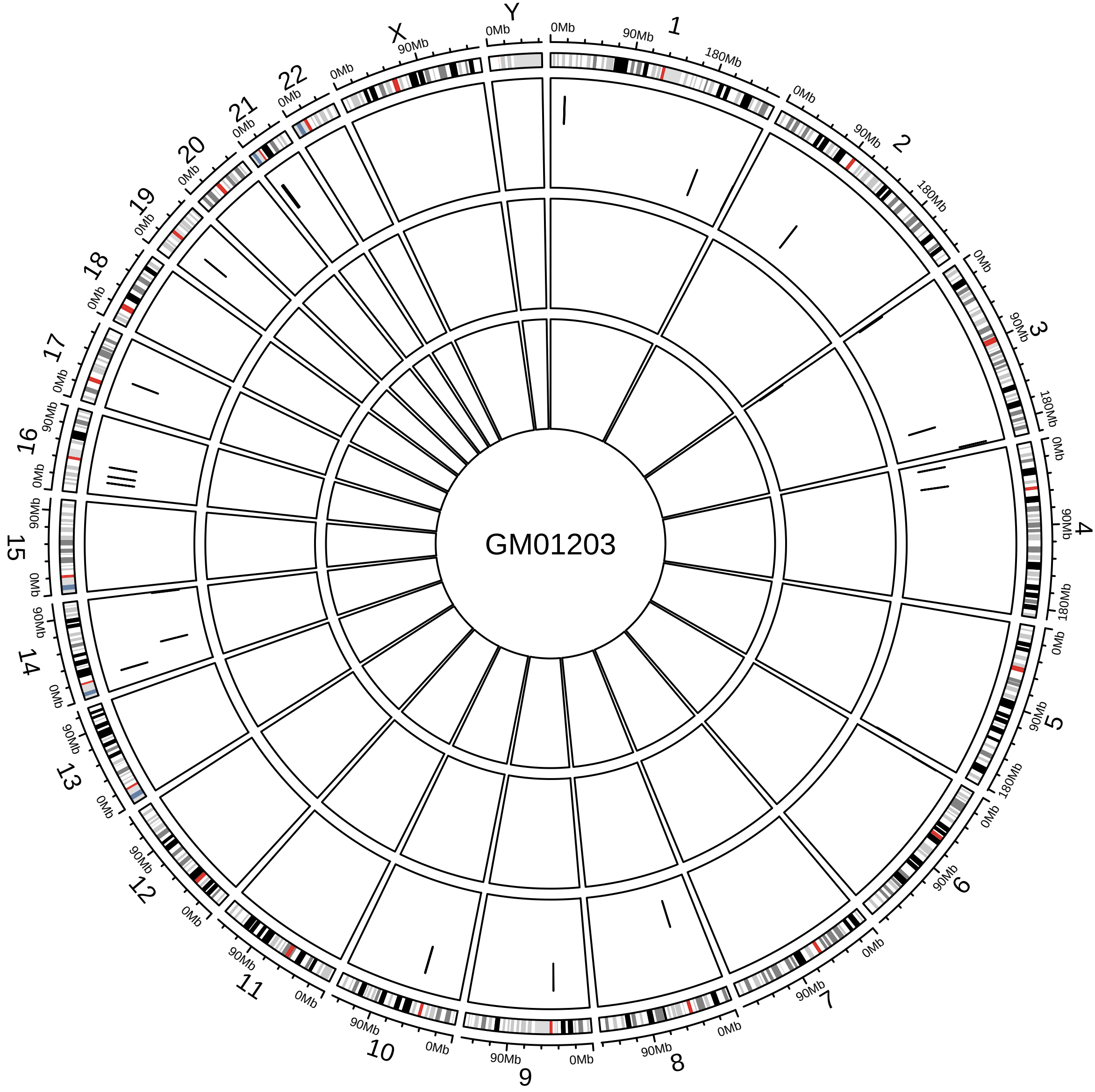
GM01203

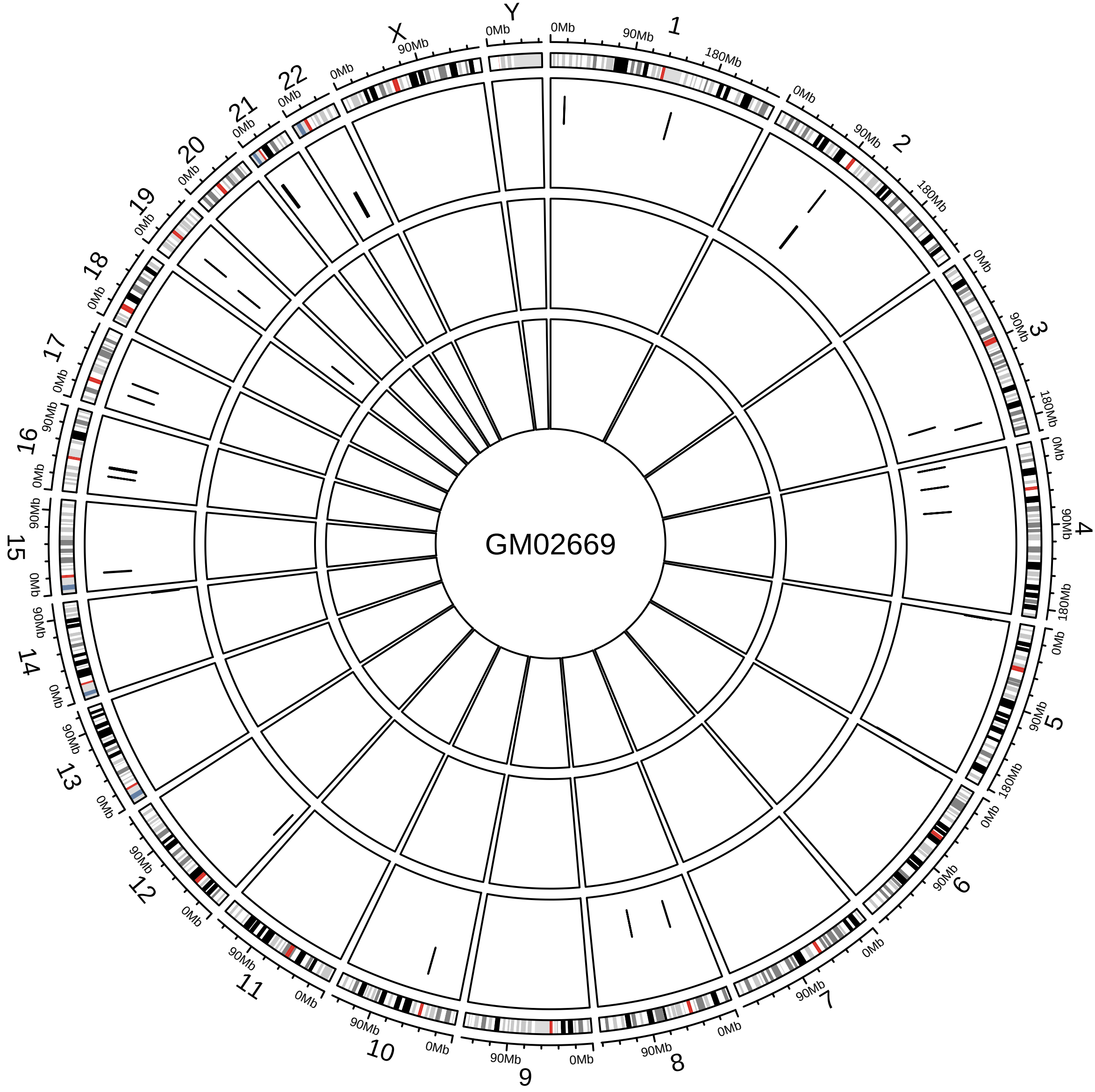

GM02669
X
Y
0Mb
90Mb
180Mb

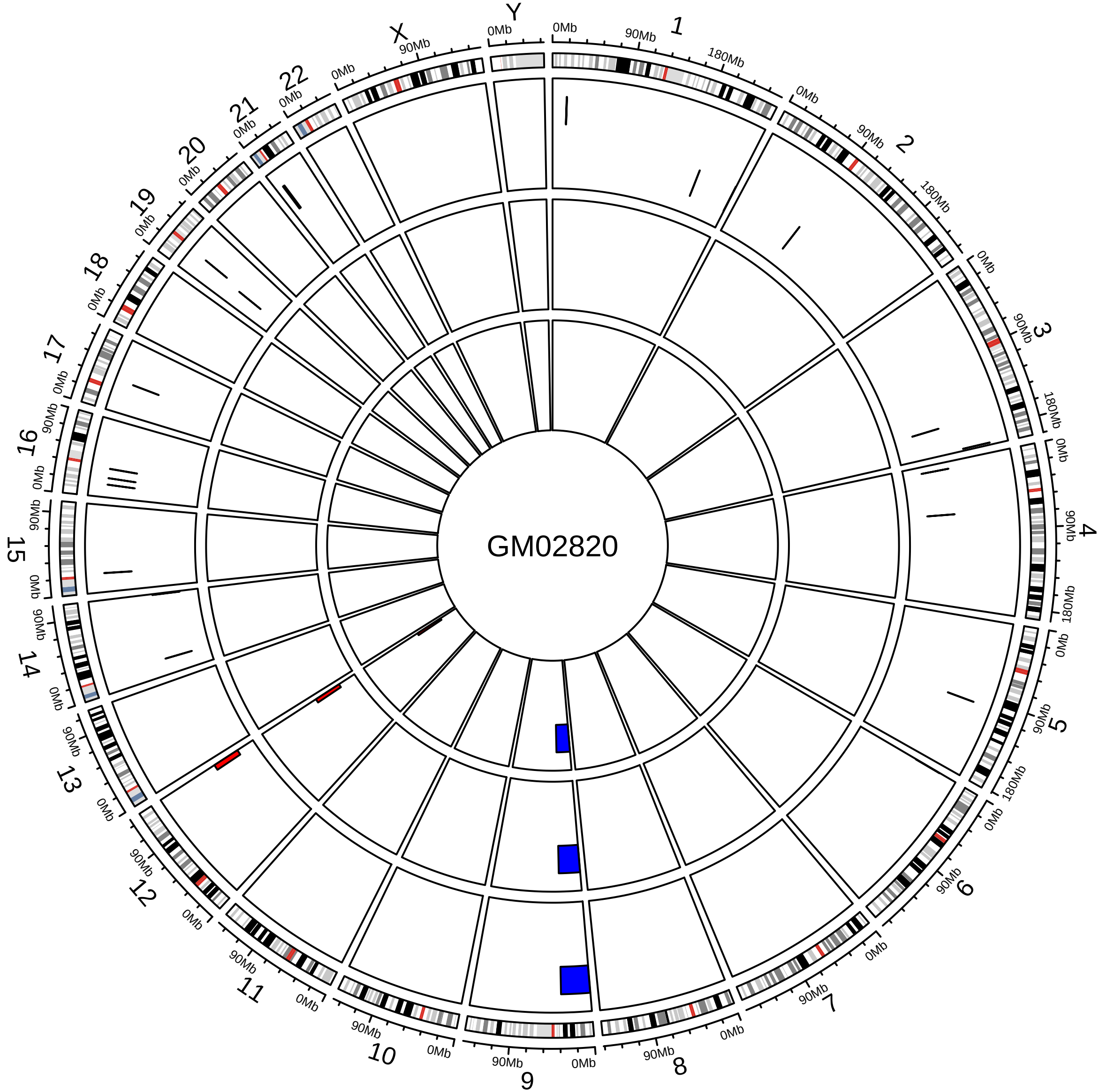

GM02820
X
Y
0Mb
90Mb
180Mb

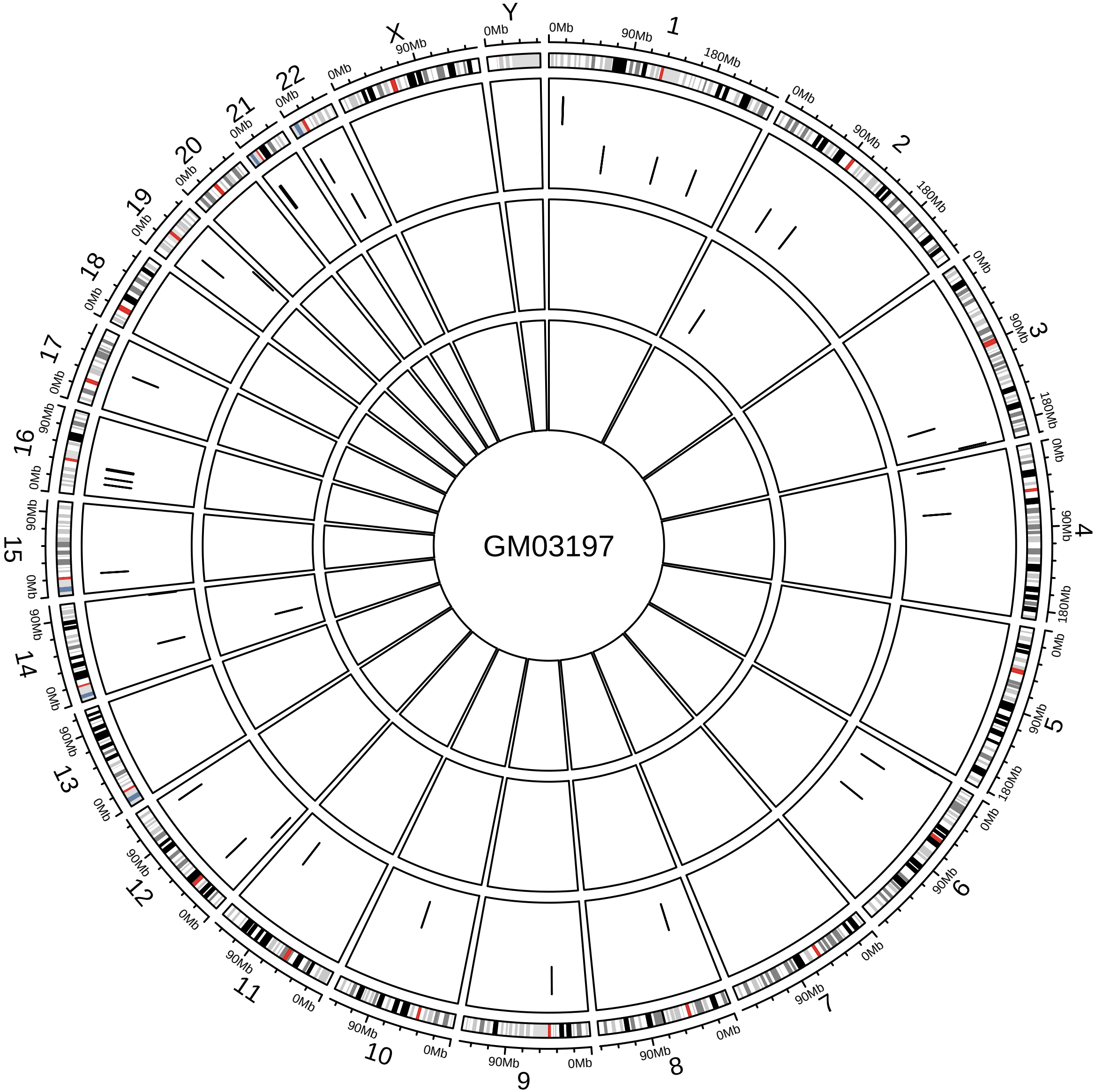

GM03197
X
Y
0Mb
90Mb
180Mb

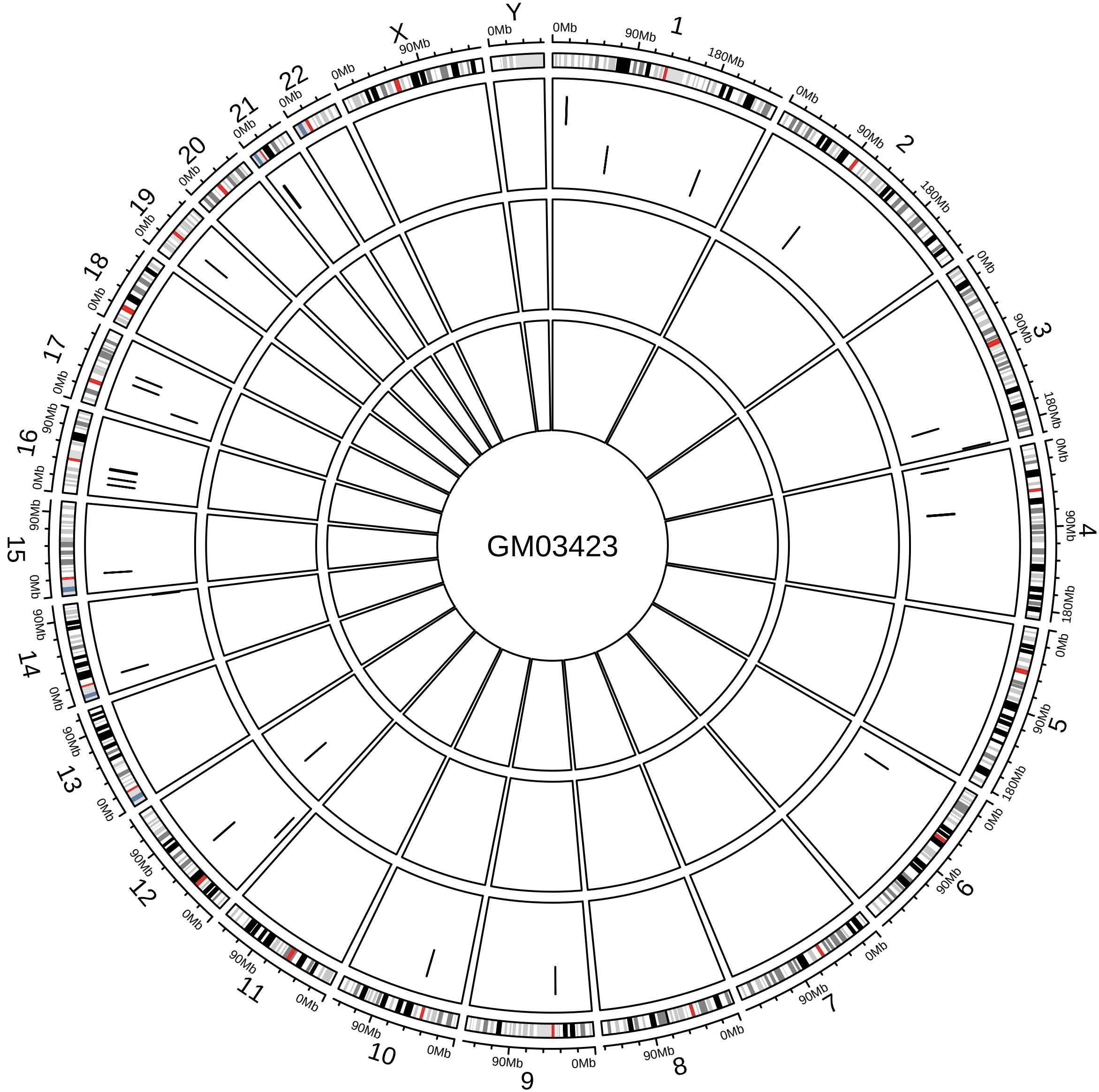
GM03423
X
Y
0Mb
90Mb
180Mb

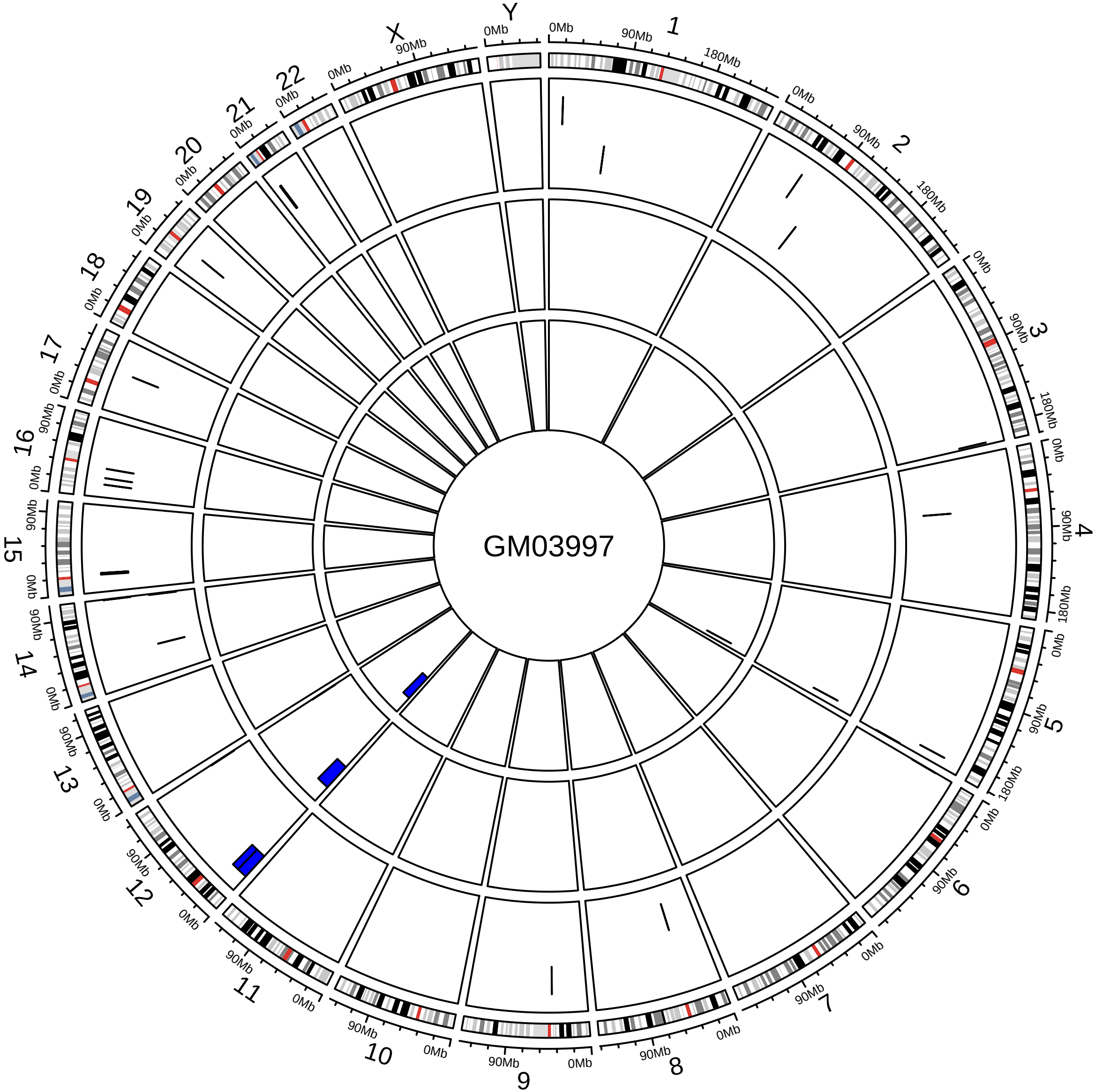

GM03997
X
Y
0Mb
90Mb
180Mb

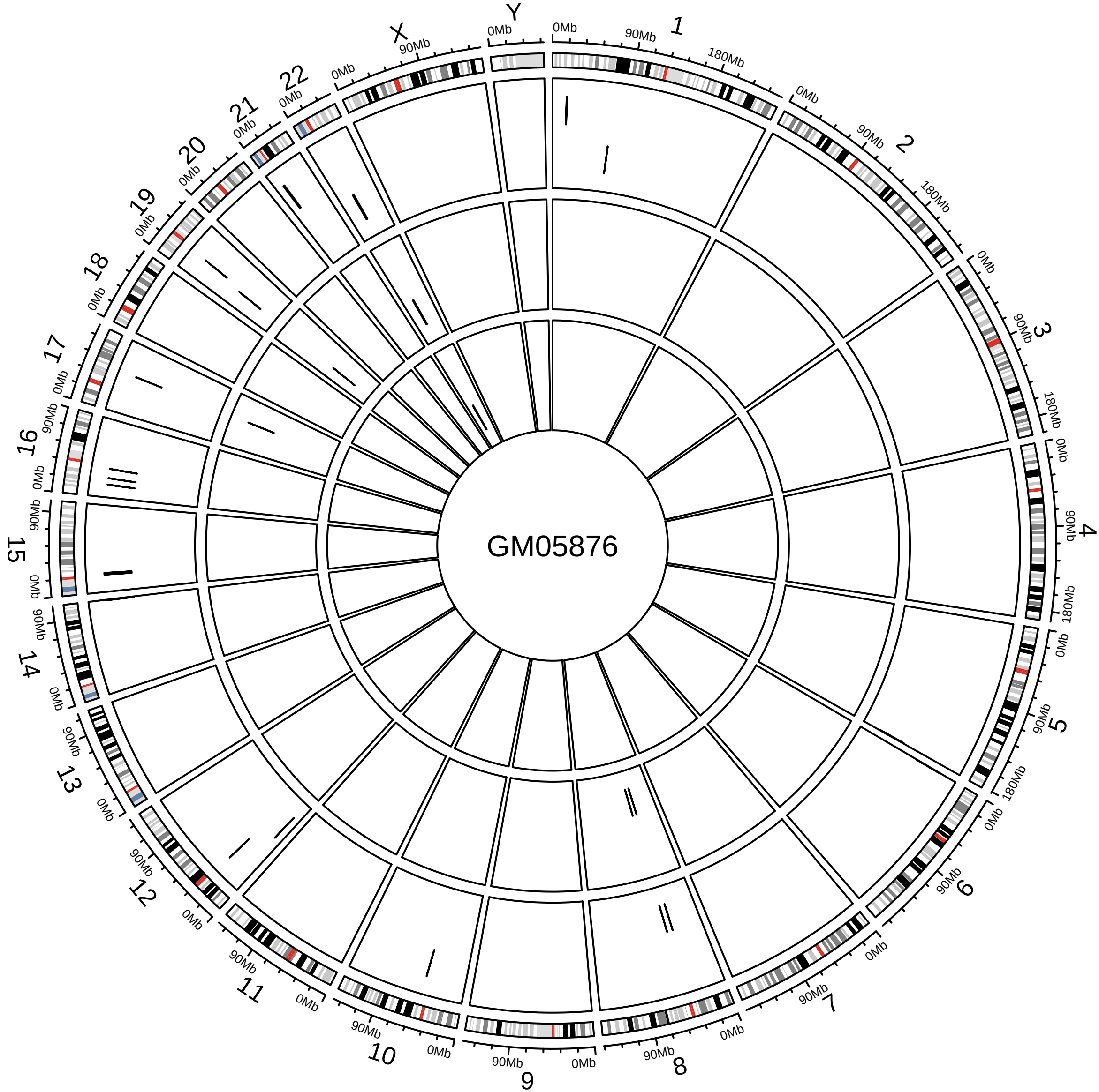
GM05876
X
Y
0Mb
90Mb
180Mb

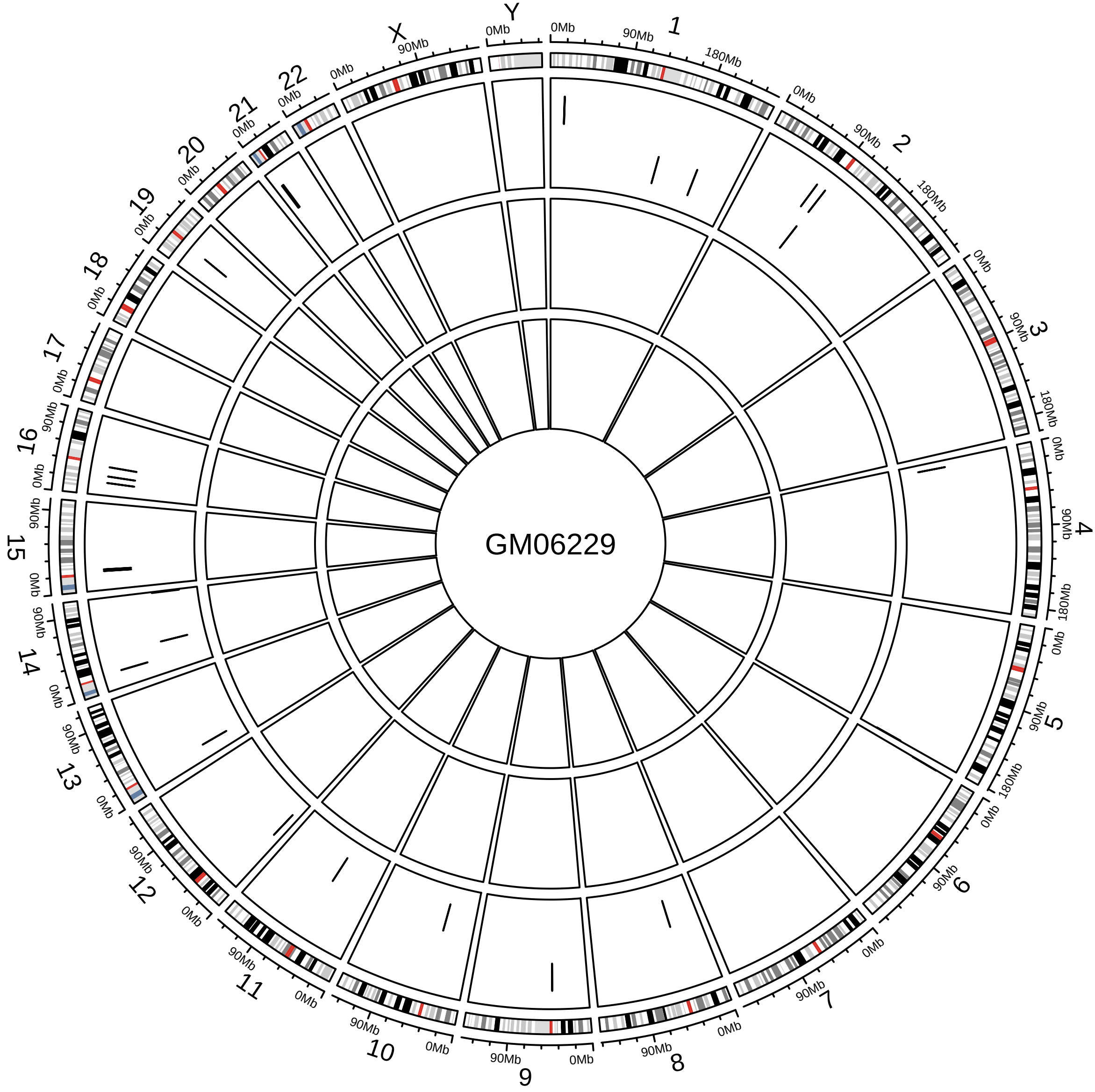
GM06229
X
Y
0Mb
90Mb
180Mb

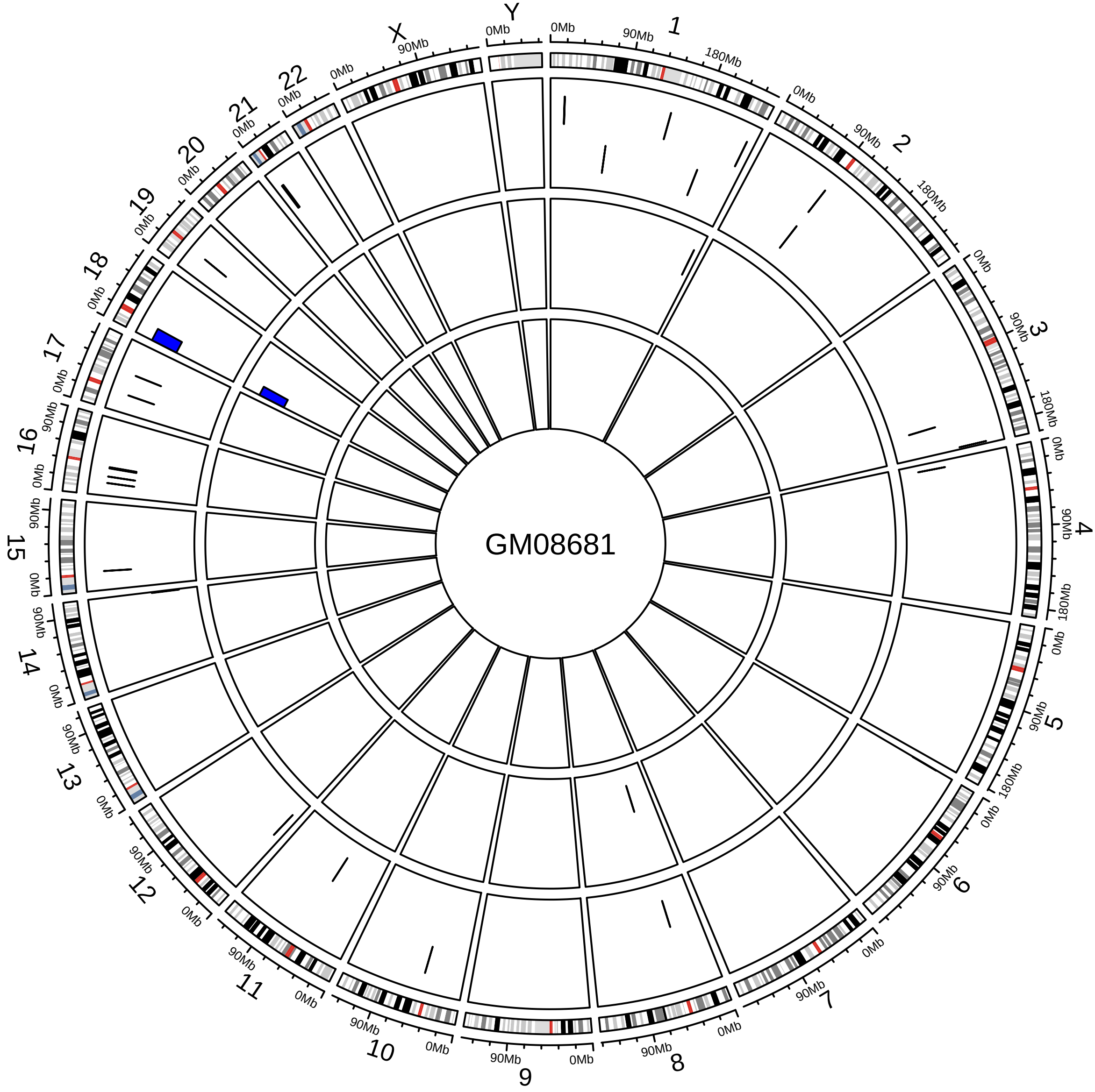
GM08681
X
Y
0Mb
90Mb
180Mb

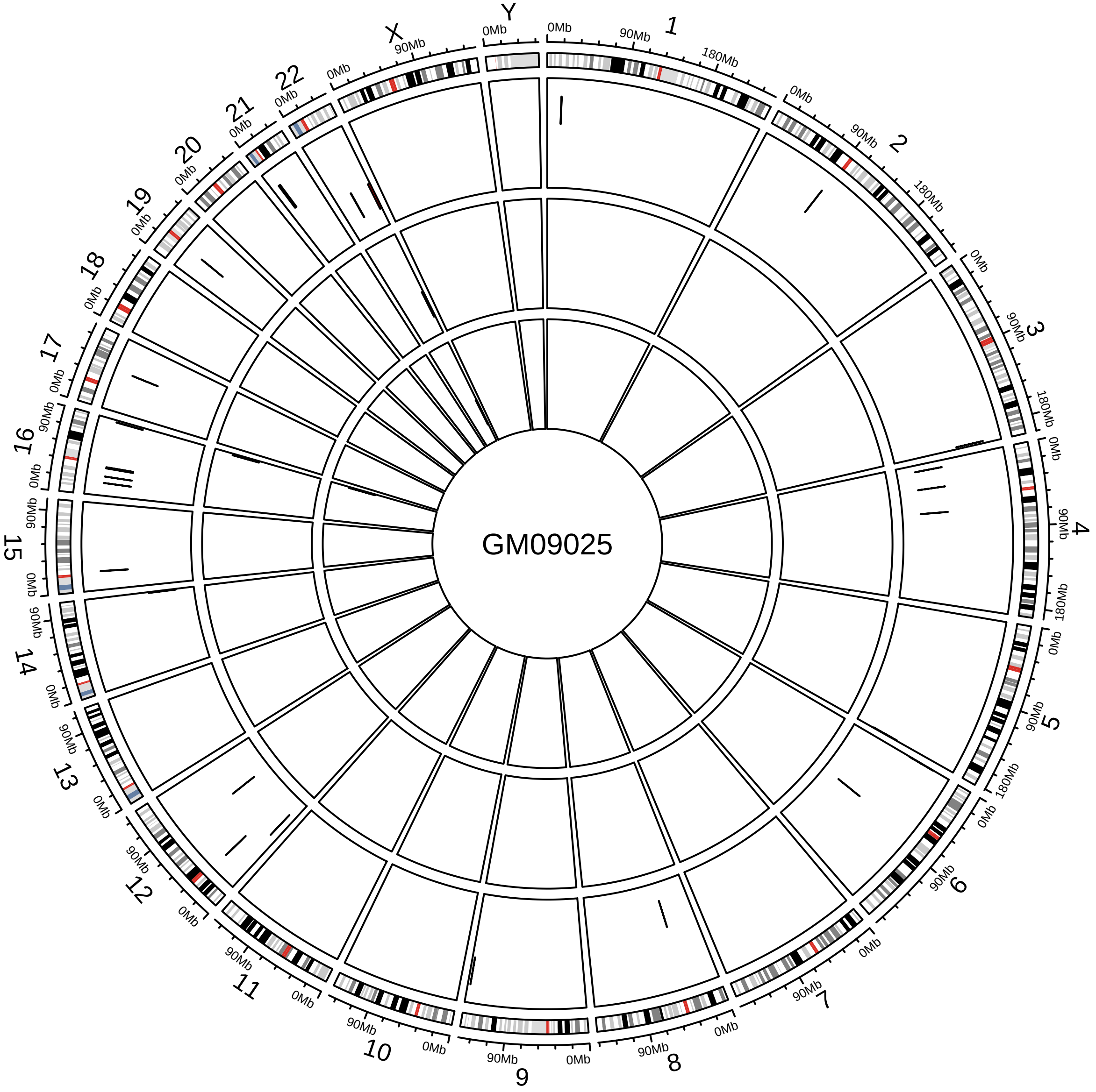
GM09025
X
Y
0Mb
90Mb
180Mb

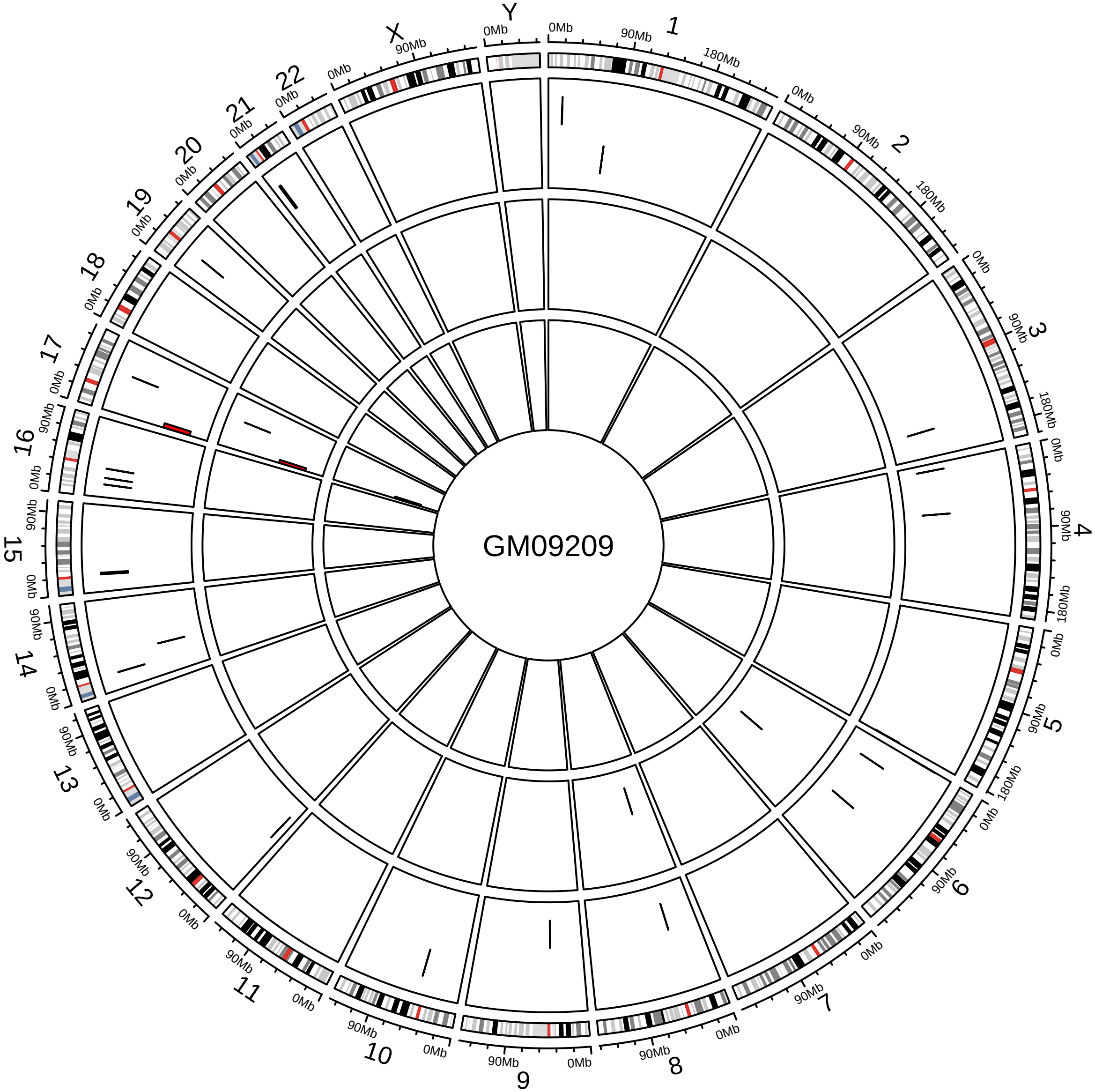
GM09209
X
Y
0Mb
90Mb
180Mb

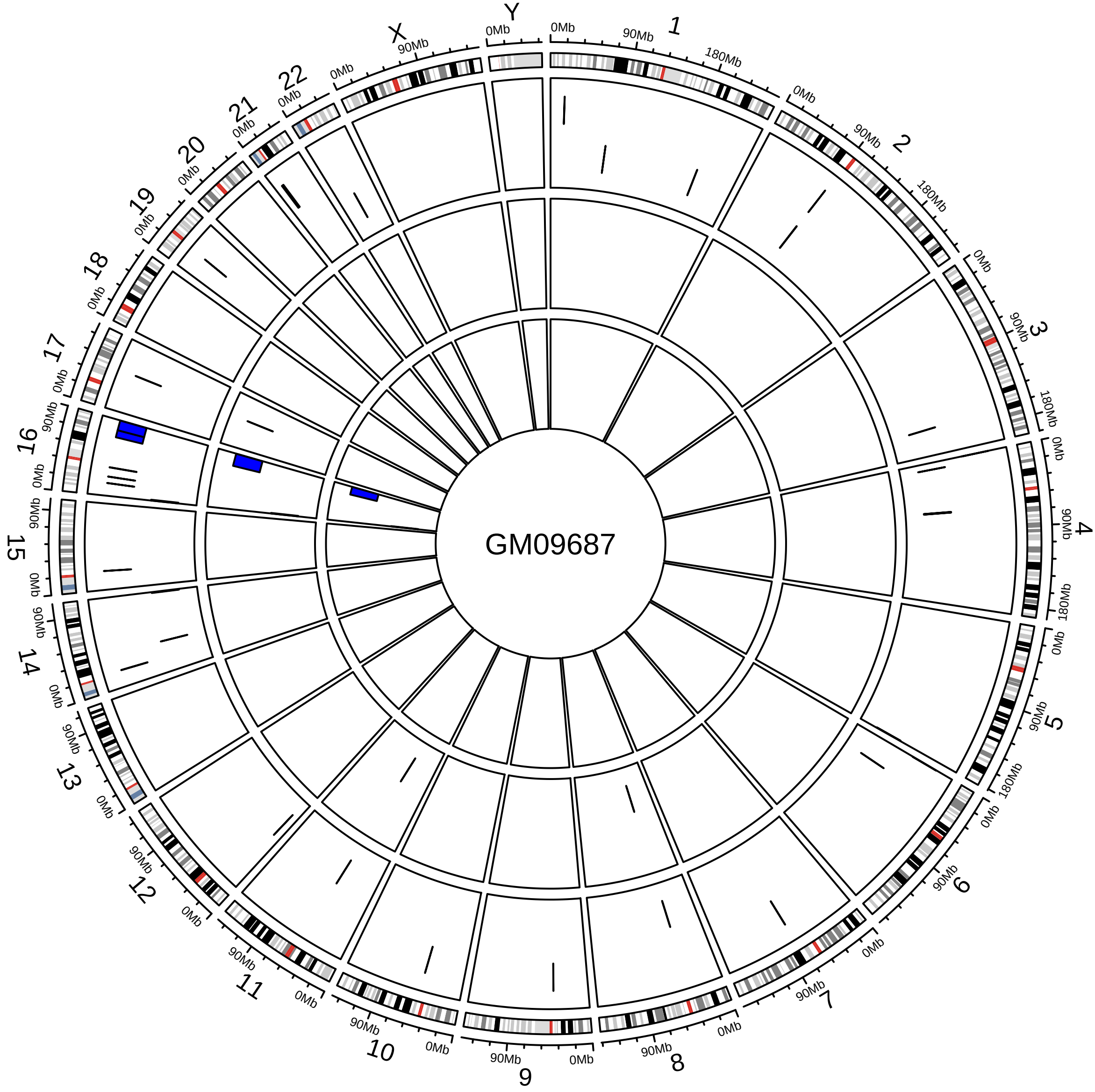
GM09687
X
Y
0Mb
90Mb
180Mb

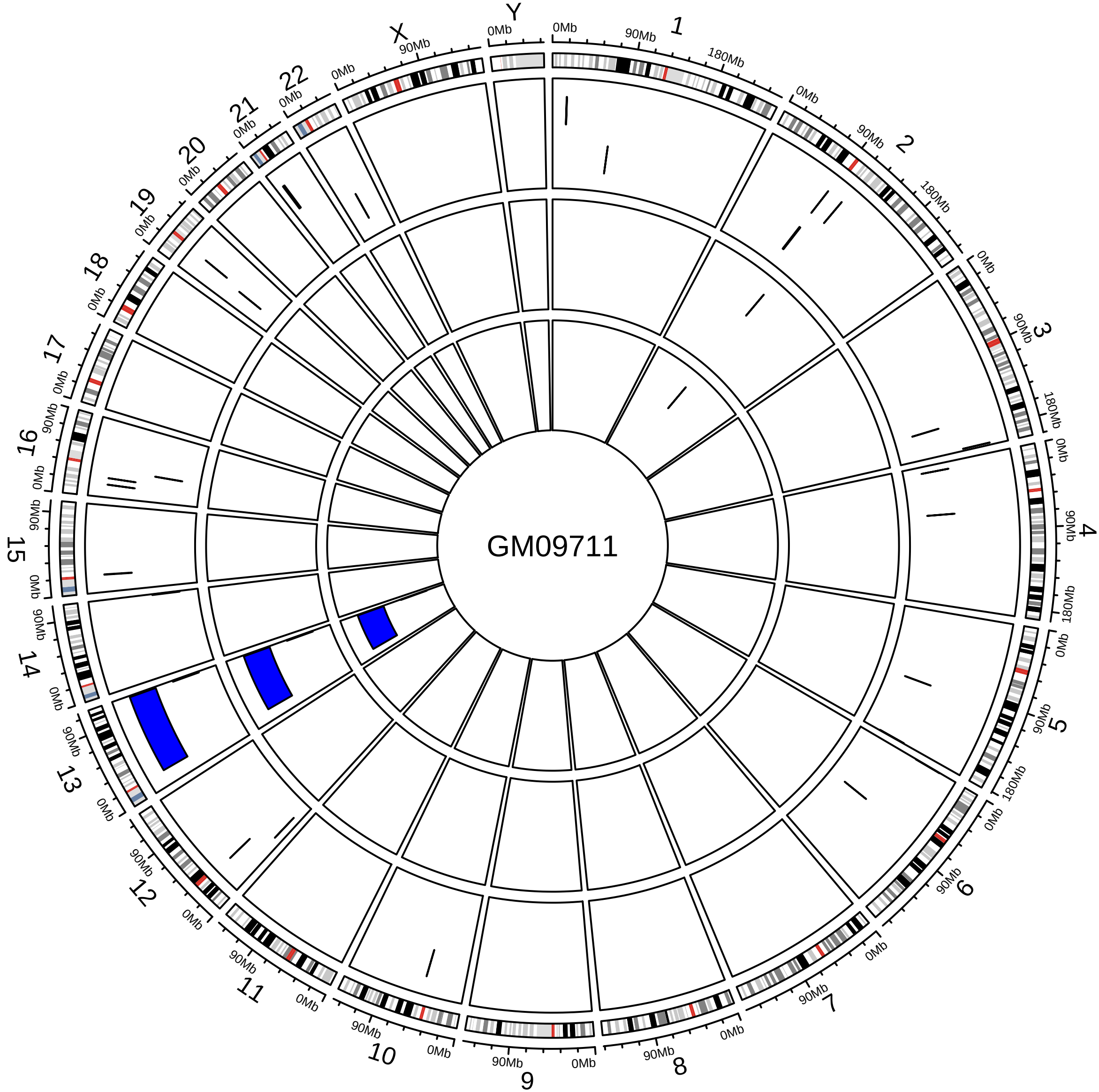
GM09711
X
Y
0Mb
90Mb
180Mb

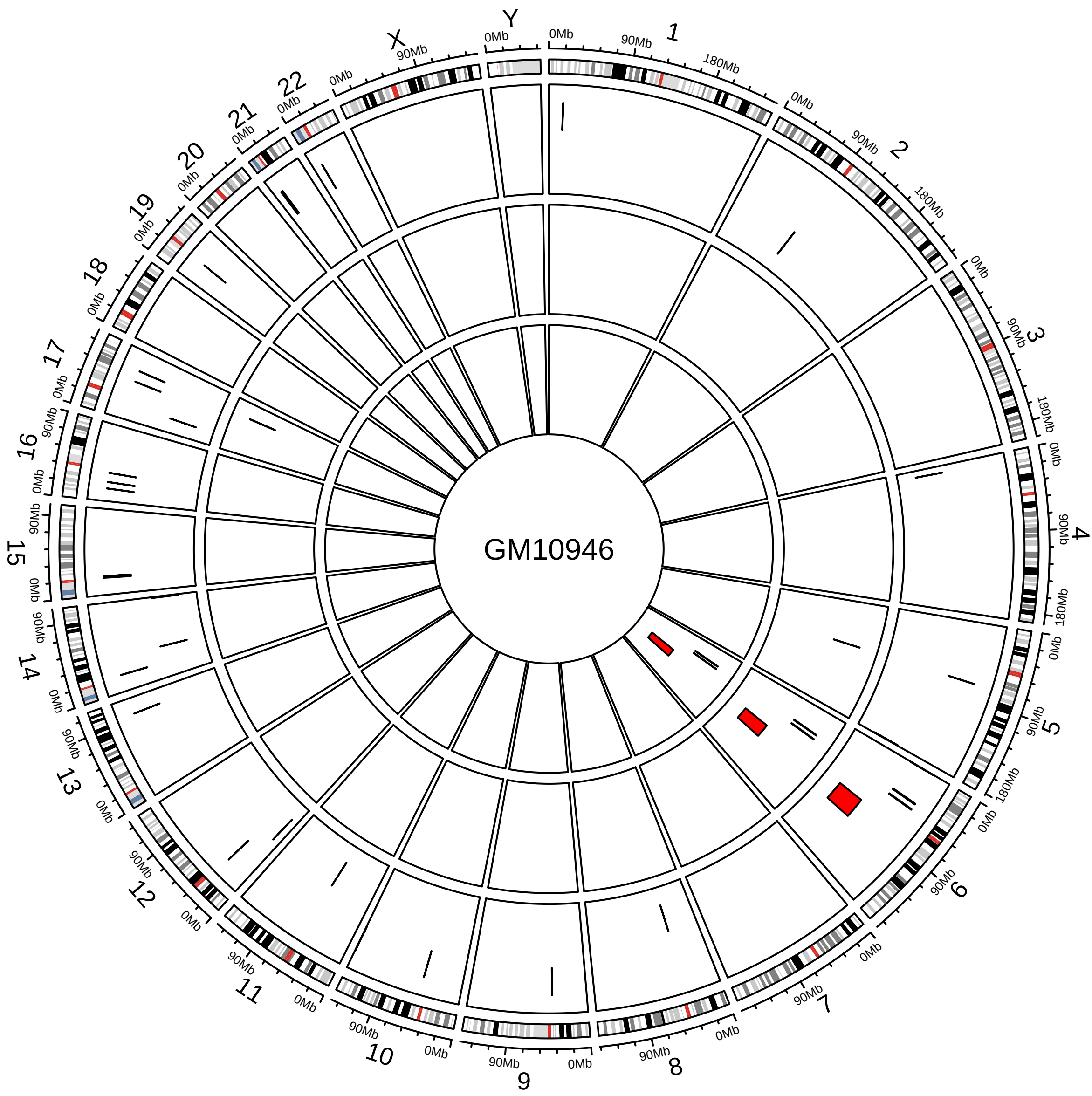
GM10946

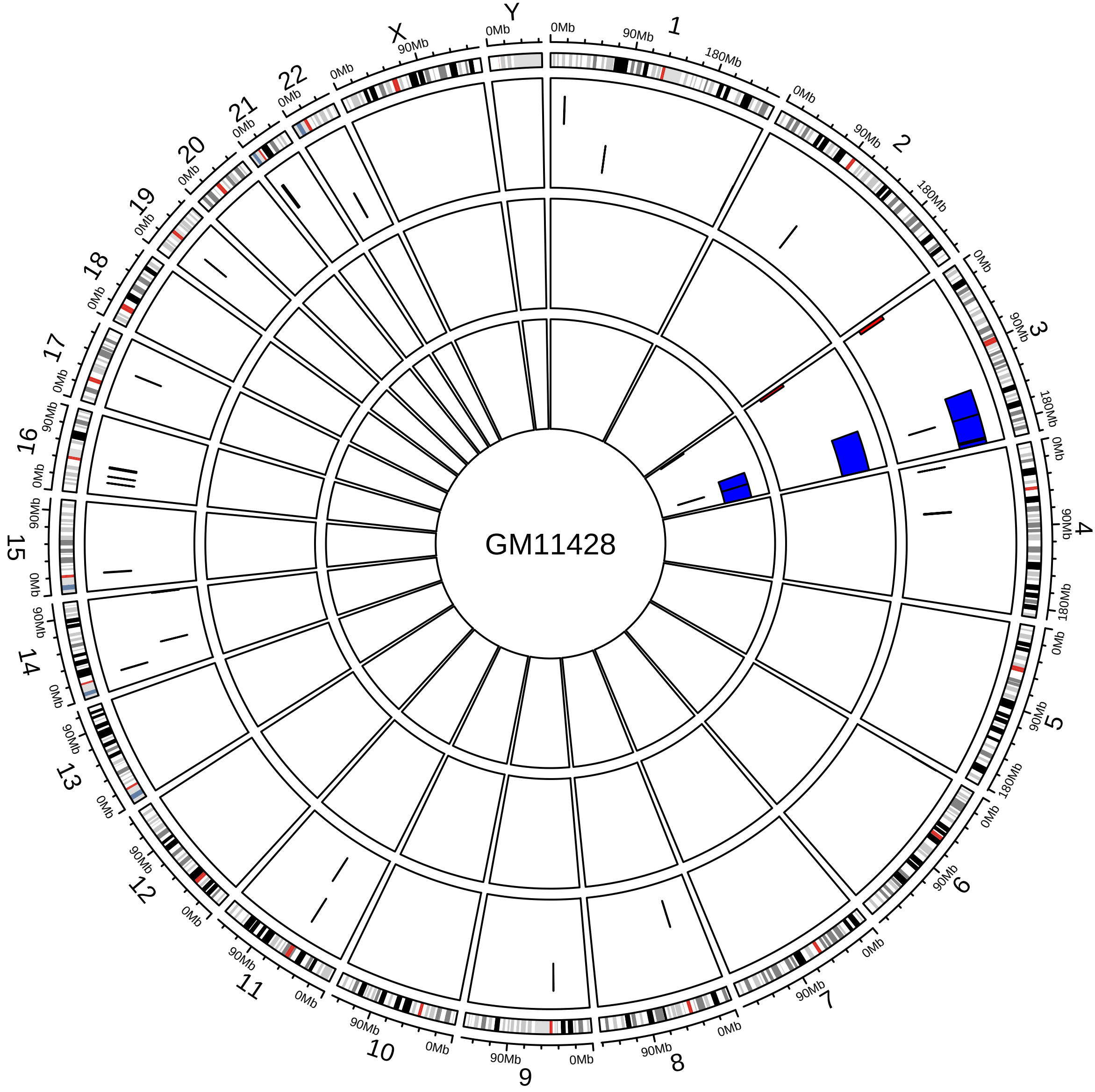
GM11428
X
Y
0Mb
90Mb
180Mb

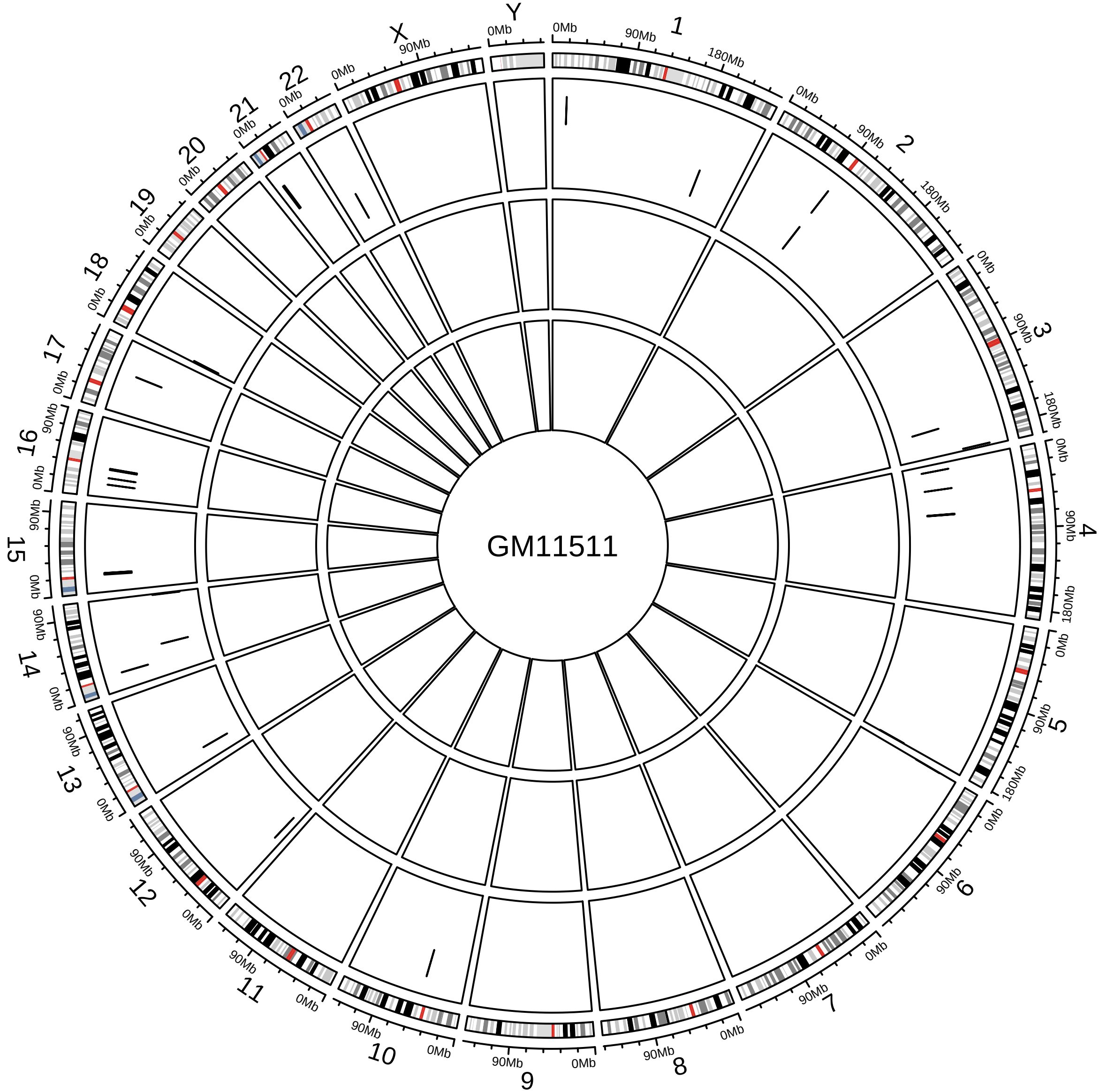
GM11511
X
Y
0Mb
90Mb
180Mb

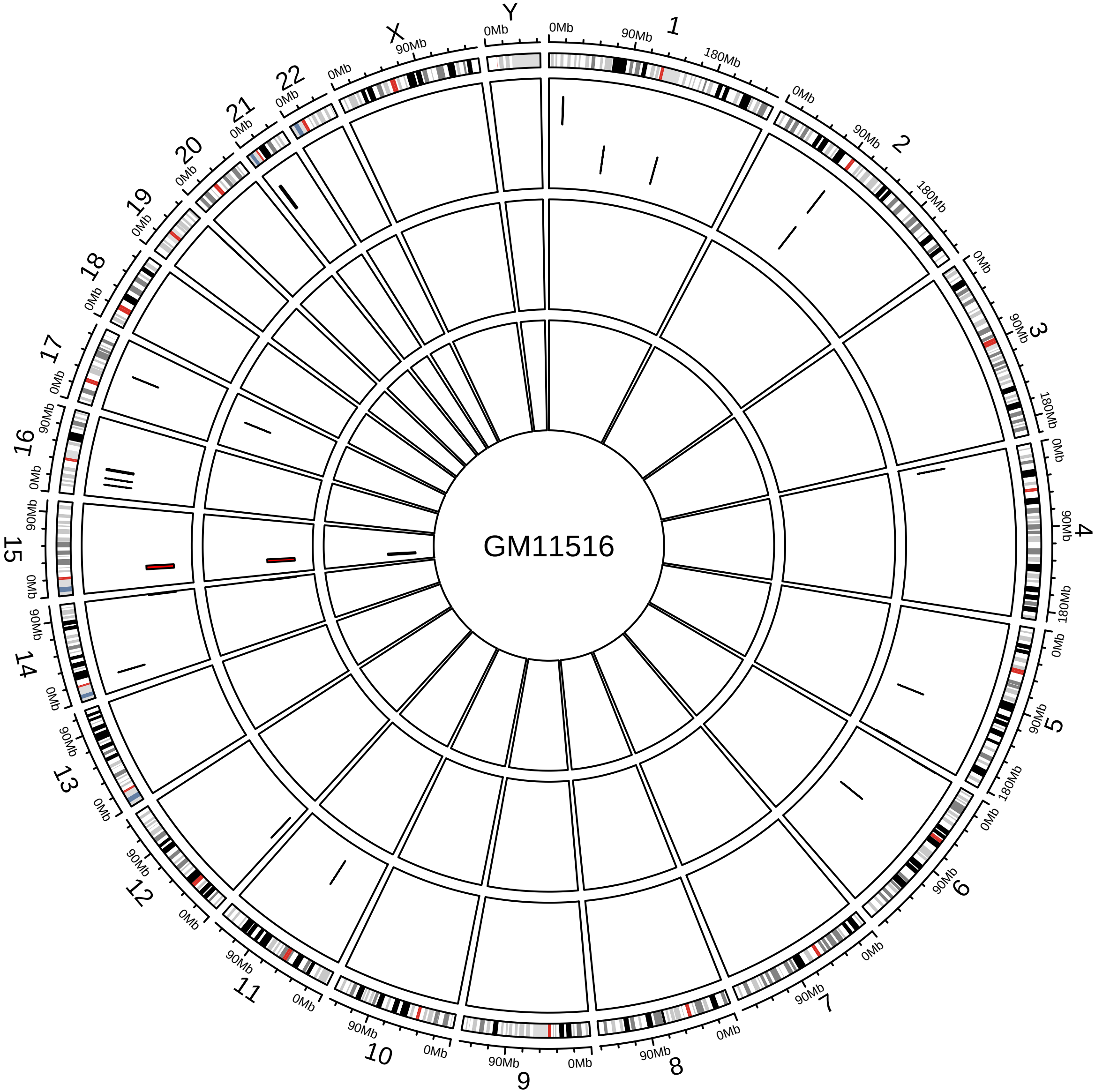
GM11516
X
Y
0Mb
90Mb
180Mb

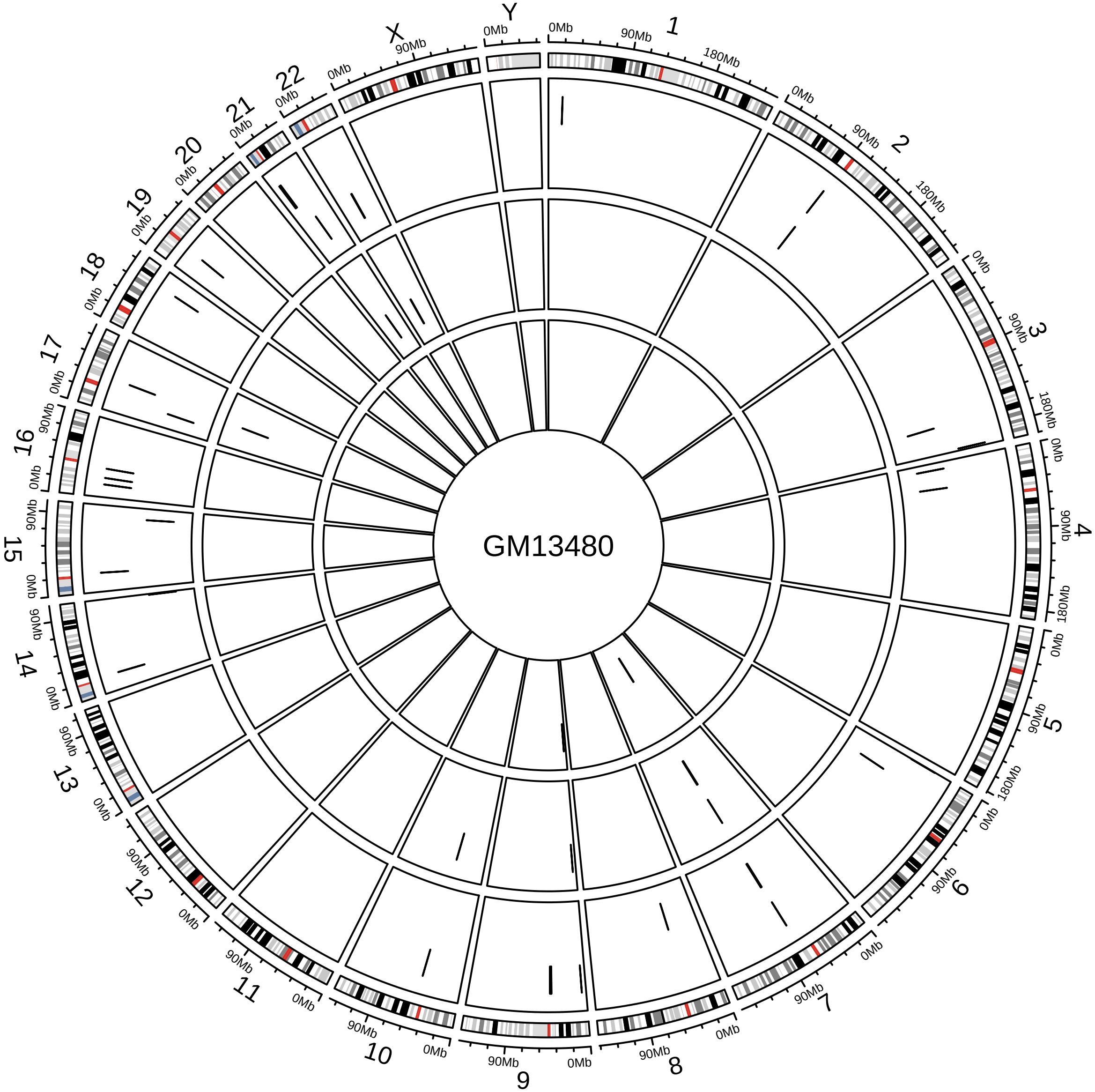
GM13480
X
Y
0Mb
90Mb
180Mb

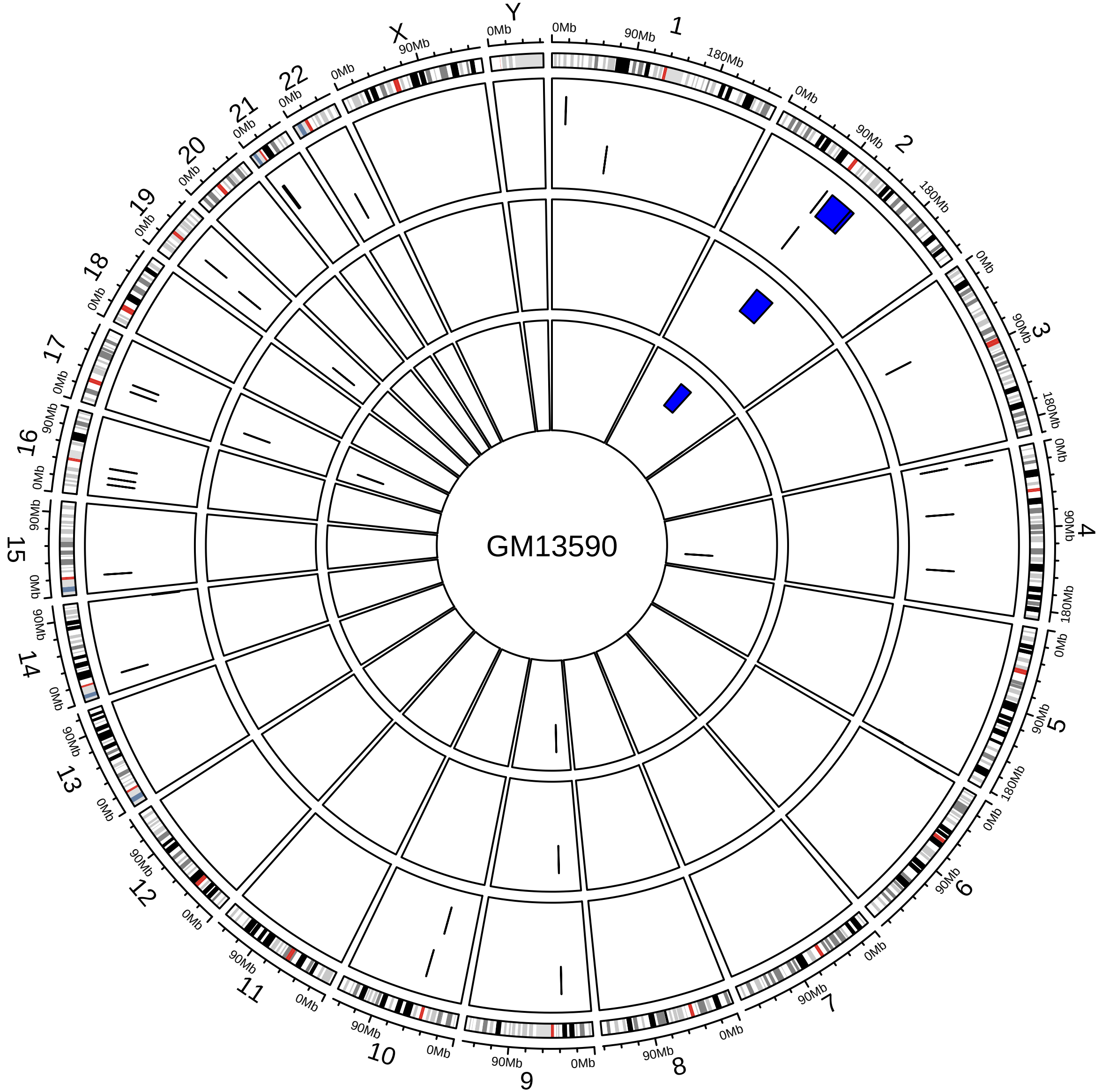
GM13590
X
Y

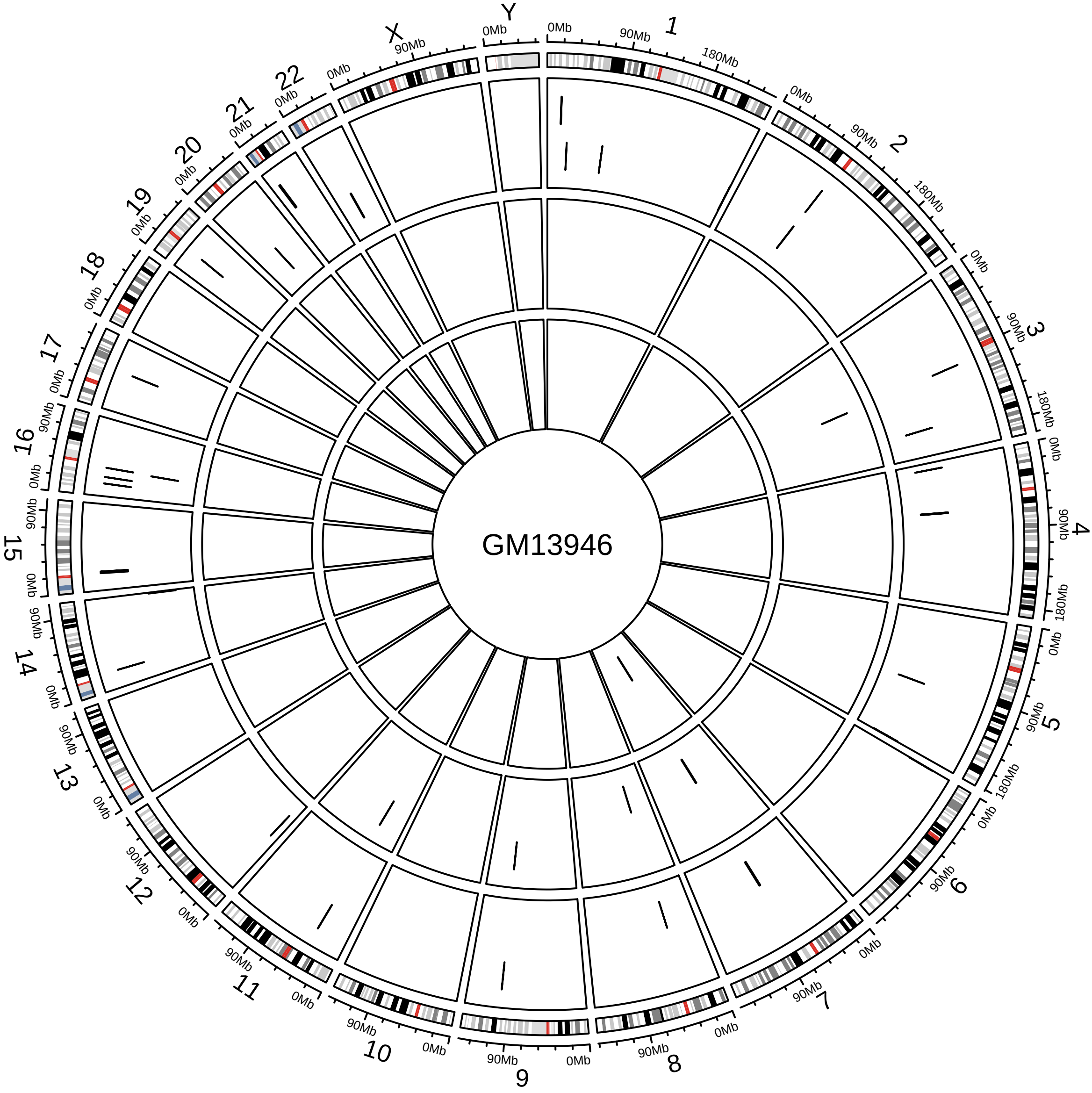

GM13946
X
Y
0Mb
90Mb
180Mb

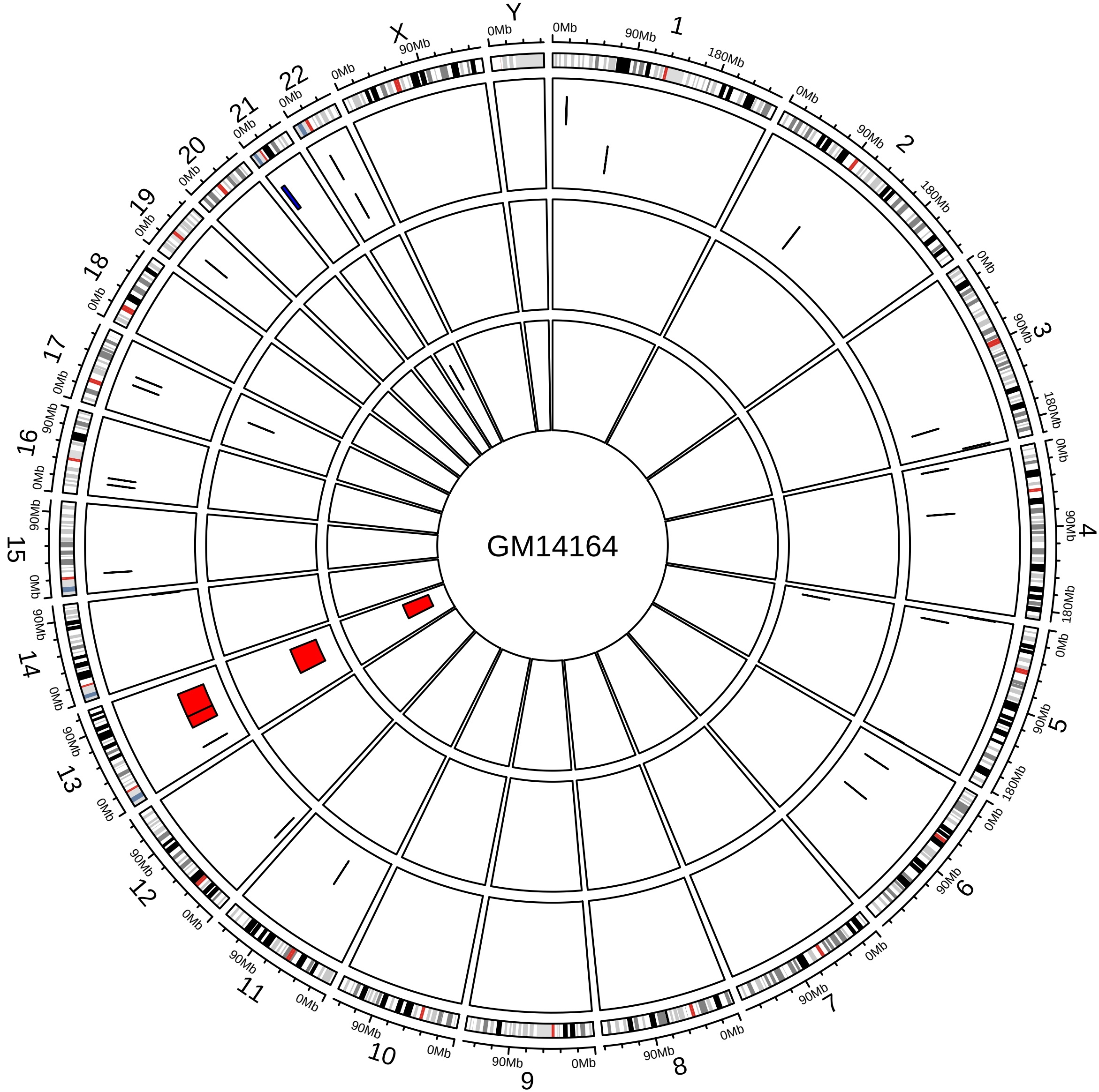

GM14164
X
Y
0Mb
90Mb
180Mb

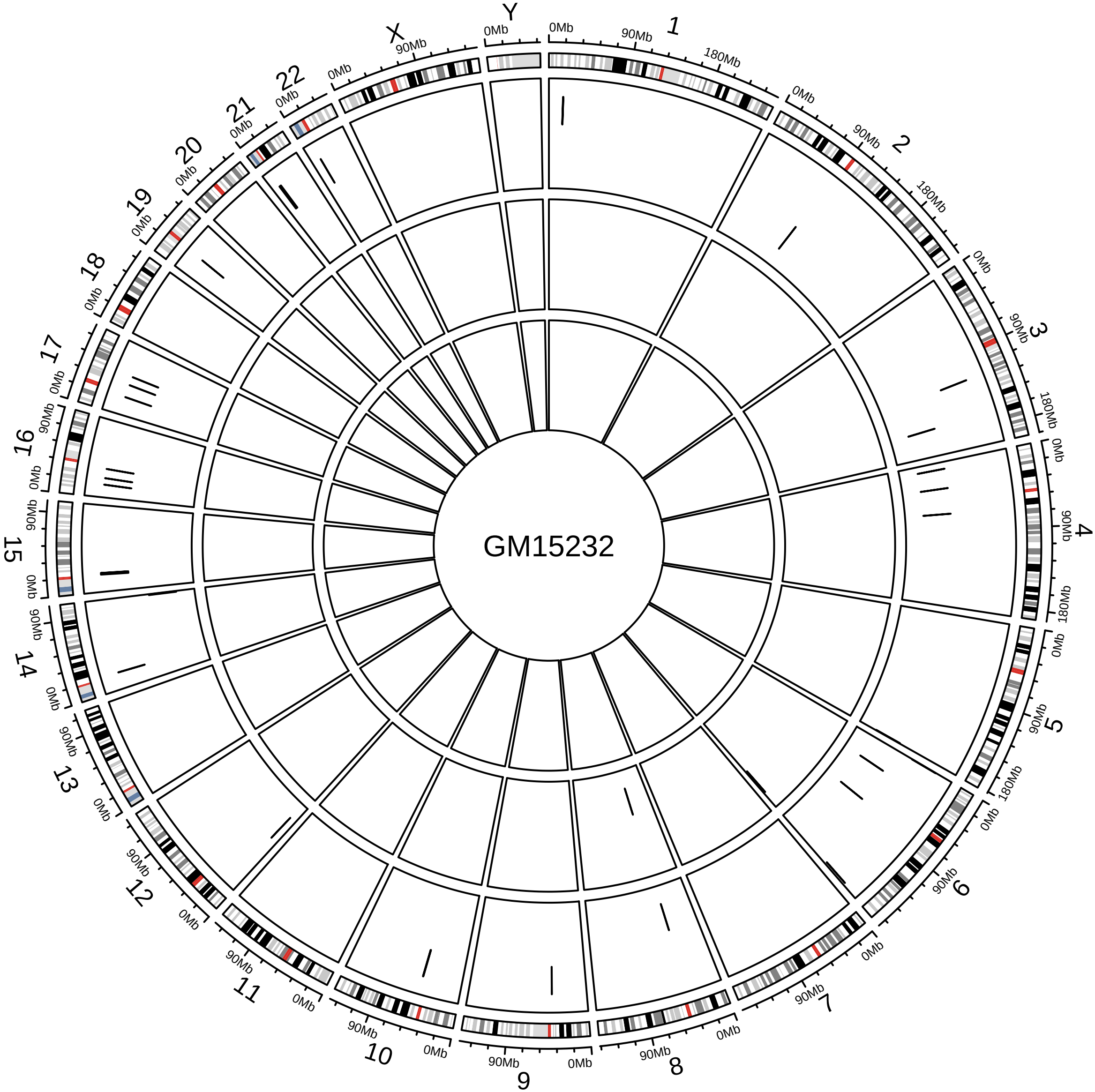
GM15232
X
Y
0Mb
90Mb
180Mb

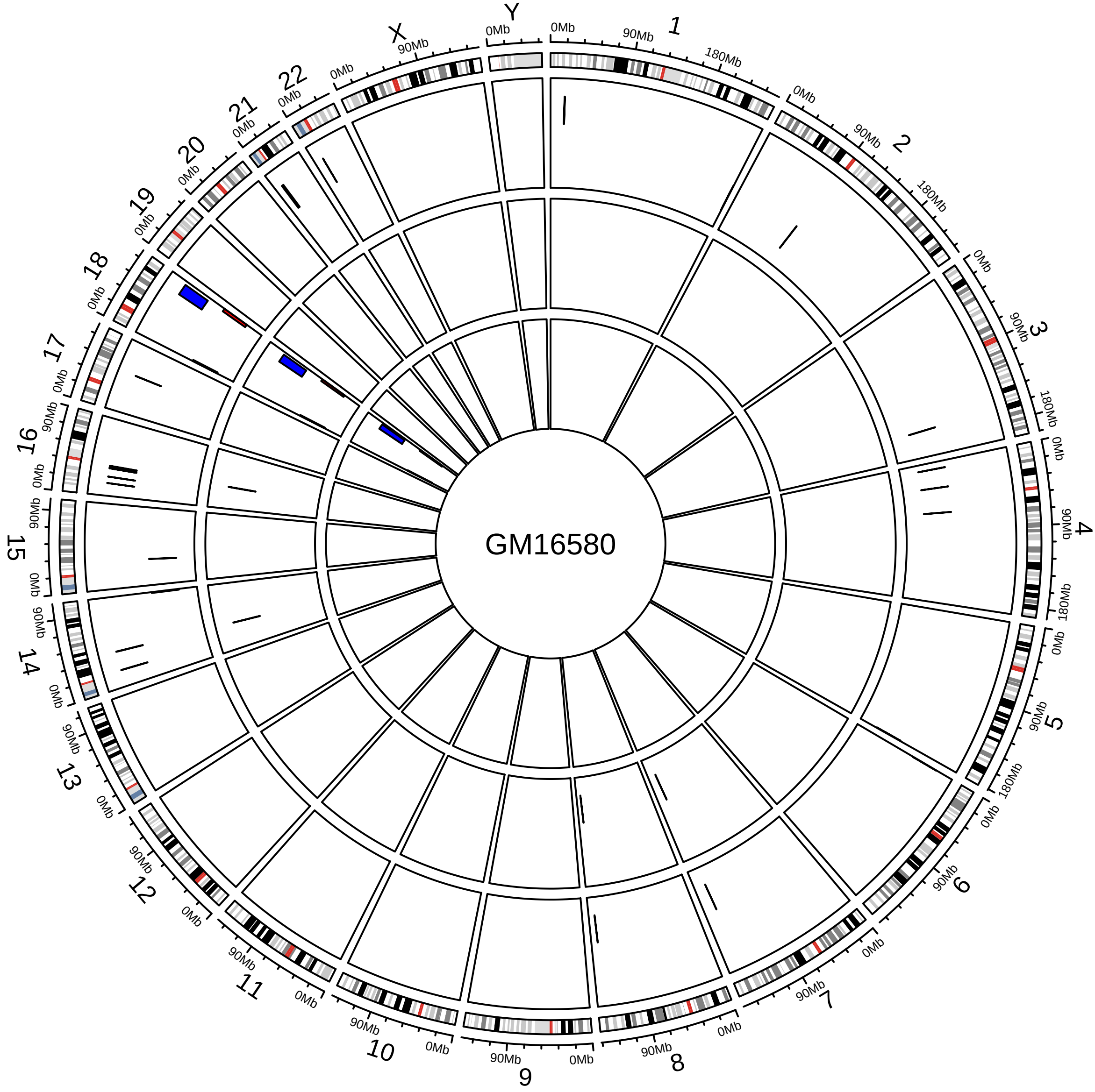

GM16580

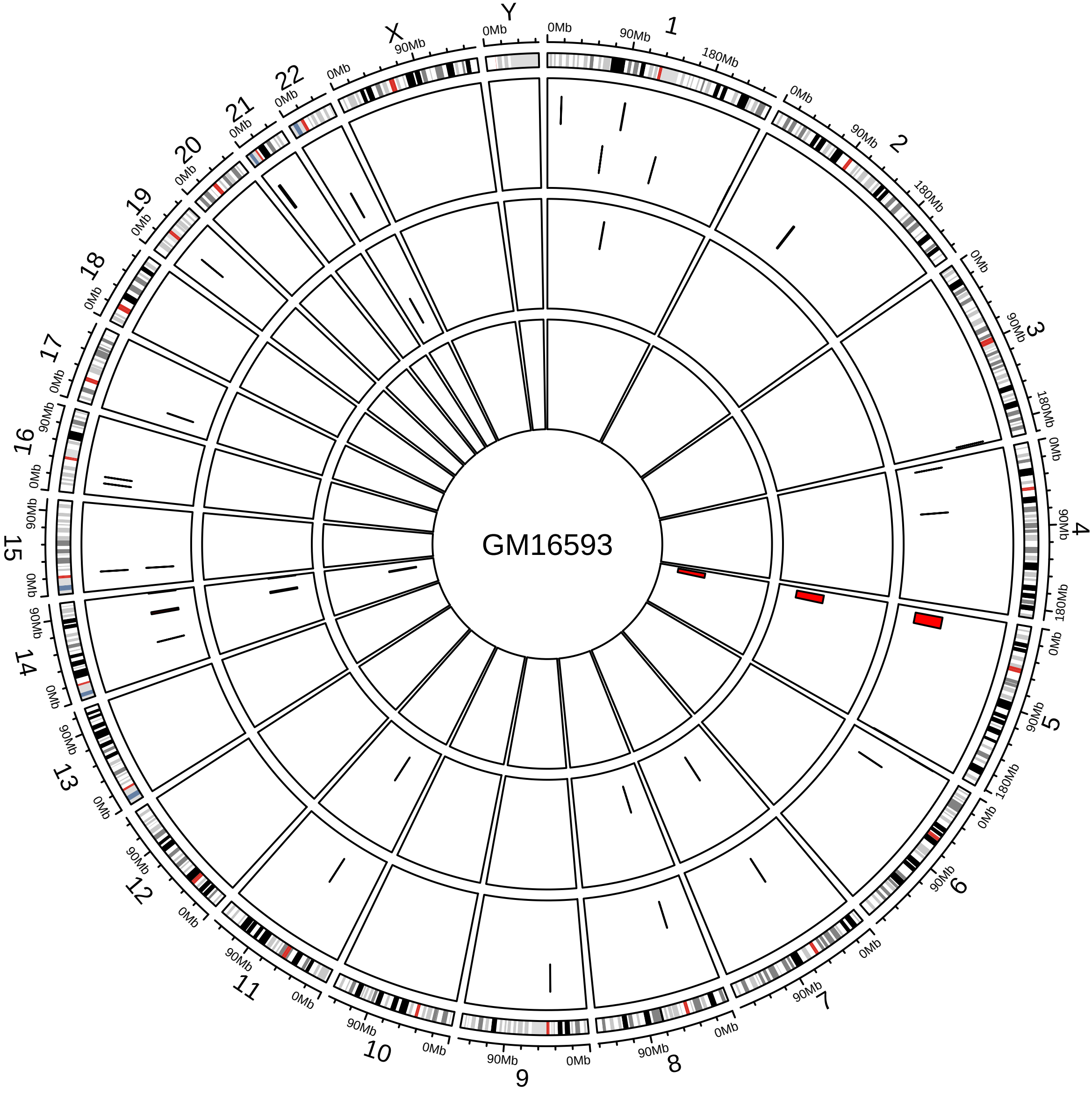
GM16593

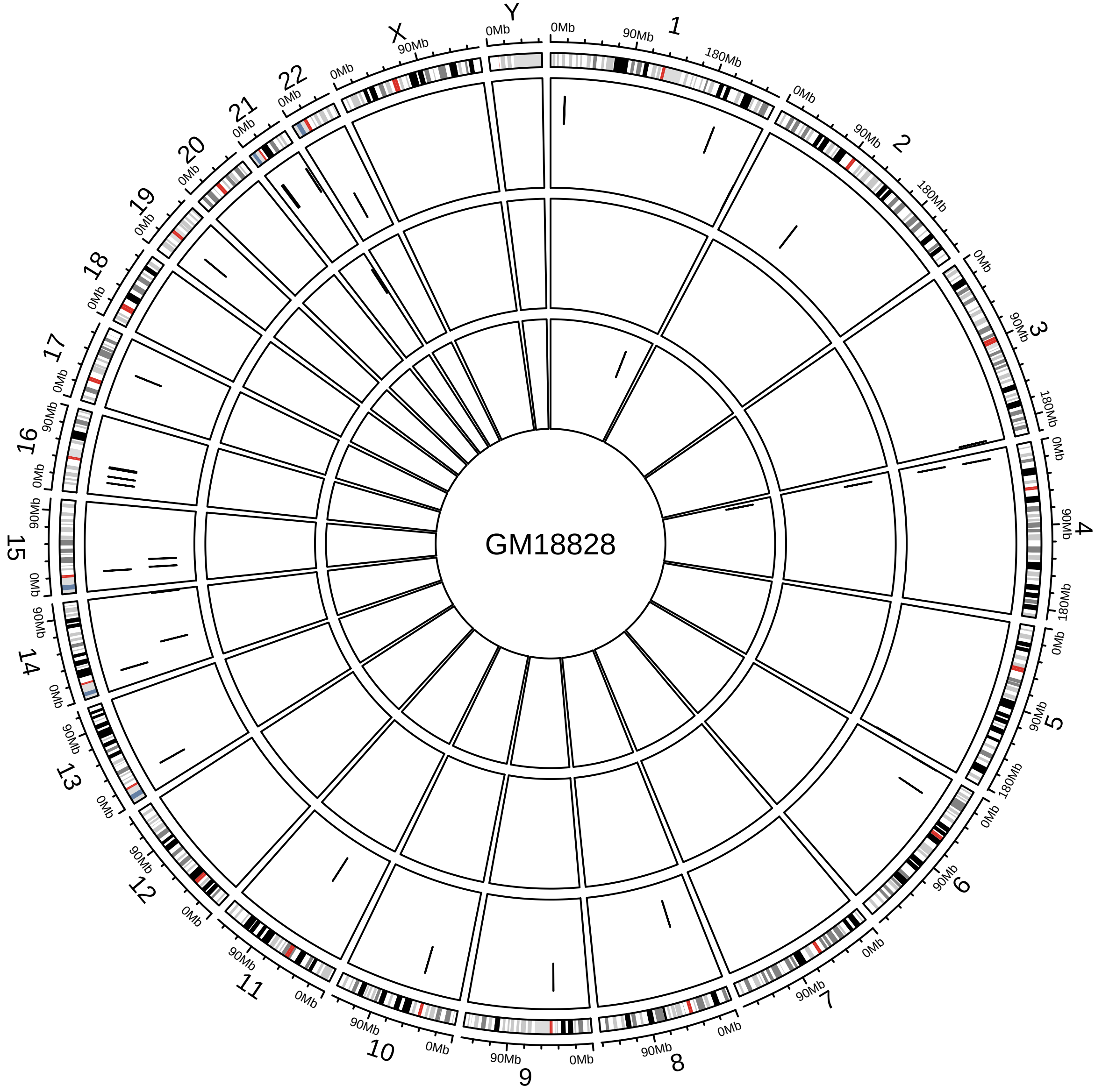
GM18828
X
Y
0Mb
90Mb
180Mb

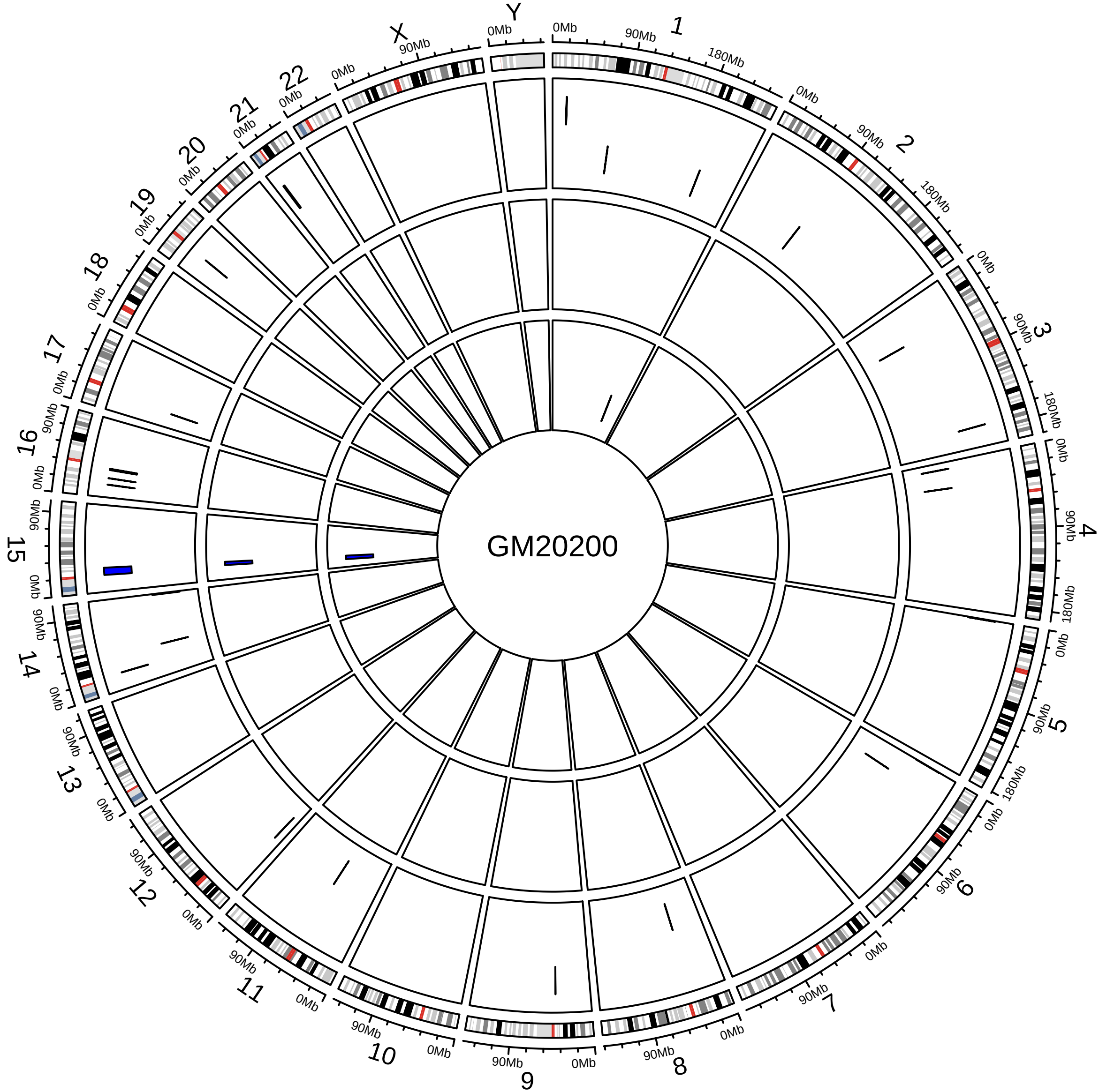
GM20200
X
Y

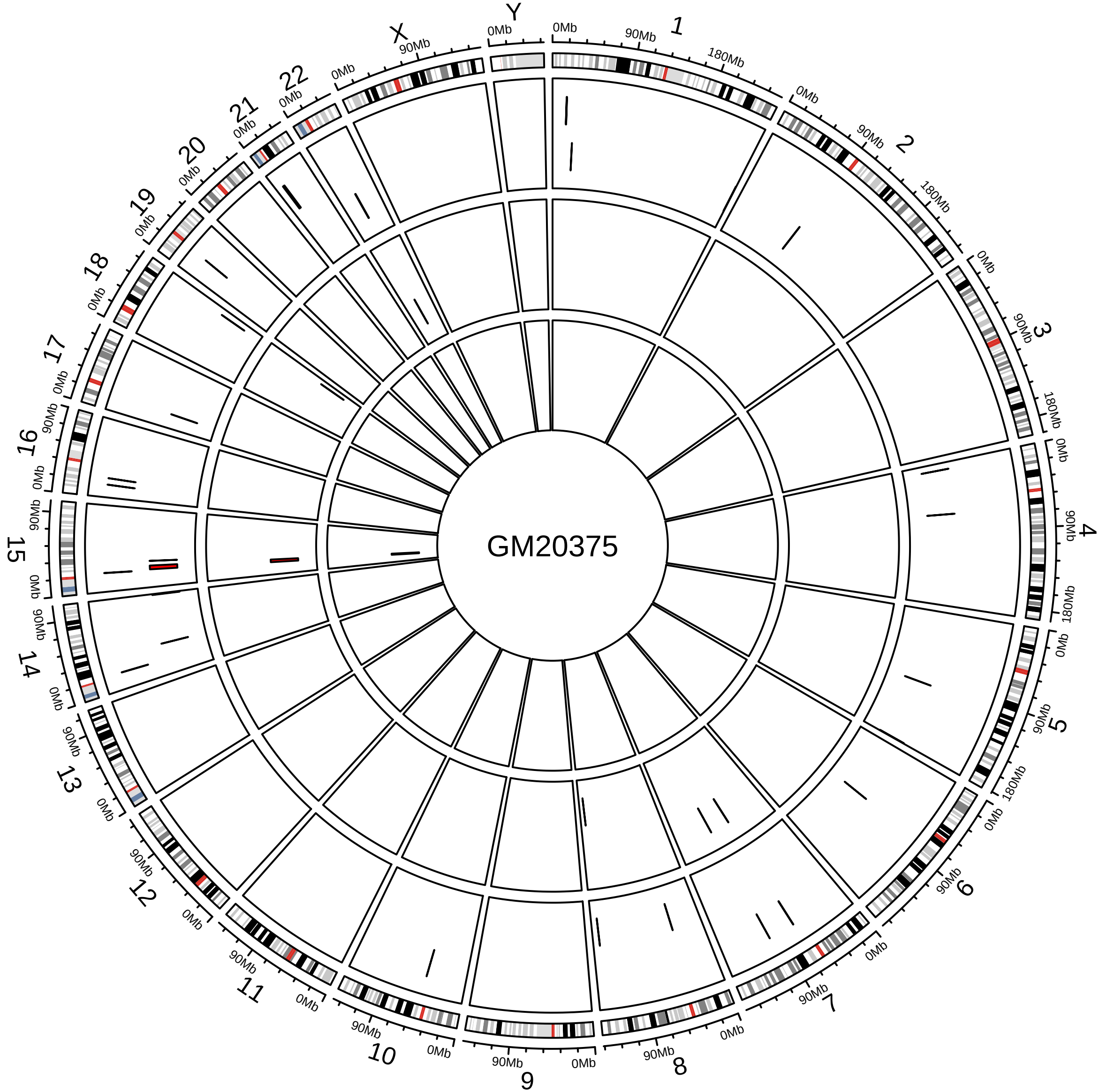
GM20375
X
Y
0Mb
90Mb
180Mb

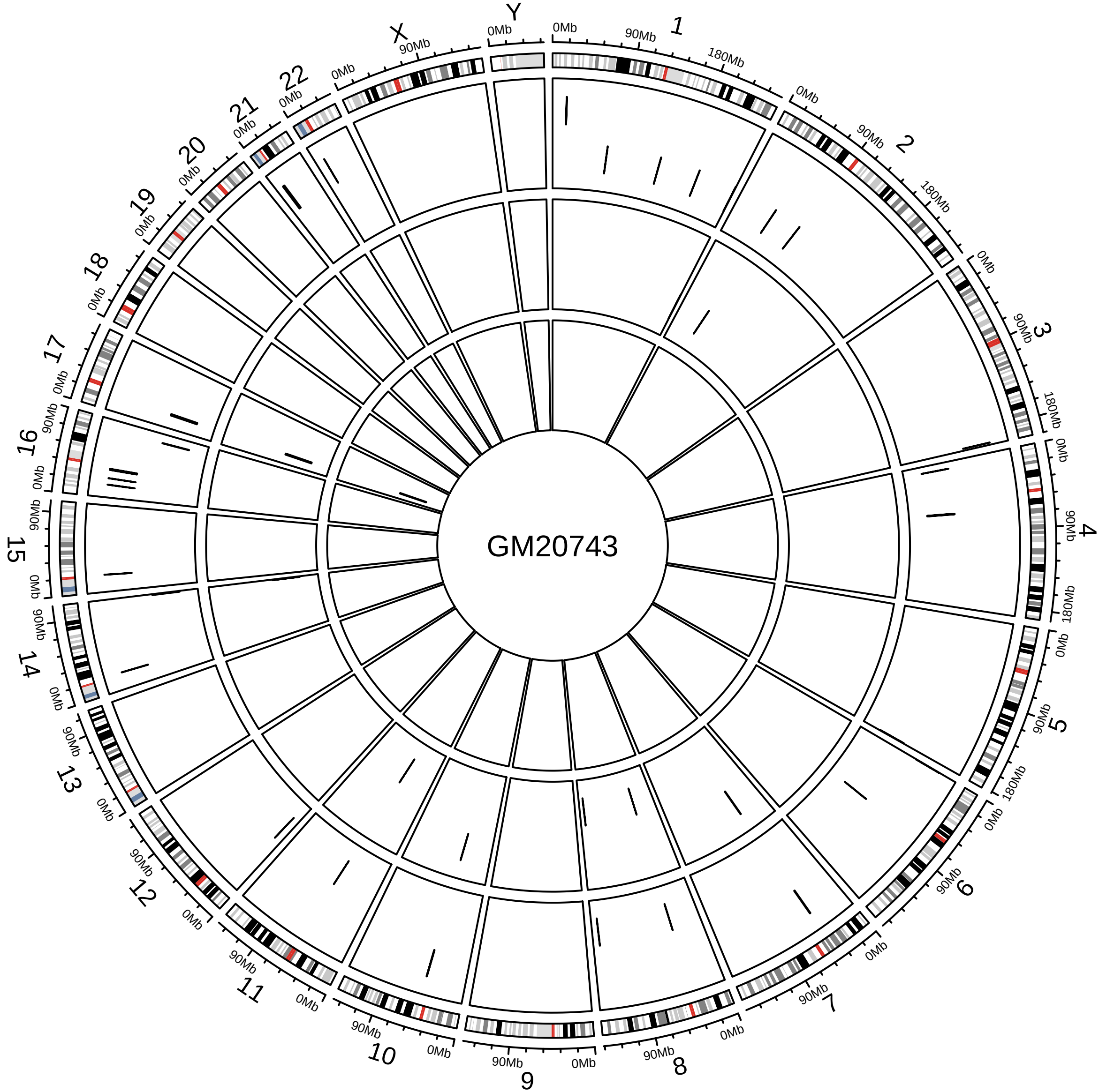
GM20743
X
Y
0Mb
90Mb
180Mb

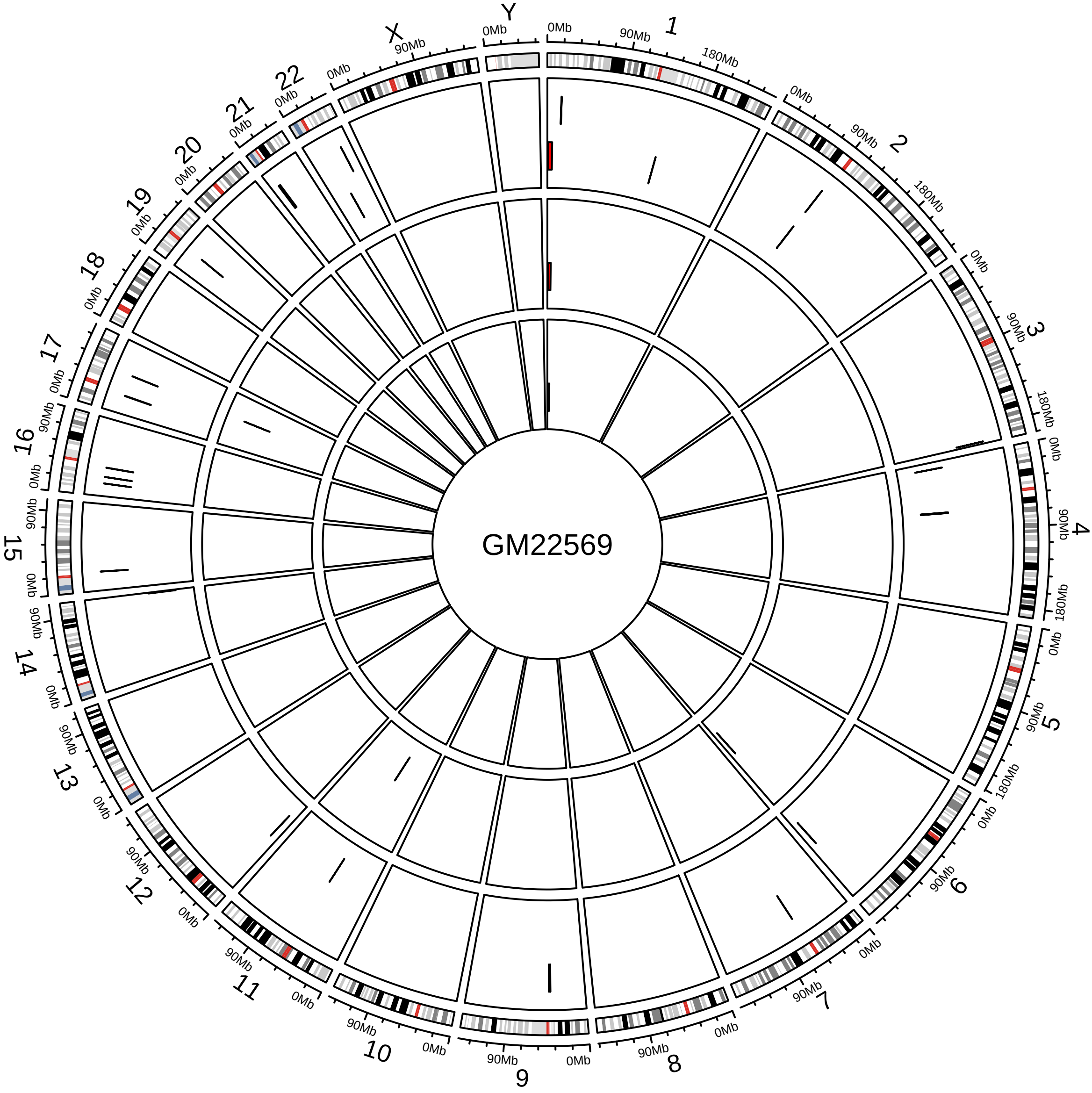
GM22569
X
Y

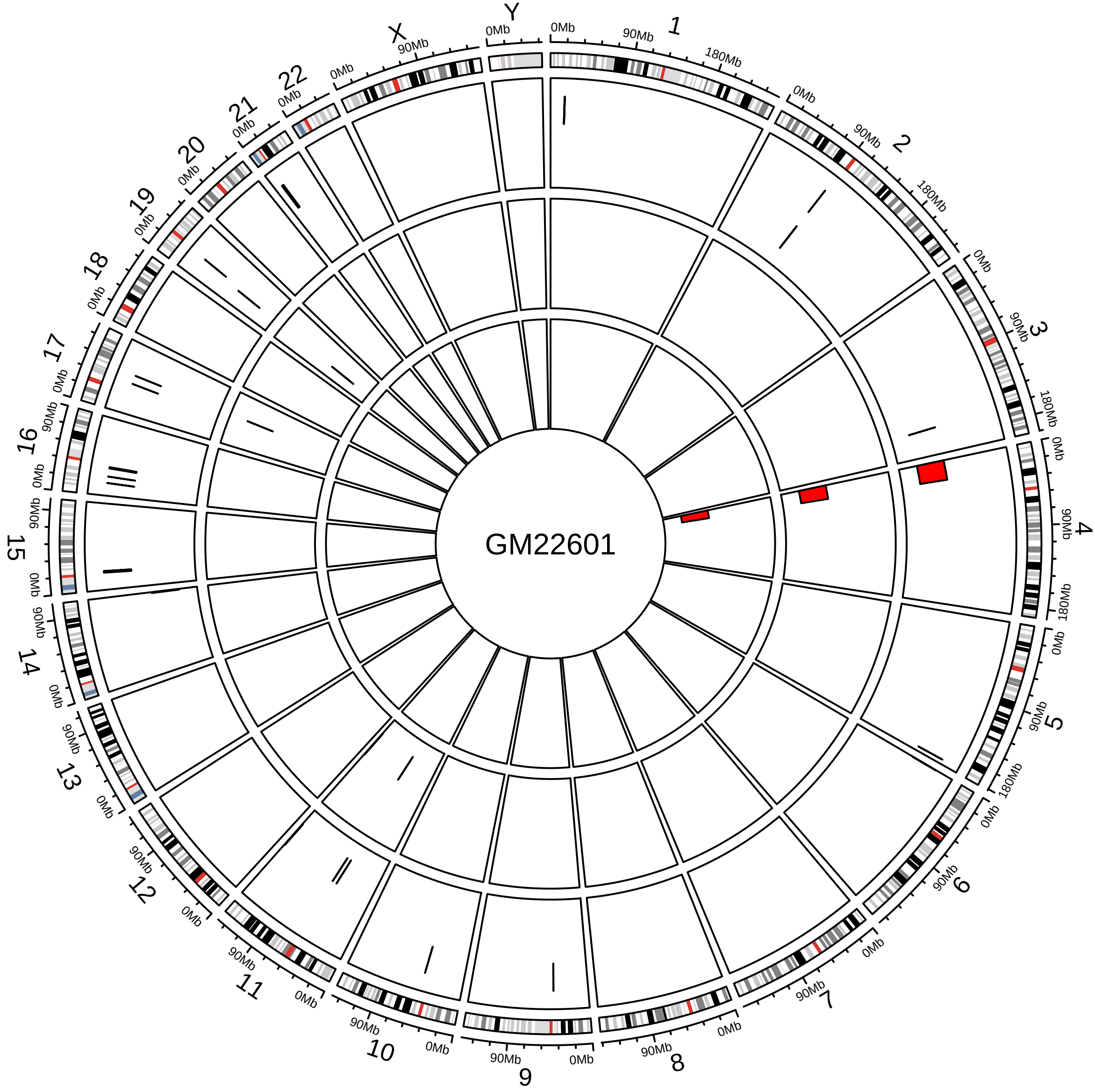

GM22601
X
Y
0Mb
90Mb
180Mb

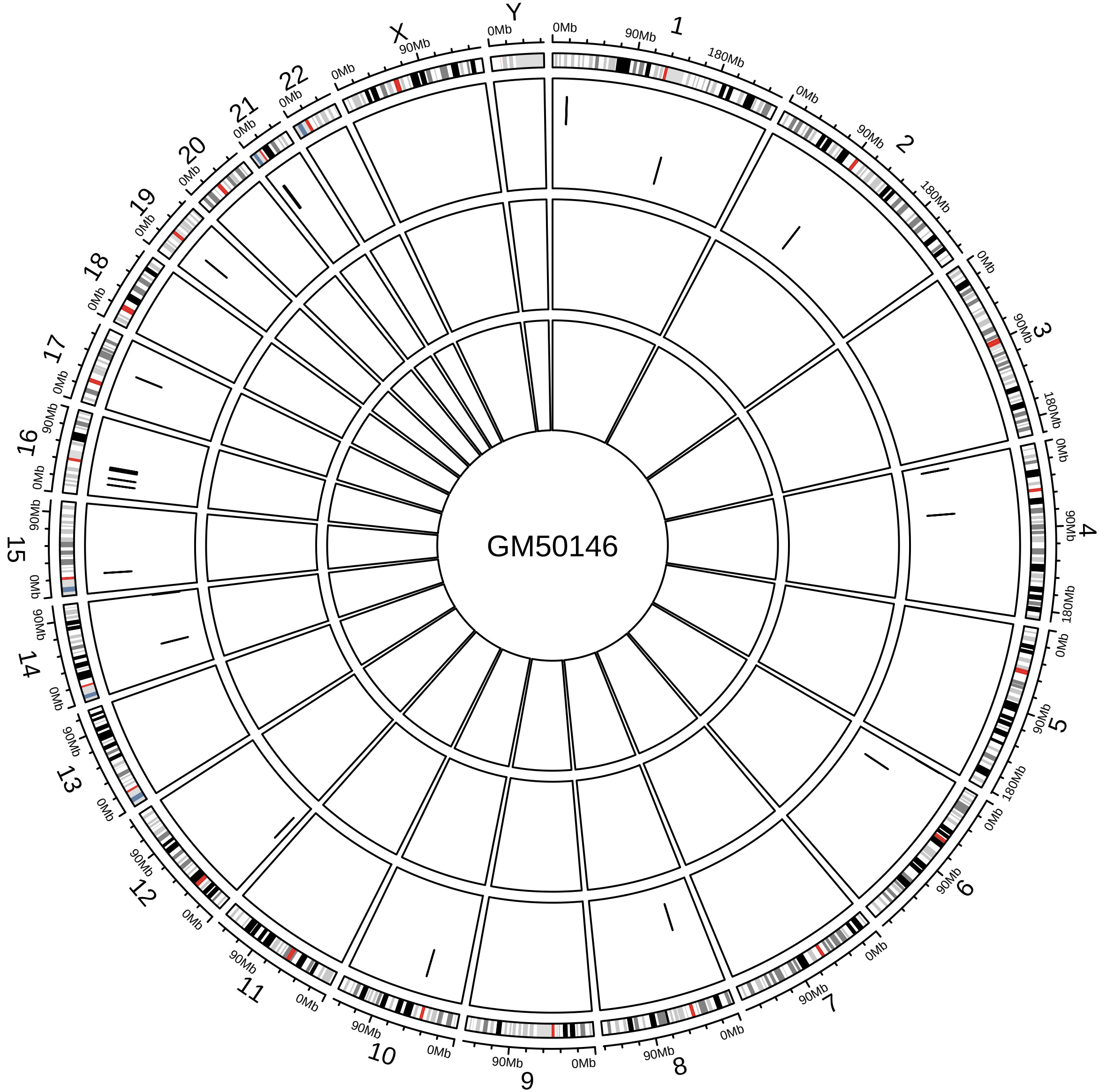
GM50146
X
Y

### Figure S3

Three deletions and three duplications that did not meet the benchmark of 50% reciprocal overlap with the JAX-CNV calls, but they were still located in the same regions with either smaller or larger size ones.

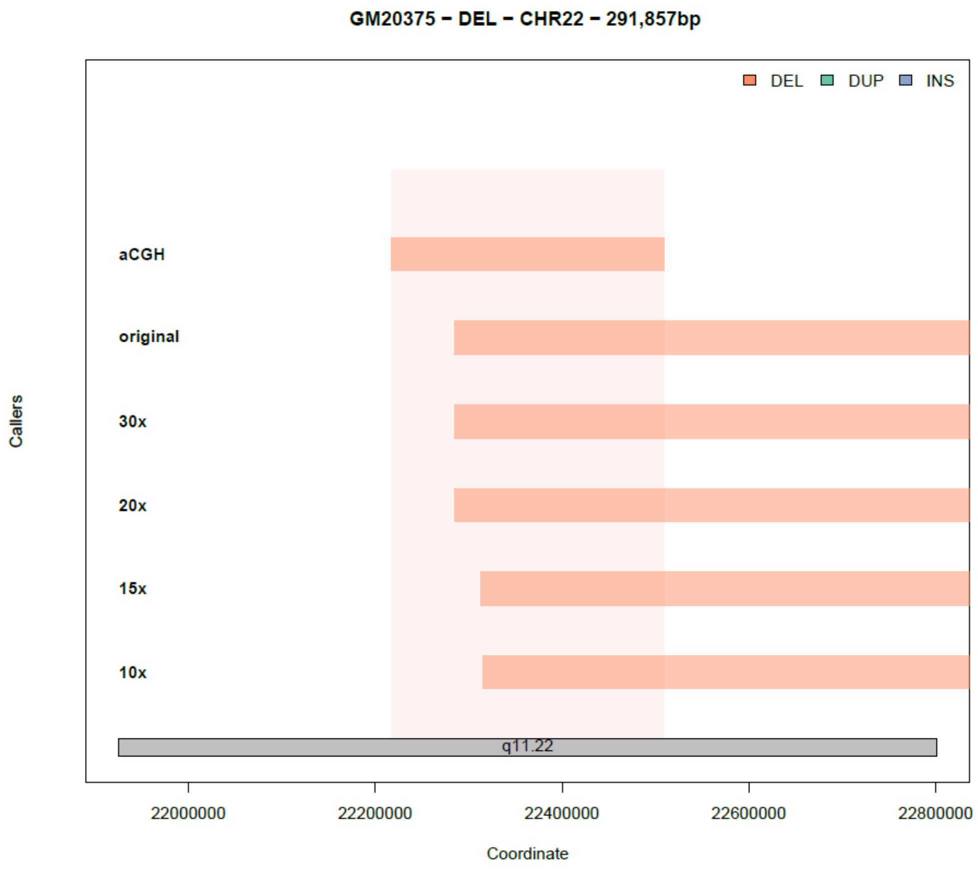

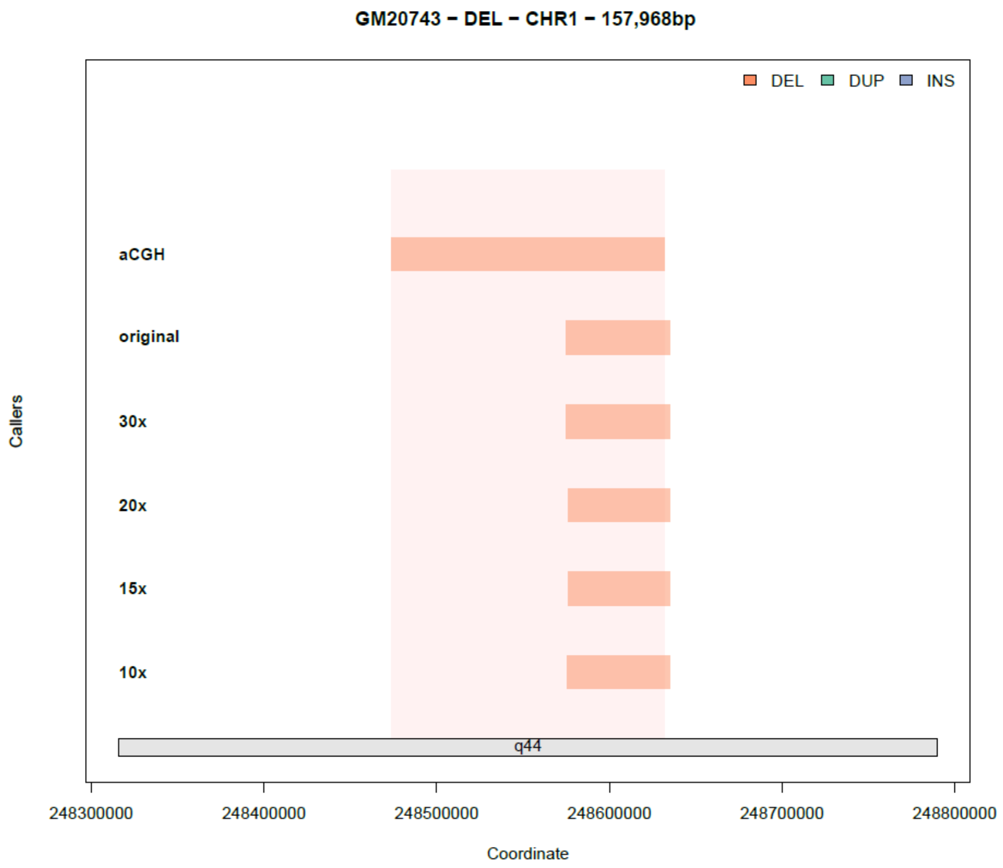
GM20743 – DEL – CHR1 – 157,968bp
DEL
DUP
INS
aCGH
original
30x
20x
15x
10x
q44
248300000
248400000
248500000
248600000
248700000
248800000
Coordinate
Callers

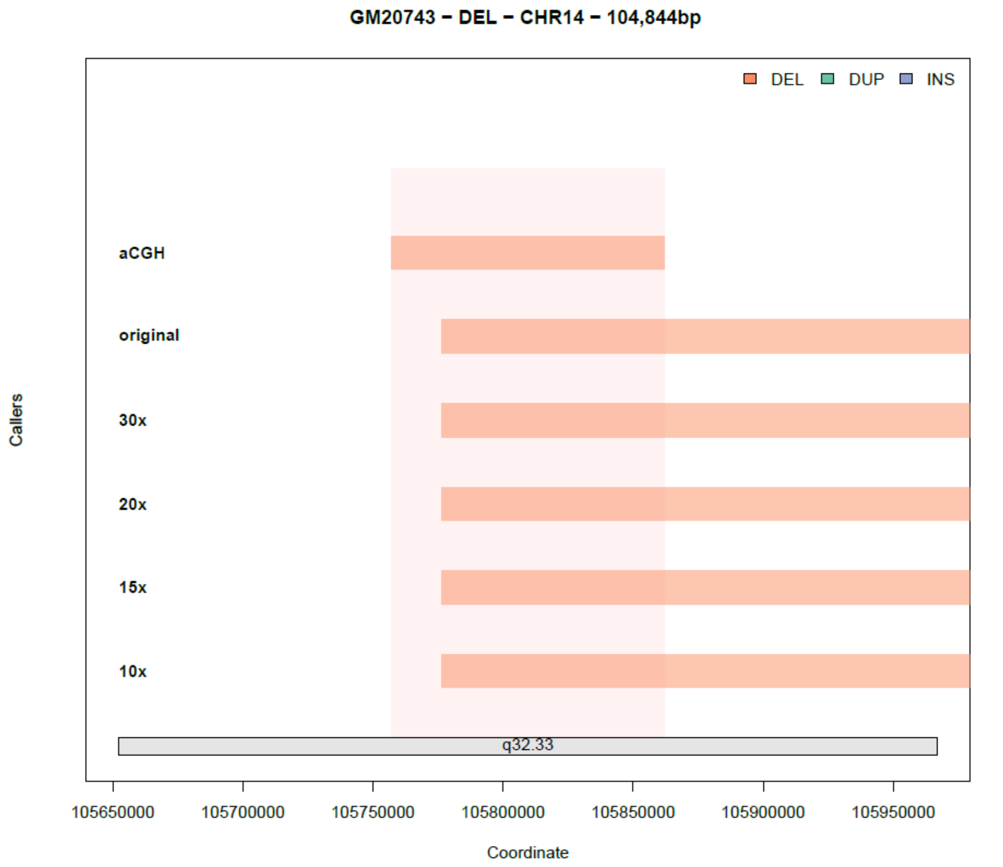
GM20743 – DEL – CHR14 – 104,844bp
DEL
DUP
INS
aCGH
original
30x
20x
15x
10x
q32.33
105650000
105700000
105750000
105800000
105850000
105900000
105950000
Coordinate
Callers

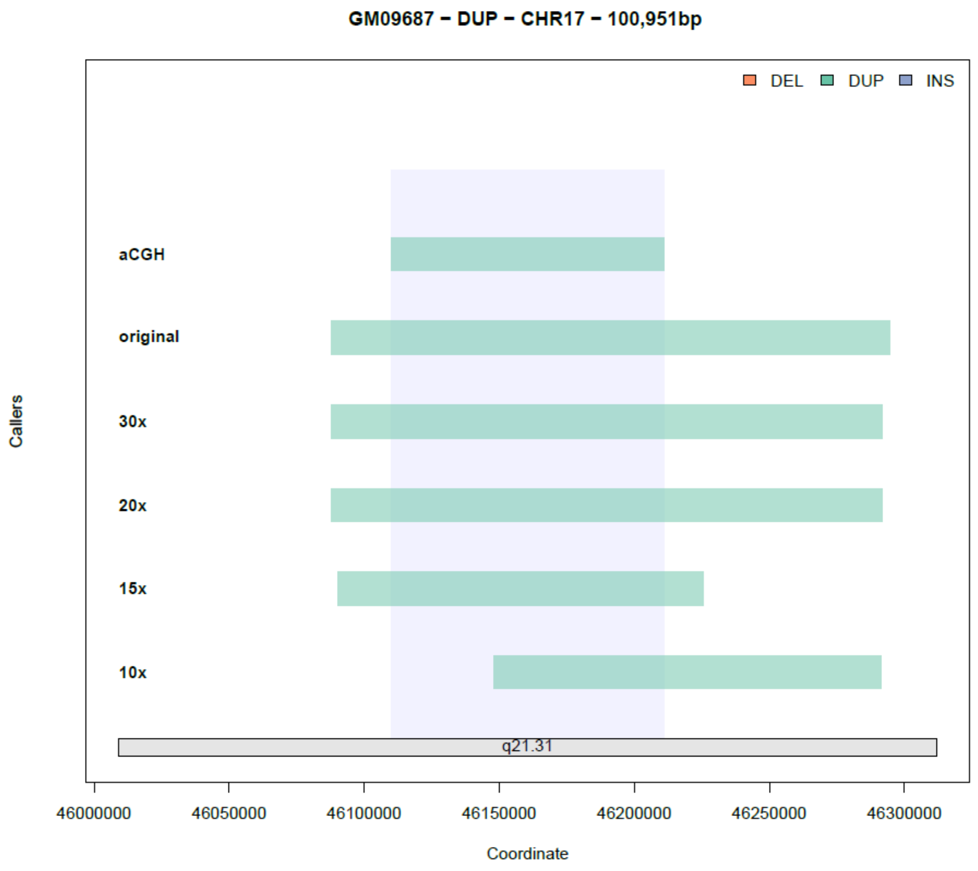
GM09687 − DUP − CHR17 − 100,951bp
DEL
DUP
INS
aCGH
original
30x
20x
15x
10x
q21.31
Callers
46000000
46050000
46100000
46150000
46200000
46250000
46300000
Coordinate

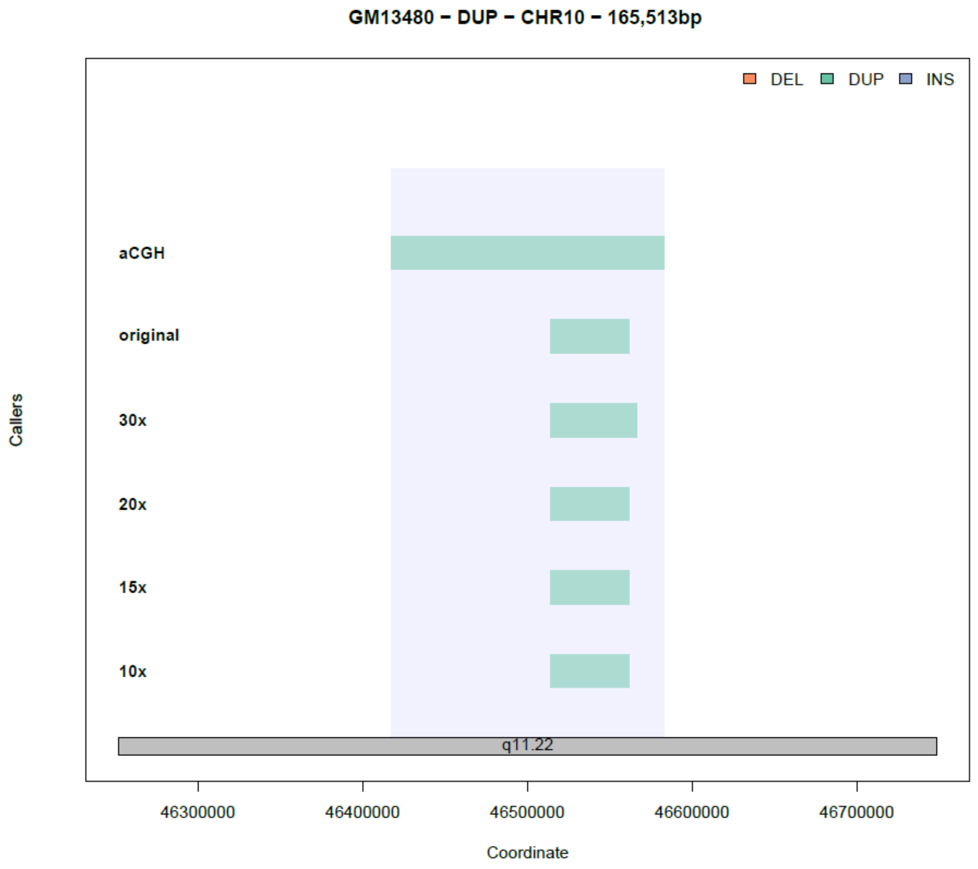
GM13480 − DUP − CHR10 − 165,513bp
DEL
DUP
INS
aCGH
original
30x
20x
15x
10x
q11.22
Callers
46300000
46400000
46500000
46600000
46700000
Coordinate

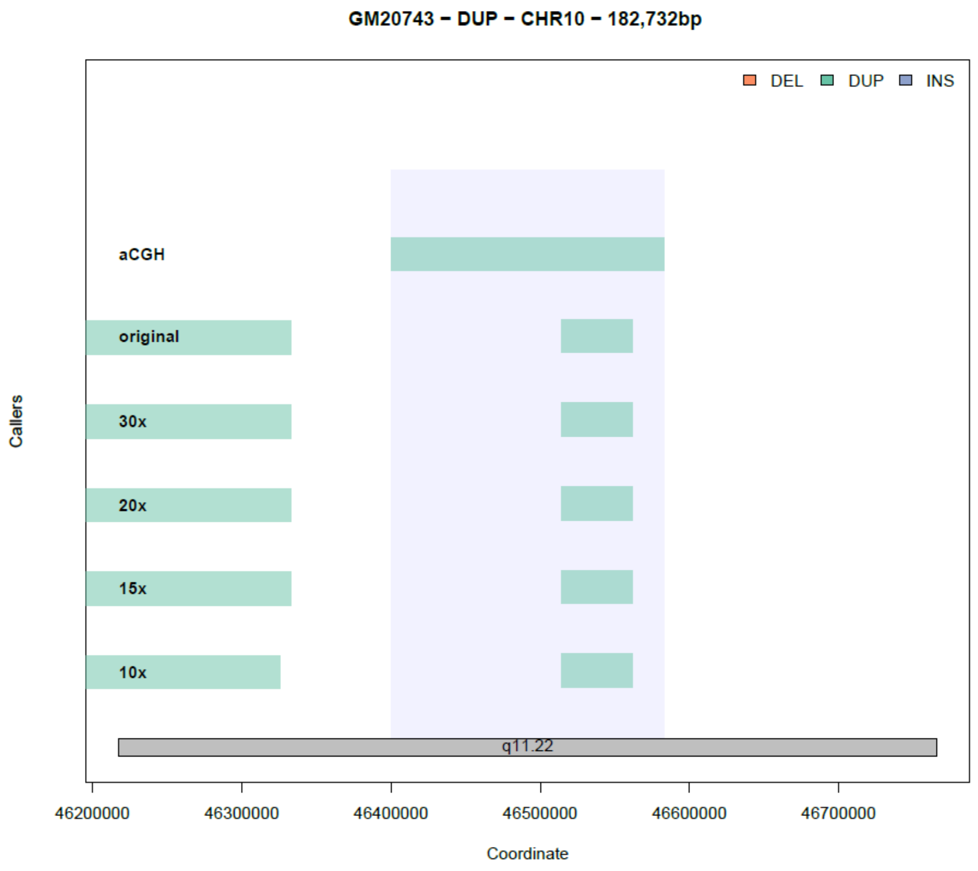

GM20743 – DUP – CHR10 – 182,732bp
DEL
DUP
INS
aCGH
original
30x
20x
15x
10x
q11.22
Callers
46200000
46300000
46400000
46500000
46600000
46700000
Coordinate

### Figure S4

20.8Mb Duplication of GM03997 that JAX-CNV uses two calls to cover it entirely.

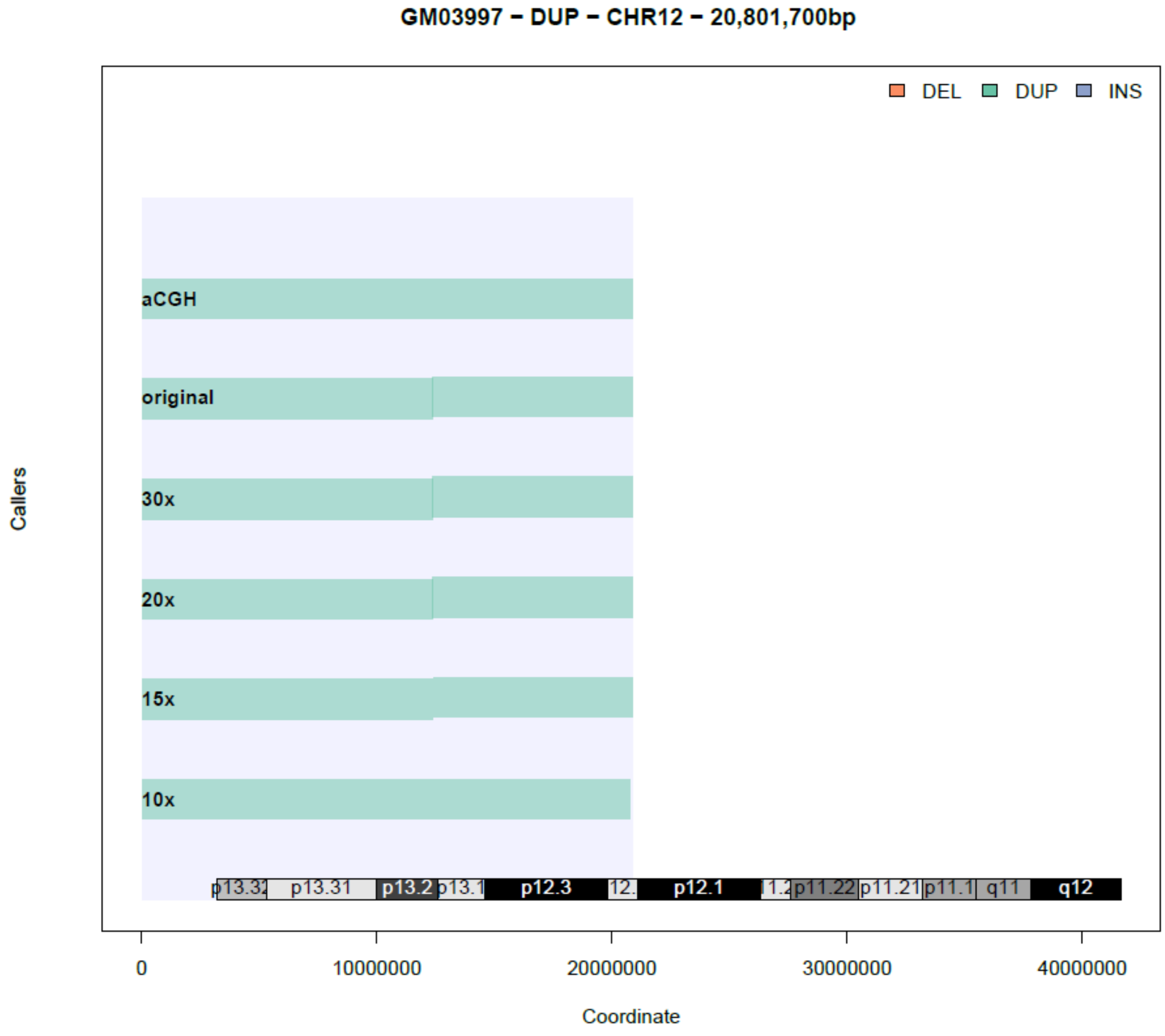

### Figure S5

A CNV that JAX-CNV cannot detect at 15x.

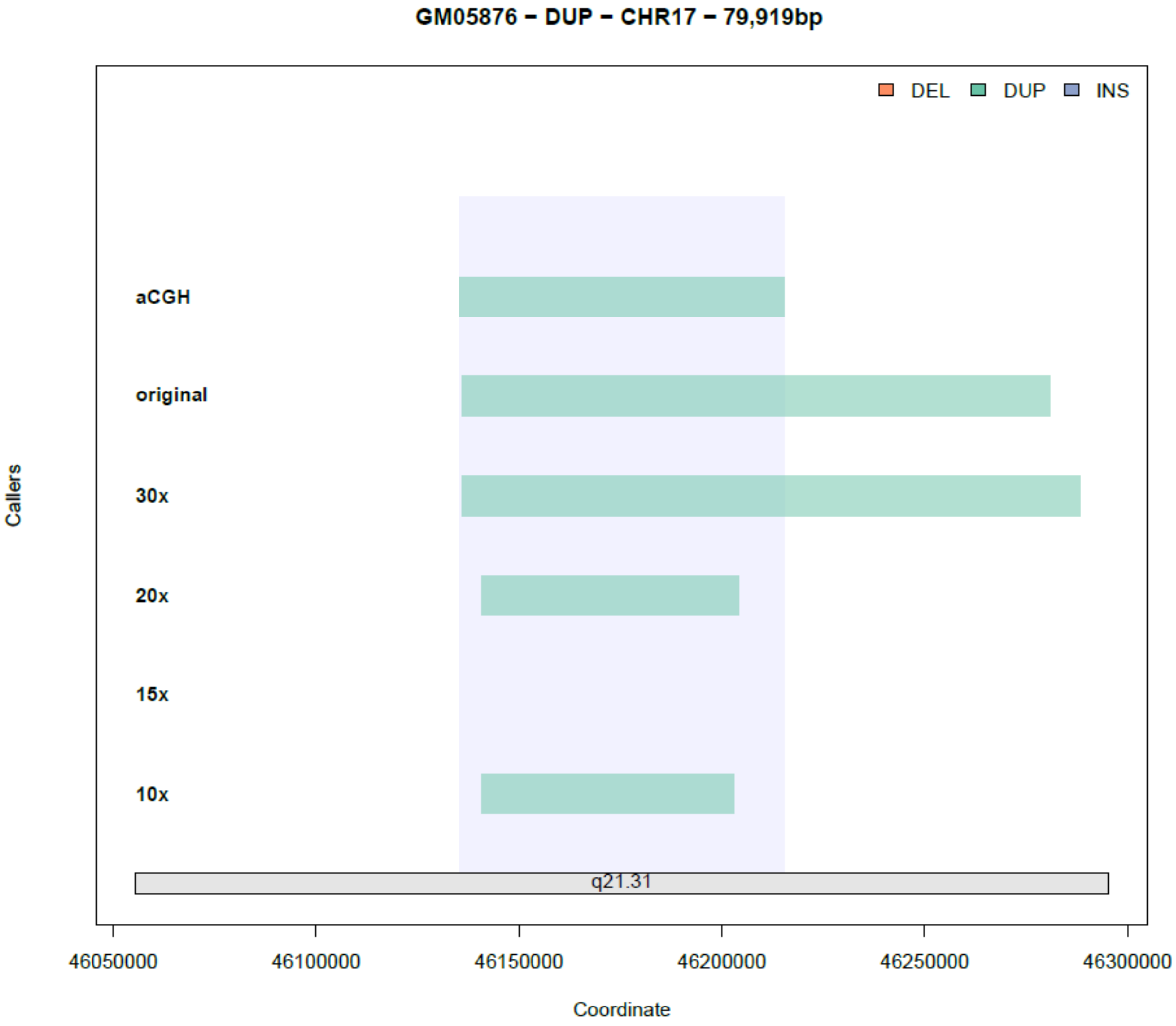

### Figure S6

CNVs (one deletion and six duplications) that JAX-CNV cannot detect at 10x.

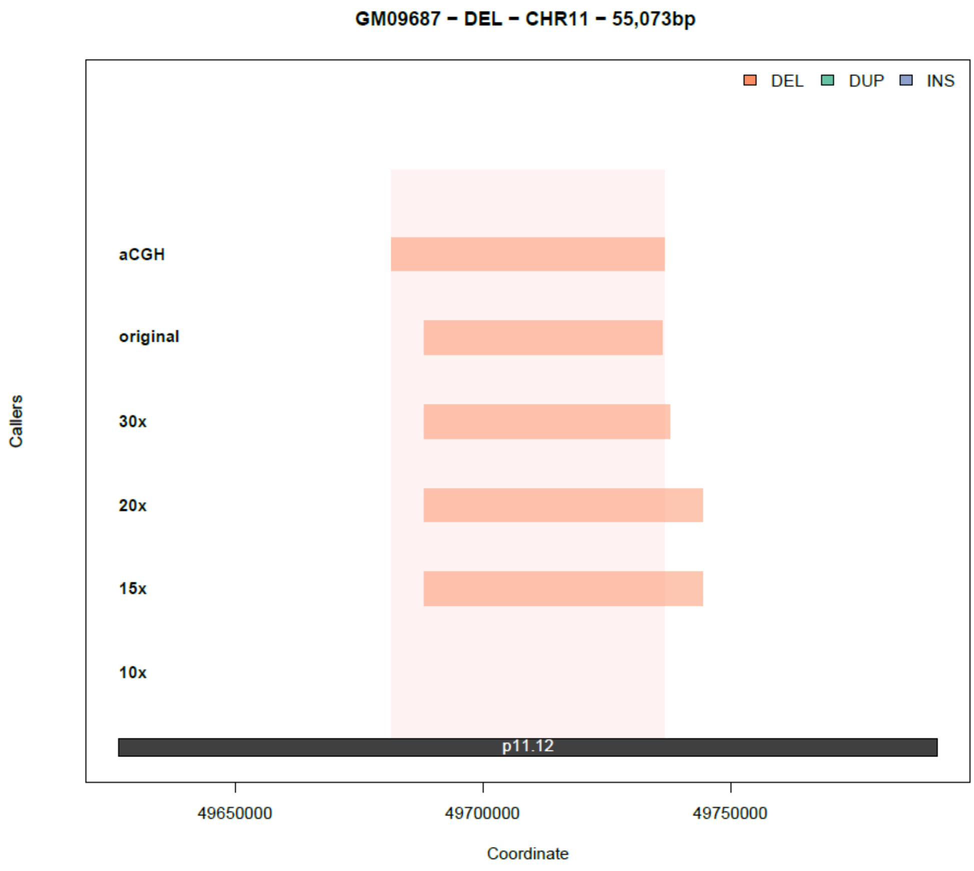

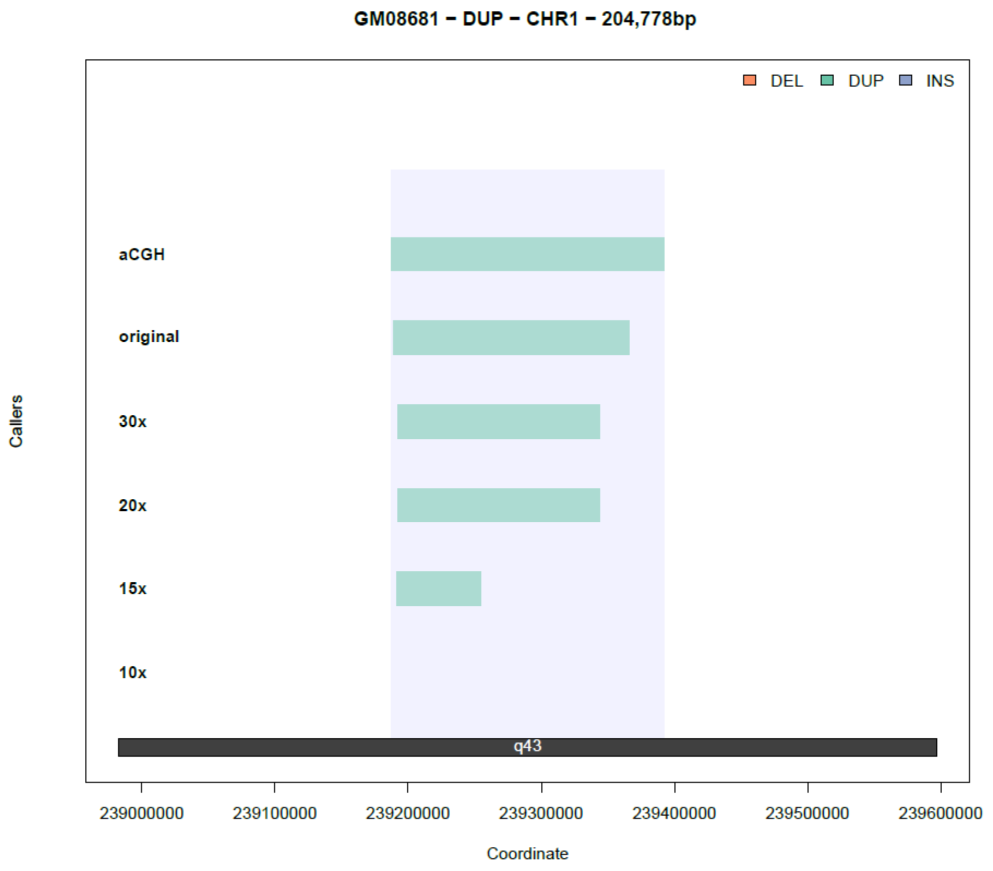

GM08681 – DUP – CHR1 – 204,778bp
DEL
DUP
INS
aCGH
original
30x
20x
15x
10x
q43
Callers
239000000
239100000
239200000
239300000
239400000
239500000
239600000
Coordinate

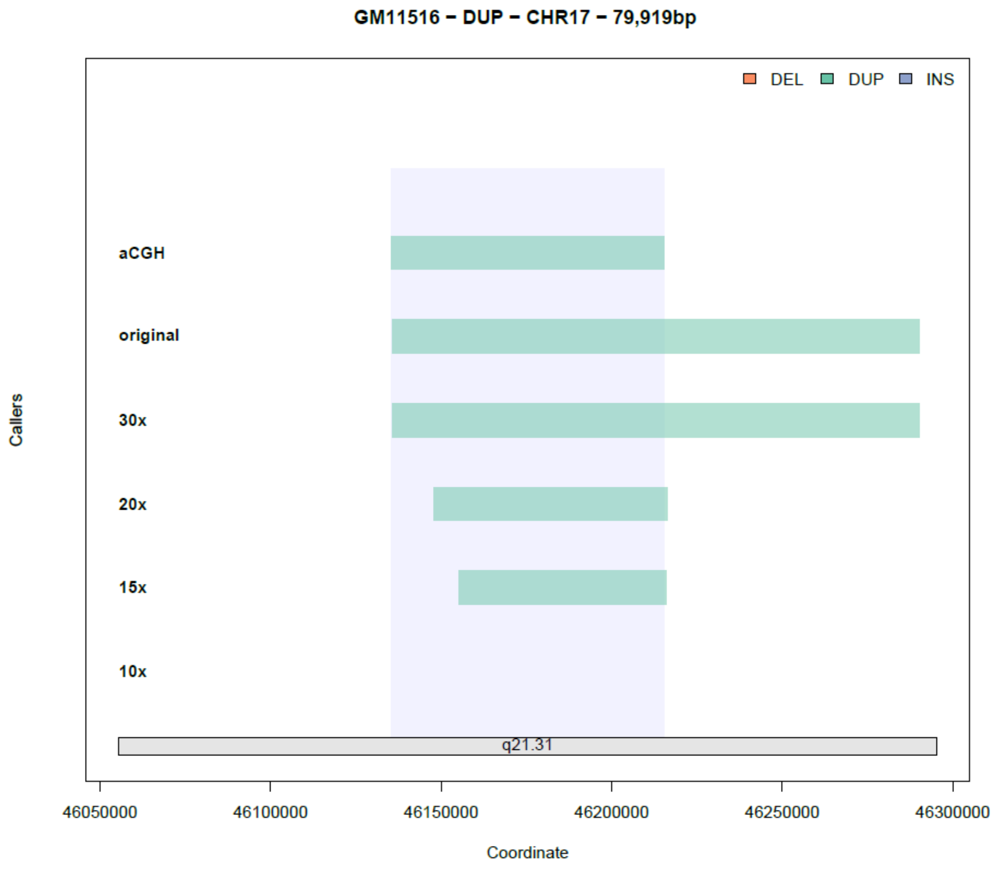

GM11516 – DUP – CHR17 – 79,919bp
DEL
DUP
INS
aCGH
original
30x
20x
15x
10x
q21.31
Callers
46050000
46100000
46150000
46200000
46250000
46300000
Coordinate

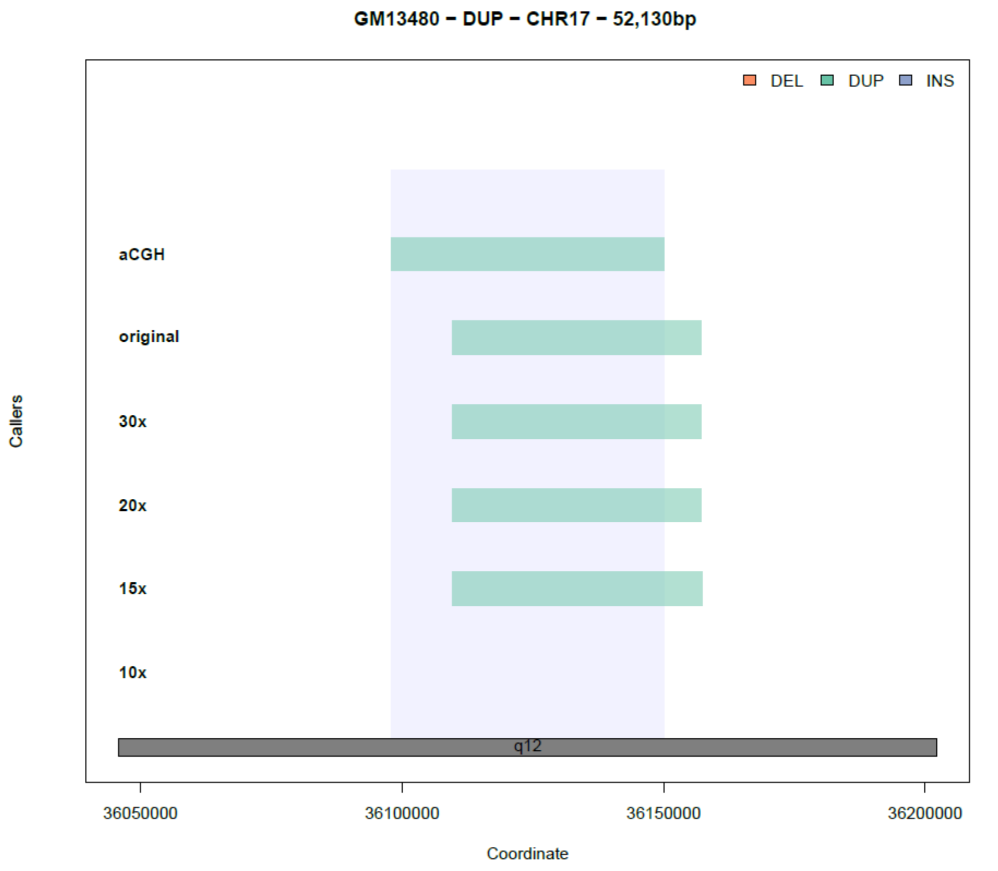
GM13480 − DUP − CHR17 − 52,130bp
DEL
DUP
INS
aCGH
original
30x
20x
15x
10x
q12
Callers
36050000
36100000
36150000
36200000
Coordinate

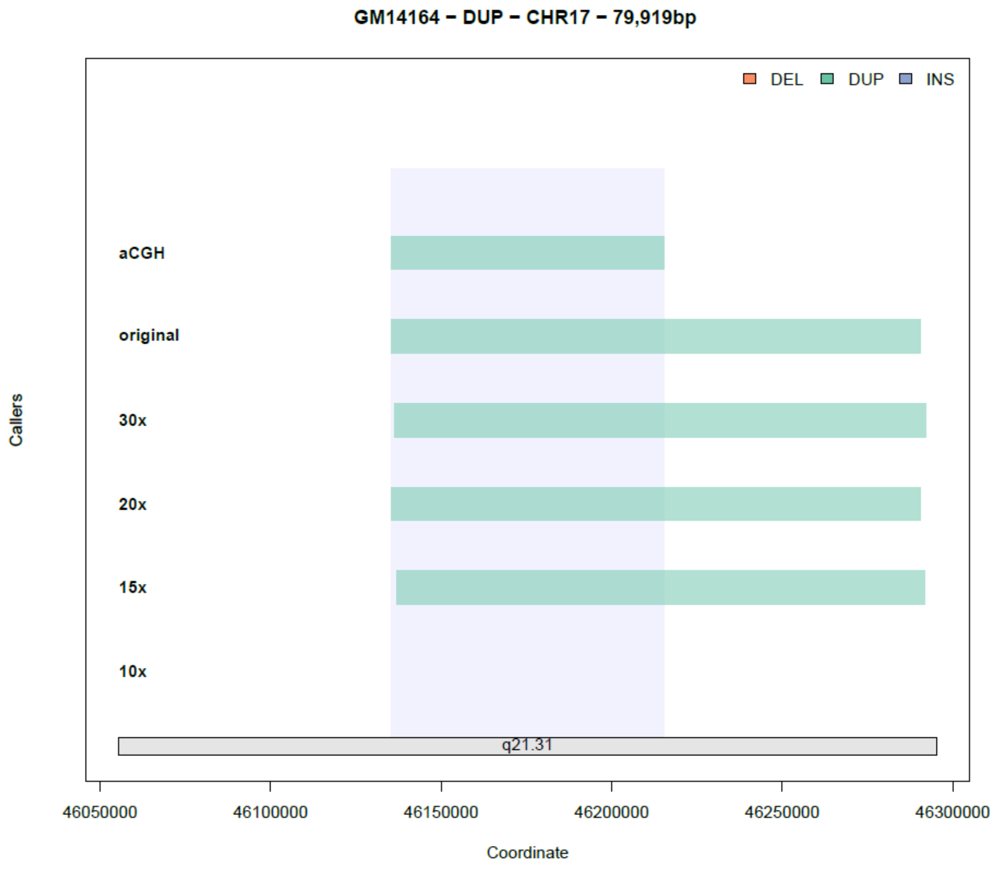
GM14164 − DUP − CHR17 − 79,919bp
DEL
DUP
INS
aCGH
original
30x
20x
15x
10x
q21.31
Callers
46050000
46100000
46150000
46200000
46250000
46300000
Coordinate

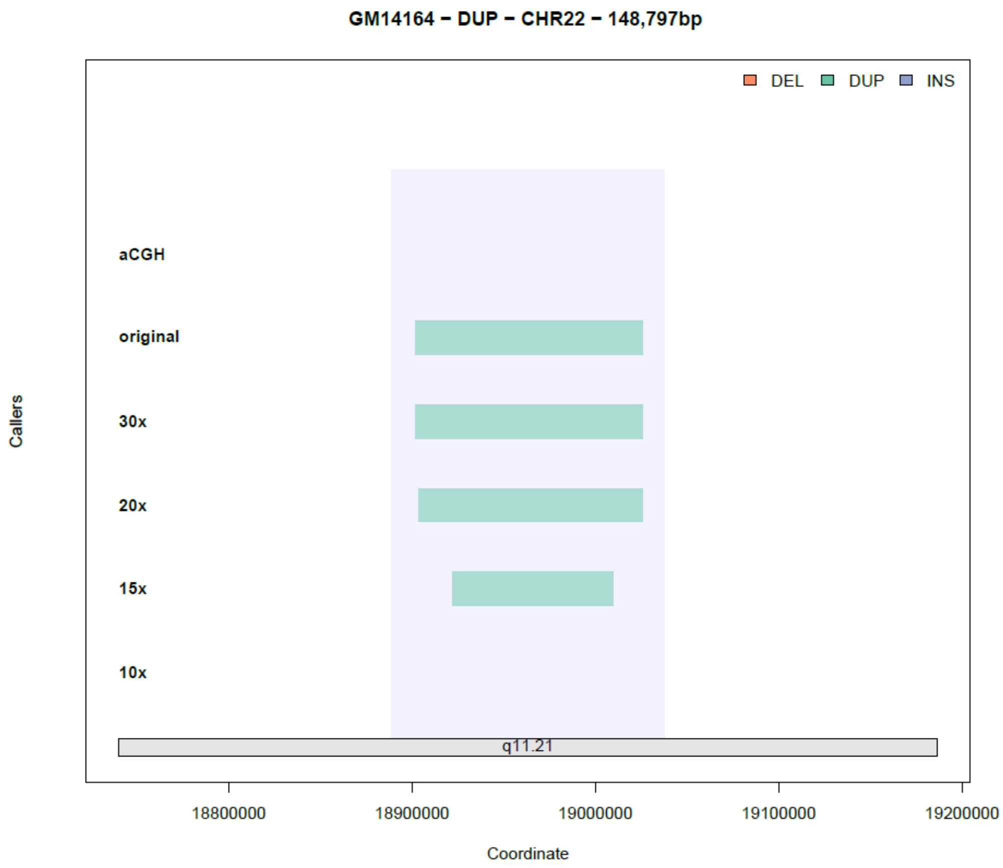
GM14164 – DUP – CHR22 – 148,797bp
DEL
DUP
INS
Callers
aCGH
original
30x
20x
15x
10x
q11.21
18800000
18900000
19000000
19100000
19200000
Coordinate

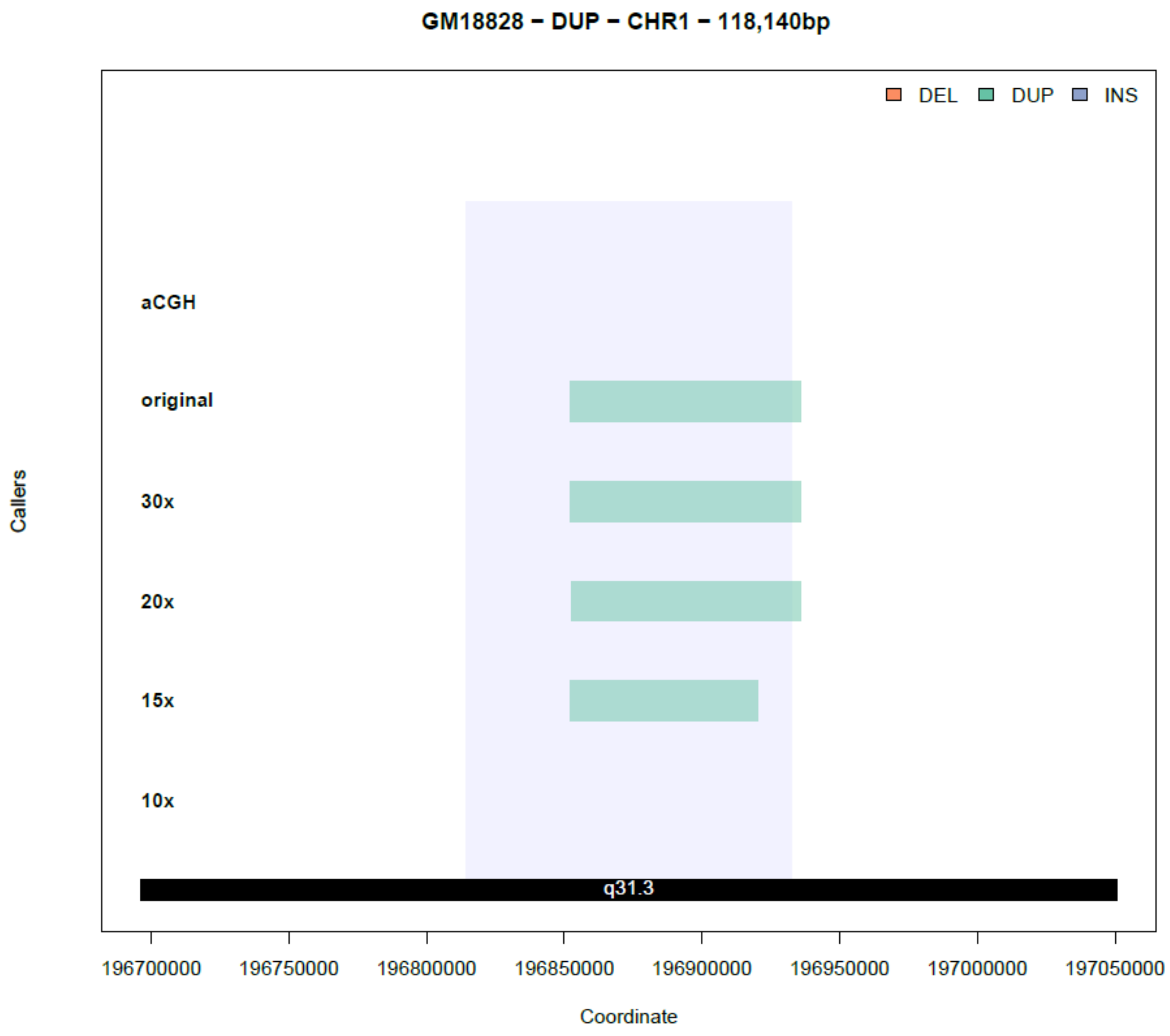
GM18828 – DUP – CHR1 – 118,140bp
DEL
DUP
INS
aCGH
original
30x
20x
15x
10x
q31.3
Callers
196700000
196750000
196800000
196850000
196900000
196950000
197000000
197050000
Coordinate

### Figure S7

**A.** The recalled CNVs in the truth set of each algorithm. **B.** Numbers of total calls reported by each algorithm.

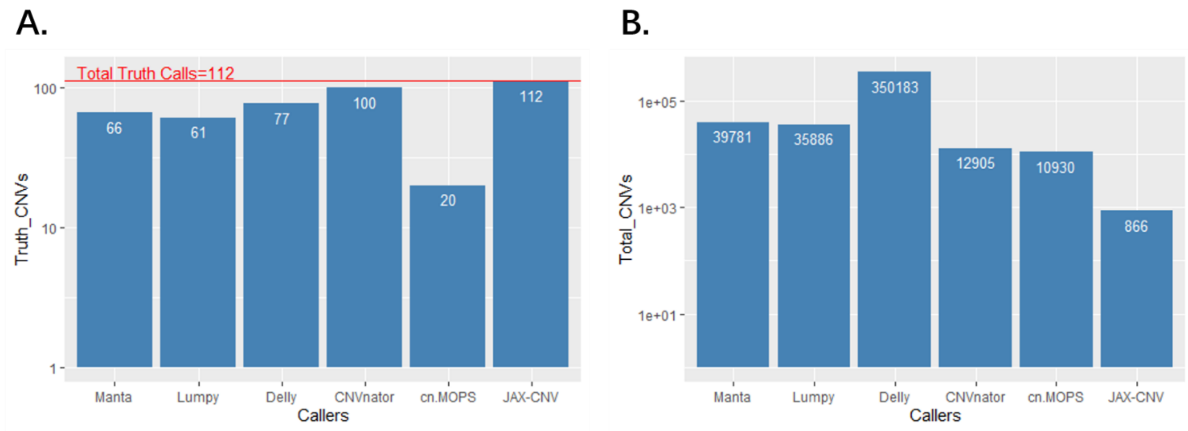
