## Supplemental Materials and Methods for "JAX-CNV: A whole genome sequencing-based algorithm for copy number detection at clinical grade level"

1. Precision Medicine Center, The First Affiliated Hospital of Xi’an Jiaotong University, Xi'an 710061, China
2. The Jackson Laboratory for Genomic Medicine, Farmington, CT 06032, USA
3. School of Cyber Science and Engineering, Xi'an Jiaotong University, Xi'an 710049, China
4. Department of Pathology and Laboratory Medicine, Perelman School of Medicine, University of Pennsylvania, Philadelphia, PA 19104, USA
5. School of Computer Science and Technology, Faculty of Electronic and Information Engineering, Xi'an Jiaotong University, Xi'an 710049, China
6. MOE Key Lab for Intelligent Networks & Networks Security, Faculty of Electronic and Information Engineering, Xi'an Jiaotong University, Xi'an 710049, China
7. Department of Life Sciences, Ewha Womans University, Seoul 03760, South Korea
8. Corresponding authors

*: These authors contributed equally to this work.

Content

### K-mer selection

Using larger k-mer will increase uniqueness (higher sensitivity) of each k-mer. However, the improvement will saturate and processing larger k-mer requests more computational resource. For example, Jellyfish may use more than 60GB memory for 30-mer.

We choose 25-mer since it maintains good sensitivity and consumes reasonable computational resource. Below is the figure to show the percentage of unique k-mer (gray colored) by applying different k value. Using 5-mer or 10-mer, there is no unique kmer against the human reference genome. From 15-mer to 20-mer, we see a significant increase of unique k-mers that saturates from 25-mer to 30-mer. Since the sensitivity of JAX-CNV does not reply on single factor of uniqueness of a k-mer, the sensitivity was not affected using k-mer larger than 25-mer.

|  | 2^0 | 2^1 | 2^2 | 2^3 | 2^4 | 2^5 | 2^6 | 2^7 | 2^8 | 2^9 | 2^10 | 2^11 | 2^12 | 2^13 | 2^14 | 2^15 | 2^16 | 2^17 | 2^18 | 2^19 |
| --- | --- | --- | --- | --- | --- | --- | --- | --- | --- | --- | --- | --- | --- | --- | --- | --- | --- | --- | --- | --- |
| 5mer | 0 | 0 | 0 | 0 | 0 | 0 | 0 | 0 | 0 | 0 | 0 | 0 | 0 | 0 | 0 | 0 | 578869 | 3169190 | 46460986 | 56553898 |
| 10mer | 0 | 30 | 835 | 9887 | 178875 | 1155671 | 2865023 | 16760752 | 59822879 | 67965400 | 151394980 | 683944650 | 871720105 | 470394196 | 192729150 | 81946849 | 71169379 | 39055631 | 26246468 | 2187465 |
| 15mer | 163920459 | 350468682 | 617833859 | 574589193 | 324629161 | 166582851 | 96390322 | 65481295 | 53160926 | 47474605 | 46490414 | 50790238 | 50565850 | 38844029 | 27520221 | 28733922 | 23875721 | 14228962 | 1331864 | 0 |
| 20mer | 2008119761 | 193942668 | 77398660 | 53358732 | 44027424 | 39241604 | 37200052 | 34939754 | 33176902 | 35466584 | 38010242 | 43274070 | 35790060 | 25156243 | 17544093 | 18794778 | 4737322 | 4146546 | 130668963 | 0 |
| 25mer | 2189224507 | 138460167 | 62798433 | 43638381 | 35933207 | 31883042 | 30222722 | 27662831 | 26462814 | 29725979 | 32753925 | 35309173 | 26567686 | 14628926 | 11058147 | 6954616 | 529509 | 131174973 | 0 | 0 |
| 30mer | 2282436171 | 129205464 | 55787733 | 37769058 | 30047767 | 26675857 | 24919247 | 21583997 | 20758934 | 25546421 | 28754286 | 29199080 | 17614601 | 8671580 | 4135161 | 131508752 | 370041 | 0 | 0 | 0 |
| 35mer | 2352664426 | 122290226 | 49959763 | 32911665 | 25254943 | 22055775 | 19437488 | 16467937 | 17779372 | 22420688 | 24945300 | 22423695 | 11552548 | 3664916 | 130723850 | 426604 | 0 | 0 | 0 | 0 |
| 40mer | 2407668032 | 115893200 | 44825424 | 28602320 | 20972327 | 17759233 | 14687694 | 12864344 | 15591906 | 20463881 | 21440584 | 16883961 | 136417090 | 697998 | 206515 | 0 | 0 | 0 | 0 | 0 |
|  | 0.698476 |  |  |  |  |  |  |  |  |  |  |  |  |  |  |  |  |  |  |  |

### DBSCAN

Specifically, we first sort the candidate CNV fragments based on their positions. Then, we separate the fragments into different raw clusters by two conditions: a. the distances of any two continuous fragments are < 3,000,000bp; b. the type (e.g. loss, gain) of all fragments located in the raw cluster region are the same. Next, for each raw cluster, we calculate the distance of every continuous fragment pair fi and fi+1 as di,i+1=(ei+1-si)/(li+li+1), where ei+1is the end position of fi+1, siis the start position of fi, and li and li+1 are the length of fi and fi+1. We also calculate the mean distance of the raw cluster as dmean=(E-S)/i=1Nli, where E is the end position of the raw cluster, S is the start position of the raw cluster, and N is the number of fragments in the raw cluster. To overcome the cluster bias on the raw clusters with small and sparse fragments, we set the distance of a continuous fragment pair with d>3 and discontinuous fragment pair as dmean+1. Finally, we apply dbscan function (in dbscan R package) on the distance matrix of each raw cluster with parameters eps = dmean and minPts = 2 to obtain clusters. Afterwards, we update distance matrix and dmean and iteratively apply dbscan function until the cluster results are not change. For the raw clusters with only two fragments (denoted as f1 and f2, and the position of f1 is smaller than that of f2), which can not be clustered by dbscan, we calculate three variables: y1=(s2-e1) / mean(l1,l2), y2=(s2-e1) / min(l1,l2), and y3=(s2-e1) / max(l1,l2). We cluster f1 and f2 when one of the following two conditions is satisfied: a. y1<1 and y2<3; b. y3<0.1. Each final cluster is a CNV and its type is set as the type of fragments in the corresponding raw cluster.

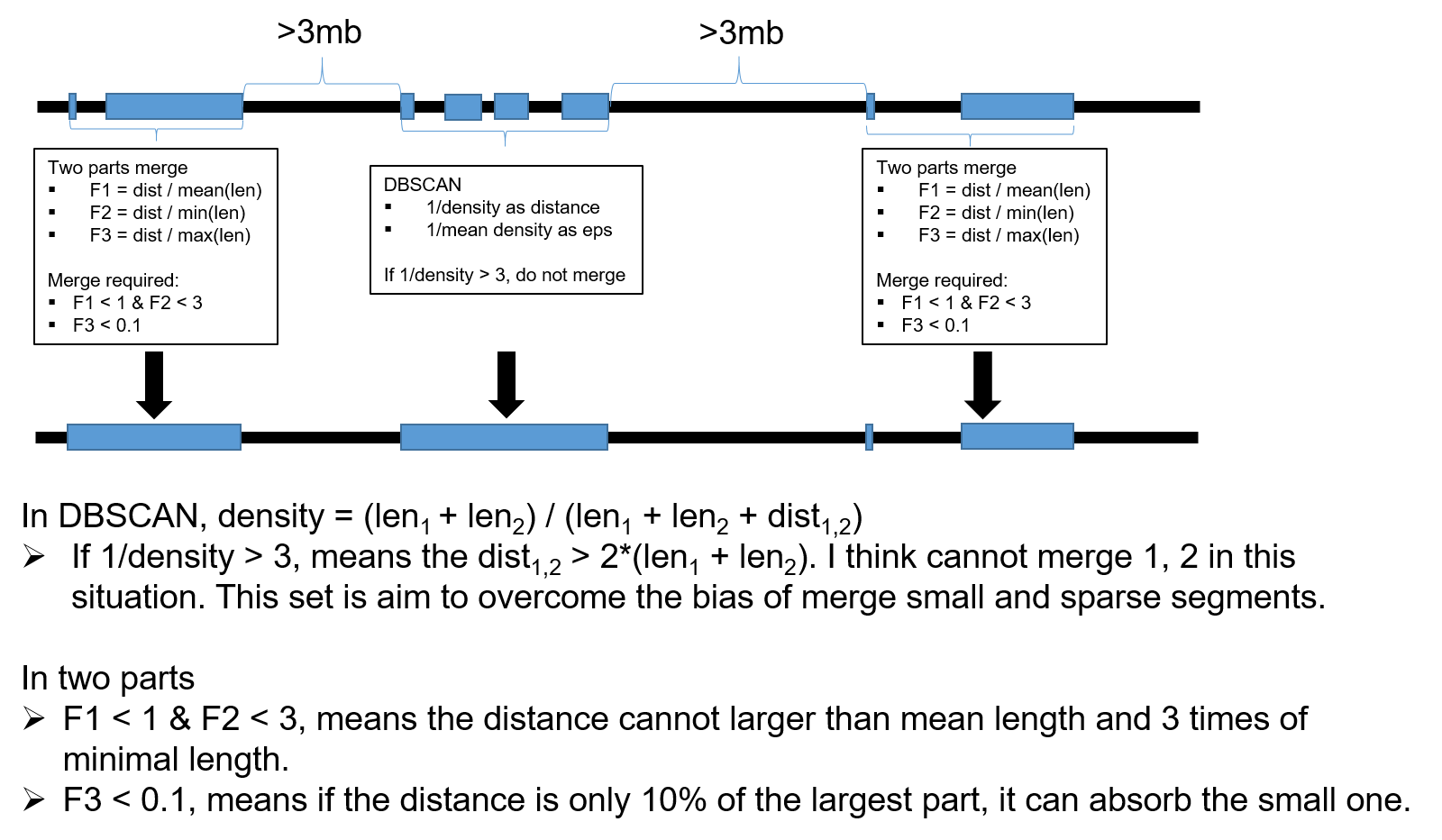

### Droplet Digital PCR (ddPCR) Validation

The ddPCR reactions were created following the Bio-Rad QX200™ system manufacturer protocol. A total of 10ng DNA template was mixed with a 2X ddPCR SuperMix for Probes (no dUTP), HindIII-HF enzyme (2U/reaction) (New England BioLabs, MA, USA), 20X primer/probe, (both FAM and HEX-labeled probes) and water to a final volume of 22 μL. Droplets were generated on the AutoDG™ using the manufacturer’s instructions in a 96-well plate, then heat-sealed with a foil seal. PCR amplification was performed using a C1000 Touch thermal cycler with the following conditions for CNV detection: enzyme activation at 95°C for 10 minutes, denaturation and extension at 94°C for 30 seconds and 60°C for 1 minute for a total of 40 cycles, enzyme deactivation at 98°C for 10 minutes, finished with a 4°C hold. Once completed, the 96-well PCR plate was loaded on the QX200™ Droplet Reader. All experiments had at least two normal controls, and a no-template control (NTC) with water. All samples and controls were run in duplicate, and data from any well with less than 8,000 droplets was treated as failed QC and excluded for downstream analysis. Analysis of the ddPCR data utilized QuantaSoft™ software.

### Scripts

Manta v.1.5.0

manta-1.5.0.centos6_x86_64/bin/configManta.py \

--config manta-1.5.0.centos6_x86_64/bin/configManta.py.ini \

--bam $BAM \

--referenceFasta $FA \

--runDir $WORK_DIR

python $WORK_DIR/runWorkflow.py -m local -j 8

Lumpy v. 0.2.13

lumpy-sv/bin/lumpyexpress -B $BAM -o $OUT_DIR/$SAMPLE.vcf

Delly v. 0.7.9

```
delly call \
  -g $FA \
  -x $DELLY/excludeTemplates/human.hg38.excl.tsv \
  -o $WORK_DIR/$SAMPLE.bcf \
  $BAM
```

CNVnator v.0.3.3

CNVNATOR=CNVnator_v0.3.3/src/cnvnator

PARAM="-chrom chr1 chr2 chr3 chr4 chr5 chr6 chr7 chr8 chr9 chr10 chr11 chr12 chr13 chr14 chr15 chr16 chr17 chr18 chr19 chr20 chr21 chr22 chrX chrY -d refs/hg38/chromosomes"

$CNVNATOR -root $SAMPLE.root -tree $BAM -unique $PARAM

$CNVNATOR -root $SAMPLE.root -his 1000 $PARAM

$CNVNATOR -root $SAMPLE.root -stat 1000 $PARAM

$CNVNATOR -root $SAMPLE.root -partition 1000 $PARAM

$CNVNATOR -root $SAMPLE.root -call 1000 $PARAM > $OUT_DIR/$SAMPLE.cnvnator

cn.MOPS

We were using SVE (<https://github.com/TheJacksonLaboratory/SVE>) to run cn.MOPS.

SVE/bin/sve call -a cnmops -g hg38 -r $FA -o $OUT_DIR/ $BAM

JAX-CNV v. 0.1.0

```
JAX-CNV GetCnvSignal \
  -b $BAM -f $FA -k $KMER \
  > $OUT_DIR/$SAMPLE.bed \
  2> $OUT_DIR/$SAMPLE.err

Rscript --vanilla JAX-CNV/JaxCNVMerge.R -i $OUT_DIR/$SAMPLE.bed
```
